# Genome-wide meta-analysis of insomnia in over 2.3 million individuals implicates involvement of specific biological pathways through gene-prioritization

**DOI:** 10.1101/2020.12.07.20245209

**Authors:** Kyoko Watanabe, Philip R. Jansen, Jeanne E. Savage, Priyanka Nandakumar, Xin Wang, 23andMe Research Team, David A. Hinds, Joel Gelernter, Daniel F. Levey, Renato Polimanti, Murray B. Stein, Eus J.W. Van Someren, August B. Smit, Danielle Posthuma

## Abstract

Insomnia is a heritable, highly prevalent sleep disorder, for which no sufficient treatment currently exists. Previous genome-wide association studies (GWASs) with up to 1.3 million subjects identified over 200 associated loci. This extreme polygenicity suggested many more loci to be discovered. The current study almost doubled the sample size to over 2.3 million individuals thereby increasing statistical power. We identified 554 risk loci (confirming 190 previously associated loci and detecting 364 novel), and capitalizing on this large number of loci, we propose a novel strategy to prioritize genes using external biological resources and information on functional interactions between genes across risk loci. Of all 3,898 genes naively implicated from the risk loci, we prioritize 289. For these, we find brain-tissue expression specificity and enrichment in specific gene-sets of synaptic signaling functions and neuronal differentiation. We show that the novel gene prioritization strategy yields specific hypotheses on causal mechanisms underlying insomnia, which would not fully have been detected using traditional approaches.

## Main

Insomnia is a sleep disorder characterized by difficulty falling or remaining asleep. It is highly prevalent in the population^1^ and associated with high morbidity, mortality^2^ and societal costs^3^. It is moderately heritable (*h*^*2*^=38-59%^4^), and recent genome-wide association studies (GWASs) have led to improved understanding of the complex polygenic etiology of insomnia^5–7^. A recent GWAS based on a sample size of >1.3 million individuals reported over 200 genomic loci linked to the risk of insomnia^6^. Together, these loci explained around a quarter of the estimated heritability of insomnia, implicated the involvement of several neurobiological processes, cell types, brain areas, and circuitries in insomnia, and showed considerable overlap with genetic risk factors for psychiatric disorders^6,7^.

Previous genetic studies have, however, also shown that insomnia is one of the most polygenic traits^8^, predicted to require at least 50 million individuals to detect SNPs at the level of genome-wide significance (*p*<5e-8) to explain at least 90% of the genetic variance (SNP heritability estimated from GWAS summary statistics)^8^. With the current rapidly increasing sample sizes, we may expect to reach a sample size of 50 million in the next decade. Yet, even if we achieve this, it will be far from straightforward to separate the true causal variants and genes from the ones that are statistically associated due to being in linkage disequilibrium (LD) with the true causal ones (‘LD by-products’). Efficient separation requires additional *in silico* post-GWAS analyses followed by wet-lab functional experimentation, to advance our understanding of how the combined effects of truly causal variants disrupt biological systems and ultimately lead to insomnia.

*In silico* strategies to prioritize causal variants may focus on improving LD resolution by comparing results from cohorts with different LD patterns (i.e. due to ancestry)^9^, while wet lab strategies could involve conducting large-scale screening of candidate variants using CRISPR/Cas9 technology within a locus^10^. These strategies, however, are not always feasible due to a lack of data accessibility or a lack of informative readouts for the functional experiments. We propose an alternative *in silico* approach in which we combine statistical fine-mapping with cross-locus linking of genes, for which external data are available, and which is aimed to lead to more specific hypothesis that can be followed-up in wet-lab experiments. This approach is especially suited for GWAS’s that identify hundreds of loci. We here perform a meta-analysis of insomnia GWASs in the UK Biobank and 23andMe Inc. cohorts including over 2.3 million participants, increasing the previous largest GWAS for insomnia by 1 million. We find 554 loci, implicating 3,898 genes using standard functional annotation and gene-based methods. Using per locus fine-mapping and cross-locus linking of genes, we prioritize 289 genes from 239 loci and show association with brain specific gene expression and enrichment in gene-sets related to synaptic signaling functions and neuronal development. Since many other traits are highly polygenic like insomnia, and future GWAS studies will increasingly lead to reliable results with ever-larger sample sizes, we believe this strategy will be highly relevant for other traits as well.

We conducted a GWAS meta-analysis of insomnia in 2,365,010 individuals, including data from the UK Biobank Study (UKB; N=386,988) and 23andMe, Inc. (N=1,978,022). In the UKB cohort, insomnia was assessed using a single question (field ID 1200), which was dichotomized following previous studies^6,11^ (**Methods**), and which was validated as being a reliable proxy of insomnia disorder previously^11^. We performed a GWAS on 386,988 unrelated European subjects (109,548 cases and 277,440 controls, using UKB release May 2018), correcting for age, sex, genotyping array and the first 10 ancestry principal components (see **Methods**). In the 23andMe cohort, insomnia was defined based on multiple questions completed by online surveys (**Methods** and as previously described in the study of Jansen *et al*.^6^). The GWAS in this cohort was performed on 1,978,022 unrelated research participants of European ancestry who consented to participate in research, correcting for age, sex and the first 5 ancestry principal components (484,176 cases and 1,493,846 controls; **Methods**). In both cohorts, analyses were limited to SNPs with a minimum minor allele count (MAC) of 100. Details regarding sample size, prevalence and SNP heritability for each cohort (and sex-specific samples) are provided in **Supplementary Table 1**.

The meta-analysis included in total 593,724 cases and 1,771,286 controls and was performed using a fixed-effect model in METAL^12^, weighting SNP effects by sample size (**Methods**). In addition, we performed sex-specific meta-analyses to evaluate whether results may differ between males and females.

The GWAS on the UKB cohort identified 14 independent risk loci, the SNP heritability (*h*^*2*^_*SNP*_) was 8.23% (SE=0.36%), with a *λ*_*1000*_=1.00, an intercept of 1.02 (SE=0.0086) and a ratio (LD score intercept-1 divided by mean *χ*^*2*^-1) of 0.043 estimated with LD score regression (LDSC)^13^ (**Methods**). The 23andMe cohort GWAS identified 477 independent risk loci, with an *h*^*2*^_*SNP*_ of 8.15% (SE=0.0021), a *λ*_*1000*_=1.00, an intercept of 1.15 (SE=0.016) and a ratio of 0.084 (SE=0.009). *h*^*2*^ estimates agree well between the two cohorts, and both indicate as much as 95.7% and 91.6% of the observed inflation could be ascribed to true polygenicity and large sample size. The genetic correlation between the cohorts was 0.66 (SE=0.0179, *p*=2.7×10^−292^). This is relatively low, as was also noted in the previous GWAS for insomnia^6^. This between-cohort discrepancy may be due to the lower accuracy of the 23andMe insomnia phenotype as compared the UKB insomnia phenotype, which was also shown when both their measures were benchmarked to the same independent cohort (using the Netherlands Sleep Registry (N=1,918): where we previously showed sensitivity 98% and 84%, and specificity 96% and 80%, for the items used in UKB and 23andMe, respectively^6,11^). These results suggest that the items used in both cohorts are good predictors of insomnia, although what they ascertain is slightly different phenotypically. In addition, insomnia is a heterogeneous trait and might consist of multiple subdomains of symptoms. In the meta-analysis, associations that are similar across the two cohorts will be amplified, while dissimilar results are less likely to show significant association, supported by both cohorts, in the meta-analysis. There, the relatively low genetic correlation is expected to decrease our statistical power, yet the large sample size partly counterbalances this.

For the meta-analysis, LDSC^13^ estimated *h*^*2*^_*SNP*_ of 7.2% (SE=0.17%), a *λ*_*1000*_ of 1.00, an intercept of 1.15 (SE=0.0158) and a ratio of 0.079 (SE=0.08). The latter indicates that at most 92.1% of the observed inflation is due to high polygenicity of insomnia. We estimated that current GWS SNPs explain 17.3% of the total *h*^*2*^_*SNP*_ and over 57 million subjects will be required to detect SNPs at genome-wide significance (*p*<5e-8) that explain at least 90% of the SNP heritability (**Supplementary Note 1**). Insomnia is currently estimated to be the 3rd most polygenic trait, following major depressive disorder (MDD) and educational attainment (EA)^8^ (**Supplementary Note 1**).

We found that polygenic risk scores (PRS) calculated for 10,000 individuals from the UKB sample (using an independent hold-out sample) explained 2.0% of the phenotypic variation at most (**Supplementary Note 2, Extended Data Fig. 1** and **Supplementary Table 2**). We observed a considerable decrease of the predictive power using a non-UKB independent cohort as target sample (the Million Veteran Program cohort MVP, N=183,944) in which the PRS explained 0.66% of the phenotypic variation at most (**Supplementary Note 2** and **Supplementary Table 2**). This discrepancy between the UKB hold-out sample and the MVP sample may be due to dissimilarity in the phenotype as measured in the MVP cohort compared to the combined UKB and 23andme cohorts, or to ancestral mismatches. Using LDSC^13^, we estimated the genetic correlation (*r*_*g*_) between the GWAS results for males and females to be 0.92 (*p*<1e-323), 0.85 (*p*=3.2e-64) and 0.91 (*p*<1e-323) in the meta-analysis, UKB and 23andMe cohort specific GWAS, respectively, consistent with the previous report on partly overlapping data^6^. Sex-specific results are available in the supplementary materials, we will here focus on results obtained in from the sex-combined meta-analysis.

**Figure 1.**
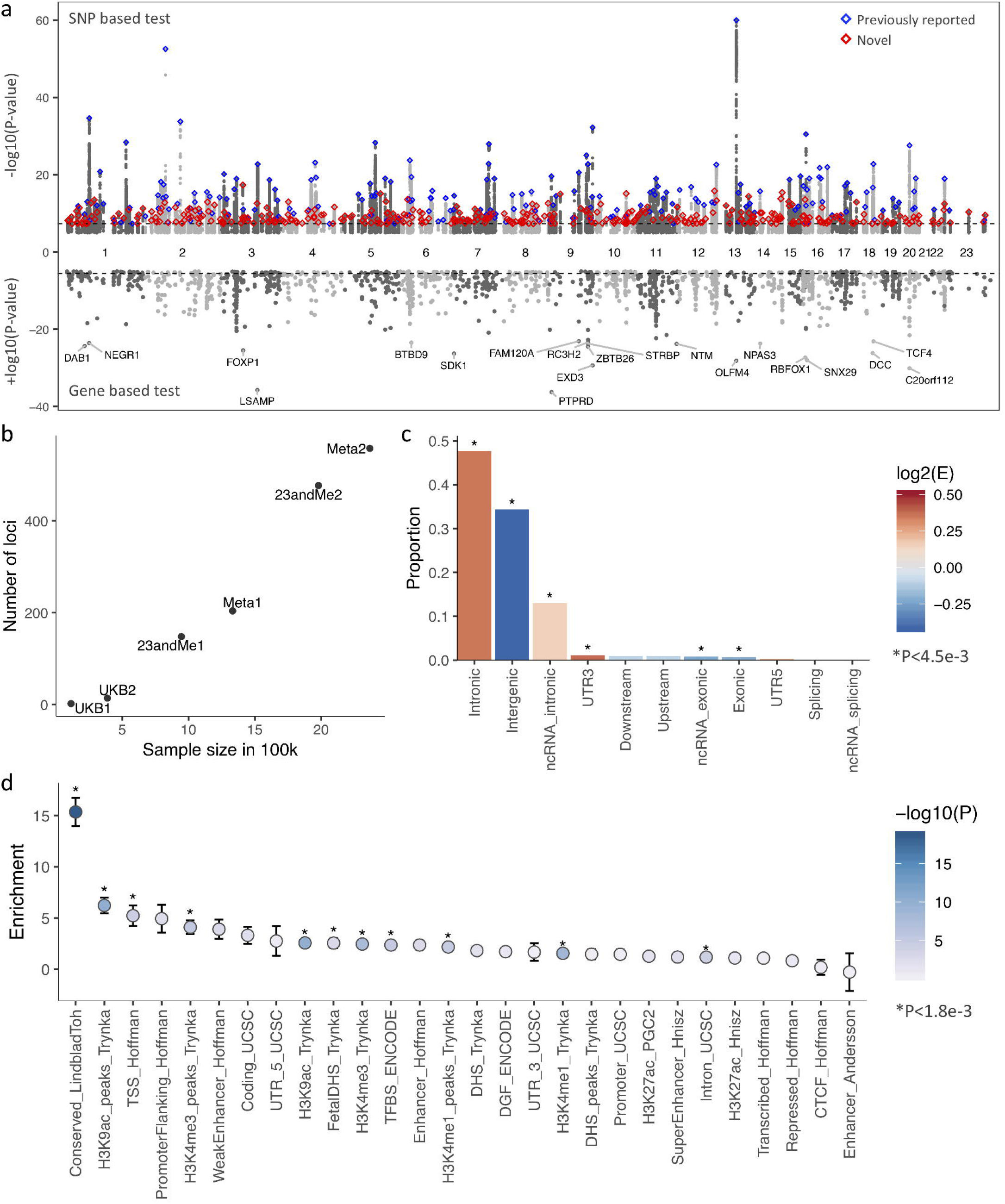
GWAS meta-analysis of insomnia in 2,365,010 individuals. (**a**) Manhattan plot for SNPs (top) and genes (bottom, based on MAGMA gene analysis). The top SNPs of previously identified loci are labeled in blue, novel loci are in red. (**b**) Number of risk loci identified by insomnia GWAS with different sample sizes. (**c**) Proportion of genome-wide significant (GWS) SNPs in each functional category. Bars are colored by log2 transformed enrichment value (the proportion of GWS SNPs in a category divided by the proportion of all analyzed SNPs in the same category). Stars represent significant enrichment or depletion compared to all analyzed SNPs based on Fisher’s exact test (two-sided). (**d**) Enrichment of SNP heritability in 28 annotation categories computed by LD score regression. Error bars indicate standard errors, and data points are colored by -log10 P-value. Stars represent significantly enriched annotations.

### SNP and gene-based findings from the meta-analysis

The meta-analysis yielded 51,876 genome-wide significant SNPs, residing in 554 distinct loci containing 791 independent lead SNPs (*r*^*2*^<0.1; **Methods, Extended Data Fig. 2-3, Supplementary Table 3-4** and **Supplementary Note 3**). Of the top 538 SNPs (i.e. SNPs with the lowest P-value in each locus), 97.1% showed concordance in direction of effect between the two cohorts. Out of 554 loci, 11 loci were genome-wide significant in the UKB, and 419 also in the 23andMe cohort-specific GWASs, while 9 loci were identified in both cohorts (**Supplementary Table 4**). The total summed length of risk loci was 145.2Mb, which is 4.9% of the genomic regions containing known SNPs in the entire genome. Of the 554 loci, 190 loci overlapped with previously identified risk loci^5–7,11^ and 364 were novel (**Methods, Fig. 1a** and **Supplementary Table 4**). Of the loci previously reported in Hammerschlag *et al*.^11^, Lane *et al*. (2017)^5^, Lane *et al*. (2019)^7^, and Jansen *et al*.^6^, 1/2, 3/5, 25/57 and 18/202 loci were no longer significant in the current meta-analyses, respectively (**Supplementary Table 4, Supplementary Note 4** and **Extended Data Fig. 4**). We show that the number of risk loci increases almost linearly as a function of sample size (**Fig. 1b**), and that both newly identified risk loci and un-replicated risk loci from the previous GWASs showed significantly higher P-values compared to other risk loci, as expected (**Supplementary Note 5** and **Extended Data Fig. 5**).

**Figure 2.**
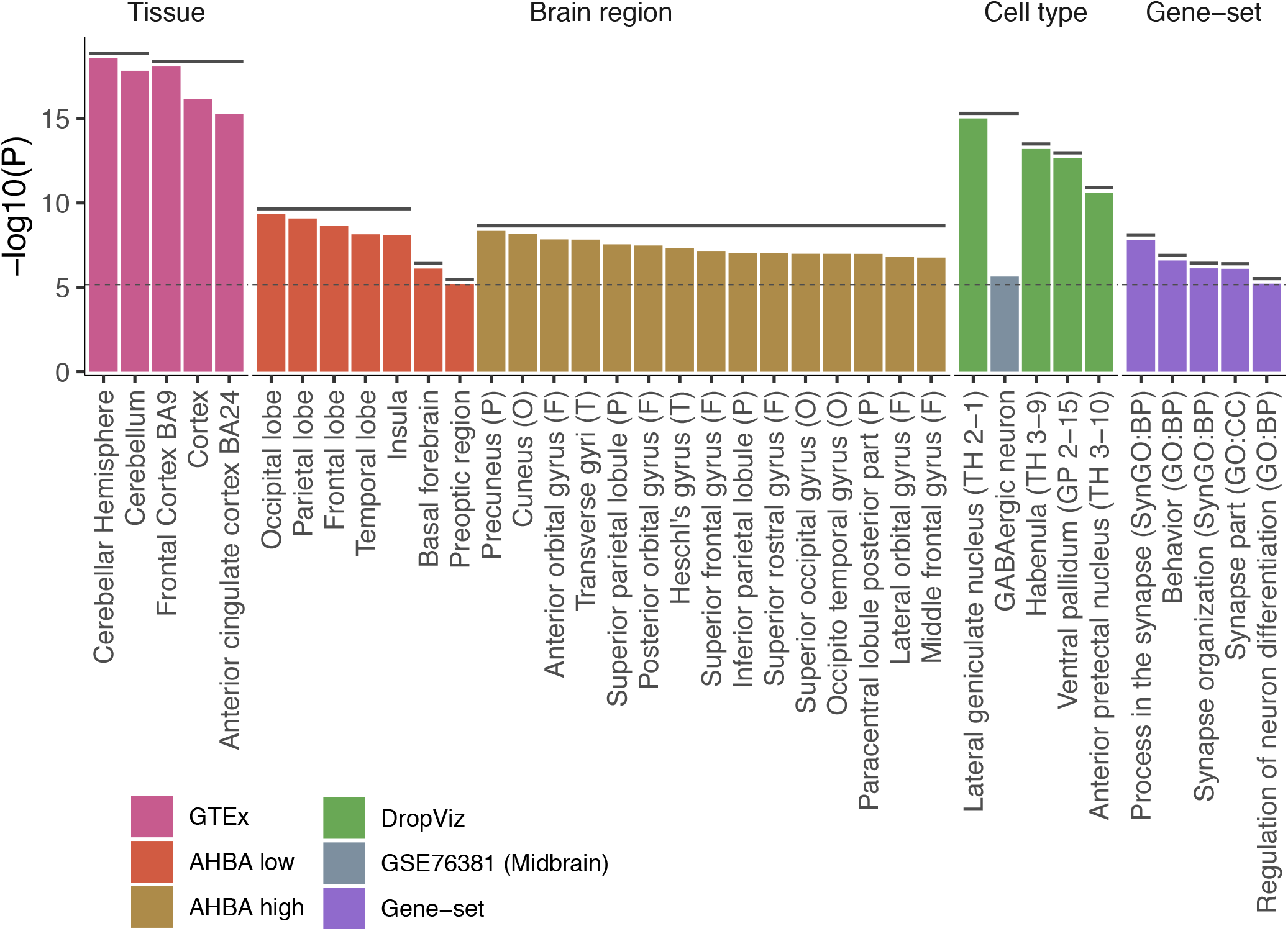
Tissues, brain regions, cell types and gene-sets associated with insomnia based on MAGMA analyses. P-values of significantly associated tissues, brain regions, cell types and gene-sets. Confounders after pairwise conditional analyses per dataset are not displayed (full results are available in **Supplementary Tables 13-21**). Horizontal solid lines above bars represent clusters of items which are either collinear or jointly associated with insomnia. Bars are colored by datasets. A letter in the parenthesis for brain regions from the AHBA high dataset refers to parent brain regions from the AHBA low dataset (F: frontal lobe, O: occipital lobe, P: parietal lobe and T: temporal lobe). Labels in parenthesis for cell types from DropViz dataset refer to the cluster label from the original study. BP: biological process, CC: cellular components. The dashed line indicates the Bonferroni corrected threshold for statistical significance (*p*=0.05/5974).

**Figure 3.**
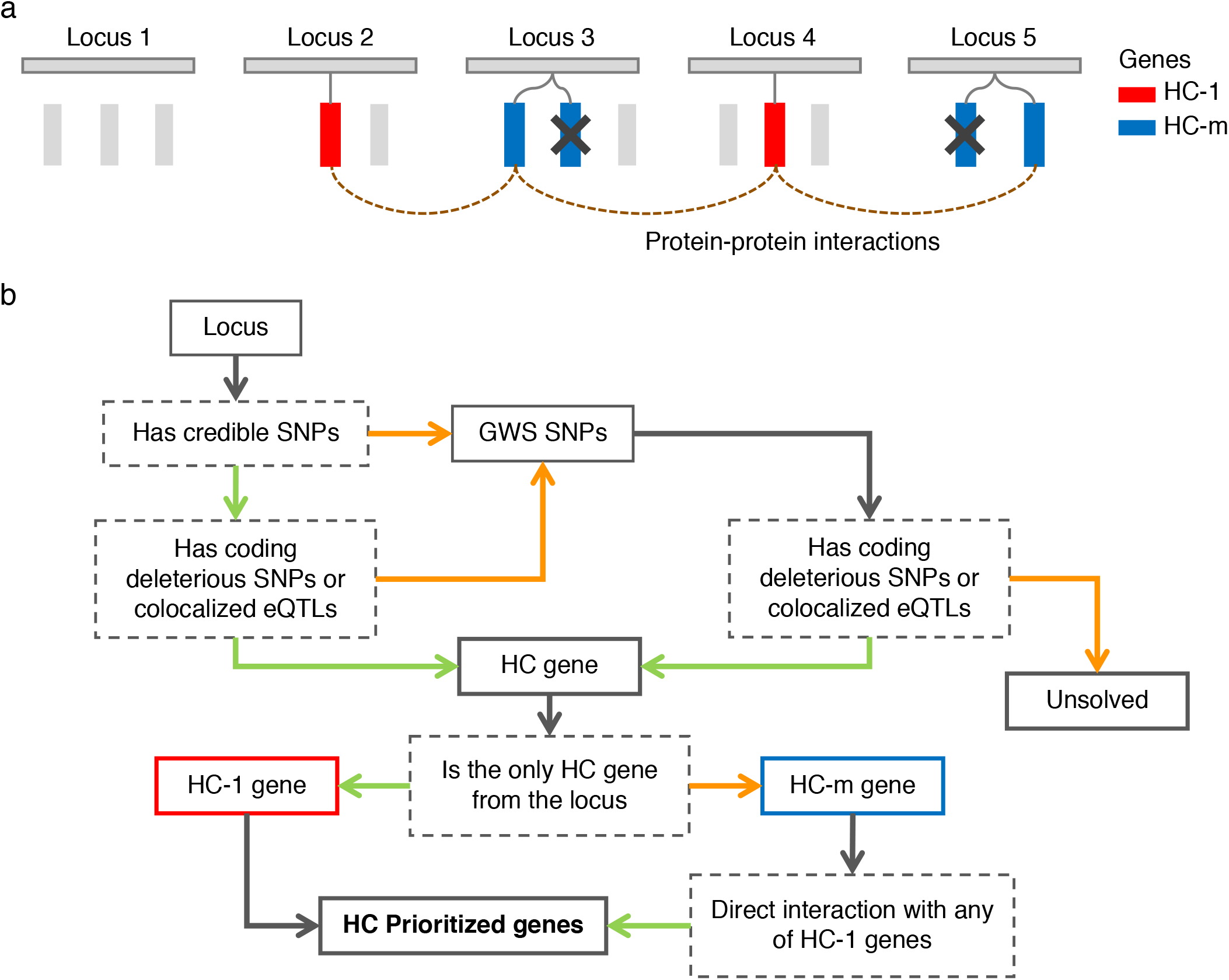
Schematic overview of gene prioritization strategy from risk loci. (**a**) a conceptual illustration of gene-prioritization based on multi-loci information. Rectangles represent genes colored by prioritization status, HC-1: high confidence, single gene from a locus; HC-m: high confidence in locus with multiple HC-m genes. Genes colored grey are located within or close to insomnia risk loci but were not implicated either due to a lack of functional evidence or because other genes in the same locus had higher priority. Genes that do not have a direct protein-protein interaction to HC-1 genes are filtered out (indicated by a cross). (**b**) a flowchart of gene prioritization using credible SNPs and genome-wide significant (GWS) SNPs. Solid boxes represent inputs or outputs, dashed grey boxes represent conditions, green arrows represent paths where an originated condition is positive, orange arrows represent paths when an originated condition is negative, grey arrows represent paths which do not have any other option.

**Figure 4.**
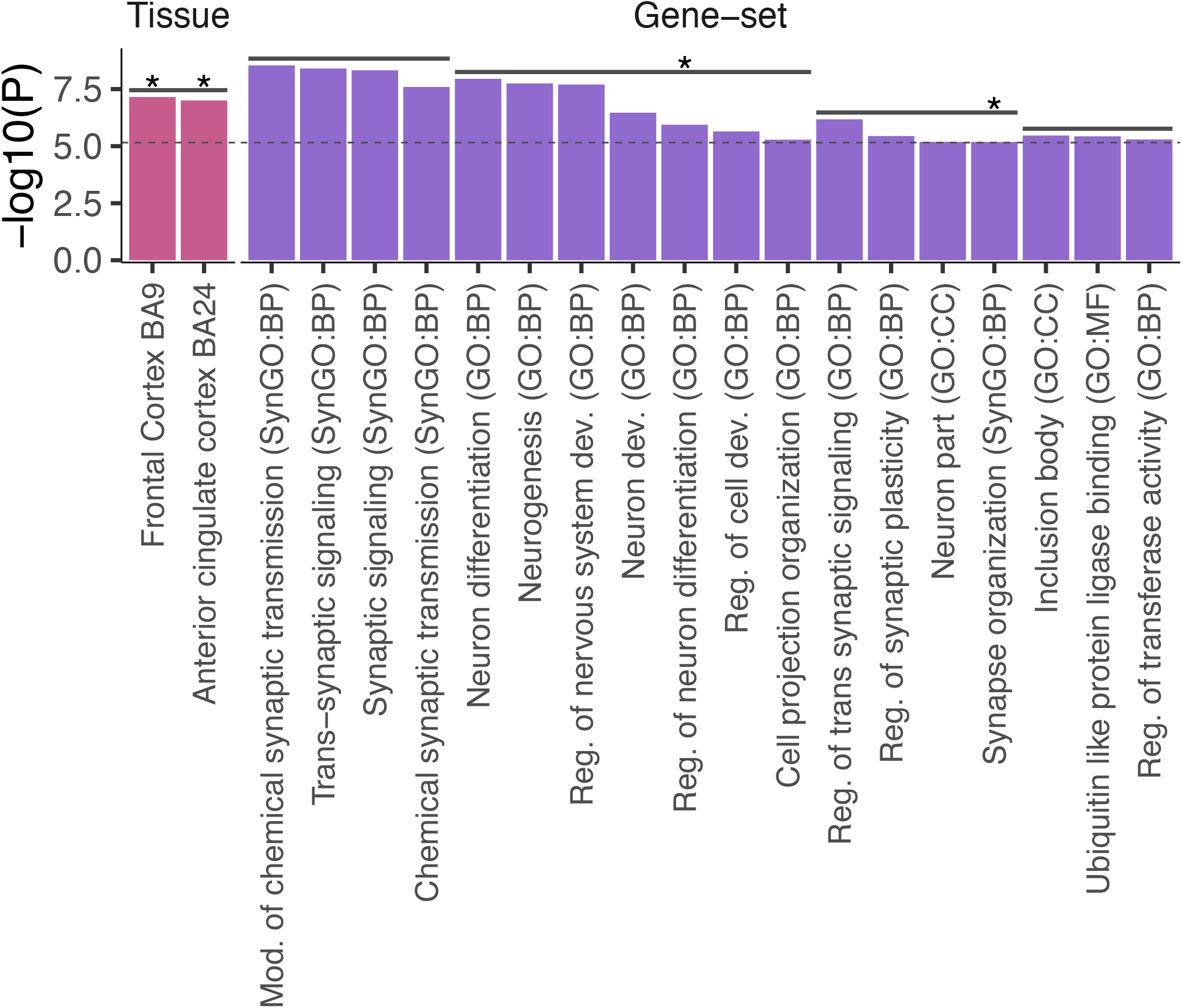
Tissues and gene-sets associated with the high-confidence prioritized genes. P-values of significantly associated tissues, brain regions, cell types and gene-sets. Confounders after pairwise conditional analyses per dataset are not displayed. Horizontal solid lines above bars represent cluster of items which are either collinear or jointly associated with insomnia. Stars represent items that also showed significant association with insomnia by using the full GWAS results. Bars are colored by datasets. BP: biological process, CC: cellular components, MF: molecular function.

The 51,876 genome-wide significant SNPs were most abundant in intronic regions (47.7%) and significantly enriched compared to all analyzed SNPs (enrichment (E)=1.28; *p*<1e-323, two-sided Fisher’s exact test; **Methods**). Intergenic regions (34.4%) were the second most common, although these showed a significant depletion compared to all analyzed SNPs (E=0.75, *p*<1e-323; **Fig. 1c** and **Supplementary Table 5**). GWS SNPs in exonic regions (0.73%) showed a significant depletion (E=0.86, *p*=3.6e-3) compared to all analyzed SNPs, and included 168 nonsynonymous, 1 stop gain, 192 synonymous and 1 non-frameshift substitution. SNPs in 3’ untranslated regions (UTR; 1.1%), representing significant enrichment (E=1.30, *p*=6.9e-10), while proportions of SNPs in other flanking regions did not differ from all analyzed SNPs (**Fig. 1c** and **Supplementary Table 5**). Stratified heritability analyses^14^ with 28 annotations (**Methods**), showed that SNP heritability was most strongly enriched in conserved regions, followed by multiple chromatin modification markers, consistent with the previous meta-analysis^6^ (**Fig. 1d** and **Supplementary Table 6**). Next, we performed a gene-based test using MAGMA gene analysis by assigning SNPs to genes with 2 kb upstream and 1 kb downstream windows. Of 19,751 protein-coding genes analyzed, 1,429 genes reached genome-wide significance (0.05/19751=2.53e-6; **Fig. 1a** and **Supplementary Table 7**). The most significant gene was *PTPRD* (*p*=7.2e-37) which has been associated previously with insomnia ^6^, restless leg syndrome^15^, type 2 diabetes^16^, and coronary artery disease^17^. The second-most significant gene was *LSAMP* (*p*=2.8e-36) which was not significant in the previous insomnia GWAS but has been associated with major depressive disorder (MDD)^18^ and suicidal behavior^19^, which are highly genetically correlated to insomnia. The most significantly associated genes from the previous insomnia GWASs, *MEIS1*^11^ and *BTBD9*^6^, were also supported in the current study (*p*=1.2e-14 and 4.8e-24, respectively). Risk loci and gene analyses for sex-specific meta-analyses are summarized in **Supplementary Note 6, Supplementary Table 7-11** and **Extended Data Fig. 6**.

The 51,876 GWS SNPs were mapped to 3,526 genes (of which 1,455 are located within the risk loci) using positional, eQTL and chromatin interaction mapping strategies^20^ (**Methods** and **Supplementary Table 12**). Together with genes significantly associated with insomnia based on formal gene-based analysis in MAGMA, 3,898 unique genes were implicated. We observed significant genetic correlations with 350 traits out of 551 traits tested, including multiple cardiovascular, metabolic, and psychiatric traits, in agreement with previous reports^6^ (**Methods, Supplementary Note 7, Supplementary Table 13** and **Extended Data Fig. 7**). Of 554 insomnia risk loci, 282 loci were colocalized with one of the 350 traits indicating shared causal variants between the traits (**Supplementary Note 7**). In addition, a clustering of these 282 loci based on the colocalization pattern across 350 traits suggested presence of locus heterogeneity where we observed distinct clusters of loci: one was mainly colocalized with metabolic traits and the other was mainly colocalized with psychiatric traits (**Supplementary Note 7, Supplementary Table 14-18** and **Extended Data Fig. 8**). We performed extensive post-GWAS analyses to test for convergence of genetic association signals in tissue types, brain regions, cell types and biological pathways associated with insomnia, based on the total genome-wide distribution of genetic associations, weighted by statistical significance using MAGMA gene-set analyses^21^ (**Methods** and **Supplementary Note 8**). Based on the full GWAS distribution, we found evidence for enrichment in brain tissue (specifically the cerebral cortex), neurons in four brain specific areas (lateral geniculate nucleus, habenula, ventral pallidum, and anterior pretectal nucleus), GABAergic neuron and biological pathways involved in synaptic functions, behavior and neuron differentiation (**Fig. 2, Supplementary Note 8, Extended Data Fig. 9 and Supplementary Tables 19-30**).

### Prioritization of high confidence genes from 554 risk loci using multi-locus information

Due to LD, many non-causal SNPs will show a statistical association with a trait simply because they are correlated with causal SNPs due to LD. We call these non-causal significant SNPs ‘LD by-products’. Since we do not know which SNPs are the causals ones and which are the LD by-products, post-GWAS annotation will provide information on many SNPs that are likely not causally related to the trait, and therefore conventional post-GWAS analysis testing for convergence may still contain considerable noise from these LD by-products. Multiple studies have already proposed to conduct fine-mapping per locus and prioritize credible SNPs and genes from each locus before testing for convergence^22–25^. Here we propose to prioritize genes additionally based on cross-locus connections.

We assume the following: (i) Credible SNPs can be indicated using *in silico* fine-mapping strategies; (ii) SNPs that have a structural or regulatory effect on a gene product are more likely to be causal than SNPs that have no such (known) effect; (iii) genes that are implicated as the only gene in a locus are likely to be the gene responsible for the statistical result at the locus, and are therefore likely a true causal gene, and (iv) if insomnia is influenced by hundreds of genes, and at least part of those are functionally related (**Fig. 3**).

We first defined credible SNPs by performing statistical fine-mapping for each of the 554 insomnia risk loci, using FINEMAP^23^ (**Methods**). For each locus, the 95% credible sets were extracted, resulting in a total of 26,016 unique SNPs (see **Supplementary Note 9** and **Extended Data Fig. 10** for detailed results). More than 94.5% of the SNPs in 95% credible sets had a posterior inclusion probability (PIP) ≤0.1, suggesting that those SNPs were ‘unsolved’ by FINEMAP, which can be due to high LD and small effect sizes. We retained only the 1,423 credible SNPs with PIP>0.1 distributed over 429 loci. Association statistics and functional annotation of these credible SNPs are provided in **Supplementary Table 31**. Credible SNPs were then mapped to genes if they were deleterious coding SNPs (non-synonymous, stop-gain, stop-loss or splicing SNPs) (**Supplementary Table 32**) or if they were eQTLs from GTEx v8^26^, PsychENCODE^27^ and eQTLGen^28^ resources that were colocalized with insomnia GWAS summary statistics (**Methods** and **Supplementary Table 33**). This resulted in labeling 314 genes from 178 loci as having a credible SNP that was either nonsynonymous coding or co-localized with eQTLs (**Supplementary Table 34**). For 376 of the remaining insomnia loci, no genes were present with the above criteria. This was either due to a lack of credible SNPs or a lack of credible SNPs that were deleterious coding or were colocalized eQTLs. For these loci, we used GWS SNPs instead of credible SNPs and again evaluated whether they were deleterious coding SNPs or colocalized with eQTLs. This resulted in an additional 257 genes from 103 loci (**Supplementary Table 34**). Together, these 571 genes from 281 loci were called high confidence (HC) genes. A single locus could contain multiple HC genes, but some were mapped to a single gene. In the case of a single HC gene, that gene was considered the only likely causal candidate from the locus. We labeled these single HC genes as HC-1 (with the highest confidence) and the remaining genes as HC-m (i.e., HC genes but with multiple genes in the same locus and therefore unclear whether they are all causal, whether only one is causal, or whether a subset is causal; **Fig. 3** and **Methods**). We obtained 164 HC-1 genes from 166 loci (2 genes were mapped from two loci due to distal eQTLs) and 407 HC-m genes from 116 loci (**Supplementary Table 34**).

Assuming that the HC-1 genes are the most likely causal genes, and that functional relationships exist among the set of causal genes, we then used the set of HC-1 genes to select the most likely causal genes from the set of HC-m genes. To assess functional relations, we used protein-protein interactions (PPI) using the InWeb database^29^, and identified HC-m genes that have a direct interaction with HC-1 genes (**Methods**). We choose to use PPI for this, and not for example co-expression, because we aimed to identify genes whose products are known to form a protein complex, increasing the likelihood that they have a common function. Alternate measures of connections between genes (such as spatial or temporal co-expression) can of course also be used in this step. Of 407 HC-m genes, 125 genes from 74 loci were found to have a direct PPI with an HC-1 gene. We then defined 289 genes (164 HC-1 and 125 HC-m genes connected to HC-1) from 239 loci as the ‘high confidence prioritized’ (HCP) genes (**Supplementary Table 34**). The HCP genes are the genes we believe to be most likely functionally associated to insomnia based on the associated loci. Out of 239 loci, 202 were linked to single HCP genes and the maximum number of genes from a single locus was 5. We consider these 239 loci to be ‘resolved’ and the remaining 315 loci to be unsolved due to lack of biological evidence or information at this moment. Although we still cannot identify genes from 56.7% of the insomnia loci, we assume that these loci are randomly distributed, and thus we expect the HCP genes to explain some of the underlying biological mechanisms of insomnia as the HCP genes are more refined and likely to contain fewer false positives compared to 3,526 genes mapped by all GWS SNPs.

The HCP genes include *NEGR1* which has been reported to be associated with multiple traits such as body mass index^30^, MDD^31^, cognitive traits^32,33^ as well as in the previous insomnia GWAS^6^ and its effect on neuronal growth and behavior has been suggested^34,35^. The HCP genes also include *EP300* known to be involved in circadian rhythm^36^ which regulates sleep timing (see **Supplementary Table 34** for the full list of the HCP genes).

To look for convergence in biological functions of the HCP genes, we assessed enrichment in tissues, brain regions, cell types and biological pathways (**Methods**).

The HCP genes showed significant joint associations with frontal cortex BA9 and anterior cingulate cortex BA24 (**Fig. 3** and **Supplementary Table 35-36**). These tissue enrichments seen with HCP genes were also detected when using the full GWAS results (**Supplementary Table 37-39** and **Supplementary Note 8**).

On the other hand, some tissue enrichments that were significant when using the full GWAS, are no longer detected when using the HCP gene list. This is because genes driving these associations in the full GWAS were not prioritized, which suggests that these initial enrichments were less likely to be based on true causal genes. (**Supplementary Note 10 and Extended Data Fig. 11**).

From the gene-set enrichment analyses, the HCP genes showed significant enrichment in 18 gene-sets clustered into four independent groups (**Fig. 4, Extended Data Fig. 12** and **Supplementary Table 40**). The most significantly enriched gene-sets from each cluster were ‘modulation of chemical synaptic transmission’ (SynGO:BP), ‘neuron differentiation’ (GO:BP), ‘regulation of trans synaptic signaling’ (GO:BP) and ‘inclusion body’ (GO:CC). We identified not completely overlapping and more gene-sets when using the prioritized genes than when using the full GWAS results. This may indicate that, by filtering LD by-products genes to reduce noise in associated genes, we were able to identify gene-sets specific to the most likely causal genes prioritized from insomnia associated loci, which are missed by using full GWAS results.

In this study, we performed a GWAS meta-analysis for insomnia including over 2.3 million subjects. We identified 554 insomnia risk loci, almost doubling the number identified by the previous largest study with 1.3 million subjects^6^. We demonstrated a novel strategy using known biological functions of SNPs and multi-locus functional relations of genes to prioritize the most likely causal genes, and based post-GWAS analyses for convergence on these genes. Applying this strategy, we identified 289 HCP genes from 239 loci and compared associated tissue and cell types, as well as gene-sets based both on the set of prioritized genes and all genes implicated in the GWAS. We found that the former is less likely to contain LD by-products, as it provided more specific results, pointing towards specific synaptic functions in specific brain regions. Of course, the ultimate test of whether the HCP genes are actually causally involved still lies in functional follow-up experiments.

There are several limitations of the current strategy. First, the proposed strategy using multi-locus information is only feasible for polygenic traits with a reasonable number of independent risk loci identified by GWAS and a reasonable number of loci with single implicated genes. Second, the prioritization procedure depends on the availability and accuracy of functional annotations of SNPs and genes. For example, we defined an HC-1 gene as the only gene from a single locus with high confidence biological evidence.

However, in the future, more SNPs may be known to be deleterious (by increasing accessible (rare) SNPs in GWAS) or colocalized eQTLs (by increasing statistical power to detect eQTLs and availability of them in specific cell types), which may change the current results and may allow us to identify additional high confidence genes from loci. Third, cross-locus linking of genes depends on the availability and reliability of biological information (PPI, co-expression networks, or any other gene-correlation matrix deemed relevant), which is currently not abundantly available and still imperfect. We do believe using cross-locus information greatly aids in making sense of the multitude of associated genes, and the current study shows that this strategy implies a role for more specific biological functions in insomnia. In conclusion, the current study provides further insights into the possible genetic pathways contributing to insomnia using a novel strategy in prioritizing genes and generating hypotheses that can be tested in functional experiments.

## Methods

### Sample cohorts

#### UK Biobank

The UK Biobank (UKB) study is designed to curate genotype and phenotype information for ∼500,000 adult participants in the UK^37^. All participants provided written informed consent (the UKB received ethical approval from the National Research Ethics Service Committee North West-Haydock), and all study procedures were in accordance with the World Medical Association for medical research. The current study was carried out using the UK Biobank resource under application number 16406. We used genotype data released in May 2018 by UK Biobank (UKB) in which genotyping was completed in 489,212 individuals, and 488,377 individuals passed genotype quality controls performed centrally by UKB. These genotypes were imputed based on the Haplotype Reference Consortium (HRC) reference panel^38^ and a combined reference panel of UK10K^39^ and 1000 Genome projects Phase 3(1000G)^40^ panels. The imputed genotypes were available for 487,422 individuals. Imputed variants with INFO score >0.9 were converted to hard-calls at a certainty threshold of >0.9. Although UKB provides self-reported ancestry for each participant, we determined European ancestry by projecting 1000G genetic principal components on the UKB genotypes and assigned ancestry based on the closest Mahalanobis distance from the 1000G population average (see ref^6^ for more details). This resulted in 460,527 individuals assigned to the European population, of whom 387,614 unrelated individuals were included in the current analyses.

#### 23andMe

23andMe Inc. is a personal genetics company. Customers of 23andMe have the option to consent to participate in research and answer survey questions on-line about a wide variety of phenotypes. We obtained insomnia GWAS summary statistics from 23andMe Inc. research participants who are of European ancestry. All participants included in the study provided informed consent, and the research study and data collection procedures were approved by an AAHRPP-accredited private institutional review board, Ethical and Independent Review Service.

DNA was extracted from saliva samples and each sample was genotyped by either by Illumina HumanHap550+ BeadChip (∼560k SNPs), Illumina OmniExpress+ BeadChip (∼950 SNPs), Infinium Global Screening Array (∼640 SNPs) or fully customized array containing ∼560k SNPs. Samples that failed to reach 98.5% call rate were re-analyzed. Individuals whose analyses failed repeatedly were re-contacted by 23andMe to provide additional samples. Genotypes were phased by Finch (23andMe’s inhouse tool modified from Beagle^41^) or Eagle2^42^ (for samples genotyped by the custom array). Subsequently, genotypes were imputed against combined panel of 1000G^40^ and UK10K^39^ merged by Minimac3^43^. Ancestry of individuals was determined by Ancestry Composition^44^. The reference population data was derived from public datasets (the Human Genome Diversity Project^45^, HapMap^46^ and 1000G^40^) and 23andMe customers who have reported having four grandparents from the same country.

A maximal set of unrelated European individuals was used for each association analyses, resulted in 1,038,003, 1,200,179 and 1,978,022 for male-only, female-only and sex-combined GWASs, respectively.

### Phenotype assessment

#### UK Biobank

Participants were asked the question ‘Do you have trouble falling asleep at night or do you wake up in the middle of the night?’ (Data-Field 1200) and answers were selected from the following 4 options; ‘never/rarely’, ‘sometimes’, ‘usually’ and ‘prefer not to answer’ on a touchscreen. If the participant opened the ‘help’, the message ‘If this varies a lot, answer this question in relation to the last 4 weeks.’ was displayed. There were 3 assessments available for insomnia (f.1200.0.0 - f.1200.0.2). For subjects with a missing value in the first assessment (f.1200.0.0), the second assessment (f.1200.0.1) was used if that contained a non-missing value. Subsequently, for subjects with NA in both first and second assessments, the third assessment (f.1200.0.2) was used, again only if there was a non-missing value. Note that for subjects with non-missing values in multiple assessments, only the first assessment was used. We excluded individuals who answered ‘prefer not to answer’ resulting in a total of 386,988 subjects. We then dichotomized the phenotype where subjects who answered ‘usually’ were considered as cases, otherwise they were considered controls, following previous strategy^6^.

#### 23andMe

Participants were asked to answer one or more questions related to seven sleep-related traits. Participants with positive response to any of the following questions were considered as cases: 1) ‘Have you ever been diagnosed with, or treated for, insomnia?’, 2) ‘Were you diagnosed with insomnia?’, 3) ‘Have you ever been diagnosed by a doctor with any of the following neurological conditions?’ (Sleep disturbance), 4) ‘Do you routinely have trouble getting to sleep at night?’, 5) ‘What sleep disorders have you been diagnosed with?

Please select all that apply.’ (Insomnia, trouble falling or staying asleep), 6) ‘Have you ever taken these medications?’ (Prescription sleep aids) and 7) ‘In the last 2 years, have you taken any of these medications?’ (Prescription sleep aids). Participants who did not provide either positive or uncertain answer (‘I don’t know’ or ‘I am not sure’) in any of the 7 questions nor the followings are considered as controls: 1) ‘Have you ever been diagnosed with, or treated for, any of the following conditions?’ (Insomnia; Narcolepsy; Sleep apnea; Restless leg syndrome), 2) ‘In the past 12 months, have you been newly diagnosed with any of the following conditions by a medical professional?’ (Insomnia; Sleep apnea; Migraines), 3) ‘Have you ever been diagnosed with or treated for any of the following conditions?’ (Post-traumatic stress disorder; Autism; Asperger’s; Sleep disorder), 4) ‘Have you ever been diagnosed with or treated for a sleep disorder?’, and 5) ‘Have you ever been diagnosed with or treated for any of the following conditions?’ (A sleep disorder). A detailed flowchart of the case/control decisions is provided in the previous study^6^.

### Genome-wide association analysis

#### UK Biobank

We performed genome-wide association analysis (GWAS) for insomnia with PLINK 2.0^47^, using a logistic regression model with age, sex, genotype array and the first ten ancestry genetic principal components (computed based on unrelated European subjects defined above, based on 145,432 independent SNPs (*r*^*2*^<0.1, MAF>0.01, INFO=1) using FlashPCA^48^) as covariates. We analyzed autosomal, pseudo-autosomal and X chromosomes. For X chromosome, we used a model where genotype was coded [0, 2] for males (with a PLINK flag --xchr-model 2) for consistency with the 23andMe cohort. The genotype missingness was set to <0.05 and SNPs were limited to those with a minor allele count (MAC) >100. For sex-specific GWASs, we followed the same criteria except sex was excluded from the covariates. The number of analyzed SNPs and sample sizes are summarized in **Supplementary Table 1**.

#### 23andMe

Summary statistics were obtained from 23andMe Inc. where association tests were performed using logistic regression with age, sex, genotype array, and the first 5 genetic ancestry principal components. We first extracted SNPs which passed quality control by 23andMe. When there were both genotyped and imputed genotypes available for a single SNP, the imputed SNP was kept. We then further extracted SNPs with MAC (MAF x sample size x 2) >100. The analyses were performed for unrelated European subjects for sex-combined (N=1,978,022), male-only (N=1,038,003) and female-only (N=1,200,179) separately. Note that related subjects were excluded for each analysis separately, therefore, the sum of subjects in male- and female-only analyses is not equal to the subjects in sex-combined analysis. The number of available SNPs and sample sizes are summarized in **Supplementary Table 1**.

### GWAS meta-analysis

A meta-analysis on the GWAS summary statistics from UKB and 23andMe cohorts was performed using METAL software^12^, based on the fixed-effect model with SNP P-values weighted by sample size. The meta-analysis was performed for the sex-combined and each sex-specific GWASs separately. We used a unique marker ID consisting of chromosome, position, and alphabetically ordered alleles to match SNPs between cohorts. Since indels were coded as I/D in the 23andMe cohort, exact alleles were assigned for indels based on information from the UKB cohort. We assigned alleles to indels in the 23andMe only when there were bi-allelic indels on the same position in the UKB cohort. The number of analyzed SNPs and sample sizes are summarized in **Supplementary Table 1**.

We converted Z-statistics to standardized effect sizes (and their standard error) as a function of MAF and sample size as below^49^.

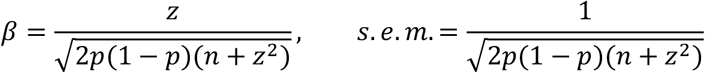

where *p* is MAF, *n* is sample size for a given SNP.

### SNP heritability and stratified heritability estimate

SNP heritability was estimated for sex-combined and sex-specific GWASs on UKB, 23andMe, and meta-analyses, using LD score regression (LDSC)^13^. Pre-computed LD scores for 1000 Genome Phase 1 European subjects were downloaded from https://data.broadinstitute.org/alkesgroup/LDSCORE/. The analyses were limited to

HapMap3 SNPs and the MHC region (chr6:26Mb-34Mb) was excluded. Additionally, SNPs with chi-square statistics >80 were excluded. To compute SNP heritability on the liability scale, we provided a population prevalence of 30%^1^. Since the sample size of this study is large, *λ*_*GC*_ was scaled for 1000 cases and 1000 controls as *λ*_*1000*_ = 1 + (*λ*_*GC*_ - 1) x (1/Ncases + 1/Ncontrols) x 500.

To test whether SNP heritability was enriched in a specific category of functional annotations, we partitioned SNP heritability for binary SNP annotations^14^. The enrichment was computed as the proportion of SNP heritability explained by the SNPs with the annotations divided by the proportion of SNPs with the annotations. We obtained 28 functional annotations from https://data.broadinstitute.org/alkesgroup/LDSCORE/.

### Polygenic risk scoring

To evaluate phenotypic variance explained by our insomnia meta-analysis, polygenic score (PGS) was computed based on the SNP effect sizes using PRSice v2.2.1^50^ (http://www.prsice.info/) with default parameters. PRSice performs clumping of SNPS at *r*^*2*^=0.1. We set P-value thresholds of input SNPs at p<1.0, 0.5, 0.05, 0.01, 5-e3, 1e-3 and 1e-5. We computed PGS for three randomly selected sets of 10,000 UKB subjects, and summary statistics were re-calculated excluding those 10,000 subjects each time for the UKB cohort. We then meta-analyzed with the 23andMe GWAS which we used as training data. To compare the difference in the size of the target subjects, we also computed PGS for 3,000 subjects, 3 times by the same procedure. Chromosome X was excluded from the analyses. To evaluate the predictive power of the insomnia GWAS meta-analysis, we computed PGS for an independent cohort, the Million Veteran Program (MVP). The MVP is a research program funded by the US Department of Veterans Affairs to learn how genes, lifestyle, and military exposures affect health and illness^51^. In the MVP cohort, insomnia cases were defined as individuals answering “Yes” to any of the following three questions : “Trouble falling asleep when you first go to bed”, “Waking up during the night and not easily going back to sleep, or “Waking up in the morning earlier than planned or desired”. To be defined as a case, individuals also had to respond “Yes” to the following: “Feeling unsatisfied or not rested by your night’s sleep”. Controls were defined as not responding “Yes” to any of the first three questions and responding “No’ to the final question. This resulted in 45,355 cases and 138,589 controls. We then computed PGS using PRSice in the same way as described above, except the summary statistics of the full meta-analysis were used as training set this time.

### Definition of risk loci

Genomic risk loci were defined within FUMA (https://fuma.ctglab.nl) as described previously^20^, by first clumping SNPs with *p*<5e-8 at *r*^*2*^=0.6 using SNPs with *p*<1e-5 to define independent significant SNPs. Those independent significant SNPs were further clumped at *r*^*2*^=0.1 to define lead SNPs. The independent significant SNPs that were in LD with the same lead SNPs (*r*^*2*^>0.1) and LD blocks closer than 250kb were merged into a single locus. Each locus is represented by the most significant SNP (top SNP). A risk locus can contain multiple lead and independent significant SNPs. We manually excluded suspicious loci (e.g., a single SNP reaching genome-wide significance without any SNPs with *p*<1e-5 with *r*^*2*^>0.6) by examining LocusZoom plots (see **Supplementary Note 2** for details).

### Comparison with previously identified insomnia risk loci

Previously identified insomnia risk loci were obtained from 4 studies; 2 loci from Hammerschlag *et al*.^11^, 5 loci from Lane et al. (2017)^5^, 57 loci from Lane *et al*. (2019)^7^ and 202 loci from Jansen *et al*.^6^. We then overlapped previously identified loci with 554 loci identified from the sex-combined meta-analysis in this study. We considered risk loci which did not overlap with any of previously identified loci as novel findings.

### Comparison of insomnia GWASs with different sample sizes

Insomnia GWAS summary statistics for 6 different sample sizes were obtained as below. *UKB1* (N=113,006): The summary statistics were obtained from the study of Hammerschlag *et al*.^11^ (https://ctg.cncr.nl/software/summary_statistics).

*UKB2* (N=386,988): UKB only summary statistics from the current study. *23andMe1* (N=944,477): Only 23andMe cohort from the study of Jansen *et al*.^6^ (summary statistics is not publicly available, only the number of risk loci was obtained).

*Meta1* (N=1,331,010): The summary statistics of meta-analysis from the study of Jansen *et al*.^6^ (summary statistics is not publicly available, only lead SNPs were obtained).

*23andMe2* (N=1,978,022): 23andMe only summary statistics from the current study (not publicly available).

*Meta2* (N=2,365,010): The summary statistics of the main meta-analysis from the current study (not publicly available).

For each GWAS, risk loci were defined as described above. For 23andMe1 and Meta1, the number of risk loci were obtained from the previous study^6^ as the summary statistics are not available. For comparison of P-values of the same locus across GWASs with different sizes, we obtained lead SNPs for Meta1 from the previous study^6^ while 23andMe1 was excluded from the analysis.

To obtain matched SNPs across GWAS to compare effect sizes, we first extracted genome-wide significant SNPs for all risk loci from 5 GWASs (23andMe1 was excluded from the analysis). For Meta1, the analysis was limited to lead SNPs provided by the previous study^6^, therefore not all GWS SNPs in the risk loci were assessed. We then selected SNPs that reached genome-wide significance in at least 3 GWASs out of 5. From each locus, the single SNP with the minimum P-value was further selected (when there are multiple) for the analyses. Standardized effect sizes of those SNPs were computed based on summary statistics from each GWAS as described above. When Z-score were not available, P-values were converted to Z-score (two-sided).

### MAGMA gene, gene-property and gene-set analyses

Gene-base testing was performed using MAGMA v1.07^21^ to obtain gene P-values, using summary statistics of sex-combined meta-analysis. From 20,260 protein-coding genes, SNPs were assigned to one of the 19,751 genes within 2kb up- and 1kb downstream windows, based on the location obtained from Ensembl v92 GRCh37 using BioMart. We used the SNP-wise mean model and randomly selected 10,000 unrelated European subjects from the UKB cohort were used as a reference panel.

For tissue specificity analyses, we obtained RNA-seq data for 54 tissue types from GTEx v8^26^. Read per kilobase per million (RPKM) was log-2 transformed with pseudo count 1 followed by winsorization at 50, and average per tissue type was computed for each gene. In a gene-property analysis, the average across 54 tissue types was conditioned and the one-sided test was performed to identify positive associations of tissue specific gene expression with insomnia.

For gene expression of specific brain regions, we obtained normalized microarray data for 3,702 samples from 6 healthy donors^52^, from Allen Human Brain Atlas (AHBA; http://human.brain-map.org/static/download). From 58,692 probes, 31,098 with missing values in less than 20% of the samples were first extracted. When there were multiple probes per gene, genes with the highest variance across 3,702 samples were selected, resulting in 17,916 unique genes. Of those, 13,943 genes were mapped to unique Ensembl gene ID (v92

GRCh37). Every 3,702 samples were assigned to the structural ID of the brain where there were multiple layers of hierarchical structure^52^. We assigned the annotation of brain regions at 4th and 5th layers of the hierarchical structure where the top of the tree (‘brain’) was considered as layer 0. We limited consideration to brain regions with at least 5 samples, resulting in 54 and 106 brain regions for layer 4 and 5, respectively. For each brain region, the average expression values were computed for each gene. We then performed gene-property analysis conditioning on average across brain regions and a one-sided test was performed to identify positive associations of brain region specific gene expression with insomnia.

For cell type specificity analysis, we used 5 datasets from Linnarsson’s group that were used in the previous insomnia meta-analysis study: mouse samples from cortex and hippocampus (GSE60361, level 2 neurons)^53^, hypothalamus (GSE74672, level 2 neurons)^54^, oligodendrocytes (GSE75330)^55^, midbrain (GSE76381)^56^ and striatum (GSE97478)^57^. We additionally tested one of the most comprehensive scRNA-seq datasets for mouse brain, DropViz^58^ (http://dropviz.org/, 565 sub-clusters from 9 brain regions). In total, we obtain 728 cell types from 6 datasets. Each dataset was pre-processed as described in the previous study^59^. We performed the 3-step workflow (per dataset analysis, within-dataset and cross-dataset conditional analyses) as described previously^59^ to determine significant associations supported by multiple independent datasets..

Gene-sets for Gene Ontology (GO) terms and canonical pathways were obtained from MsigDB v6.2^60^ (http://software.broadinstitute.org/gsea/msigdb/index.jsp). We performed gene-set analyses for 5,033 gene-sets with at least 20 genes.

We evaluated all 5,974 tested items against the Bonferroni corrected threshold (0.05/5974=8.4e-6).

### Fine-mapping and credible SNPs

We performed fine-mapping of 554 risk loci using FINEMAP with the shotgun stochastic search algorithm^23^ (http://www.christianbenner.com/). For each risk locus, SNPs within 50kb from the top SNP (with the minimum P-value) or locus boundary, whichever larger, and with *p*<0.05 were used for fine-mapping. The pairwise LD matrix of SNPs was estimated based on randomly selected 100k unrelated European individuals from UKB using LDstore^61^ (http://www.christianbenner.com/). The maximum number of causal SNPs (*k*) was set to 10.

An exception was made for locus #4 which contained 7 SNPs, where *k* was set to 5. FINEMAP outputs a set of models (all possible combination of *k* causal SNPs in a locus) with posterior probability (PP) of being a causal model. A 95% credible set was defined by taking models from the highest PP until the cumulative sum of PP reached 0.95 for each locus. The 95% credible set SNPs were, therefore, defined by taking unique SNPs from 95% credible sets. In addition, a posterior inclusion probability (PIP) was calculated for each SNP as the sum of PPs of credible sets that contain that SNP. In this study, we only used credible SNPs with PIP>0.1, however, results of all credible set SNPs are described in **Supplementary Note 7** and **Extended Data Fig. 8**.

### Annotation of SNPs with FUMA

All GWS SNPs were annotated with their functions using FUMA. Functional consequences of SNPs on genes were obtained by performing ANNOVAR^62^ gene-based annotation using Ensembl genes. Enrichments of GWS SNPs in each annotation were tested by Fisher’s exact test (two-sided) by compared in with the annotations of all SNPs analyzed in the meta-analysis. To determine the deleteriousness of the SNPs, CADD v1.4^63^ score was annotated for each SNPs. In addition, RegulomeDB score (categorical score indicating how likely the SNPs are involved in regulatory elements)^64^ and 15-core chromatin state for 127 tissue/cell types obtained from Roadmap^65,66^ were annotated.

### eQTL colocalization

We colocalized insomnia summary statistics from the sex-combined meta-analysis within 554 risk loci with eQTL summary statistics using the coloc package in R^67^. We only tested genes whose significant eQTLs were overlapping with at least one of the GWS SNPs in the insomnia GWAS, and colocalization was performed for each gene for each of 51 eQTL datasets (i.e. 49 tissues from GTEx v8^26^, meta-analysis of blood samples from eQTLGen consortium^28^ and prefrontal cortex from PsychENCODE^27^). For each colocalization, we extracted SNPs available in both eQTL summary statistics of a testing gene and within 10kb of insomnia risk loci. We then performed *coloc*.*abf* function. We did not perform colocalization when there were <10 SNPs overlapping between insomnia and eQTL summary statistics.

The *coloc*.*abf* function assumes a single causal SNP for each trait and estimates the posterior probability of the following 5 scenarios for each testing region; *H*_*0*_: neither trait has a genetic association, *H*_*1*_: only trait 1 has a genetic association, *H*_*2*_: only trait 2 has a genetic association, *H*_*3*_: both trait 1 and 2 are associated but with different causal SNPs and *H*_*4*_: both trait 1 and 2 are associated with the same single causal SNP. In the case of our study, trait 1 is insomnia and trait 2 is expression of a tested gene. Since we limited the analyses to genes where there was at least one overlap of significant SNPs with insomnia GWS SNPs, this discards scenarios *H*_*0*_ to *H*_*2*_, and we are only interested in whether *H*_*4*_ is most likely. We therefore defined eQTLs of a testing gene as colocalized with insomnia summary statistics when *H*_*4*_ is >0.9. It is possible that genomic regions outside of the pre-defined risk loci can also be colocalized with eQTLs. However, we limited the analyses to within the risk loci in this study, since the primary aim was to prioritize genes linked from the insomnia risk loci.

### Gene mapping with FUMA

We used FUMA to map SNPs to genes with 3 criteria: positional, eQTL and chromatin interaction mappings^20^.

#### Positional mapping

SNPs were mapped to one of 20,260 protein-coding genes with 10kb windows on both sides.

#### eQTL mapping

Significant eQTLs in 49 tissue types from GTEx v8^26^; blood samples from the eQTLGen consortium^28^ and prefrontal cortex samples from PsychENCODE^27^ were used for the mapping. FUMA annotates those significant eQTLs with candidate SNPs and those SNPs are mapped to the gene whose expression is potentially affected by the SNPs.

#### Chromatin interaction mapping

Significant chromatin loops (FDR<1e-6) in 14 tissues defined based on HiC data from Schmitt *et al*.^68^ and pre-processed chromatin loops based on HiC of prefrontal cortex from PsychENCODE^27^ were used for the mapping. In FUMA, the candidate SNPs are required to be overlapped with one end of the loop and transcription start sites (TSS) of genes (500bp up- and 250bp downstream from the TSS) are required to be overlapped with the other end of the loop to be mapped. Since HiC is designed to measure physical interactions of two genomic regions, not all significant loops necessarily contain functional interactions. We therefore, further limited chromatin interaction mapping to those where SNPs are overlapping with enhancer regions and gene TSSs are overlapping with promoter regions predicted by Roadmap consortium^65^ (http://egg2.wustl.edu/roadmap/data/byDataType/dnase/). We used all available 113 cell types for enhancers and promoters.

We performed gene mapping for all genome-wide significant SNPs and credible SNPs, separately. We further performed further filtering outside of FUMA, as described details in the main text.

### Tissue and cell type association, and gene-set enrichment tests with prioritized genes

Associations of tissue and cell type specific gene expression with prioritized genes were tested with a linear regression model using *lm* function in R. We defined the model as follows to correct for the average expression across tissues or cell types within a dataset and gene size:

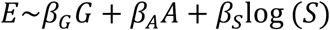

where *G* is a binary status reflecting whether the gene was prioritized (1) or not (0), *E* is a tissue/cell type specific expression value, *A* is the average expression of the gene across all available tissues/cell types in a dataset, *S* is a gene length. We performed a one-sided test (*β*_*G*_>0) which evaluates how well the prioritized gene status predicts the specificity of gene expression in a testing tissue/cell type. For tissue/cell types significantly associated with the prioritized genes, we performed conditional analyses as below and performed a one-sided test (*β*_*G_1*_>0 and *β*_*G_2*_>0).

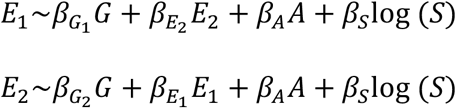

For gene-set enrichment analyses, a one-sided hypergeometric test (greater) was performed. We tested the same datasets as tested in the MAGMA gene-set analysis (53 tissues, 54 and 106 brain regions, 728 cell types, and 5,033 gene-sets). As is done in MAGMA v1.07, gene-property (gene expression value) was truncated when the value is above or below 5 standard error from the mean. The analyses were limited to genes available in each dataset out of 20,260 protein-coding genes based on Ensembl v92 GRCh 37.

We performed Bonferroni correction across all 5,974 tested items (0.05/5974=8.4e-6).

### Conditional analysis for associated tissues, cell types and gene-sets using the full GWAS results and the set of prioritized genes

To identify independent association signals in gene-set or gene-property associations, we performed conditional analyses for sets that were significantly associated with insomnia per dataset (i.e., GTEx, AHBA low/high resolution, scRNA and gene-sets) in both the analyses based on the full genome-wide distribution (using linear regression as implemented in MAGMA) and the set of prioritized genes (using linear regressions, where we used tissue or cell type specific gene expression value as an outcome of a linear regression and binary status of genes as a predictor while conditioning on average expression and gene size). Note that the conditional analyses were performed for gene-sets identified by analysis based on full GWAS results but not for the set of prioritized genes since the enrichment was tested by the hypergeometric test. To systematically identify independent and dependent associations of gene-properties (or gene-sets), and group them into clusters of independent associations, we set thresholds for conditional P-value and proportional significance (PS) of conditional P-value relative to marginal P-value. To simplify, we use cell type A and B for examples hereafter, while same applies to tissue type or gene-set. Here we denote, p_A_ and p_B_ as the marginal P-values of cell type A and B where p_A_<p_B_, p_A,B_ as the P-value of cell type A conditioning on cell type B, and PS_A_ as the proportional significance of cell type A conditioning on cell type B (computed as -log10(p_A,B_)/-log10(p_A_)). We then defined relationship between cell type A and B based on the decision rules as below (ordered according to the priority of the rules).

- When PS_A_≥0.8 and PS_B_≥0.8, associations are independent.
- When p_A,B_≥0.05 and p_B,A_≥0.05, both cell types are jointly associated and both are equally likely true signals (grouped into a same cluster).
- When PS_A_≥0.5 and p_B,A_≥0.05, the association of cell type B is confounded by cell type A and cell type B is discarded.
- When PS_A_≥0.5 and PS_B_<0.2, the association of cell type B is mostly dependent on cell type A and cell type B is discarded.
- When PS_A_≥0.2 and p_B,A_≥0.05, the association of cell type A is partially jointly explained by cell type B but the association of cell type B is confounded by A, therefore cell type B is discarded.
- When PS_A_≥0.2 and PS_B_<0.2, the association of both cell types are partially jointly explained while the association of cell type B is largely depending on cell type A, therefore cell type B is discarded.
- When PS_A_≥0.5 and PS_B_<0.5, same as above.
- When PS_A_<0.2 and PS_B_<0.2, while there are still remaining signals by conditioning on each other, both cell types are largely jointly explained (grouped into a cluster).
- When PS_A_≥0.2, PS_B_≥0.2, p_A,B_≥0.01 and p_B,A_≥0.01, although there are still small signal remaining in both cell types, associations of both cell types are considered as jointly explained (grouped into a cluster).
- When PS_A_≥0.2 and PS_B_≥0.2, both cell types are partially jointly explained while there are remaining signals, therefore cell type A and B are grouped into separate clusters.
- When cell type A and B are collinear (in a case when regression fails), they are considered as joint associations and grouped into a cluster.

The conditional analyses were performed with the most significant sets until all cell types are denoted independent, clustered or discarded.

## Genetic correlation

We first selected 551 GWASs (with 551 unique traits) which showed *h*^*2*^_*SNP*_>0.01 and Z-score>2 from 558 GWASs analyzed previously in the study of Watanabe *et al*.^8^, excluding insomnia and trouble falling asleep (depression item). We estimated genetic correlations of insomnia sex-combined meta-analysis with 551 traits using LDSC^13^. Pre-computed LD scores for 1000 Genome Phase 1 European subjects were downloaded from https://data.broadinstitute.org/alkesgroup/LDSCORE/. The analyses were limited to HapMap3 SNPs and the MHC region was excluded. Additionally, SNPs with chi-square statistics >80 were excluded.

## Colocalization of risk loci and clustering

Colocalization of 554 insomnia risk loci was performed using *coloc* package in R as described in the section of eQTL colocalization. Risk loci with a length <10kb were expanded to 10kb by centering the top SNP. Colocalization of each of 554 insomnia risk loci was tested with GWAS summary statistics of 350 traits by selecting overlapping SNPs, and loci were considered colocalized when *H*_*4*_>0.9.

We performed t-distributed stochastic neighbor embedding (tSNE)^69^ 100 times and the optimal solution was obtained by minimizing the Kullback-Leibler divergence. The clustering of traits was performed on tSNE 2D matrix using DBSCAN and the clustering cut-off was optimized by maximizing the silhouette score. One percent of the data points were allowed to be ‘un-clustered’. Since tSNE projects data into a certain number of dimensions based on the similarity of data points, traits that do not share any colocalized loci or share less than other traits can form a cluster. To distinguish this from a cluster where traits within the cluster share more colocalized loci than others, we tested for each cluster whether the number of shared colocalized loci within clusters is larger than between clusters, with Mann-Whitney U test (one-sided, greater). Clusters that did not show significant difference (*p*≥0.05) were discarded. In the same way, insomnia risk loci were projected into 2D map with tSNE and dense clusters were identified based on colocalization patterns across traits.

## Data Availability

The GWAS summary statistics for the 23andMe data set will be made available through 23andMe to qualified researchers under an agreement with 23andMe that protects the privacy of the 23andMe participants. Please visit https://research.23andme.com/collaborate/#publication for more information and to apply to access the data.

## Endnotes

## Acknowledgements

We thank both UK Biobank and 23andMe participants who consented to participate in research and researchers who collected and contributed the data. D.P. was funded by The Netherlands Organization for Scientific Research (NWO VICI 453-14-005), NWO Gravitation: BRAINSCAPES: A Roadmap from Neurogenetics to Neurobiology (Grant No. 024.004.012), and a European Research Council advanced grant (Grant No, ERC-2018-AdG GWAS2FUNC 834057). E.J.W.V.S. was funded by the European Research Council (ERC-ADG-2014-671084 INSOMNIA). The research has been conducted using the UK Biobank Resource (application no. 16406). Analyses were carried out on the Genetic Cluster Computer hosted by the Dutch National computing and Networking Services SurfSARA. We additionally thank the GTEx Portal for providing RNA-seq data. Members of the 23andMe Research Team are: Michelle Agee, Stella Aslibekyan, Adam Auton, Robert K. Bell, Katarzyna Bryc, Sarah K. Clark, Sarah L. Elson, Kipper Fletez-Brant, Pierre Fontanillas, Nicholas A. Furlotte, Pooja M. Gandhi, Karl Heilbron, Barry Hicks, Karen E. Huber, Ethan M. Jewett, Yunxuan Jiang, Aaron Kleinman, Keng-Han Lin, Nadia K. Litterman, Jennifer C. McCreight, Matthew H. McIntyre, Kimberly F. McManus, Joanna L. Mountain, Sahar V. Mozaffari, Elizabeth S. Noblin, Carrie A.M. Northover, Jared O’Connell, Steven J. Pitts, G. David Poznik, J. Fah Sathirapongsasuti, Janie F. Shelton, Jing Shi, Suyash Shringarpure, Chao Tian, Joyce Y. Tung, Robert J. Tunney, Vladimir Vacic, and Wei Wang. The research was based in part on data from the Million Veteran Program – Office of Research and Development, Veterans Health Administration – supported by awards CSP #575B and Merit 1I01CX001849.e.

## Author contributions

D.P. conceived the study. K.W. performed the analyses. J.S. performed quality control on the UKB data and wrote the analysis pipeline. P.N., D.H., X.W., and the 23andMe Research Team contributed and analyzed the 23andMe cohort data. J.G., D.F.L., R.P., and M.B.S. performed PGS analysis for the MVP cohort. E.J.W.V.S and A.B.S provided valuable discussions. K.W., P.R.J. and D.P. wrote the paper.

