## Supplemental info for "Genome-wide meta-analysis of insomnia in over 2.3 million individuals implicates involvement of specific biological pathways through gene-prioritization"

**Supplementary Note**

**1. Estimation of polygenicity and discoverability of insomnia**

To estimate the polygenicity and discoverability of insomnia, we used the causal mixture model for GWAS summary statistics (univariate MiXeR) proposed by Holland *et al.*[1,2] (https://github.com/precimed/mixer). In this model, the distribution of SNP effect sizes is treated as a mixture of two gaussian distributions for causal and non-causal SNPs as the following[1]:

$$\beta = \pi N(0, \sigma_\beta^2) + (1 - \pi)N(0,0)$$

where $\pi$ is the proportion of (independent) causal SNPs and $\sigma_\beta^2$ is the variance of the effect sizes of causal SNPs. Therefore, $\pi$ and $\sigma_\beta^2$ respectively represent polygenicity and discoverability of the trait. As recommended in the original study, we used 1000 Genome Phase 3 European subjects as a reference panel and restricted to HapMap 3 SNPs. SNPs with chi-square statistics >80 and the MHC region (chr6:26Mb-34Mb) were excluded.

We observed a total SNP heritability of 4.3% of which 17.4% (SE=8.29e-4) is explained by genome-wide significant SNPs.

MiXeR estimated polygenicity of $\pi$=4.85e-3 and discoverability of $\sigma_\beta^2$=4.34e-6. By comparing the polygenicity with previously reported traits[3], insomnia is the 3rd most polygenic trait following major depressive disorders ($\pi$=5.75e-4, $\sigma_\beta^2$=6.02e-6) and educational attainment ($\pi$=4.89e-4, $\sigma_\beta^2$=1.21e-5).

To estimate the sample size required to explain 90% of the $h2_{SNP}$, we used the output of GWAS power estimates calculated in the MiXeR software, which contains 51 data points of sample size and the proportion of SNP heritability explained. By using the *interp1* function from the pracma package in R, we estimated that ~57 million are needed to explain 90% of the $h2_{SNP}$ with GWS SNPs.

**2. Polygenic score prediction**

The semi-out of sample polygenic score (PGS) prediction was computed for three randomly selected 10,000 UKB target hold-out samples (**Methods**), which explained 2.0% of the phenotypic variance at most (**Extended Data Fig. 1** and **Supplementary Table 2**). This is slightly lower than previously reported (2.6%)[4].

We further performed PGS prediction for an independent cohort consists of 45,355 insomnia cases and 138,589 controls (the Million Veteran Program (MVP), see **Methods** for details).

Using the full meta-analysis results to compute PGS, at most 0.66% of the phenotypic variance was explained at (**Supplementary Table 2**).

Although the sample prevalence of the MVP cohort (24.7%) is somewhat close to the UKB and 23andMe combined samples (25.1%), the predictive power for the MVP cohort based on UKB and 23andMe meta-analysis was considerably lower than the subset of UKB samples based on remaining UKB and 23andMe meta-analysis. The higher prediction power of the latter analysis is likely because the training samples are from relatively homogenic cohort and part of samples from the same cohort are included in the training sets, or because there is phenotypic heterogeneity.

### 3. Suspicious loci

Using FUMA[6], we identified 558 risk loci from the sex-combined meta-analysis. Of those, 8 loci contained only a single SNP. We visually examined those loci and 4 loci were considered likely false positives as none of the nearby SNPs showed any potential signals (**Extended Data Fig. 3**). These 4 loci were marked in **Supplementary Table 3** and were excluded from any analyses except MAGMA gene/gene-set analysis, because these analyses are based on aggregate association values across many SNPs in a gene and are therefore not strongly influenced by any single SNP.

### 4. P-value threshold for genome-wide significance

Four GWAS studies have been performed for insomnia to date[4,10–12], which makes it possible to compare GWAS findings with different sample sizes. We noticed that 18 out of 202 loci (8.9%) identified from the previous GWAS meta-analysis were not replicated in the current study suggesting they may have been false positives[4]. The significance of those loci was mostly borderline in the previous GWAS; i.e. 14 loci with $p$<1e-8 and 4 with $p$<5e-10. It has been recently debated whether the 'golden-standard' of the genome-wide significance threshold, $p$<5e-8 is still applicable as the number of accessible SNPs increases with increasing sample size[7]. Although this threshold was preserved in the current study, we compared the number of independent loci with different P-value thresholds. We observed that 23.6% of loci are no-longer significant by decreasing the threshold to 1e-8 from 5e-8 and the number of loci almost exponentially decreased along with the decreasing P-value threshold (**Extended Data Fig. 4**). To evaluate whether this proportion (i.e. 23.6%) is specific to insomnia, we conducted similar analyses for three other traits with GWAS outcomes for multiple sample sizes: educational attainment[8] (https://www.thessgac.org/data), height[3] (https://atlas.ctglab.nl/traitDB/3187) and type 2 diabetes[9] (https://www.diagram-consortium.org/downloads.html). Risk loci were defined in the same way as for insomnia GWAS. In these three traits we observed 23.5%, 17.0% and 22.5% decrease of the number of risk loci for educational attainment, height and type 2 diabetes, respectively by decreasing the P-value threshold to 1e-8 from 5e-8. Thus all three traits showed very similar proportions of decreasing number of the loci along with the decreasing P-value threshold with insomnia GWAS (**Extended Data Fig. 4**).

**5. Effects of increasing sample size in insomnia GWASs**

Here we further sought to investigate changes in association P-values and effect sizes of SNPs in insomnia GWASs as a function of sample size from ~100k up to over 2 million. Using the 4 previously published GWAS studies as well as the current study, we compared the sample size and number of detected risk loci from six GWAS cohorts: UK Biobank 1st release (UKB1, N=113,006), UK Biobank latest release (UKB2, N=386,988), 23andMe 1st GWAS (23andMe1, N=944,477), meta-analysis of UKB1 and 23andMe1 (meta1, N=1,331,010), 23andMe 2nd GWAS (23andMe2, N=1,978,022) and meta-analysis of UKB2 and 23andMe2 (meta2, N=2,365,010; main analysis of the current study). We do note that several of these cohorts overlap (i.e., UKB2 includes UKB1, 23andMe2 includes 23andMe2, Meta1 is UKB1+23andMe1, and Meta2 is UKB2+23andMe2). As expected, the number of risk loci almost linearly increased with the sample size (**Fig. 1b**).

As mentioned above, several risk loci identified in the previous insomnia meta-analysis (Meta1) were not replicated in this study (Meta2). To investigate whether the distributions of P-values differ between replicated and non-replicated risk loci, we categorized risk loci into 4 types: Type 1) loci that are only detected in a single GWAS Type 2) novel loci identified in the corresponding GWAS and replicated in the GWAS(s) with larger sample size but not seen in GWAS of smaller sample size, Type 3) loci identified in the GWASs with both smaller and larger sample sizes, and Type 4) loci identified in the GWAS(s) with smaller sample size(s) but not in the GWAS with larger sample size. Note that type 1 and 4 in meta2 are potential type 2 and 3 loci, respectively. We did not include 23andMe1 in this analysis since association statistics are not publicly available. As we expected, type 1 loci tend to show P-values just below the genome-wide significance threshold (*p*=5-e8), which showed significantly higher P-value compared to other loci (*p*=7.4e-19, Mann-Whitney U test, twosided, excluding SNPs from UKB1 and meta2; **Extended Data Fig. 5a**). Novel (type 2) loci tend to show higher P-values compared to type 3 and 4 loci that replicates loci identified by GWAS with less sample sizes ($p$=2.5e-28), indicating a decreasing P-value of the GWAS loci with increasing sample sizes (**Extended Data Fig. 5a**). However, type 4 loci did not show significant difference of P-values compared to type 2 loci ($p$=0.31; **Methods**). Although type 4 loci are potential false positives (as they are no longer replicated with larger sample sizes), it is hard to distinguish from the novel (type 2) loci based on our observation.

We further investigated the changes in effect sizes of SNPs with increasing sample size. We selected SNPs which reached genome-wide significance in at least 3 GWASs (out of 5, excluding GWAS of 23andMe1 as the summary statistics are not available) and selected one SNP per locus with the minimum P-value in case there were multiple SNPs available (**Methods**). Thus, the evaluated SNPs do not necessarily represent the most significant SNP in the corresponding locus in a particular GWAS. This resulted in 140 unique SNPs. From each GWAS, standardized effect sizes were computed for each SNP, and absolute effect size was regressed on the sample size per SNP (**Methods**). We did not observe any significant increase or decrease in standardized effect sizes as a function of the sample size (**Extended Data Fig. 5b**).

**6. Results for sex specific meta-analyses**

From the male specific meta-analysis (222,753 cases and 993,280 controls), 4,781 SNPs reached genome-wide significance resulting in 100 loci consisting of 114 lead SNPs (**Supplementary Table 7-8**). 90 loci overlapped with one of the 554 loci from the sex-combined meta-analysis. From the female specific meta-analysis (390,750 cases and 1,018,386 controls), 24,181 SNPs reached genome-wide significance resulting in 303 loci consisting of 377 lead SNPs (**Supplementary Table 9-10**). 275 loci overlapped with one of the 554 loci from sex-combined meta-analysis. The larger number of risk loci identified from the female meta-analysis is most likely due to the higher effective sample size (male $N_{eff}$=727,796 vs female $N_{eff}$=1,129,585). In total, there were 10 and 28 loci specifically identified for male and female meta-analyses, respectively (**Extended Data Fig. 6a**). Subsequently, MAGMA gene analyses were performed for the sex specific meta-analyses. We identified 278 (in males) and 871 (in females) significant genes (0.05/19751=2.5e-6) and 191 genes were significantly associated in both sexes (**Supplementary Table 6**). The correlation of -log10 gene-based P-value between the sexes was considerably low ($r$=0.56,

$p$<1e-323; **Extended Data Fig. 6b**), given high genetic correlation ($r_g$=0.92), while the correlation between sex-specific and sex-combined meta-analysis was relatively high, 0.78 (for males) and 0.92 (for females), both with $p$<1e-323 (**Extended Data Fig. 6c,d**). The higher correlation of sex-combined with female-only meta-analysis than male-only meta-analysis could be due to the difference in the effective sample size, which is greater for females than males. The low correlation between sexes suggests there may be sex specific genetic causes to the insomnia, though this needs further investigation.

**7. Genetic overlap with multiple clusters of traits and heterogeneity of insomnia risk loci**

It was previously shown that insomnia is genetically correlated with multiple psychiatric and metabolic traits[4]. To further investigate these associations, we used LDSC[13] to estimate genetic correlations ($r_g$) between insomnia and 551 traits with a SNP $h^2_{SNP}$>0.01 and Z-score>2 reported by Watanabe *et al.*[3], excluding insomnia and one depression item that indexes insomnia (trouble falling asleep; **Methods**). After Bonferroni correction, 350 traits showed a significant $r_g$ with insomnia (0.05/551=9.1e-5), of which 270 traits showed positive and 80 traits showed negative correlations with insomnia (**Extended Data Fig. 7** and **Supplementary Table 13**). The strongest positive correlation was with major depressive disorder (MDD; $r_g$=0.65, $p$=2.7e-178), in line with previous findings[4]. The strongest negative correlation was with health satisfaction ($r_g$=-0.60, $p$=3.9e-110). We also observed that insomnia was positively correlated with multiple cardiovascular, metabolic, psychiatric diseases/disorders, in agreement with previous reports (**Extended Data Fig. 7** and **Supplementary Table 13**).

We next sought to identify insomnia risk loci sharing the same causal SNPs with the 350 significant genetically correlated traits by colocalizing each of the 554 risk loci with the GWAS summary statistics of the 350 traits (**Methods**). Of the 554 loci, 282 were colocalized with at least one of, in total, 227 traits (**Supplementary Table 14**). Body Mass Index showed the greatest number of colocalized loci with insomnia (41 loci), and 18 other metabolic traits colocalized with >20 loci (**Supplementary Table 15**). From other trait domains, height (27 loci), overall health rating (26 loci), educational attainment (24 loci) and neuroticism (23 loci) showed the greatest number of colocalized loci among the others (**Supplementary Table 15**). Despite the high $r_g$ with insomnia, MDD only colocalized with 5 loci.

To investigate colocalization patterns of insomnia risk loci across traits, we counted the number of shared colocalized loci between each pair of 227 traits and projected them onto a

2D-map using the t-distribution stochastic neighbor embedding (tSNE; **Methods**). Subsequently, we performed density-based spatial clustering of applications with noise (DBSCAN) on the tSNE 2D-map to identify clusters of traits. To identify clusters of traits that are more likely to share colocalized insomnia loci, we tested whether the number of shared loci within each cluster was higher than the number of shared loci between clusters with Mann-Whitney U tests (one-sided, greater). Clusters with $p \geq 0.05$ were discarded (**Methods**). We identified 5 dense clusters of traits (**Extended Data Fig. 8a**). Cluster #1 contained the greatest number of traits (82) and was dominated by metabolic traits (33) but also included traits from multiple domains such as height, intelligence, health rating, diabetes, high blood pressure and sleep duration (**Supplementary Table 15-16**). Cluster #2 consisted of 29 traits, of which 24 were psychiatric traits including neuroticism, ever smoker, risk taking and broad depression (**Supplementary Table 15-16**). Cluster #3 consisted of 21 traits, of which 7 were nutritional traits (e.g. intake of food and drinks), but the cluster also included multiple cardiovascular traits such as CAD and high blood pressure as well as body fat percentage (**Supplementary Table 15-16**). Clusters #4 and #5 consisted of 7 and 5 traits, respectively. Both clusters included traits from multiple domains and the majority of traits showed less than 3 loci colocalized with insomnia.

Next, we evaluated whether specific insomnia loci show similar colocalization patterns across 227 traits. To do so, we counted the number of shared colocalized traits for each pair of 282 loci and projected onto a 2D-map using tSNE (**Methods**). In the same way as done for the trait clustering, loci were clustered based on DBSCAN. We identified 11 dense clusters of loci (**Extended Data Fig. 8b**). These clusters were not driven by the location of loci on the genome as chromosomes of loci did not form a cluster (**Extended Data Fig. 8c**). Additionally, by projecting loci which contain prioritized genes that are part of the significantly associated gene-set (regulation of nervous system development), we did not observe a cluster specific to the gene-set (**Extended Data Fig. 8d-g**). On the other hand, clusters of loci are likely to be representing the specificity of colocalized traits. Three clusters of loci were mainly colocalized with metabolic traits (cluster #1, 3, and 9), and 3 other clusters of loci mainly with psychiatric traits (cluster #2, 4, and 6; **Extended Data Fig. 8b**). Other clusters also aggregated in trait domains, e.g. reproduction (cluster #8), cardiovascular (cluster #10 and 11), while 2 clusters were colocalized with multiple traits from multiple domains (**Supplementary Table 17-18**). Although the colocalized loci suggest there is a shared causal SNP between a pair of traits, this does not infer a causal relationship between traits. There are two possible scenarios when loci are colocalized: i) the same causal SNP is pleiotropic which directly cause both traits, ii) trait A causes trait B and GWAS on trait B captures causal SNPs of trait A or iii) misclassification of samples. It is statistically challenging to infer causal relationship (and direction) between traits as existing methods, such as Mendelian randomization, require strict assumptions[14,15] which are often violated. Nonetheless, we observed independent clusters of loci specifically colocalized with metabolic and psychiatric traits. These results suggest that there might be independent pathogenic mechanisms behind insomnia depending on the background of other metabolic and psychiatric traits.

**8. Tissue, cell type and gene-set association analyses using the full GWAS results**

We first assess whether insomnia associated genes converge into biological functions, tissue types or specific cell types, using the conventional strategy based on the full GWAS results and weighting by strength of association but without first prioritizing genes. MAGMA gene-property and gene-set analysis[16] was performed on 54 tissue types from GTEx v8[17], 54 lower and 106 higher resolution of brain regions from the Allen Human Brain Atlas (AHBA)[18], 885 brain cell types from 9 single-cell RNA-sequencing (scRNA-seq) datasets[19–25], and 6,089 gene-sets with at least 20 genes from MsigDB v7.0[26] and SynGO v1.0[27] (**Methods**). Bonferroni correction was applied across all tested items (0.05/7188=7.0e-6). In each dataset, pairwise conditional analyses were performed for significantly associated items and confounders[28]; tissues, cell types or gene-sets whose associations were mostly explained by a more significantly associated item within the same dataset are not reported (see **Methods** for details, full results are available in **Supplementary Table 19-30**). For scRNA-seq datasets, we also performed cross-datasets conditional analyses as proposed previously[29] (**Methods**).

*Tissue analyses reveal association with cerebral cortex*

Tissue specificity analysis with GTEx showed two groups of significantly associated brain regions; cerebellar hemisphere/cerebellum and cortex where the most significant association was seen in cerebellar hemisphere (P=2.7e-19) (**Fig. 2** and **Supplementary Table 19**) in line with previous findings[4]. Previously identified basal ganglia also showed significant association in the current study, however, this association was almost completely explained by the association of cerebellum (**Supplementary Table 20**). We note that the specificity of these associated brain regions was defined relative to the average expression of genes across all available tissue types in GTEx (i.e. brain and non-brain), and thus may reflect a general effect in brain. Indeed, when we conditioned on the average expression of genes across 13 brain regions, the significance of specific brain regions was largely decreased (**Extended Data Fig. 9a** and **Supplementary Table 19**). Therefore, these results using the GTEx resource implied a general association of insomnia with brain-specific expression and did not reveal enrichment in specific brain areas.

To gain insight into more specific brain regions associated with insomnia, we tested enrichment in 54 brain regions from AHBA (**Methods**). We observed the most significant associations with regions from the cerebral cortex followed by the basal forebrain and preoptic region (**Fig. 2** and **Supplementary Table 21-22**). We then assessed enrichment in 106 more specific brain regions from AHBA (**Methods**). Out of 106 regions, 32 regions, all from the cerebral cortex, showed significant associations with insomnia (**Fig. 2** and **Supplementary Table 23**). These associations were not independent and indicated a general association with cerebral cortex (**Supplementary Table 24**).

*Cell type analyses reveal associations with neuronal cell types in sub-cortical regions*

Next, we aimed to identify cell-type specificity using brain specific scRNA-seq datasets. We used 9 datasets[19–25] consisting of 885 cell types (**Methods**). The most significant association was seen in neurons from the lateral geniculate nucleus (LGN, P=9.9e-16) from DropViz which was collinear with GABAergic neurons from embryonic mouse midbrain dataset (P=2.3e-6). In addition, neurons in habenula, ventral pallidum (VP, P=2.2e-13) and anterior pretectal nucleus (APN, P=2.5e-11), all from DropViz, showed (partially) independent associations from LGN (**Fig. 2** and **Supplementary Table 25**). There were four additional neuronal cell types that reached significance from other datasets than DropViz, including the previously identified hypothalamus Vglut 2 neurons[4], however, these associations were largely dependent on the association of LGN (**Supplementary Table 25-28**). Claustrum neurons from DropViz also showed a significant association, in line with previous findings[4] while its association was almost completely explained by the association of LGN (**Supplementary Table 26**). Association of the medium spiny neurons (MSN) was not replicated in the current study, yet we believe this is due to methodological differences explained in reference[29].

We note that although both AHBA and DropViz datasets contain samples from cortical and subcortical regions, AHBA showed more significant associations for insomnia with cortex while DropViz is showed stronger associations with cell types from subcortical regions close to the thalamus and globus pallidus. There are two possible reasons for this discrepancy. First, in the AHBA datasets, associations of cortical regions might be confounded by the true causal cell types which are not available in AHBA dataset. This is difficult to test since we do not know true causal cell types, yet we do know that there are multiple specific neuronal cell types from the frontal and posterior cortex in DropViz that showed significant associations with insomnia whose associations were mostly explained by the LGN, which is not available in AHBA (**Supplementary Table 24**). Second, AHBA is based on microarray data which normally captures a lower number of genes and is limited by the probes compared to more robust RNA-seq. There were ~13,000 genes available in AHBA while there were ~15,000 genes presented in DropViz. Of these ~11,000 genes were available in both datasets. By limiting to the overlapping 11k genes in the DropViz dataset, the significance of associations and effect size of LGN, habenula, VP, and APN showed a notable decrease. The remaining 4k genes that are available in DropViz but not in AHBA showed stronger associations with insomnia than other 11k genes (**Extended Data Fig. 9b** and **Supplementary Table 26**). On the other hand, for AHBA, ~11,000 genes mostly explained the marginal association of cortical regions (**Extended Data Fig. 9b** and **Supplementary Table 26**). Thus, genes which are not available in AHBA contributed strongly to the associations of subcortical regions in DropViz which might explain the discrepancies and in particular the absence of associations with subcortical regions in AHBA.

*Gene-set analyses of functional categories reveals enrichment in synaptic and neuronal development pathways*

Gene-set analysis showed independent association with five gene sets; the most significant with 'process in the synapse' (SynGO:BP), followed by 'behavior' (GO:BP), 'synapse organization' (SynGO:BP), 'synapse part' (GO:CC) and 'regulation of neuron differentiation' (GO:BP) (**Fig. 2, Supplementary Table 29-30**). Significant association of 'behavior' has been previously reported[4], while associations of gene-sets related to functions of synapse and neuronal development are novel (**Fig. 2**).

**9. Results of Fine-mapping**

The fine-mapping was performed for 554 risk loci identified from sex-combined meta-analysis (**Methods**). The number of causal SNPs (*k*) was optimized at *k*=1 in 401 loci (72.4%) and in total 525 loci (94.8%) were optimized at *k*≤5 (**Extended Data Fig. 10a**). There were 15 loci that reached *k*=10 which indicates that either the locus contains ≥10 independent causal SNPs or it was not optimized likely due to complex structure of the local LD or subtle effect sizes. Despite the fact that the majority of loci are likely to contain a single causal SNP (as the *k* was optimized at 1), 112 loci resulted in a set of ≤10 credible SNPs (that are part of 95% credible sets, see **Methods** for details), and 66 loci contained sets of >100 credible SNPs indicating average PIP<0.01in those loci (**Extended Data Fig. 10b**). In addition, only 91 out of 554 loci contained at least one credible SNPs with PIP>0.8 (**Extended Data Fig. 10c**), which increased to 166 and 426 loci by decreasing PIP threshold to 0.5 and 0.1, respectively (**Extended Data Fig. 10d-e**). A large proportion of loci are thus unsolved, which means that the probability of being causal is distributed across relatively large number of SNPs within those loci. It has been previously shown that fine-mapping resolution can be improved using functional annotations as a prior[9], although such methods currently only support *k*=1 and careful selection of annotations to compute priors is required as they can bias the posterior probability. It is also possible that actual causal SNPs are not tagged in the current GWAS. As insomnia is one of the most polygenic traits and effect sizes of single variants are very small, the distribution of effect sizes and LD structure alone might not be sufficient to solve the statistical fine-mapping in some loci. Although PIP>0.8 is a widely employed threshold to select likely causal SNPs from fine-mapping, due to a low coverage of loci, we considered credible SNPs with PIP>0.1 for prioritization of genes from insomnia risk loci, so that each locus is allowed maximum of 10 times *k* credible SNPs.

**10. Additional results for prioritized genes from insomnia risk loci**

To obtain better insights into the distribution of the 'high confidence prioritized' (HCP) genes across insomnia risk loci, we defined 2 additional sets of genes. Of 3,526 genes mapped by FUMA using GWS SNPs, genes from the loci which have at least one of the HCP genes are grouped as 'excluded' and genes mapped from other loci are grouped as 'unsolved'. This resulted in 2,122 excluded and 1,116 unsolved genes. Note that HCP genes are not necessary a subset of 3,526 genes as credible SNPs are not required to have genome-wide significant P-value. Indeed 1 of the 289 HCP genes was outside of 3,526 genes. To investigate the conflict between significant associations observed by full GWAS results and the HCP genes, we conditioned three sets of genes (i.e. HCP, unsolved and excluded genes) on each brain regions (top 5 associations for AHBA high and low resolution), cell types (4 independent associations from DropViz) and gene sets (5 independently associated sets) that showed significant associations based on full GWAS results, using MAGMA. For brain regions (AHBA high and low datasets) and cell types (DropViz), the HCP genes showed slight increase of association P-values compared to the marginal P-value suggesting at least some contribution of the HCP genes (**Extended Data Fig. 11**). The unsolved genes showed greater contributions than HCP genes which may be due to the greater number of genes in the conditioned gene set. On the other hand, excluded genes showed the least contribution to those associations, even though it is the largest set of genes. At the same time, after conditioning either HCP or unsolved genes, there is still a substantially strong signal remaining (**Extended Data Fig. 11**). Therefore, these results suggest that the absence of specific brain region and cell type associations with the HCP genes is likely because those associations are highly polygenic where genes from unsolved loci and outside of GWAS loci are contributing, and the 289 HCP genes are not sufficient to explain these associations. For gene sets, similar pattern was observed as brain regions and cell types, except ‘behavior’ (GO:BP) which did not show notable decrease of the signal by conditioning any of the HCP, unsolved or excluded genes (**Extended Data Fig. 11**). This result suggests the association is mainly driven by genes outside of the GWAS loci. Since the prioritization of genes are limited to GWAS loci, these associations identified by full GWAS are missed from the HCP genes.

**Extended Data**

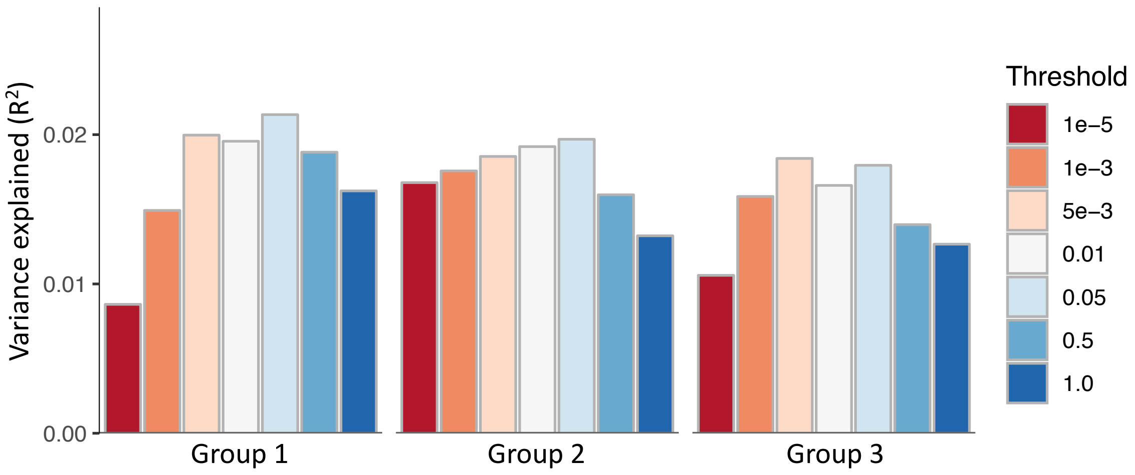

**Extended Data Fig. 1. Phenotypic variance explained by polygenic risk scoring.** Bars are colored by P-value threshold of SNPs used to computed polygenic risk score.

**Extended Data Fig. 2. Locus Zoom plots for 554 loci.** Available in a separate PDF file.

**Extended Data Fig. 3. Locus Zoom plots for 4 suspicious loci.** Available in a separate PDF file.

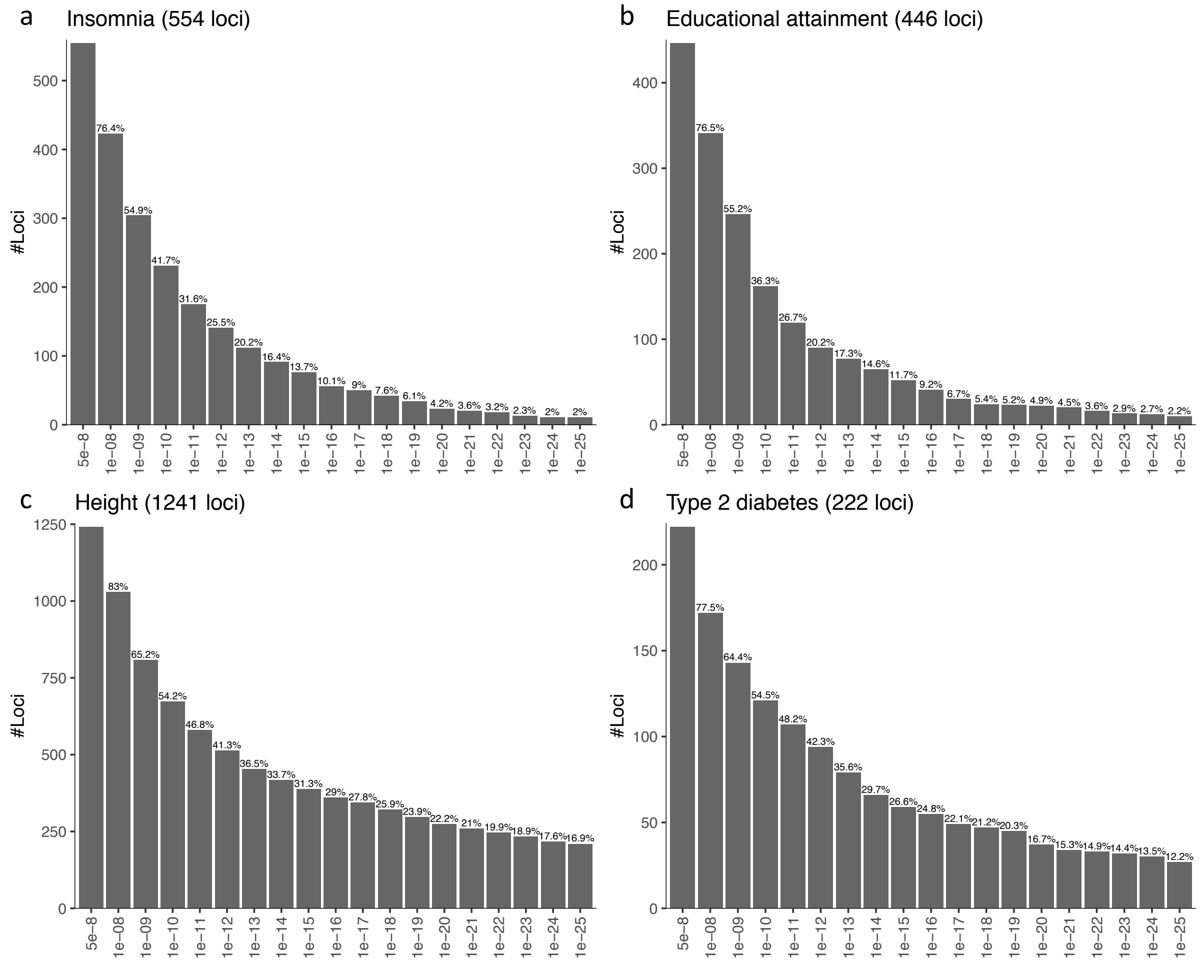

**Extended Data Fig. 4. The number of risk loci with different P-value thresholds.**

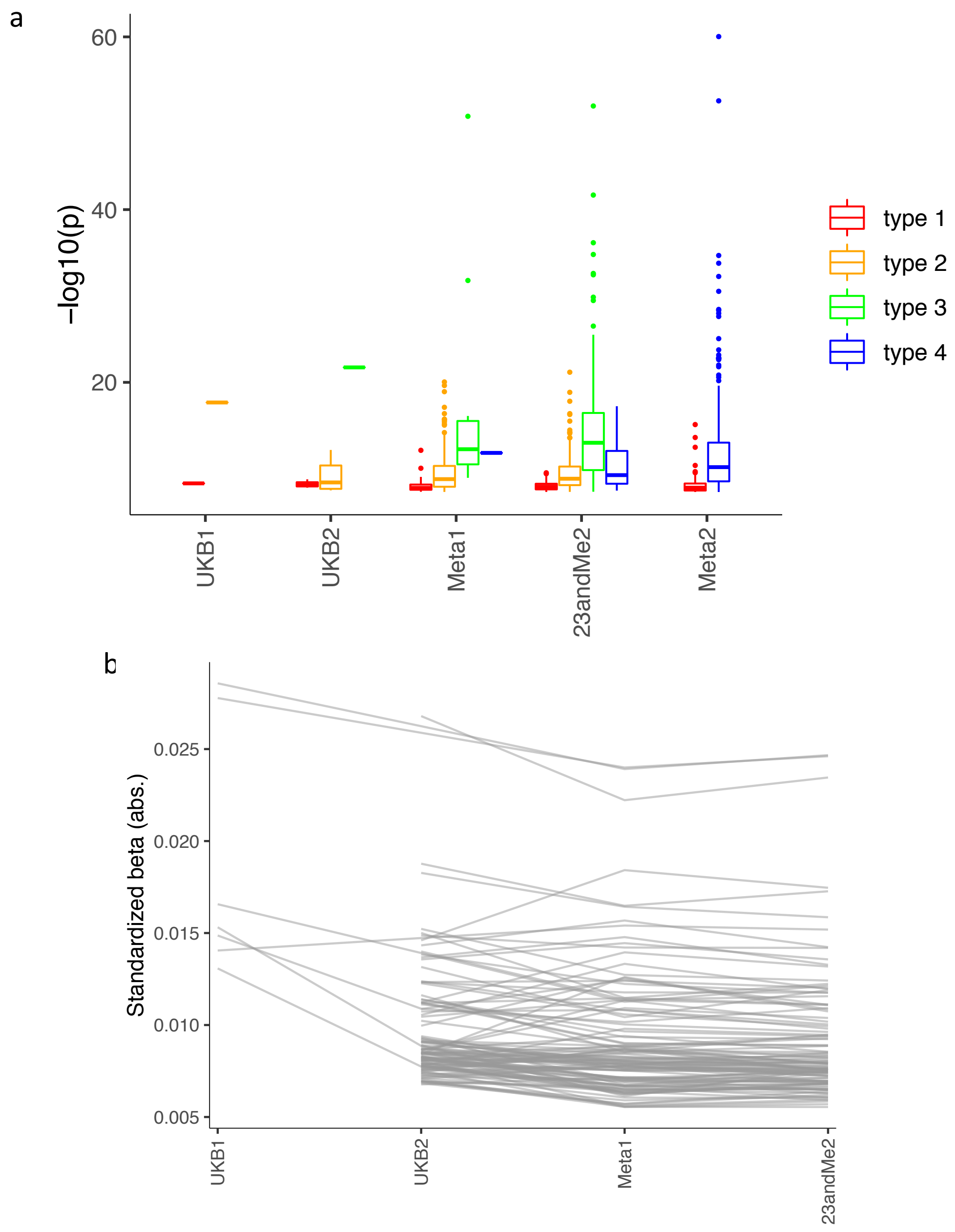

**Extended Data Fig. 5. Comparison of significance and effect sizes across insomnia GWASs with different sample sizes.** (**a**) Box and winkers plot of -log10 P-value of the top SNPs from each risk locus. Outliers are represented by dots. Type 1: loci that are detected once but not replicated by any other GWASs (smaller or larger), type 2: loci that are detected first time and subsequently replicated by a larger GWAS, type 3: loci that are previously identified with smaller GWAS and replicated by a larger GWAS, type 4: loci that are previously identified with a smaller GWAS but not replicated by larger GWAS. (**b**) Absolute standardized effect sizes of genome-wide significant SNPs. Only SNPs that are significant in at least 3 GWASs are displayed.

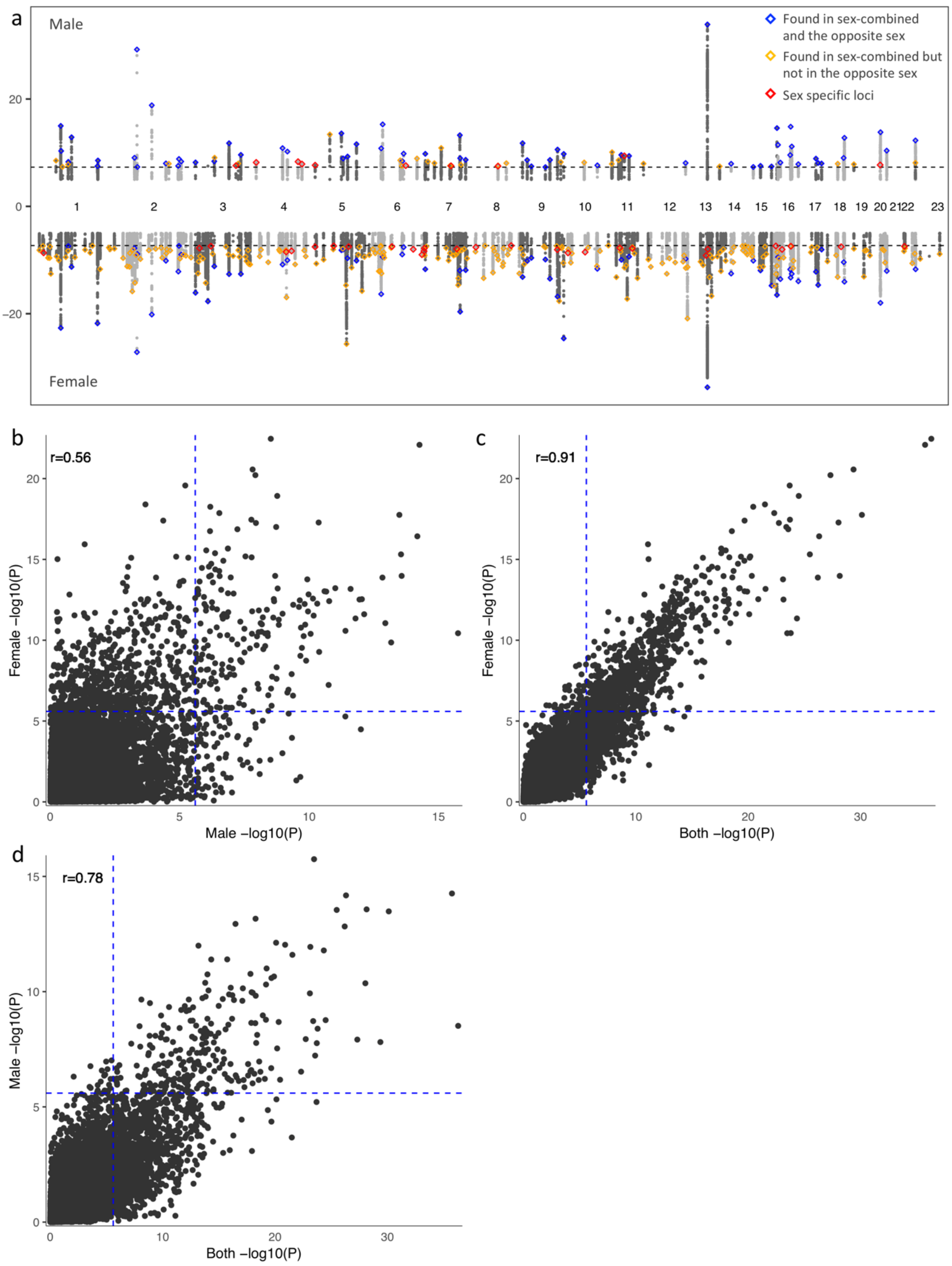

**Extended Data Fig. 6. Comparison of sex-specific insomnia meta-analyses.** (a) Manhattan plot for male only (top) and female only (top) summary statistics. SNPs with $p \geq 1e\text{-}5$ are omitted. Horizontal dashed line represents genome-wide significance (p=5e-8). **(b-d)** Comparison of gene-based P-values computed by MAGMA gene analysis. Blue dashed line represents Bonferroni corrected P-value threshold ($p$=0.05/19751).

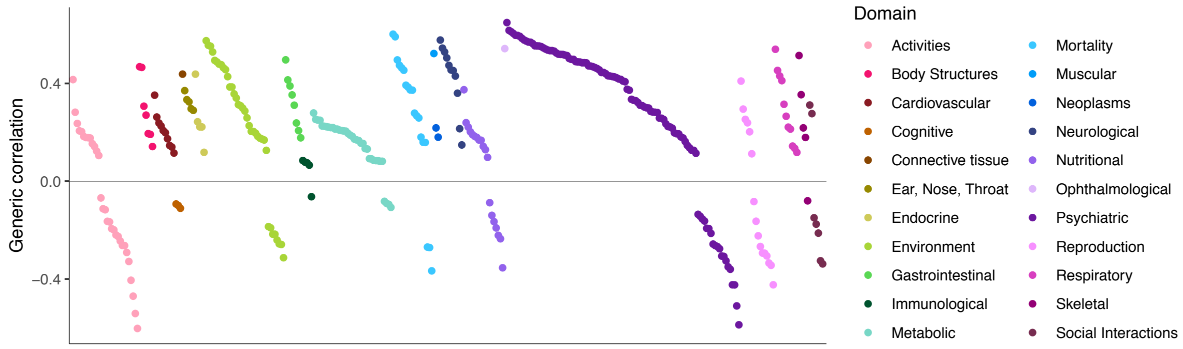

**Extended Data Fig. 7. Genetic overlap between insomnia and 350 traits.** (**a**) Significant genetic correlations with 350 traits. Each data point represents a trait and is colored by the domain category.

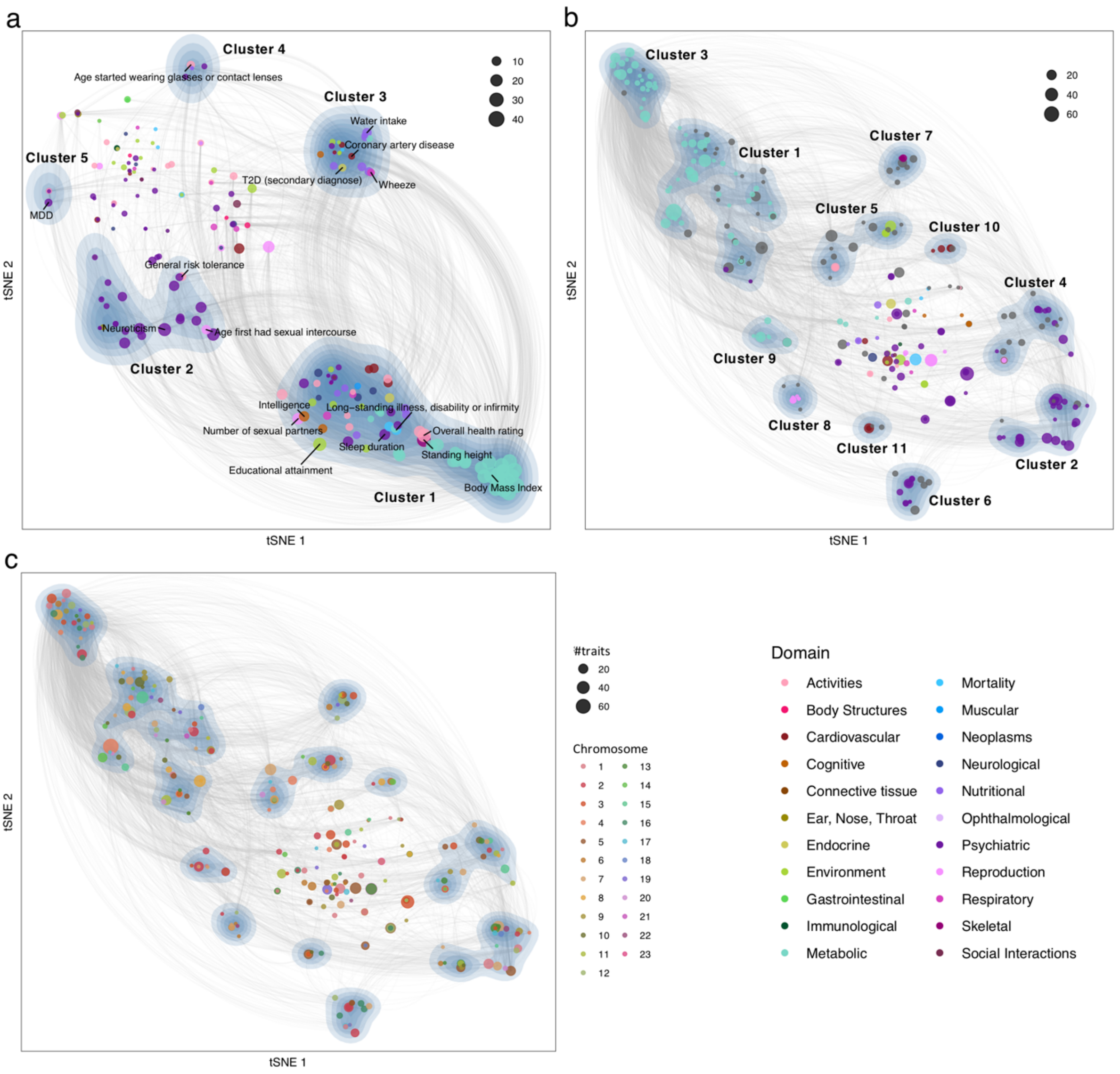

a
Cluster 4
Age started wearing glasses or contact lenses
Cluster 3
Water intake
Coronary artery disease
T2D (secondary diagnose)
Wheeze
Cluster 5
MDD
General risk tolerance
Neuroticism
Age first had sexual intercourse
Cluster 2
Intelligence
Long−standing illness, disability or infirmity
Number of sexual partners
Overall health rating
Sleep duration
Standing height
Educational attainment
Body Mass Index
Cluster 1
10
20
30
40
tSNE 2
tSNE 1
b
Cluster 3
Cluster 1
Cluster 7
Cluster 5
Cluster 10
Cluster 4
Cluster 9
Cluster 8
Cluster 11
Cluster 2
Cluster 6
20
40
60
tSNE 2
tSNE 1
c
tSNE 2
tSNE 1
#traits
20
40
60
Chromosome
Domain
Activities
Body Structures
Cardiovascular
Cognitive
Connective tissue
Ear, Nose, Throat
Endocrine
Environment
Gastrointestinal
Immunological
Metabolic
Mortality
Muscular
Neoplasms
Neurological
Nutritional
Ophthalmological
Psychiatric
Reproduction
Respiratory
Skeletal
Social Interactions

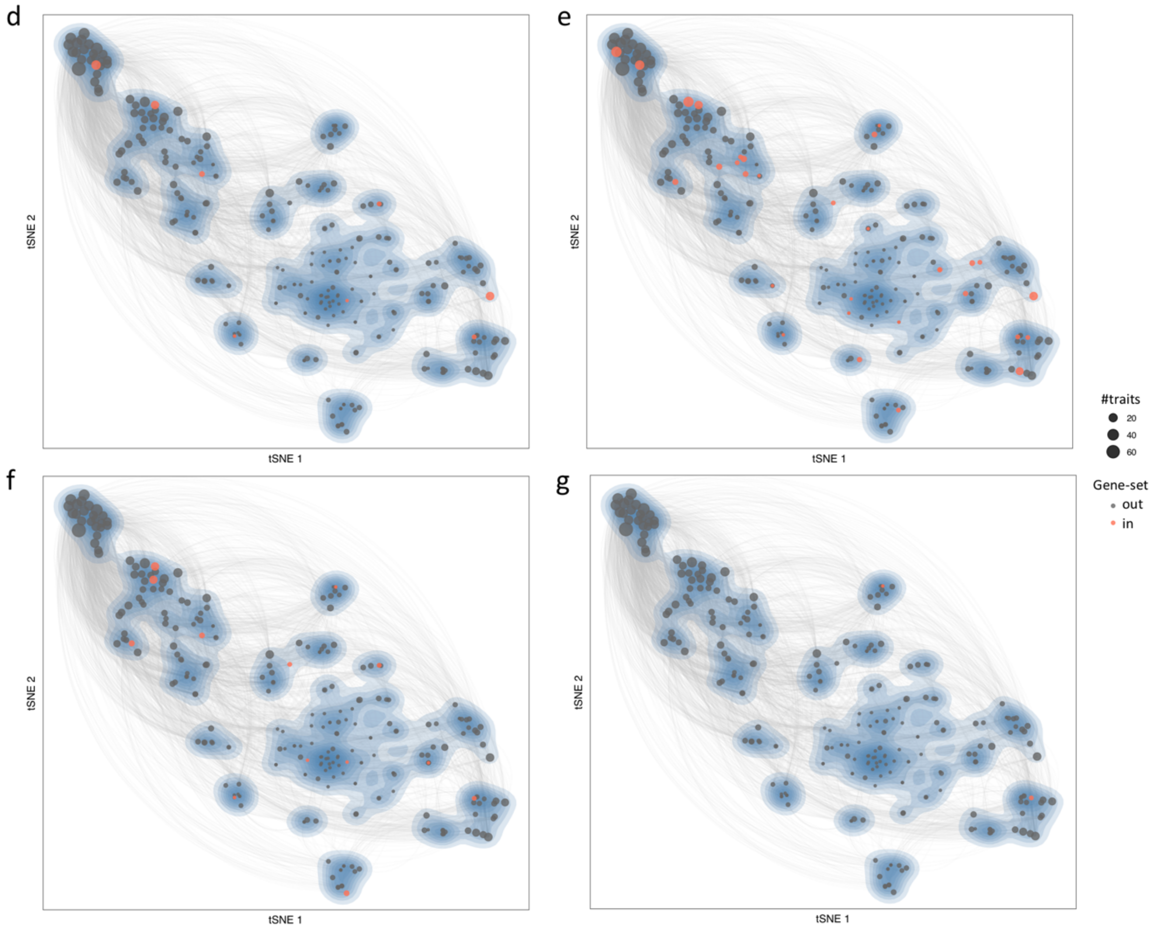

**Extended Data Fig. 8. Clusters traits and insomnia loci based on colocalization pattern.**

(**a**) t-SNE map of traits based on colocalization patterns across 554 insomnia risk loci. 227 traits that had at least one colocalized locus are displayed. Density maps indicate clusters of traits. Each data point represents a trait, colored by the domain and sized by the number of colocalized loci. Grey links between trait represents shared colocalized loci between traits from different clusters (within cluster links are omitted). (**b-g**) t-SNE map of loci insomnia loci based on colocalization patterns across 350 traits. 282 loci that were colocalized with at least one of the 350 traits are displayed. Density map indicate clusters of loci. Each data point represents a locus and sized by the number of colocalized trait. Each locus is colored by the domain of traits which account for >50% of traits colocalized with the locus, otherwise colored in grey (when none of the trait domain represent >50% of the colocalized traits) (**b**), chromosome (**c**) or significantly enriched gene-sets ((**d**) modulation of chemical synaptic transmission (SynGO:BP), (**e**) neuron differentiation (GO:BP), (**f**) regulation of trans synaptic signaling (GO:BP), (**g**) inclusion body (GO:CC)). Full results are available in **Supplementary Tables 40-43.**

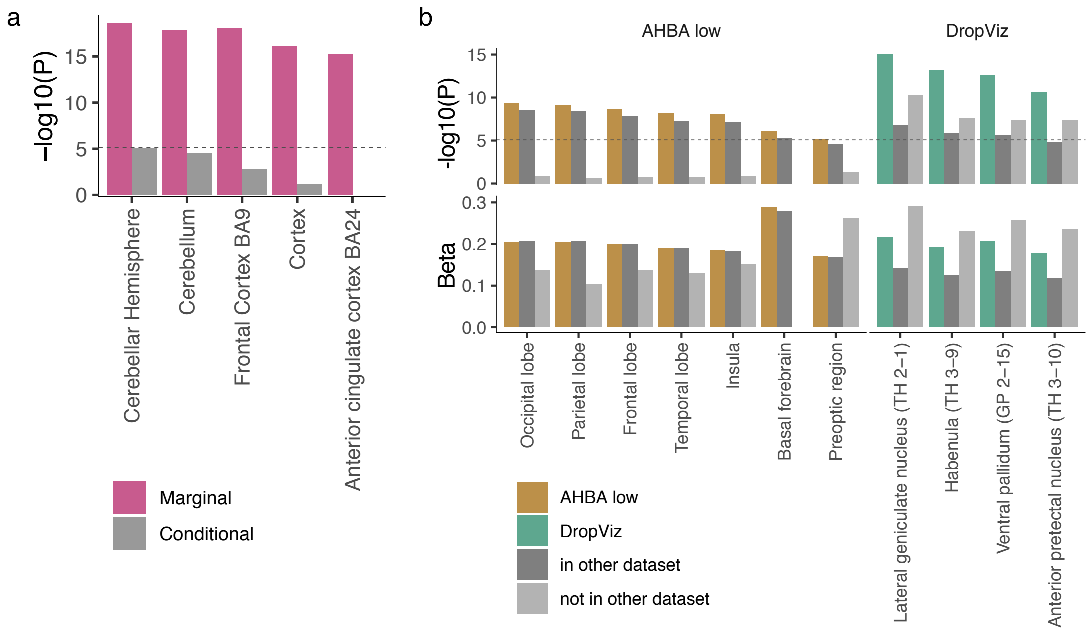

**Extended Data Fig. 9. Additional conditional analyses for MAGMA tissue and brain region association analyses.** (**a**) P-values of brain regions from GTEx, with (Conditional) and without (Marginal) conditioning on the average expression across 13 brain regions. (**b**) Comparison of AHBA (low resolution) and DropViz datasets with MAGMA gene-property analysis. P-values (top) and standardized effect size (Beta, bottom) of brain regions from AHBA low dataset and cell types from DropViz dataset. The most left bar indicates the marginal association statistics for each item. The middle bar indicates the association statistics based only on genes present in both datasets (~11,000 genes). The most right bar indicates the association statistics based only on genes that are not available in the other dataset (~2,000 for AHBA low and ~4,000 for DropViz). The horizontal dashed line indicates the Bonferroni corrected threshold for statistical significance ($p$=0.05/5974).

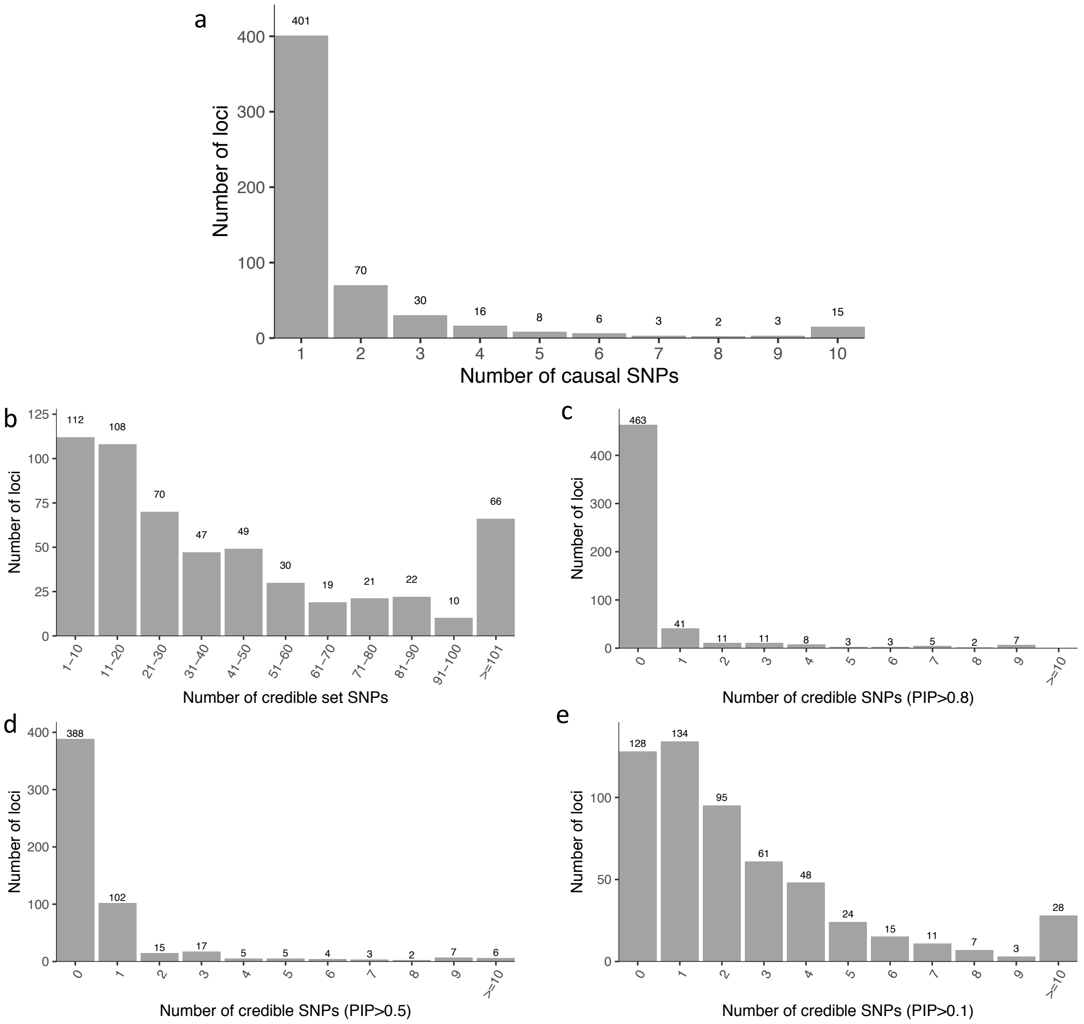

**Extended Data Fig. 10. Summary of fine-mapping results.** (**a**) Distribution of the estimated number of causal SNPs per locus. (**b**) Distribution of the number of credible set SNPs (that are in part of 95% credible sets). (**c-e**) Distribution of the number of credible SNPs with PIP>0.8 (**c**), >0.5 (**d**) and >0.1 (**e**).

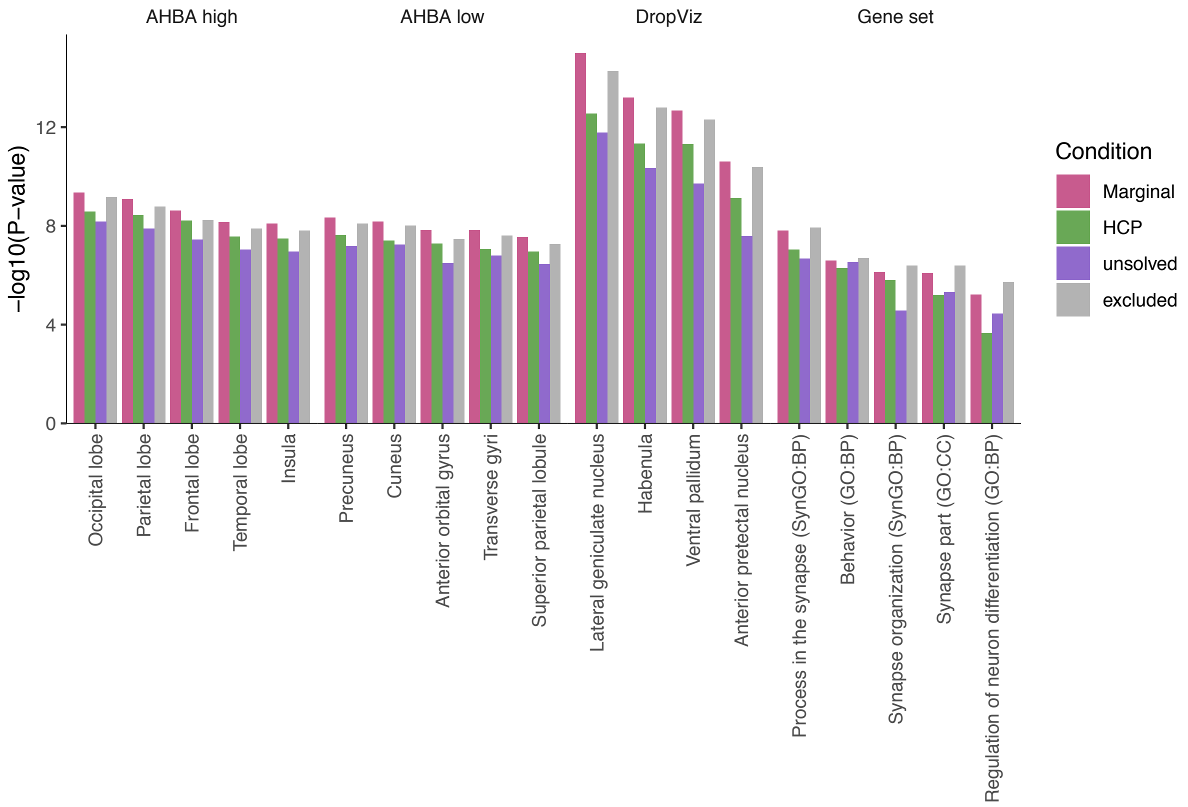

**Extended Data Fig. 11. MAGMA gene-property and gene-set analyses conditioning on sets of genes from insomnia risk loci.** The top (most significantly associated) 5 brain regions/cell types/gene-sets (referred as gene-sets hereafter) were selected for each dataset, except for DropViz where 4 independently associated cell types were selected. For each gene-sets, MAGMA was performed for conditioning 3 set of genes; high-confidence prioritized (HCP), unsolved and excluded genes.

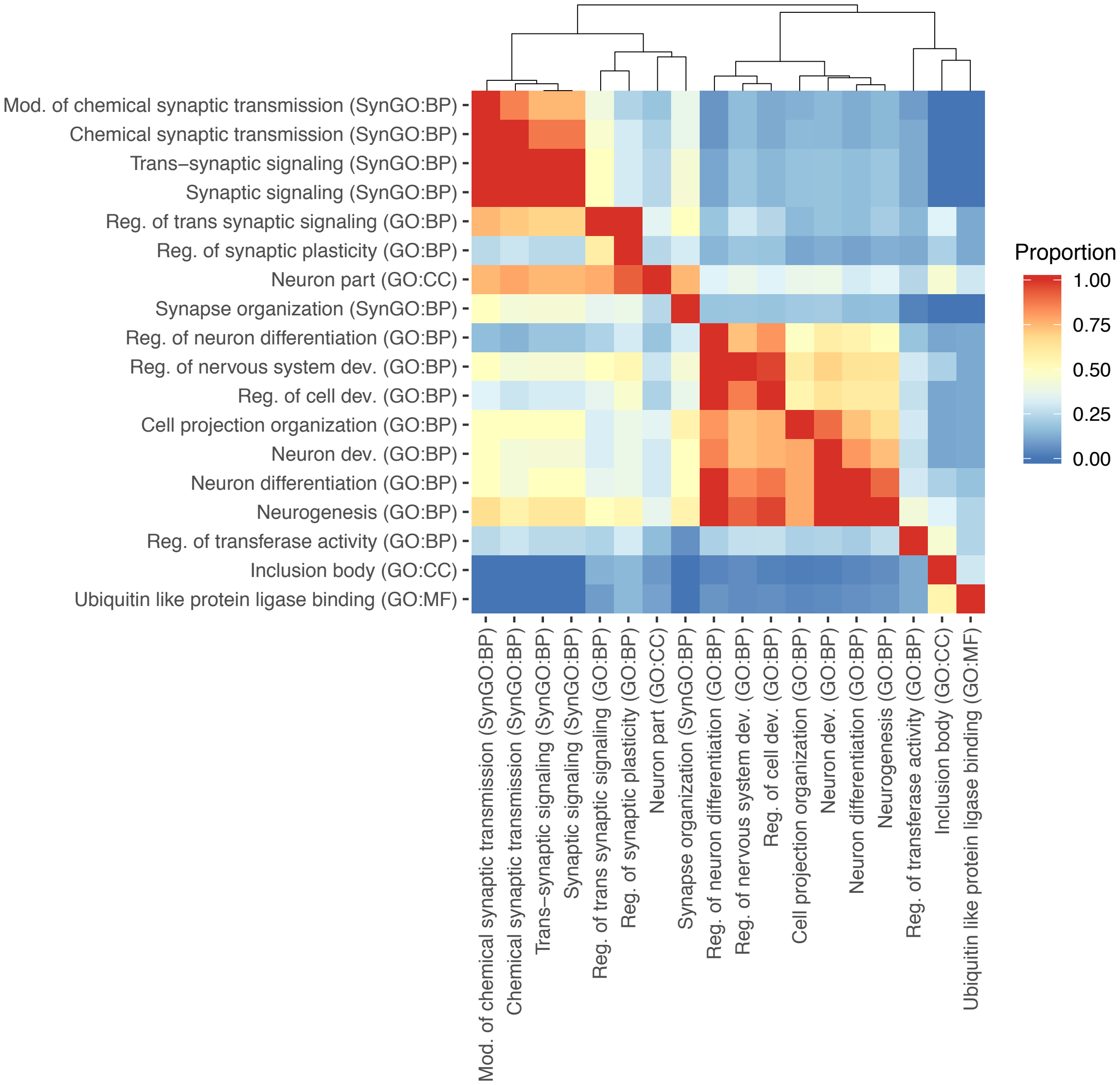

**Extended Data Fig. 12. Heatmap of the overlap of genes across significantly enriched gene-sets.** Displayed 18 gene-sets showed significant enrichment with 289 HCP genes. The heatmap is asymmetric. A cell of row *i* and column *j* represents the proportion of the prioritized genes in the gene-set *i* and *j* relative to the number of the prioritized genes in the gene-set *j*.
