## Supplementary material for "Genome-wide meta-analysis of insomnia in over 2.3 million individuals implicates involvement of specific biological pathways through gene-prioritization": Suppl Fig2

locus 1 rs528964

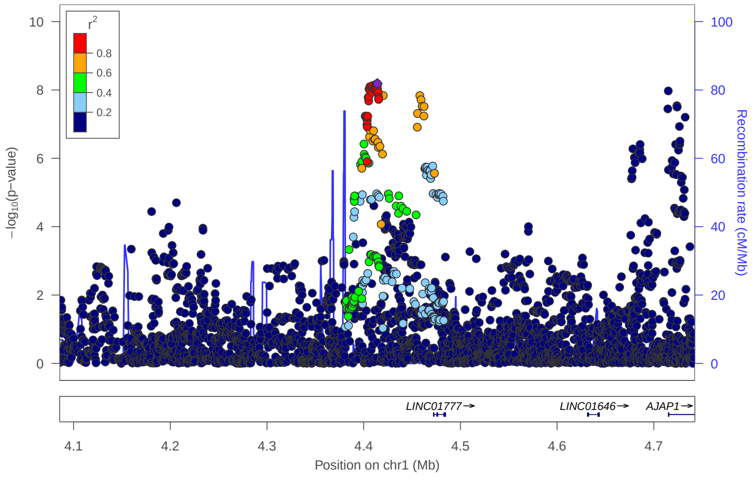


locus 2 rs148116709

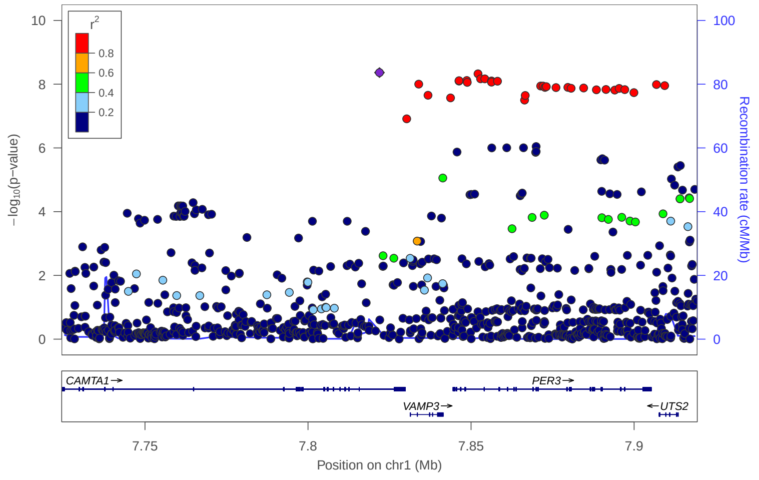


locus 3 rs10927840

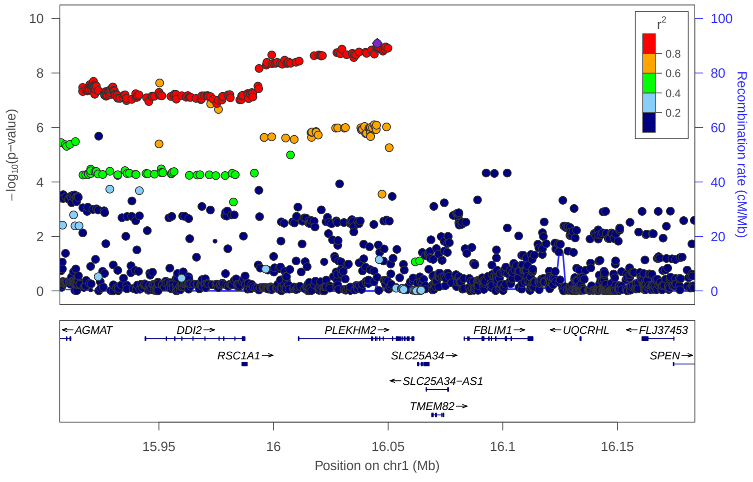


locus 4 rs112358407

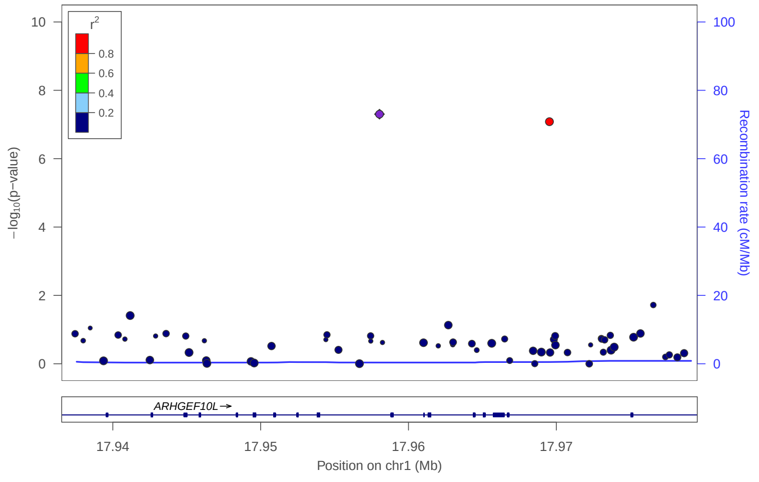


locus 5 rs55634335

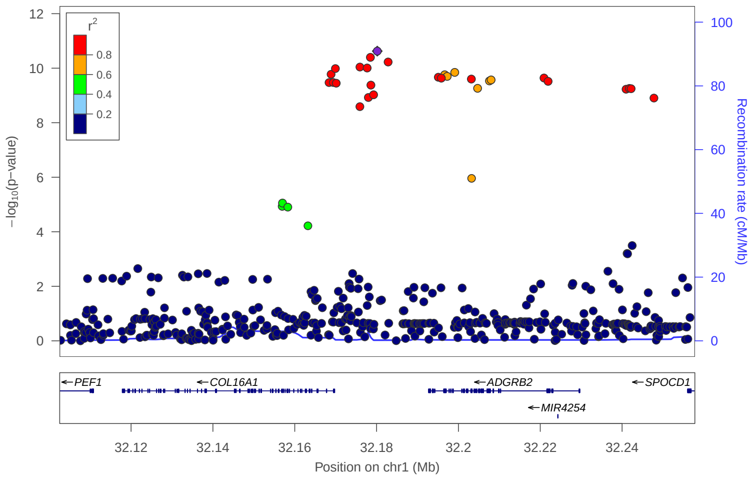


locus 6 rs11577667

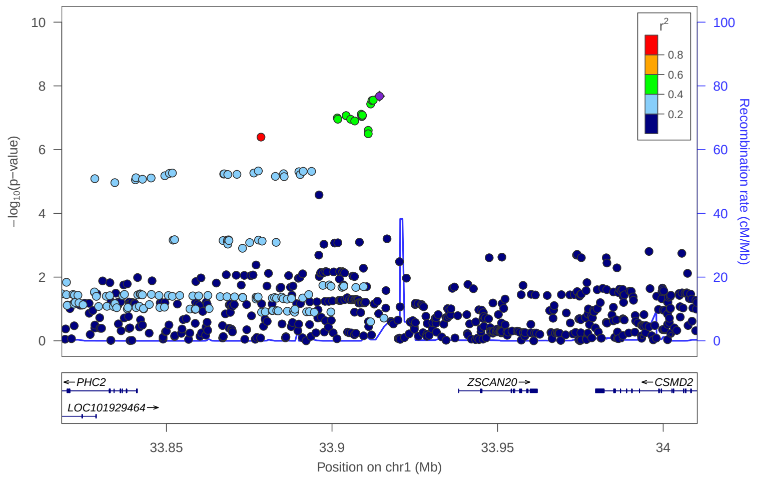


locus 7 rs72659623

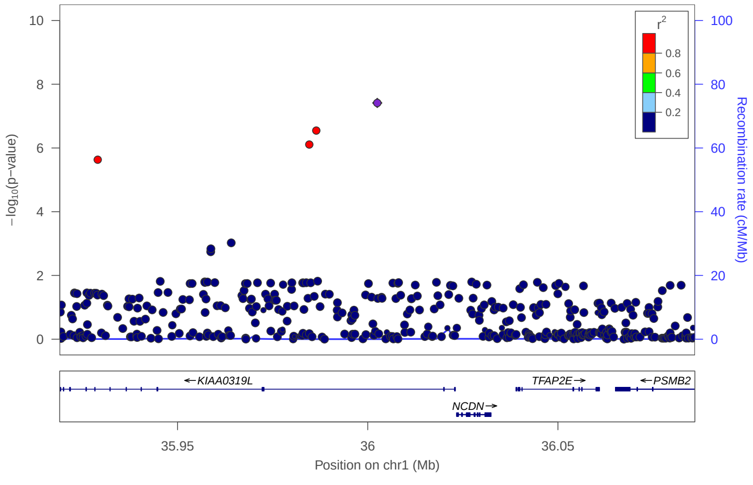


locus 8 rs218985

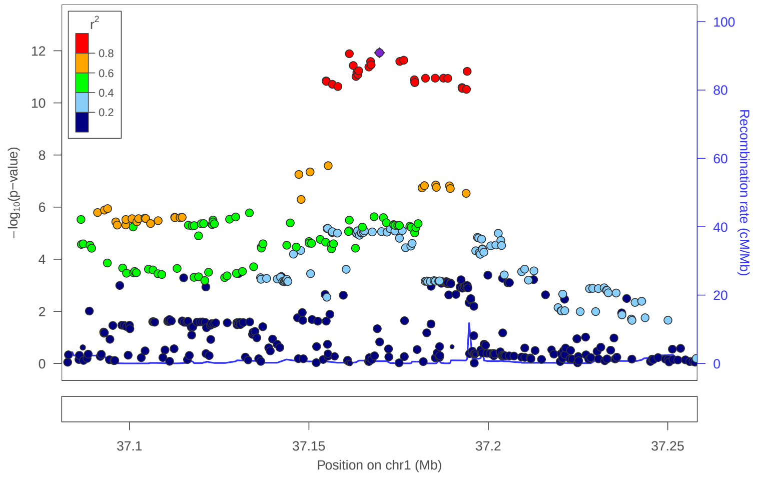

locus 9 rs1564119

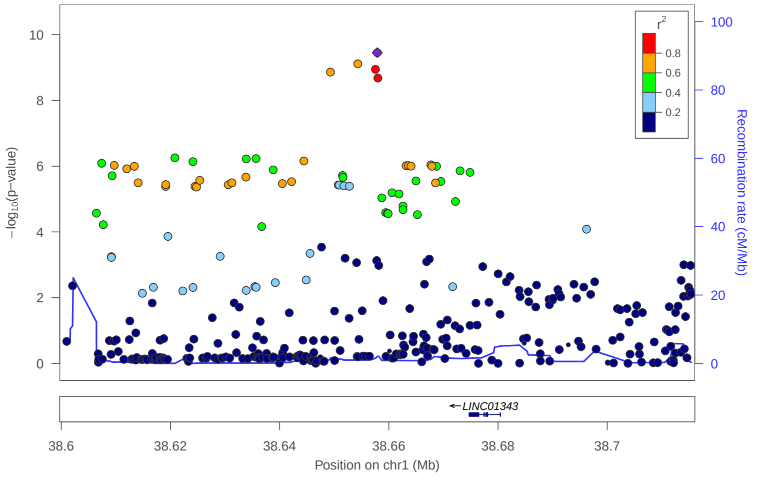


locus 10 rs579448

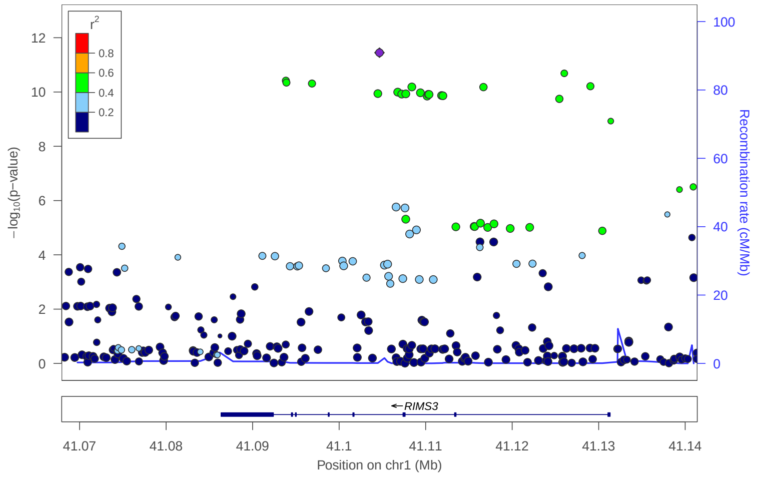


locus 11 rs4134058

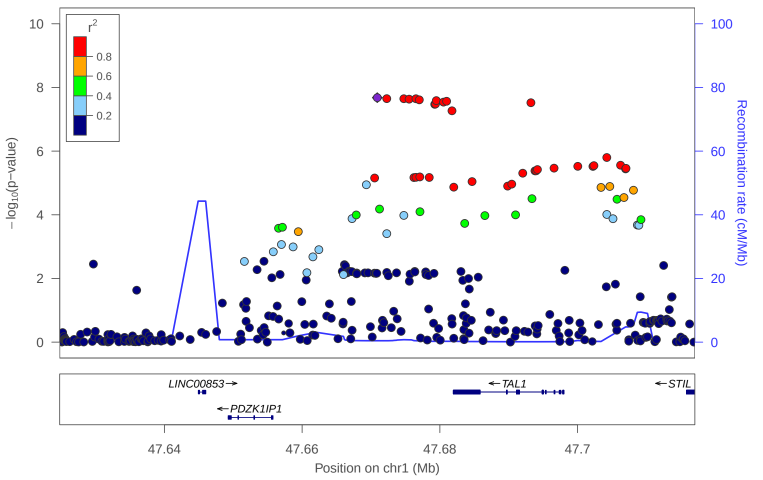


locus 12 rs12137221

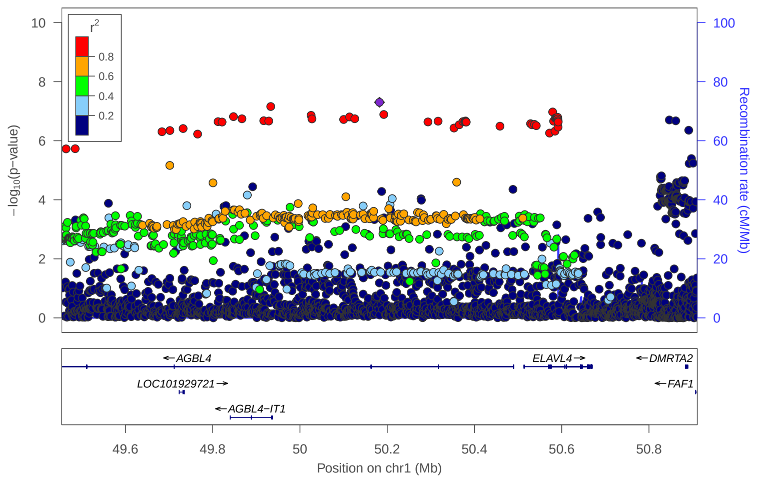


locus 13 rs543144

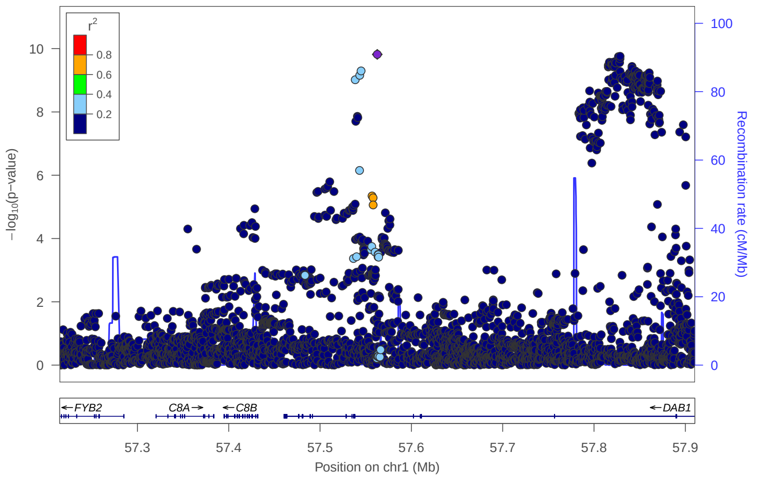


locus 14 rs61788417

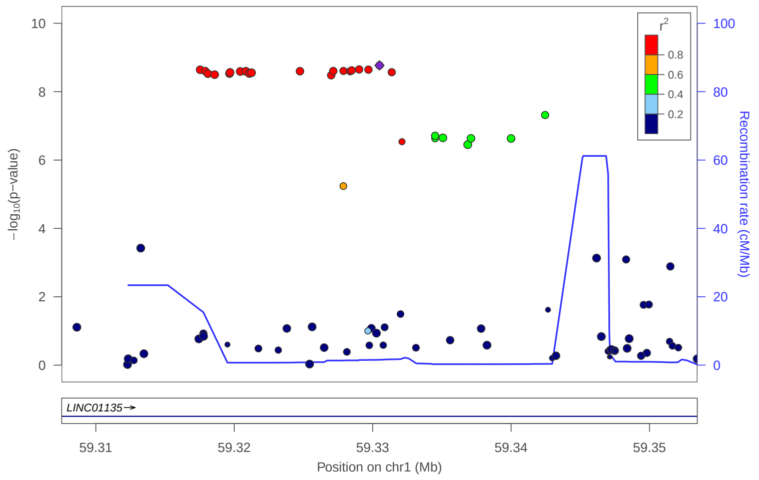


locus 15 rs2476194

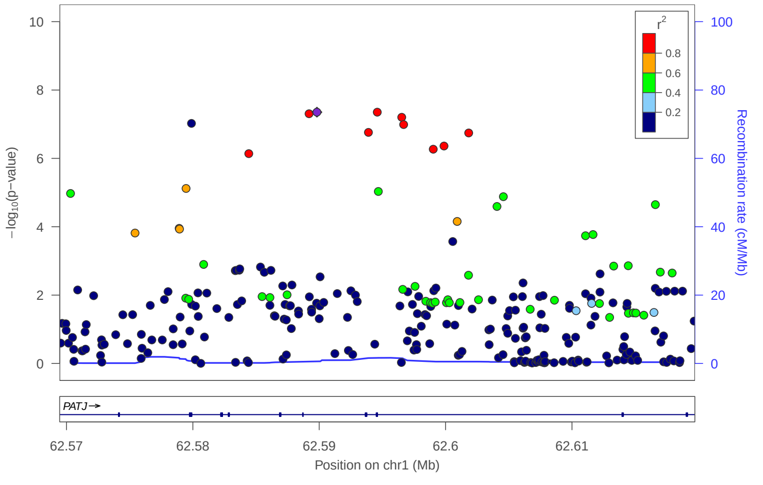


locus 16 rs6661750

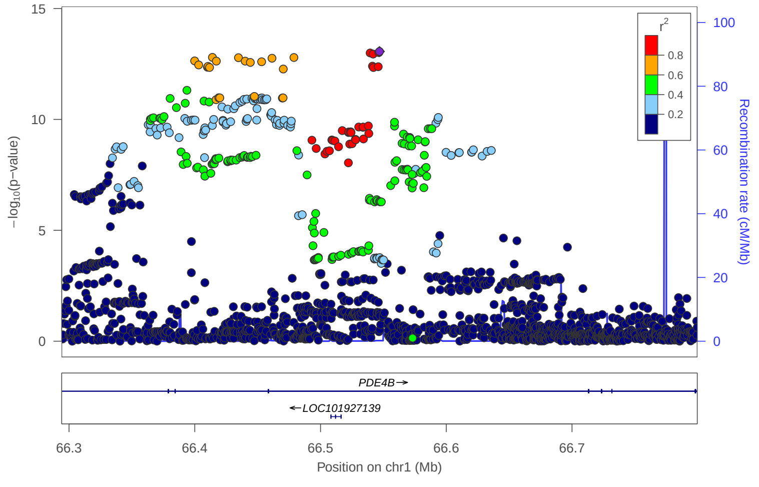

### locus 17 rs3008892

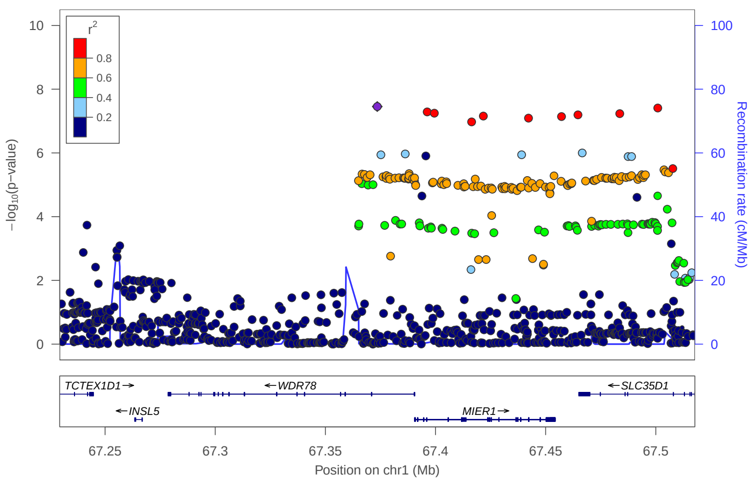


### locus 18 rs2568960

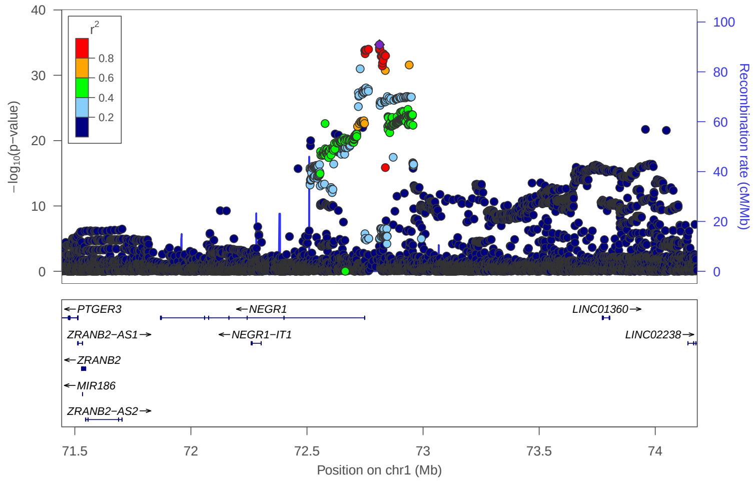


### locus 19 rs4650265

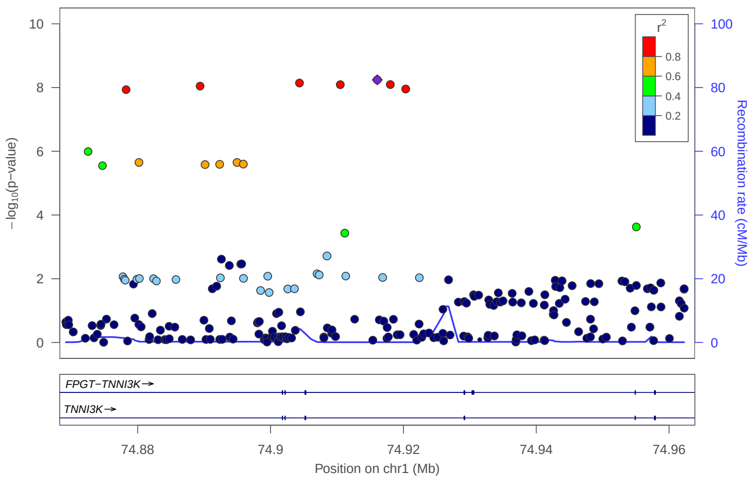


### locus 20 rs34517439

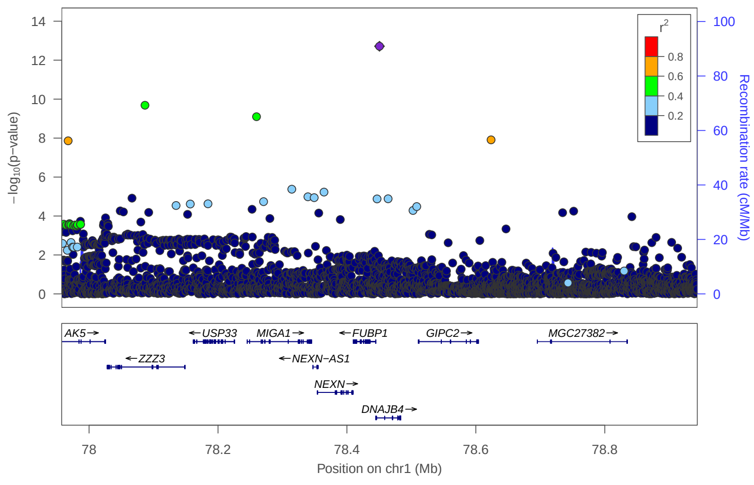


### locus 21 rs72673689

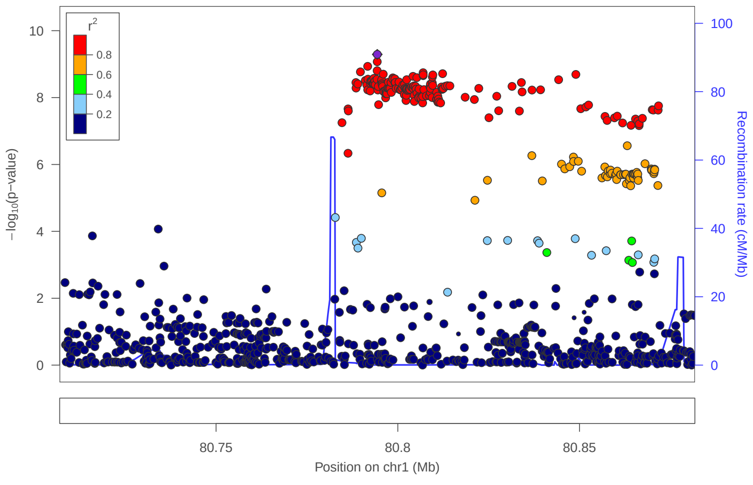


### locus 22 rs503460

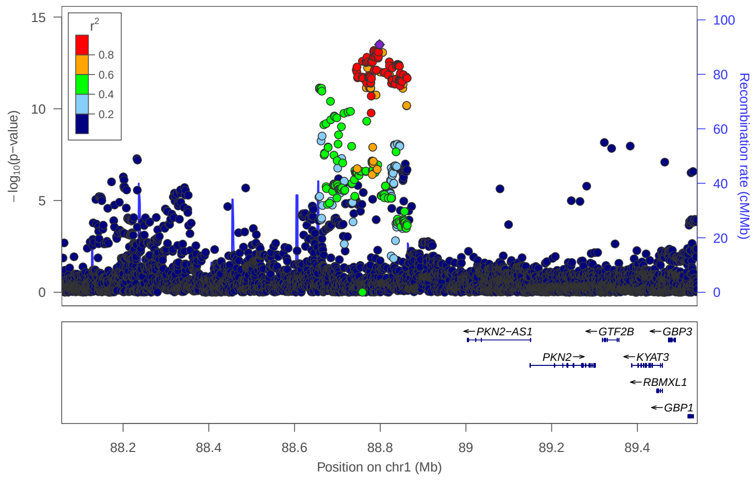


### locus 23 rs12093332

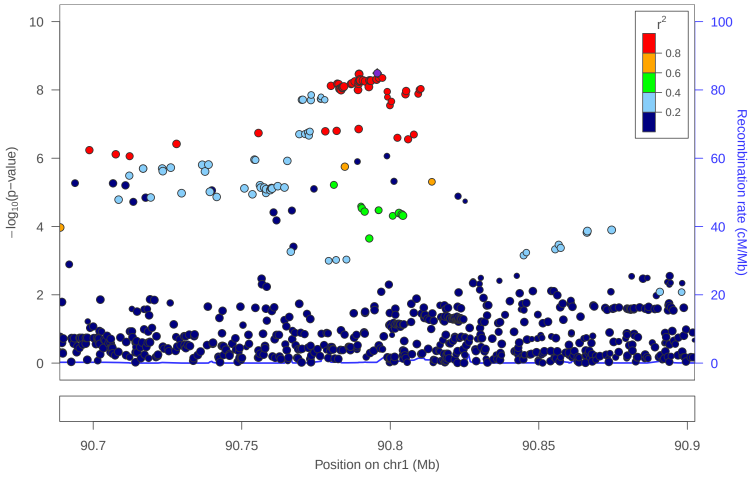


### locus 24 rs11165657

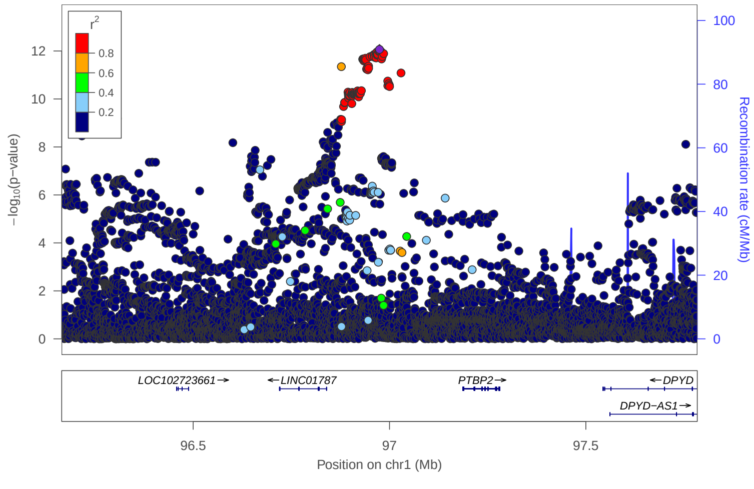

locus 25 rs12039093

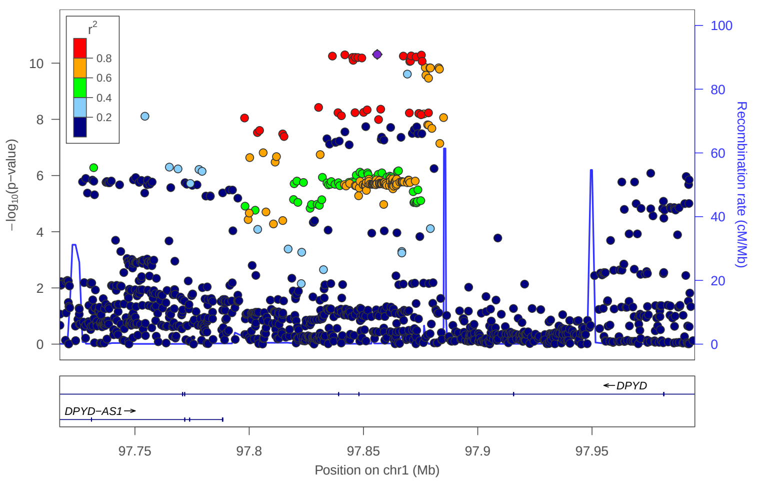


locus 26 rs9325358

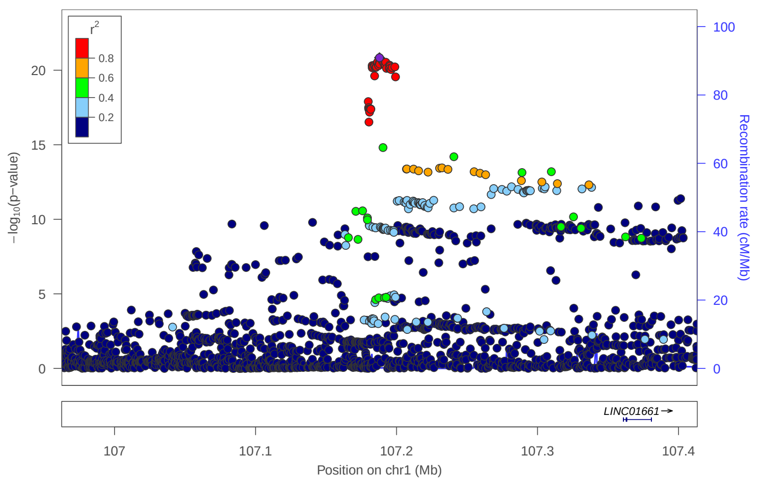


locus 27 rs1768790

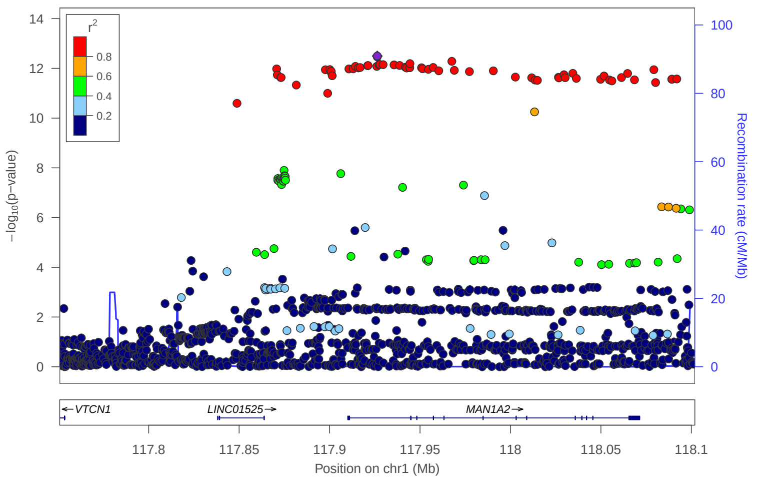


locus 28 rs834233

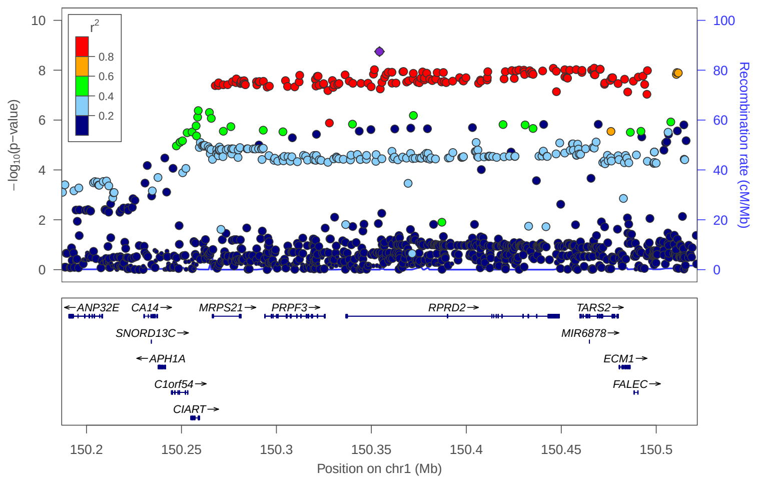


locus 29 rs11588258

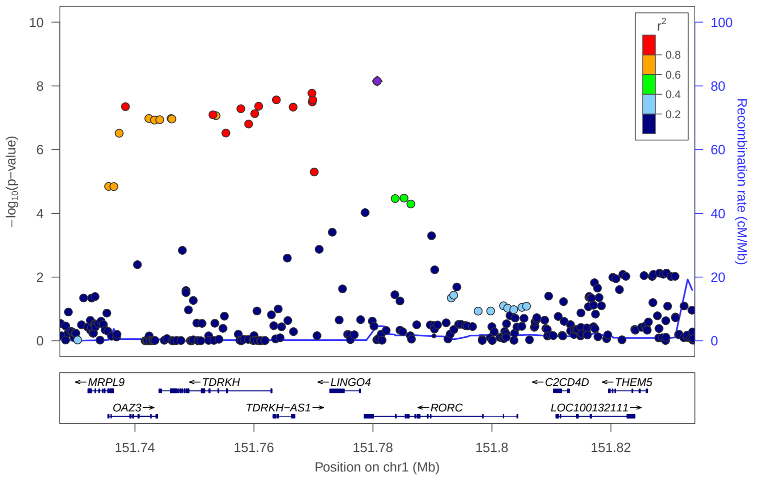


locus 30 rs12023901

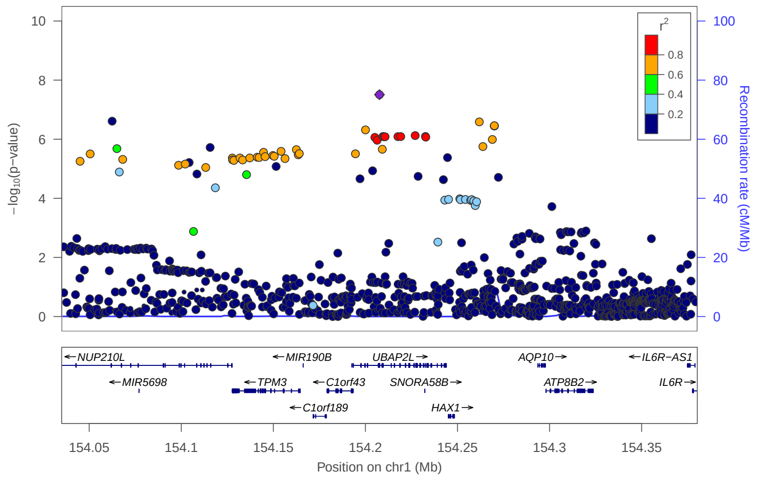


locus 31 rs7551881

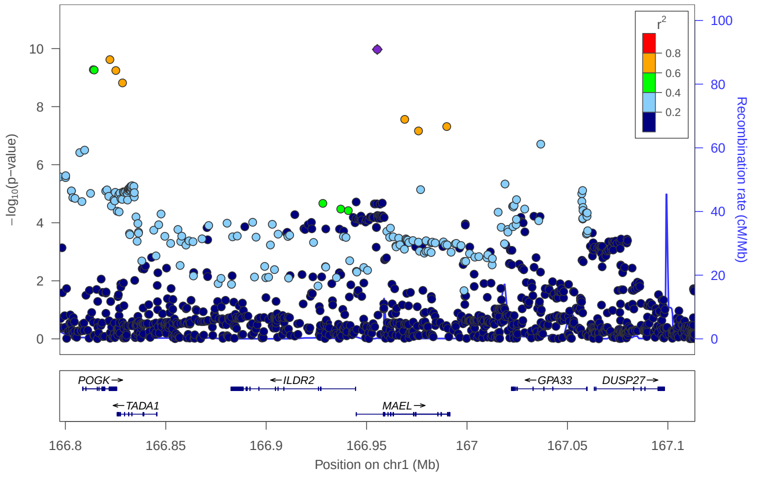


locus 32 rs56247945

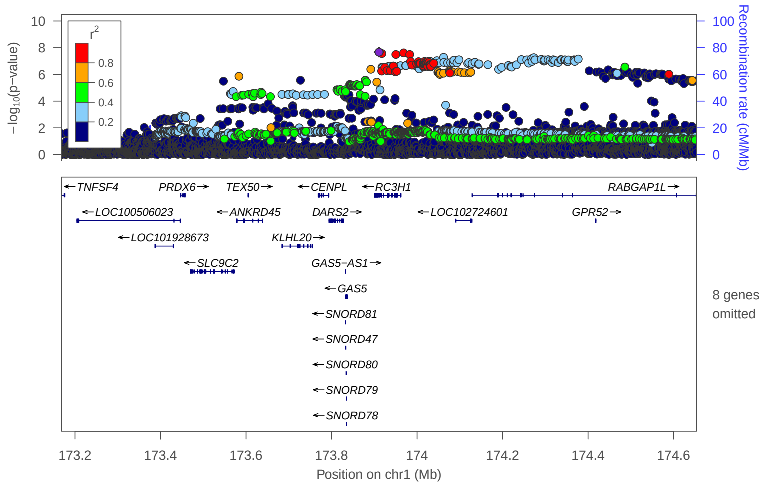

### locus 33 rs509476

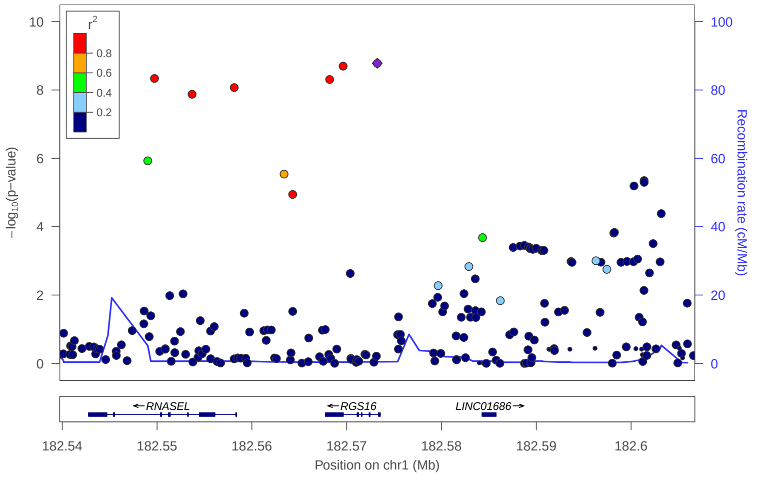


### locus 34 rs2418884

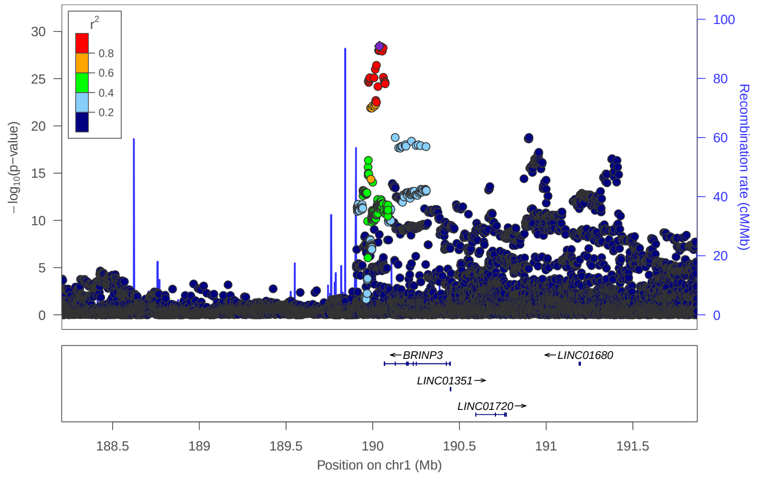


### locus 35 rs5779928

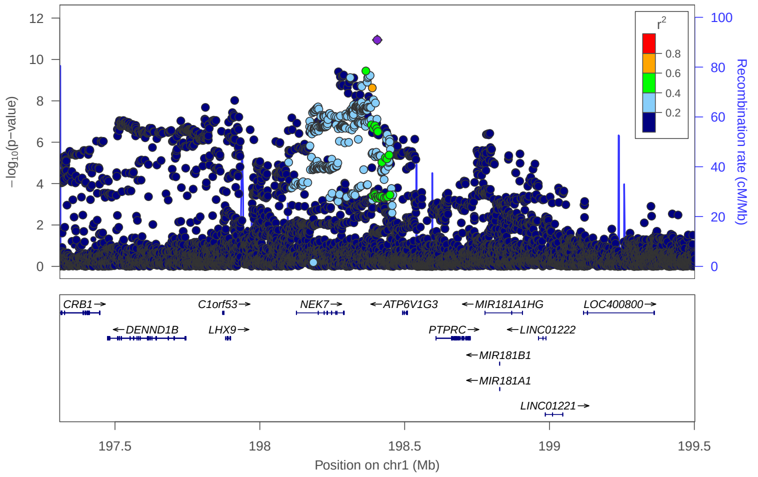


### locus 36 rs2016475

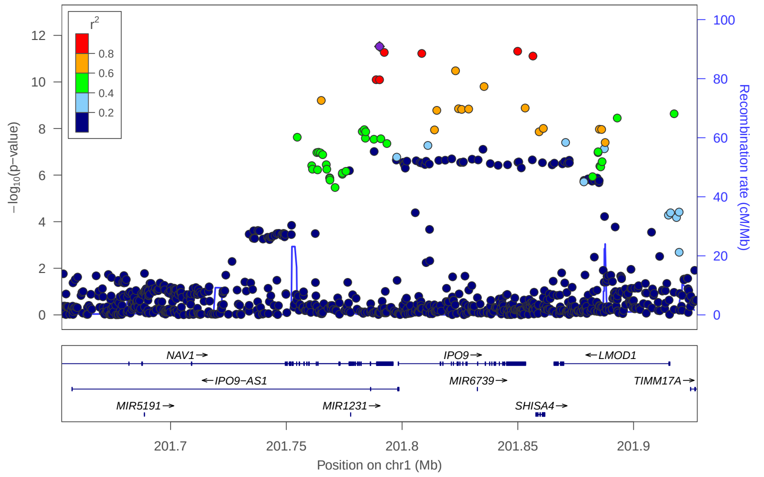


### locus 38 rs58649262

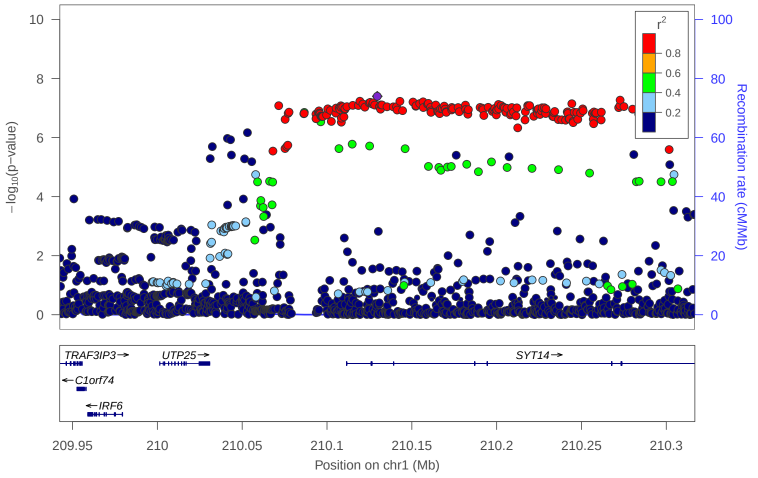


### locus 39 rs55946391

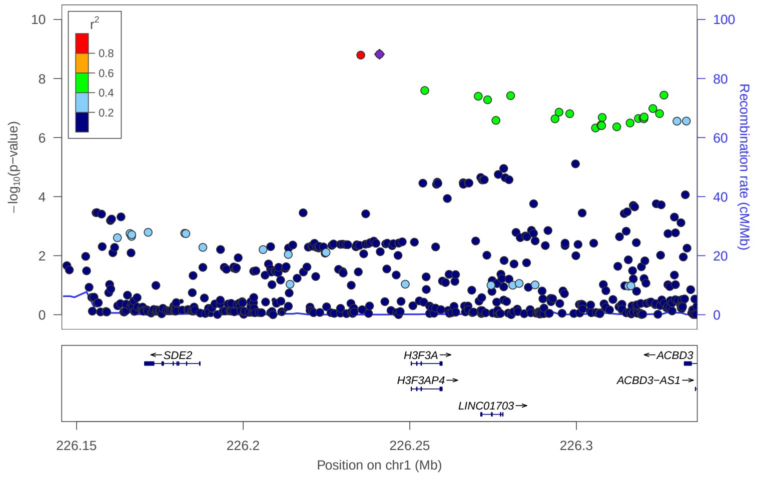


### locus 40 rs4653743

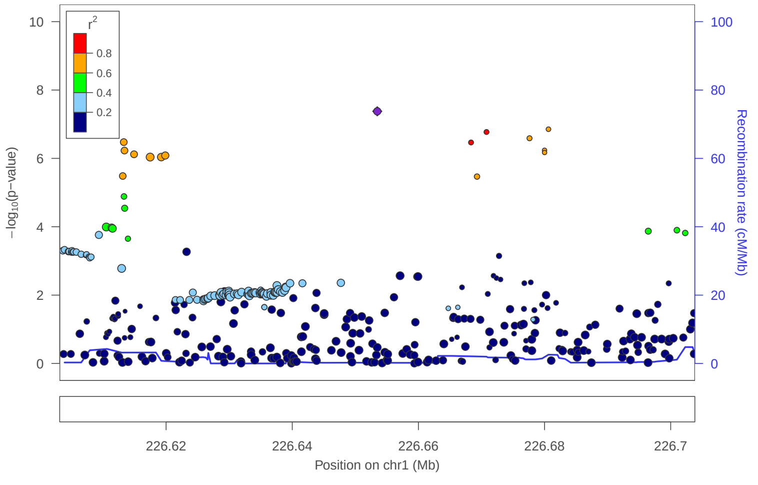


### locus 41 rs11807834

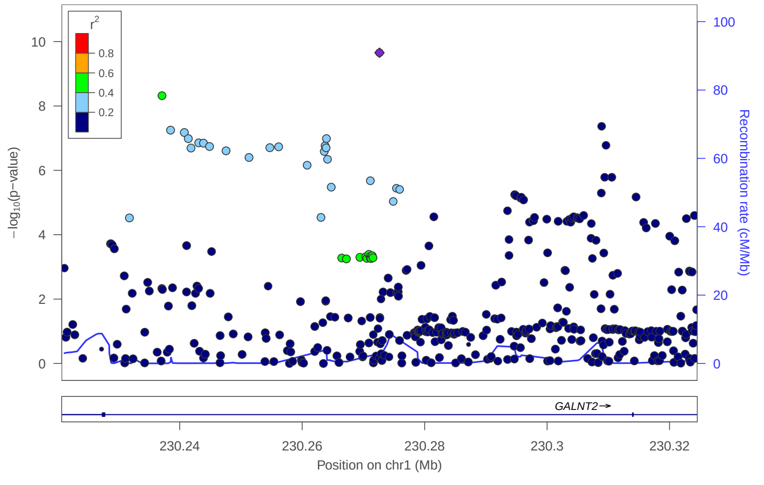

locus 42 rs10927078

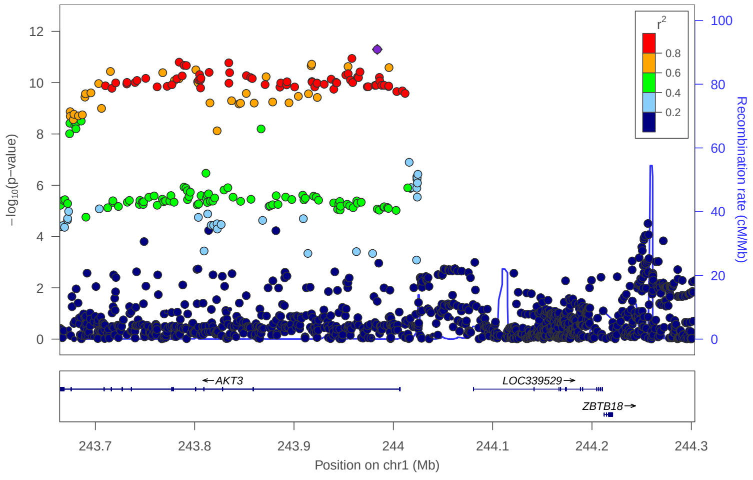


locus 43 rs823248

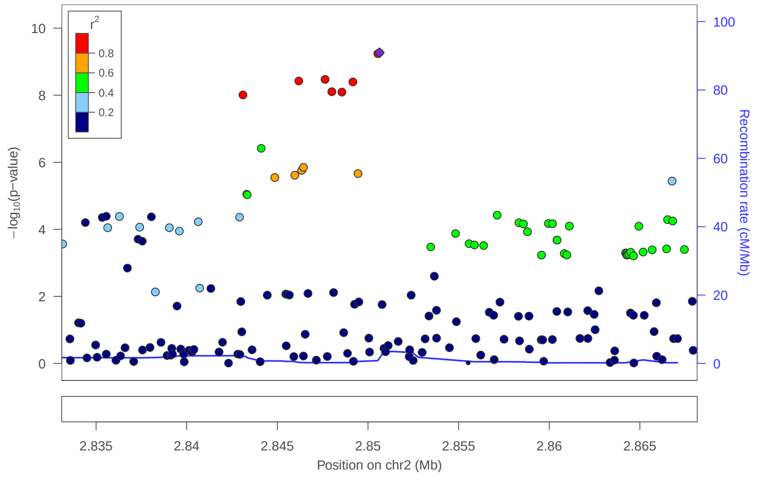


locus 44 rs10185535

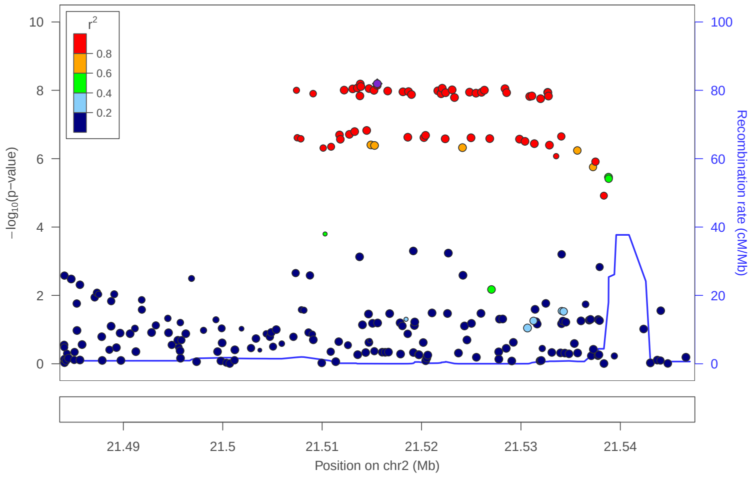


locus 45 rs12617557

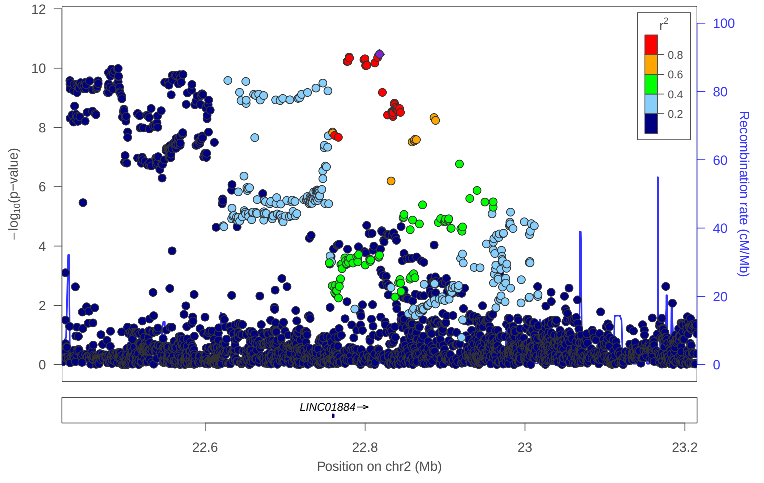


locus 46 rs10204586

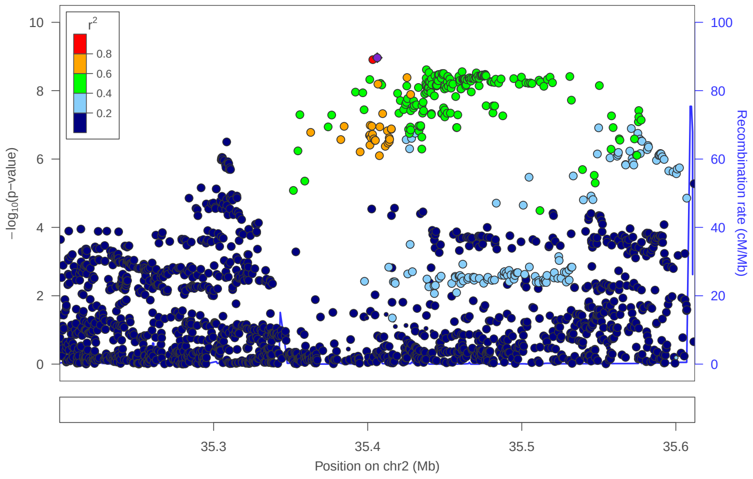


locus 47 rs11903069

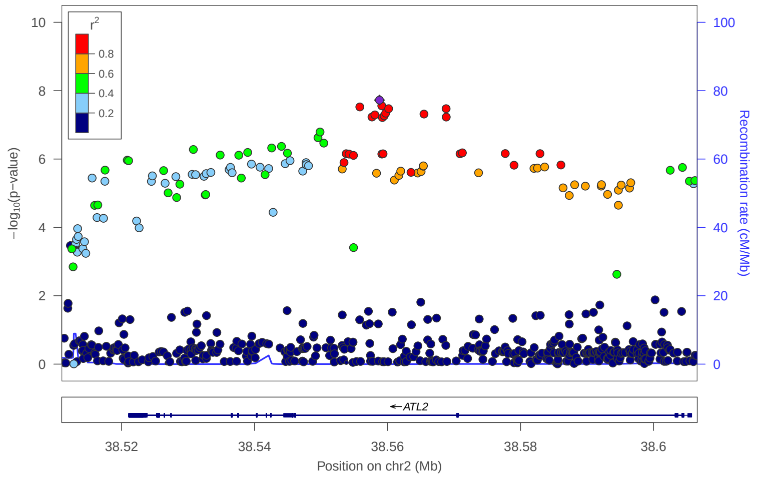


locus 48 rs2374423

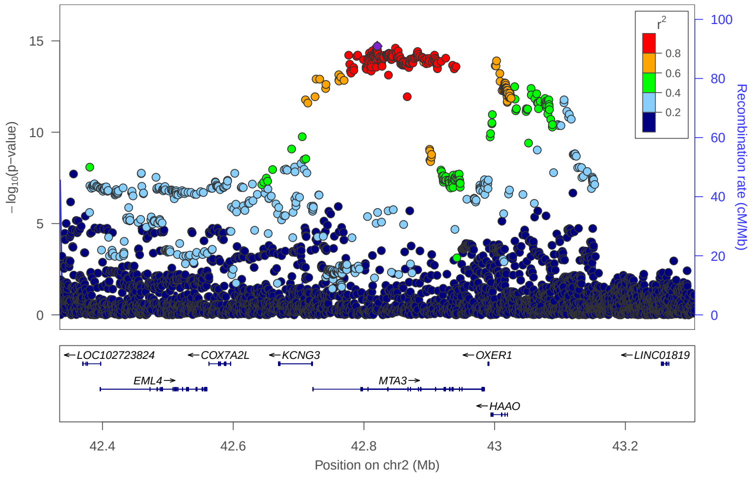


locus 49 rs6720846

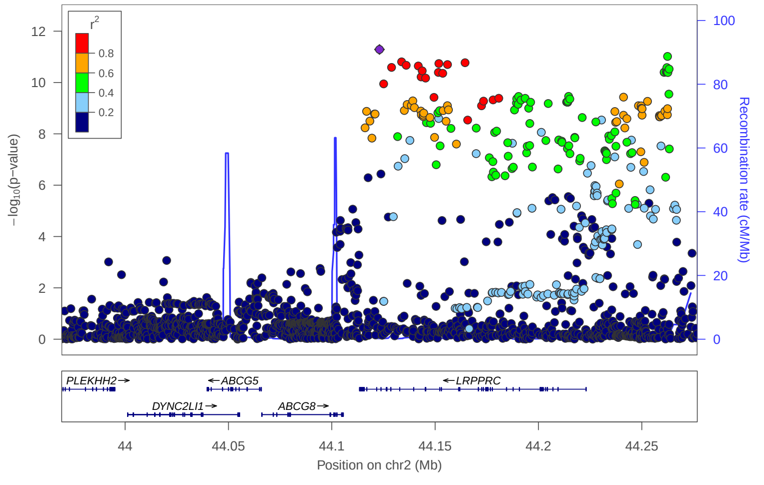

locus 50 rs2881935

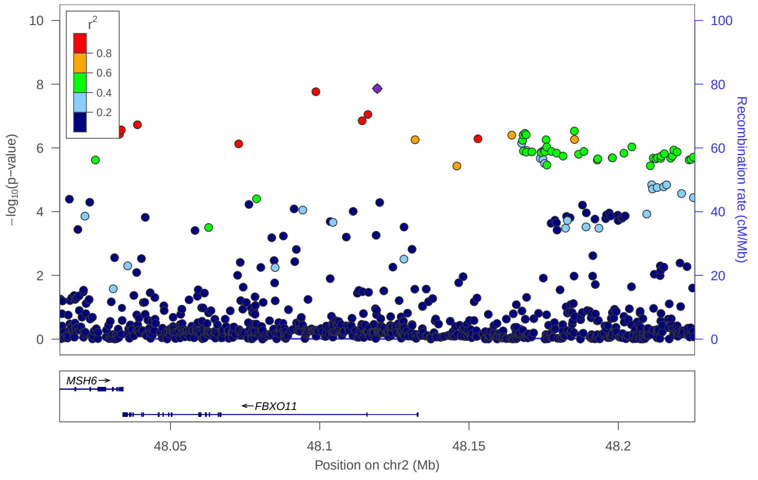


locus 51 rs74431276

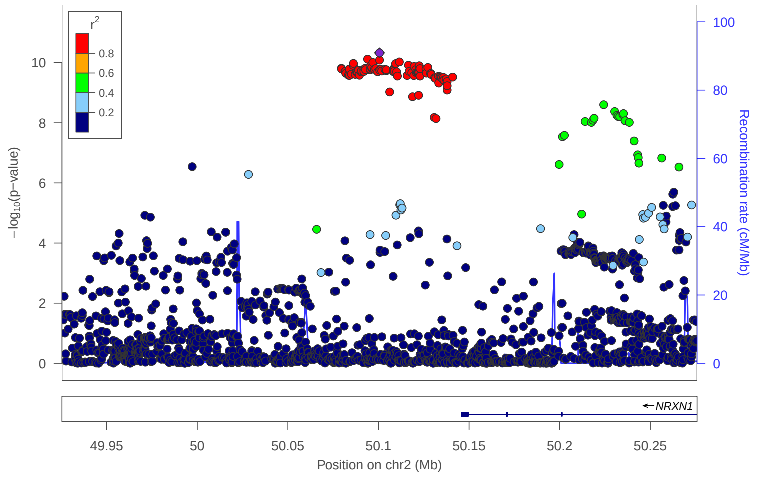


locus 52 rs72807818

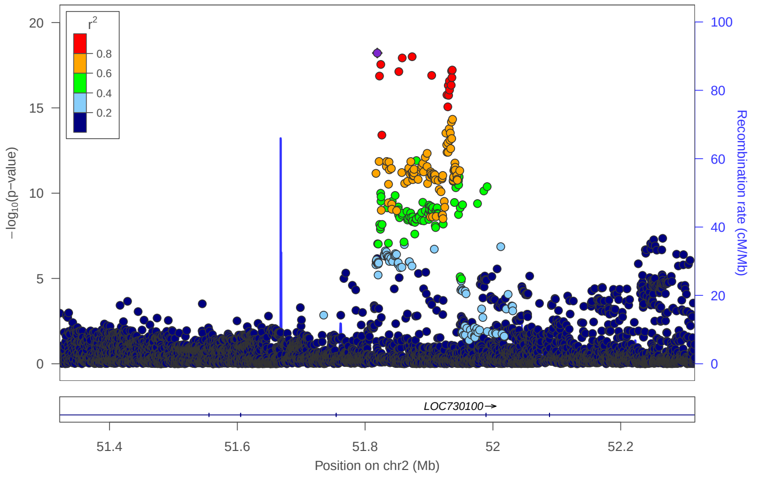


locus 53 rs13404175

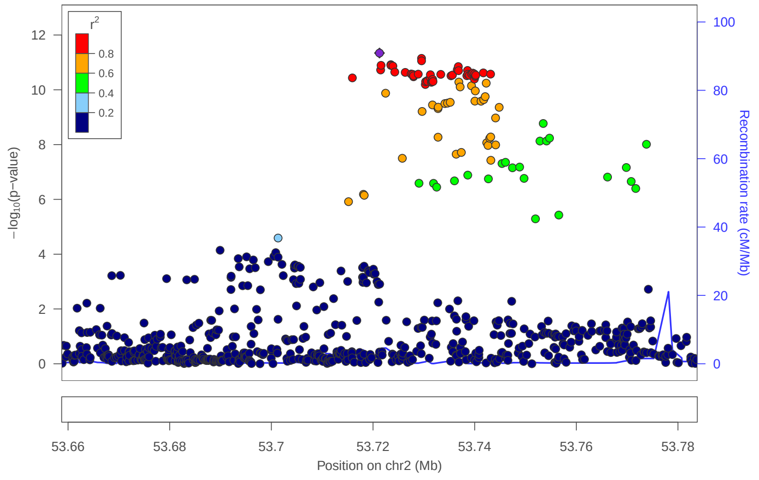


locus 54 rs12468708

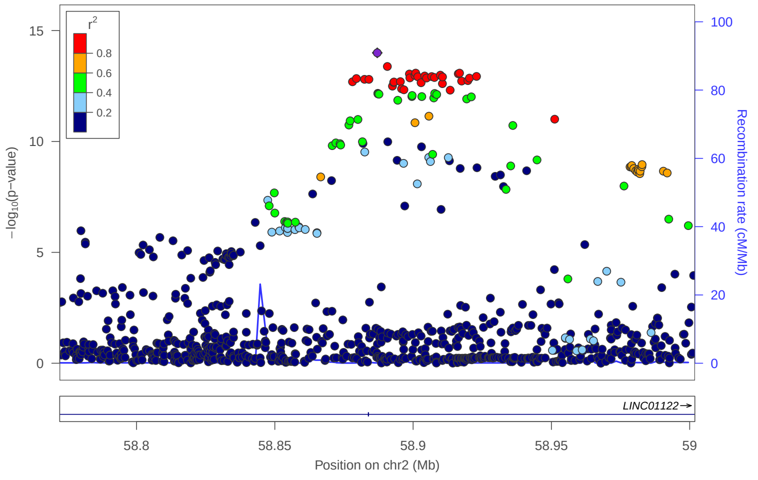


locus 55 rs359247

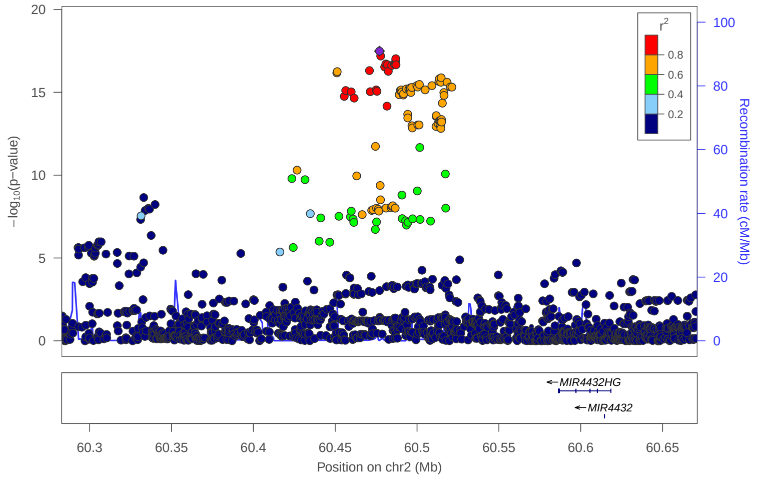


locus 56 rs13015703

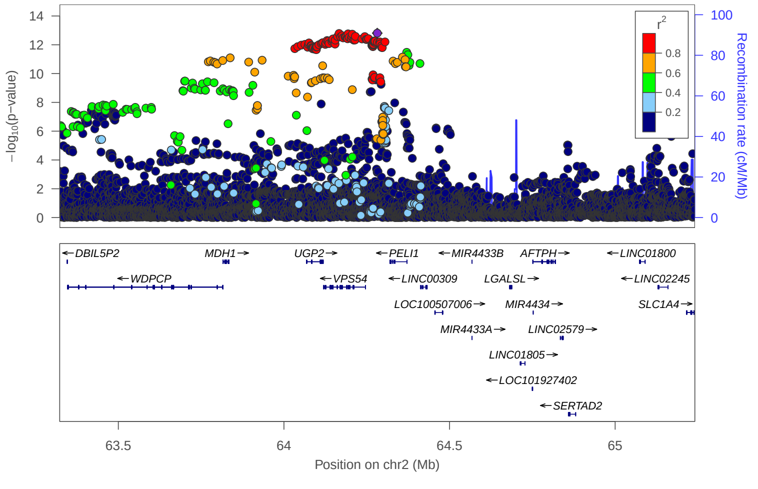


locus 57 rs62144053

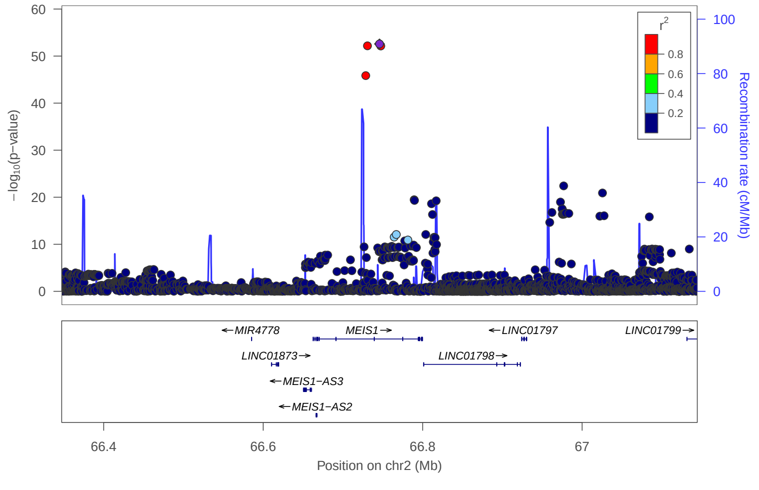

locus 58 rs2861752

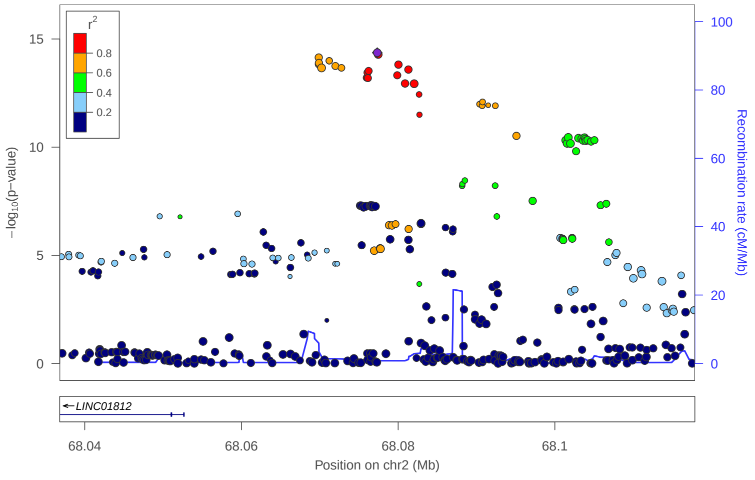


locus 59 rs62149809

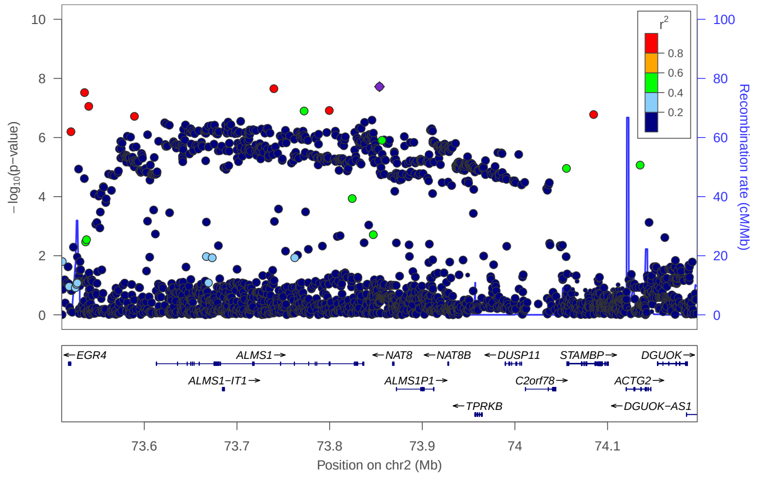


locus 60 rs4538233

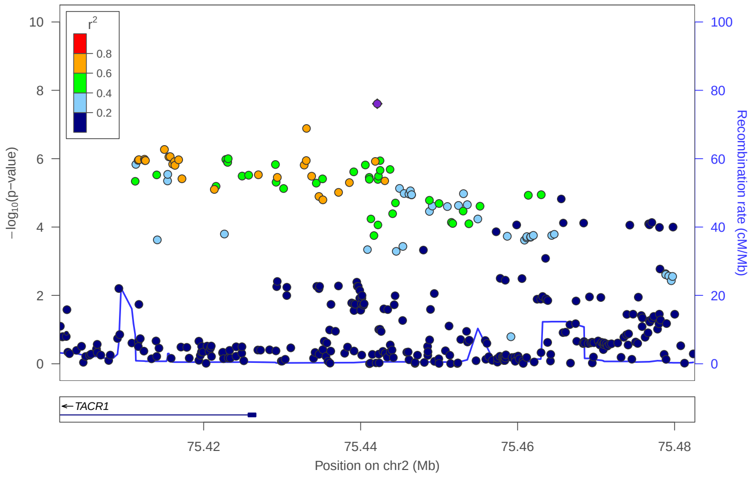


locus 61 rs6761838

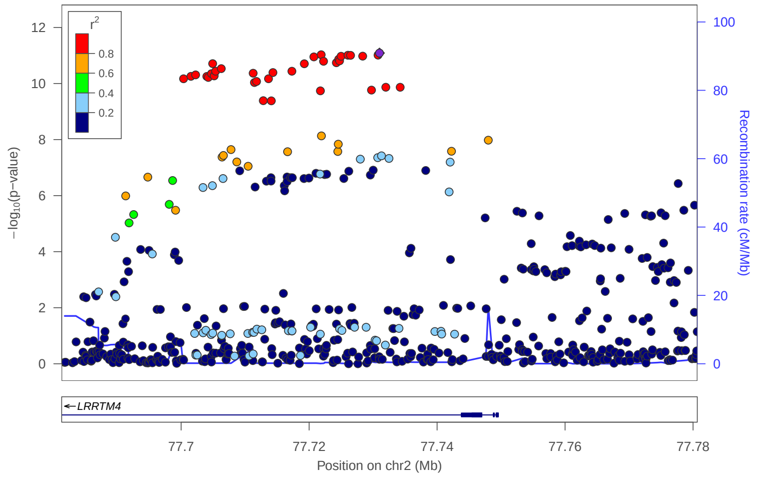


locus 62 rs6724539

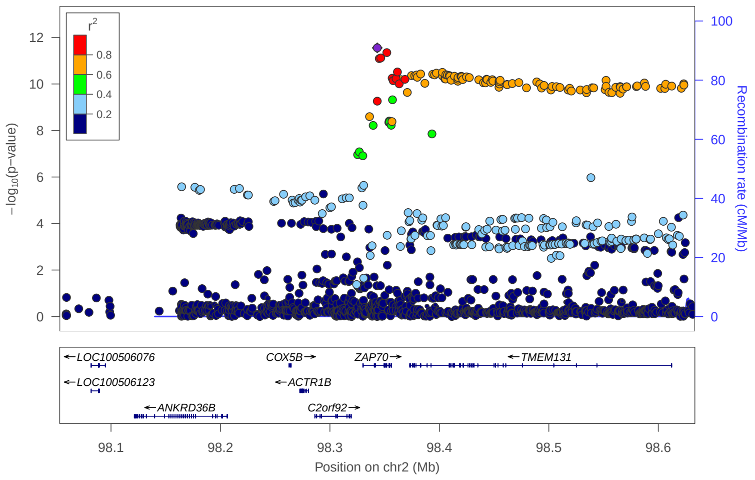


locus 63 rs6735012

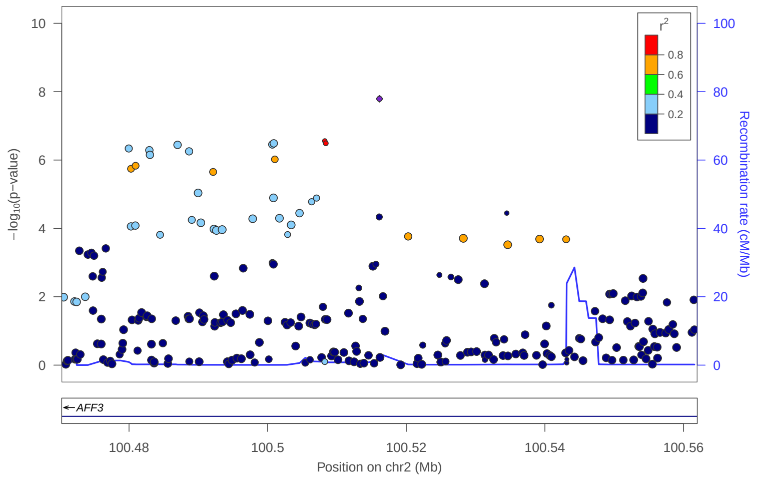


locus 64 rs4402784

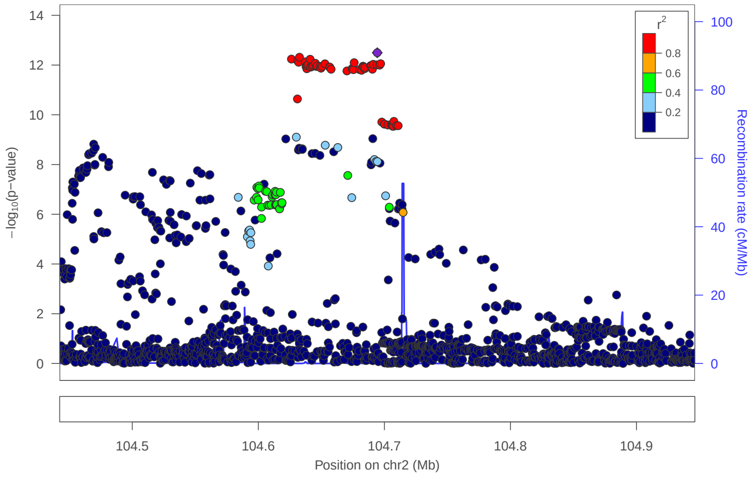


locus 65 rs62158170

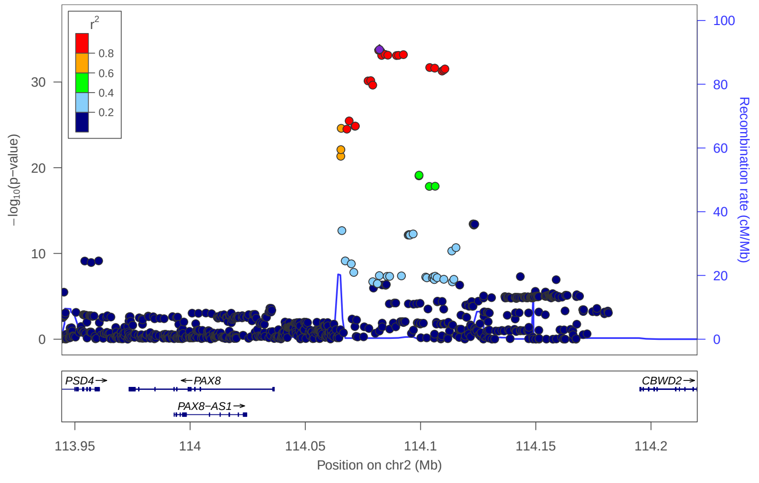

locus 66 rs2432721

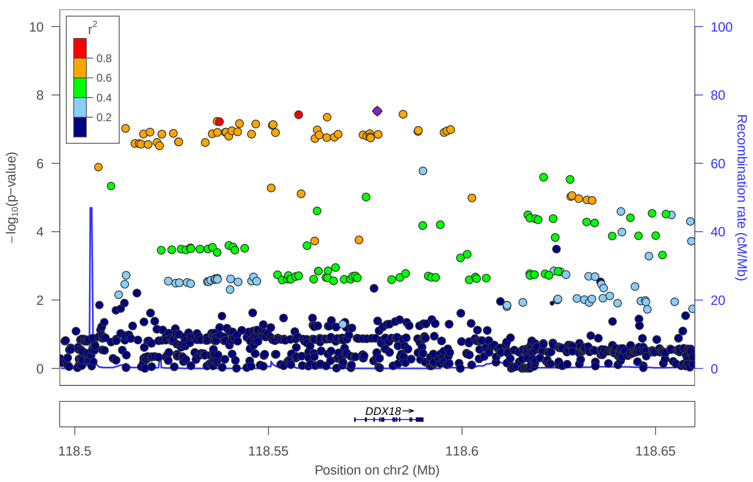


locus 67 rs4848524

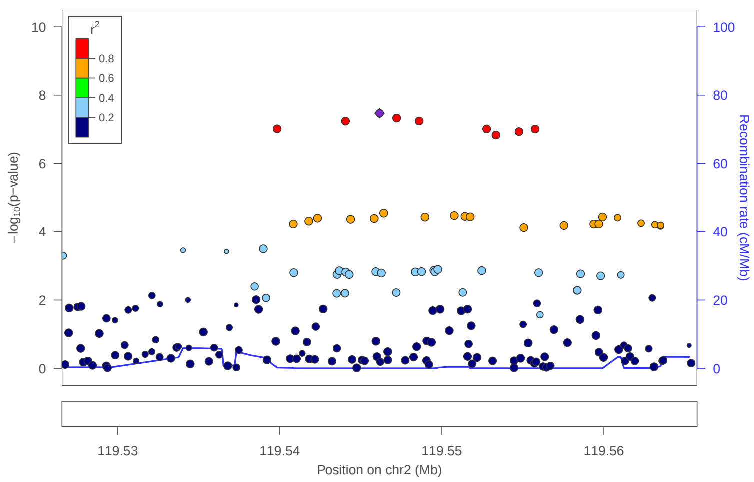


locus 68 rs12618343

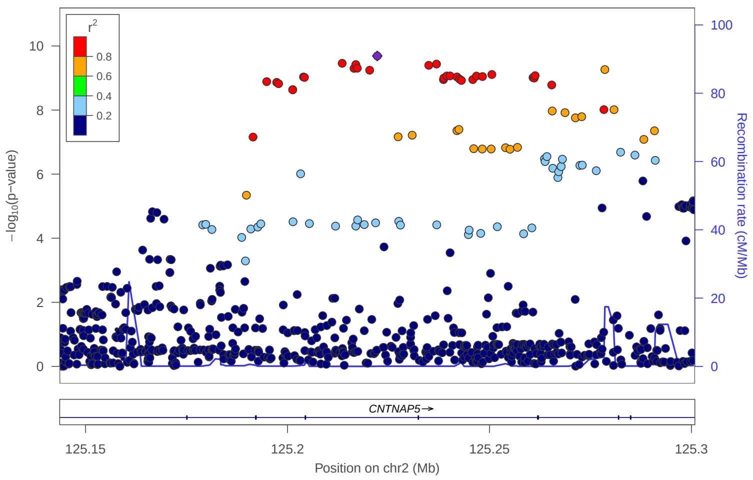


locus 69 rs773875123

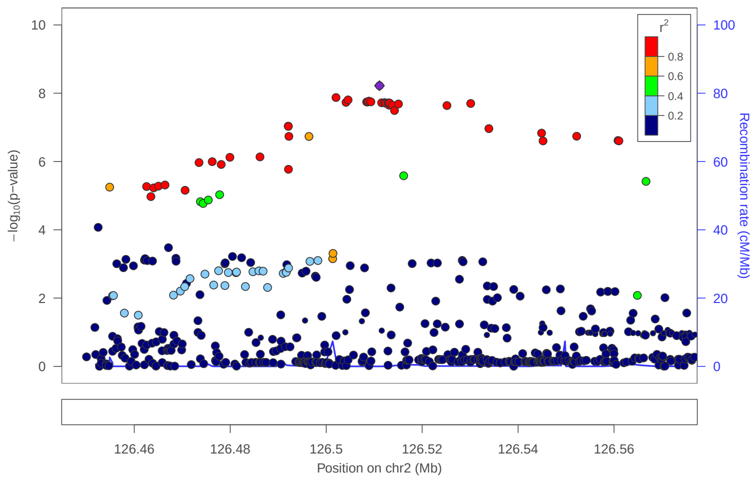


locus 70 rs13415724

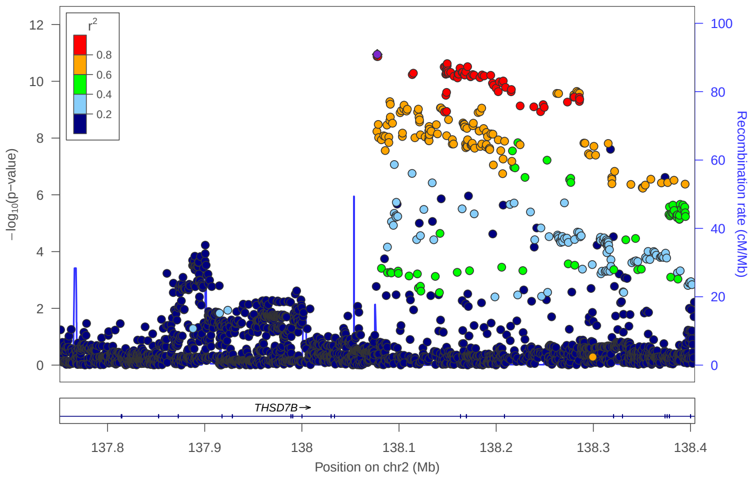


locus 71 rs78154941

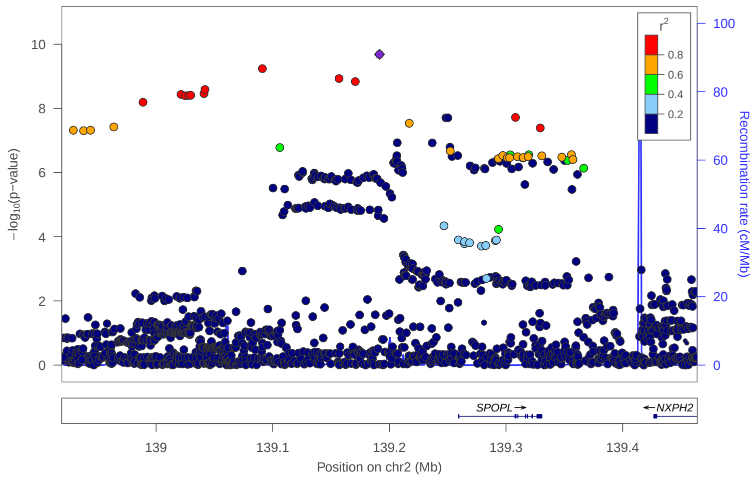


locus 72 rs1437334

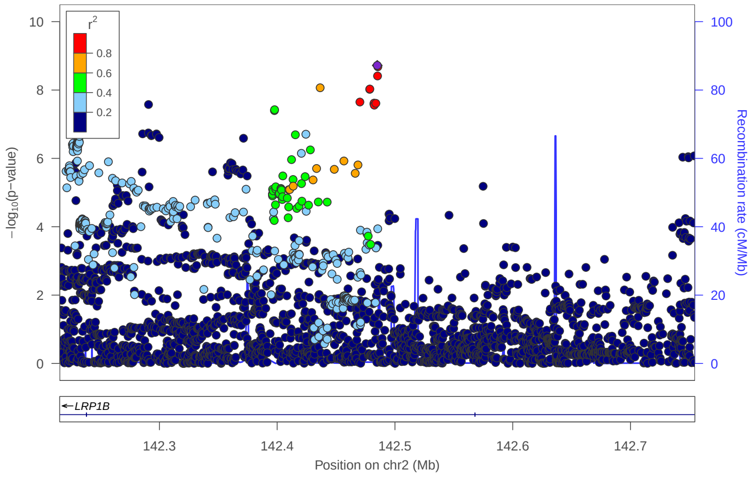


locus 73 rs13001483

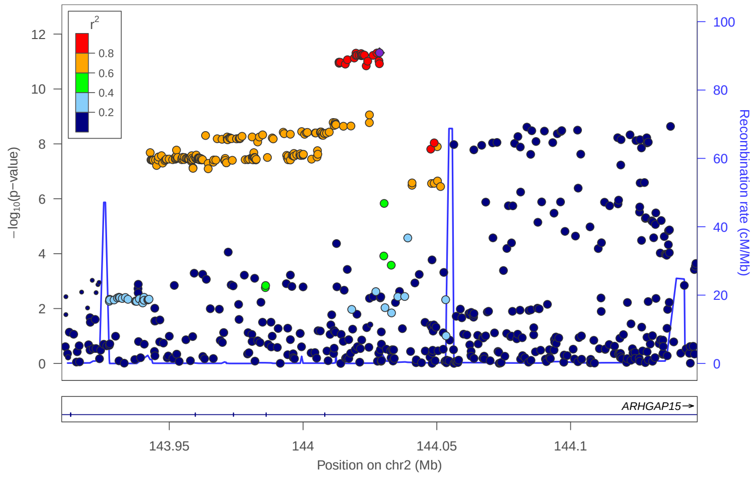

### locus 74 rs7607770

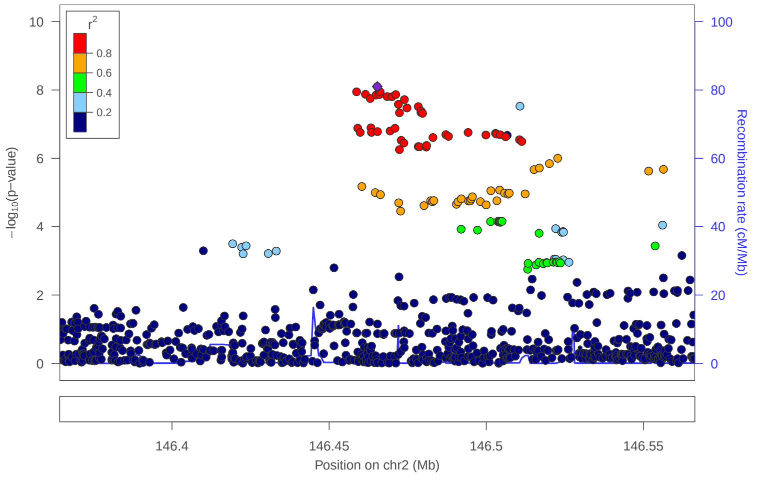


### locus 75 rs62167788

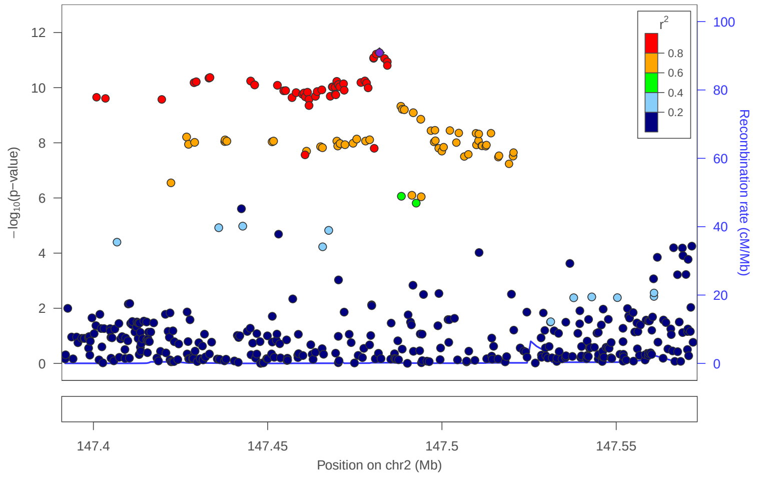


### locus 76 2:148658595:A_G

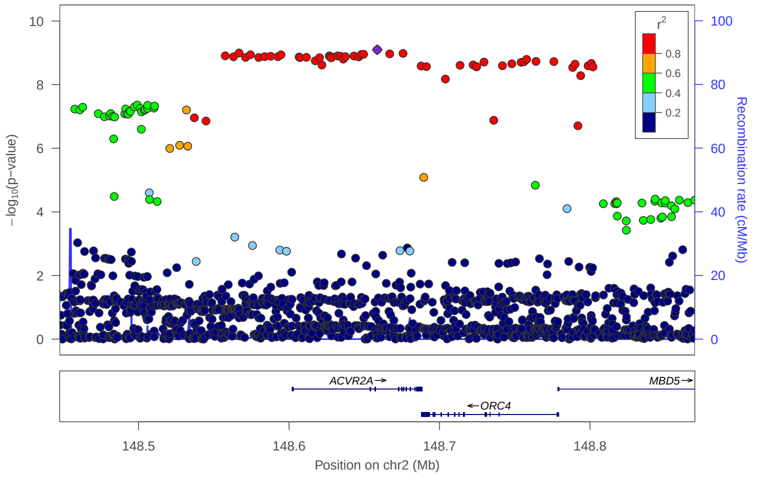


### locus 77 rs77276698

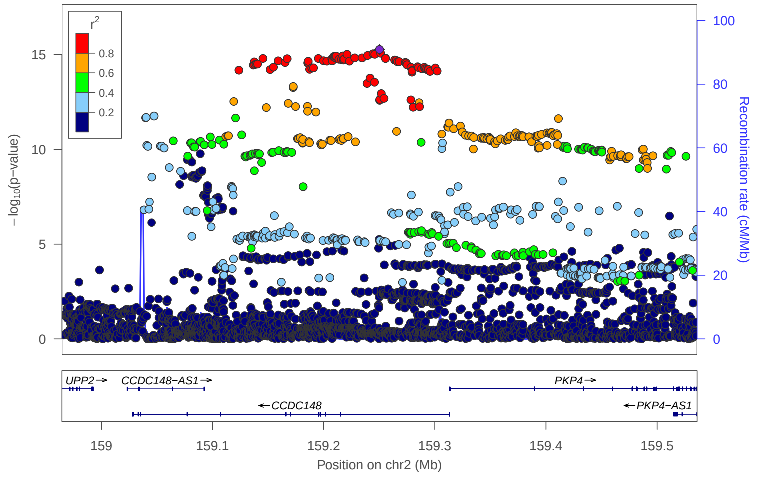


### locus 78 rs1549577

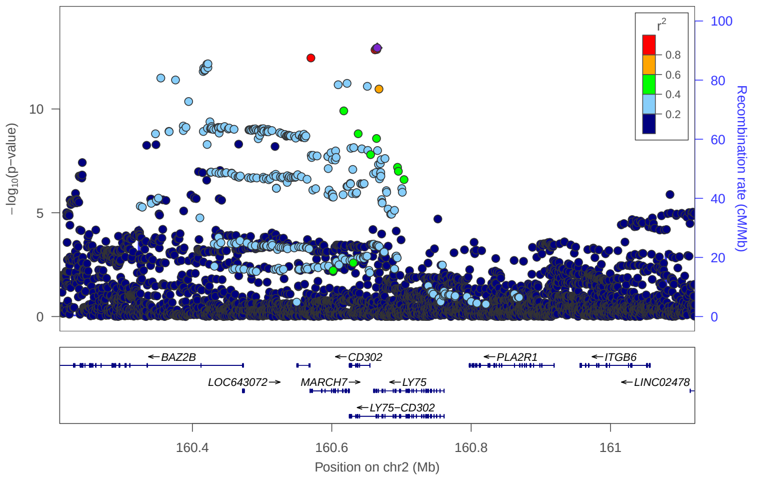


### locus 79 rs1267070

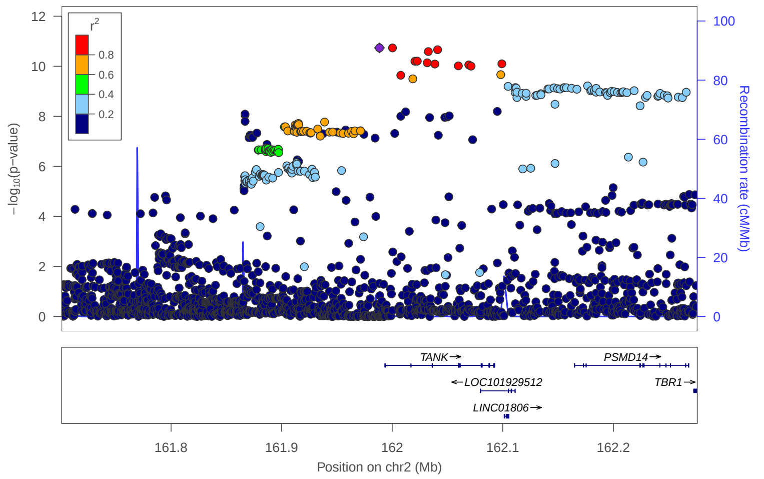


### locus 80 rs11683458

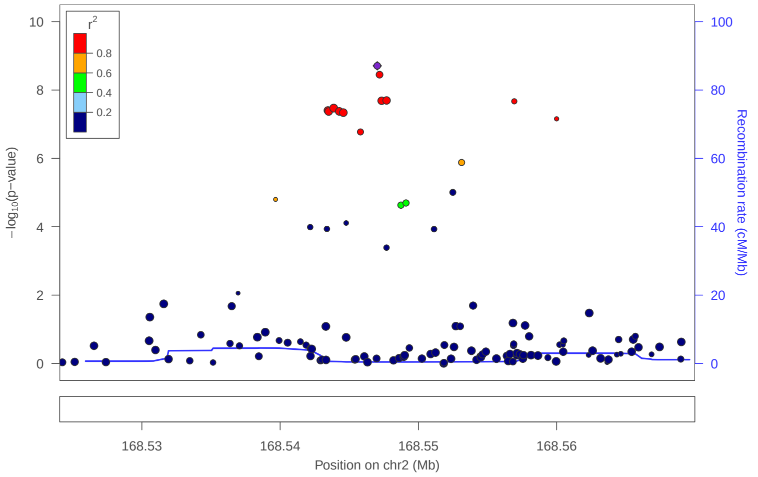


### locus 81 rs10930502

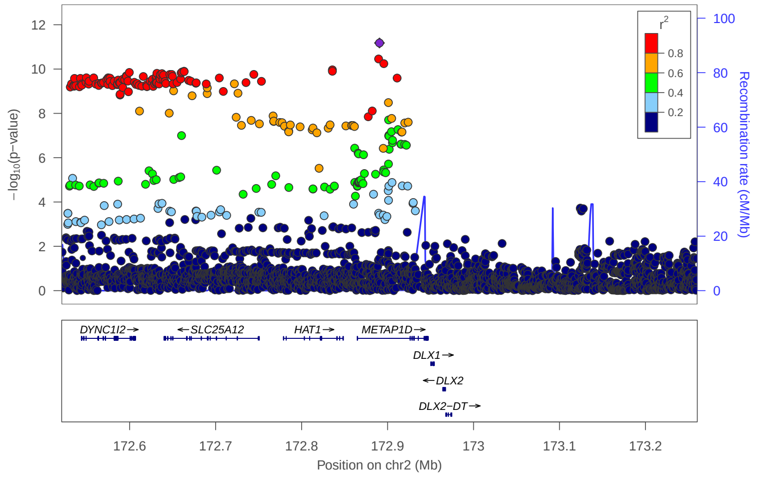

locus 82 rs836603

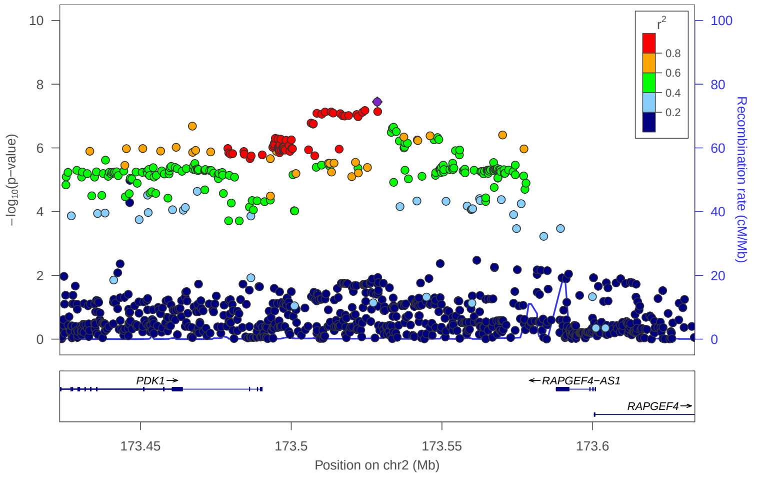


locus 83 rs2884836

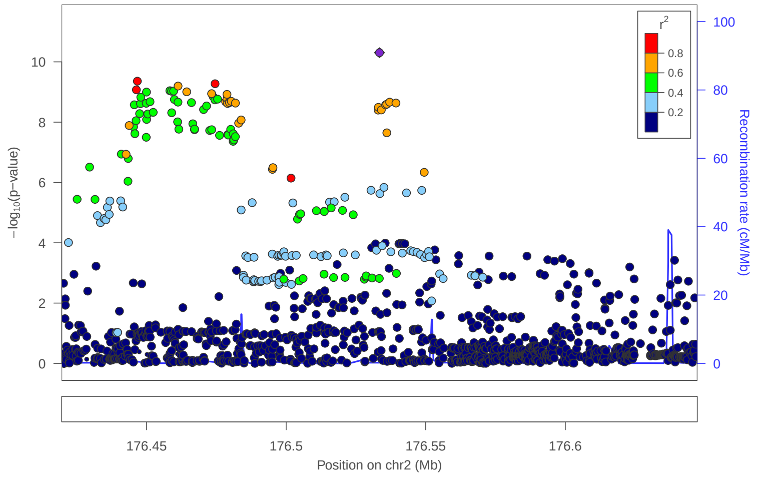


locus 84 rs2123641

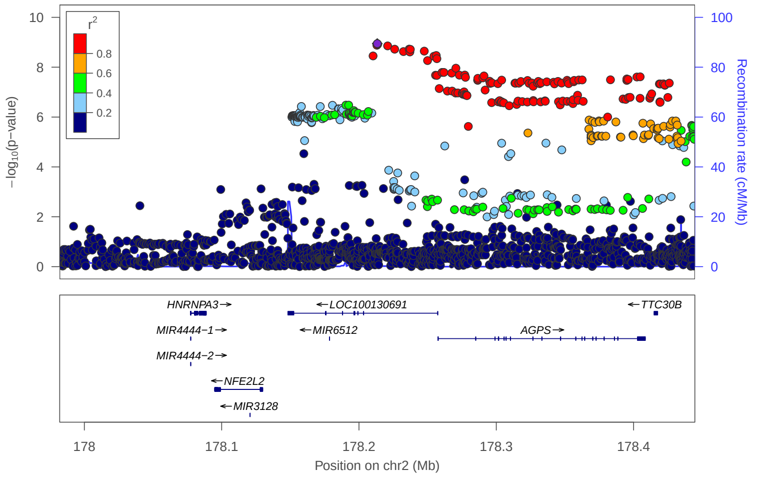


locus 85 rs10497655

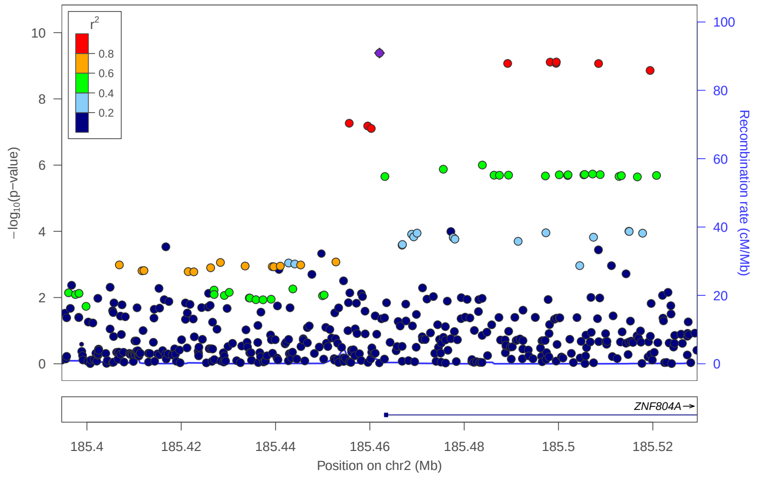


locus 86 rs1064213

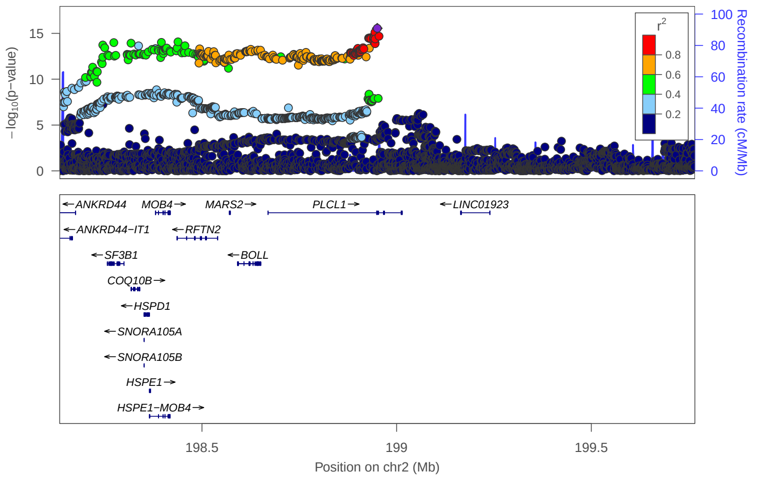


locus 87 rs60431238

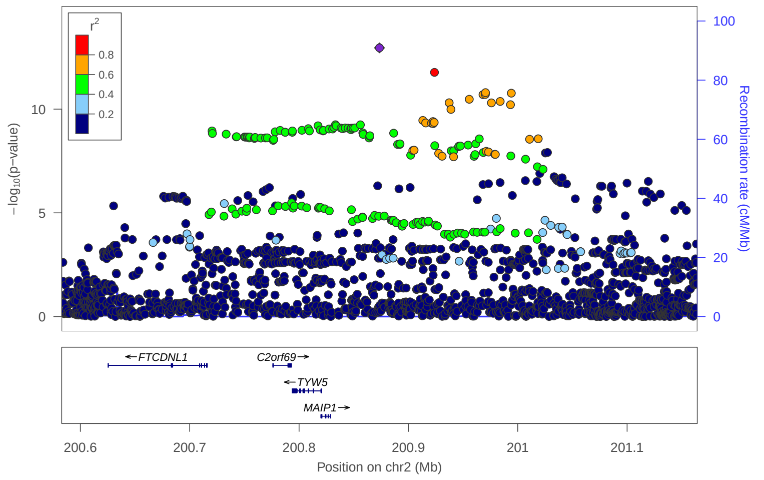


locus 88 rs145664840

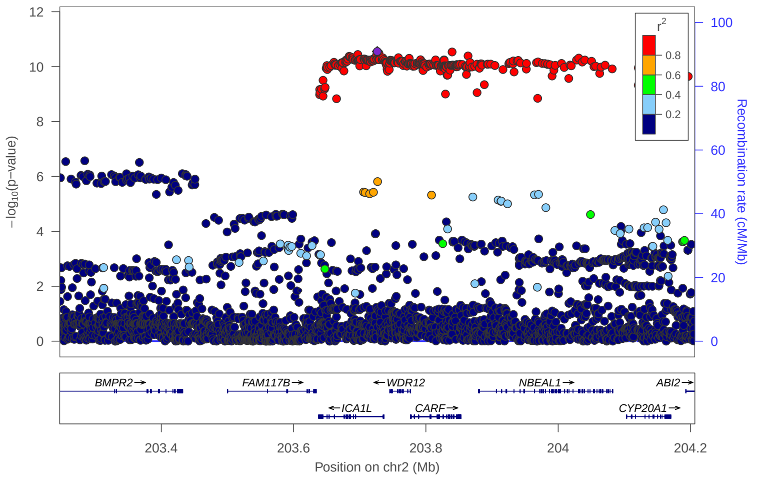


locus 89 rs10187826

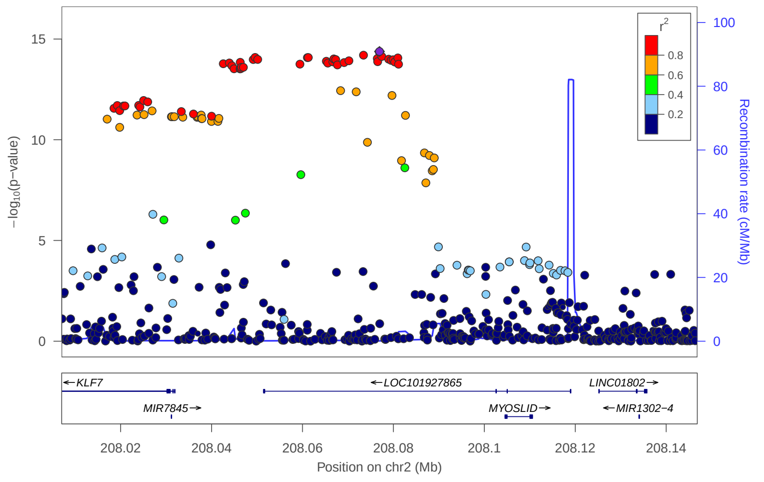

locus 90 rs62184481

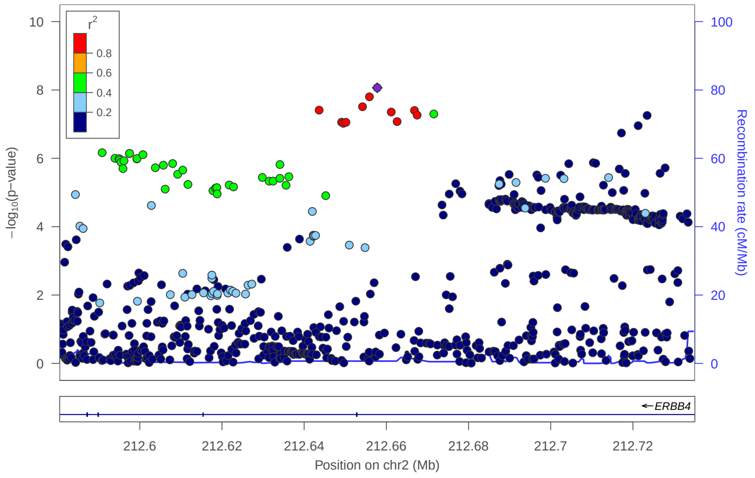


locus 91 rs34967082

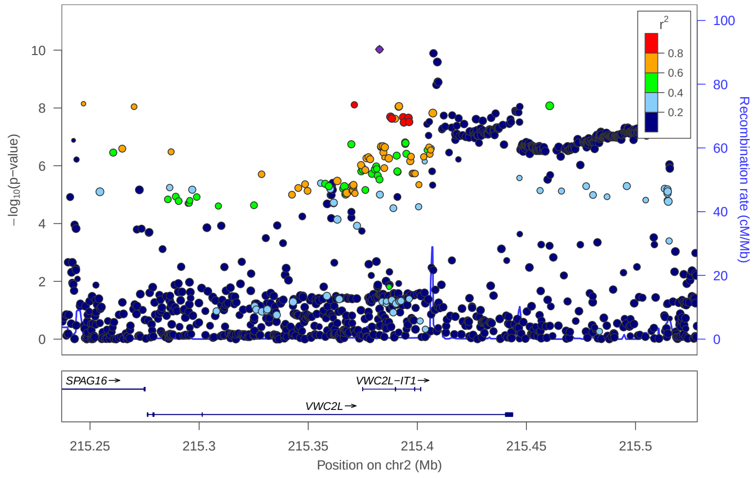


locus 92 rs62178707

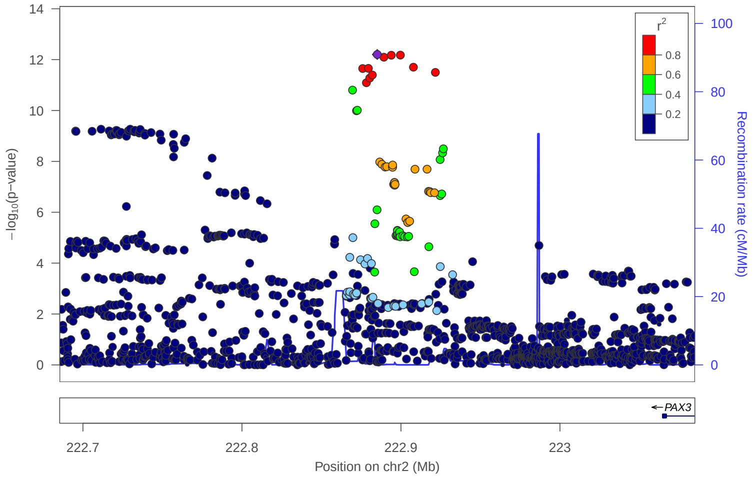


locus 93 rs11897824

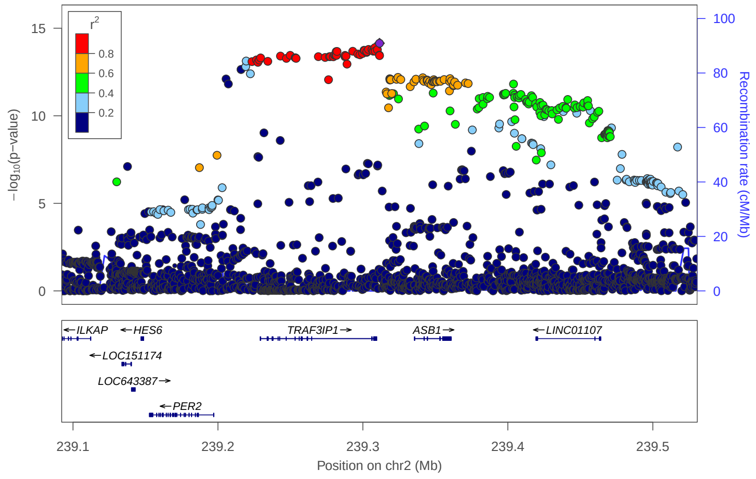


locus 94 rs4455151

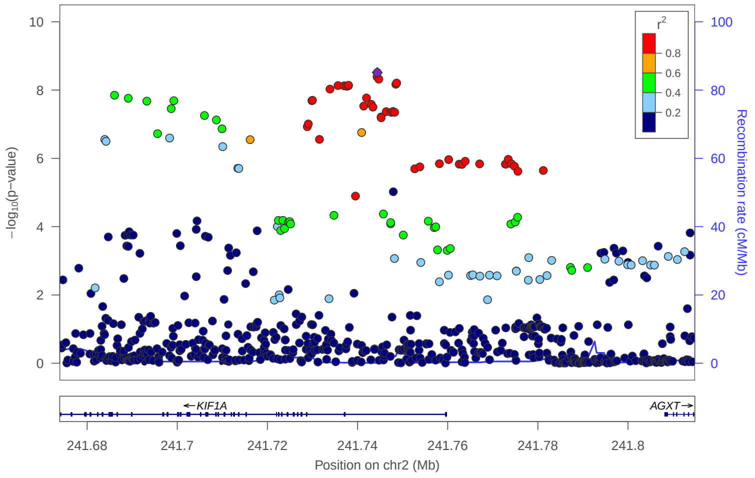


locus 95 rs2729303

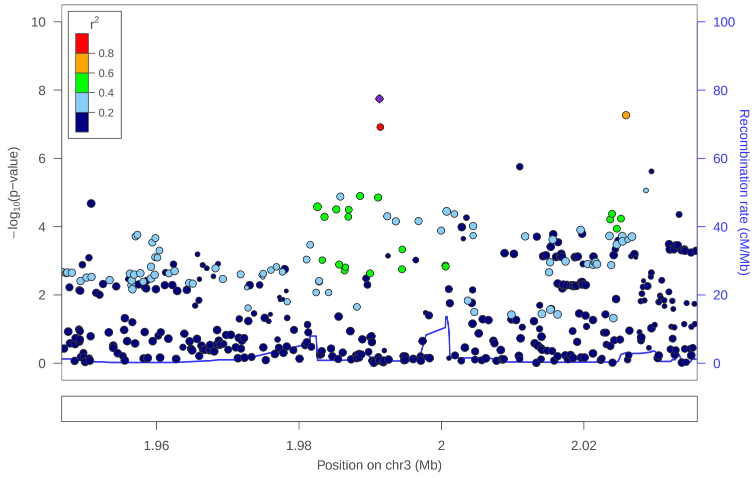


locus 96 rs76901635

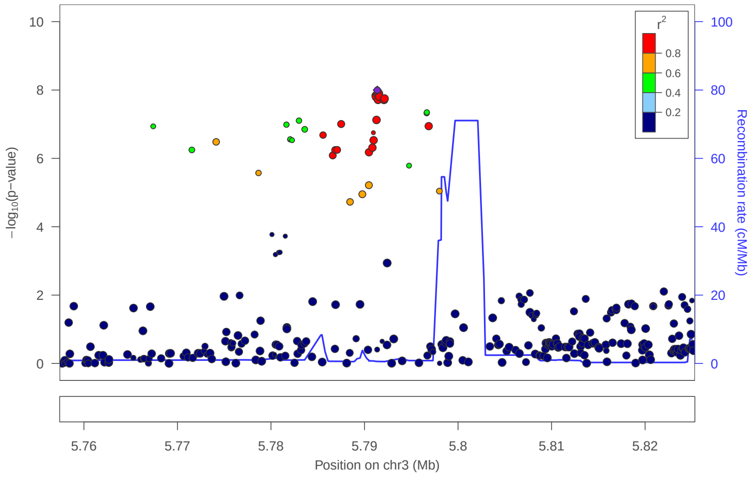


locus 97 rs78838624

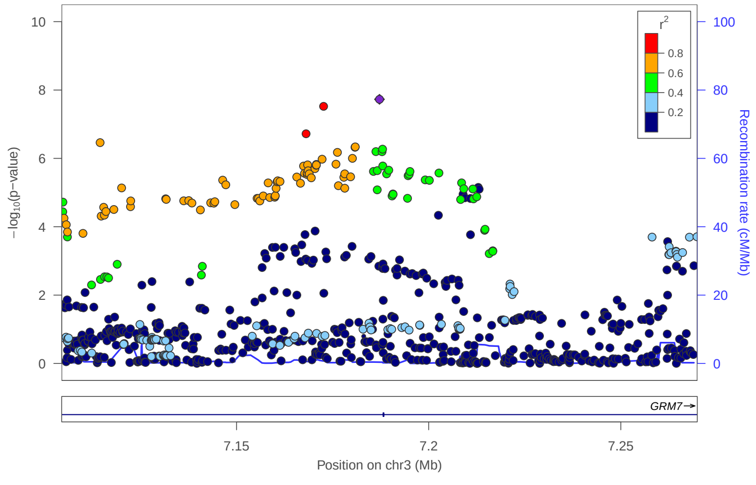

### locus 98 rs6774026

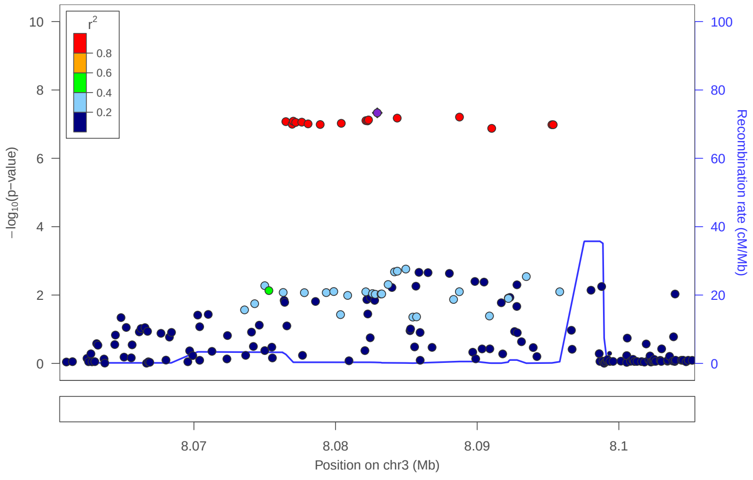


### locus 99 rs4383496

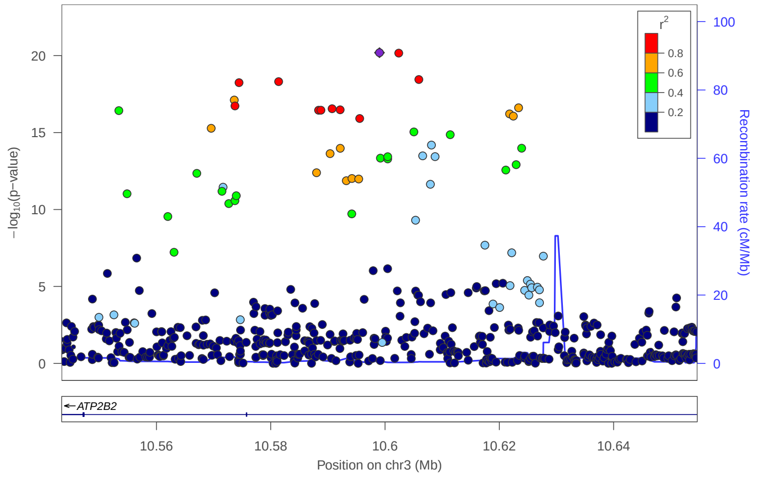


### locus 100 rs9653970

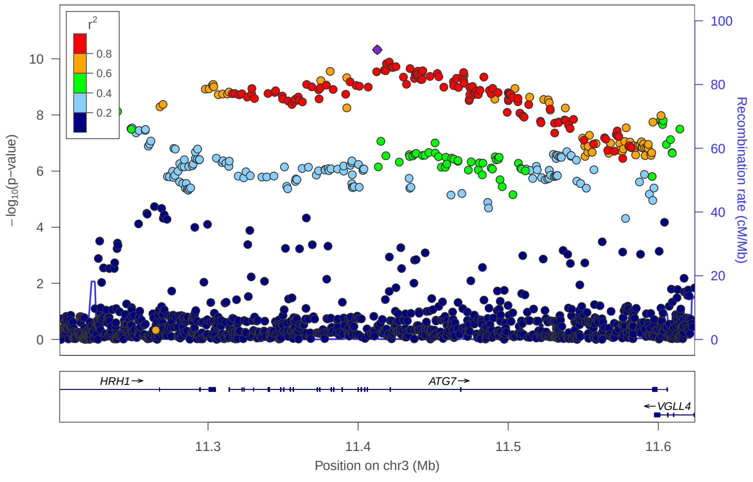


### locus 101 rs9838564

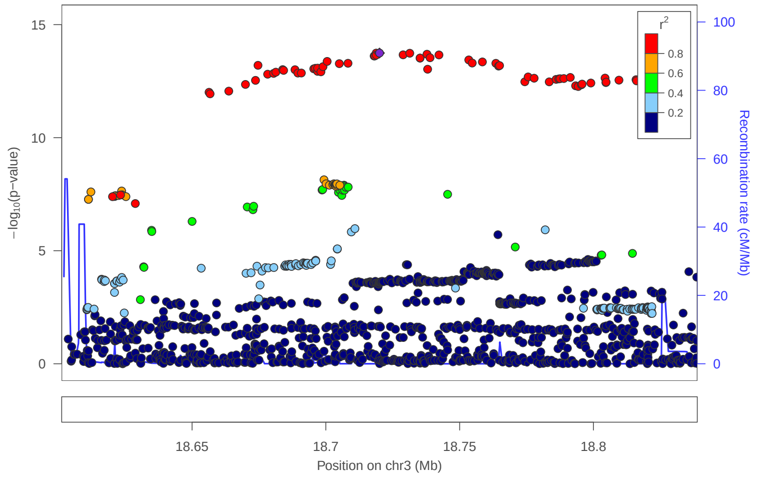


### locus 102 rs4858241

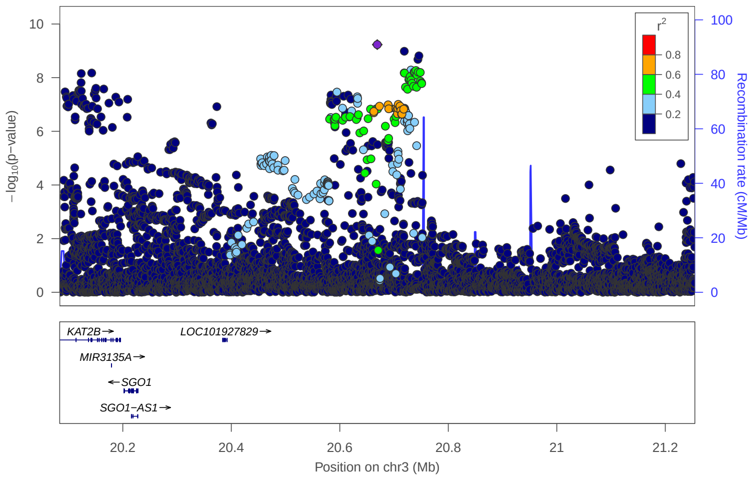


### locus 103 rs1603985

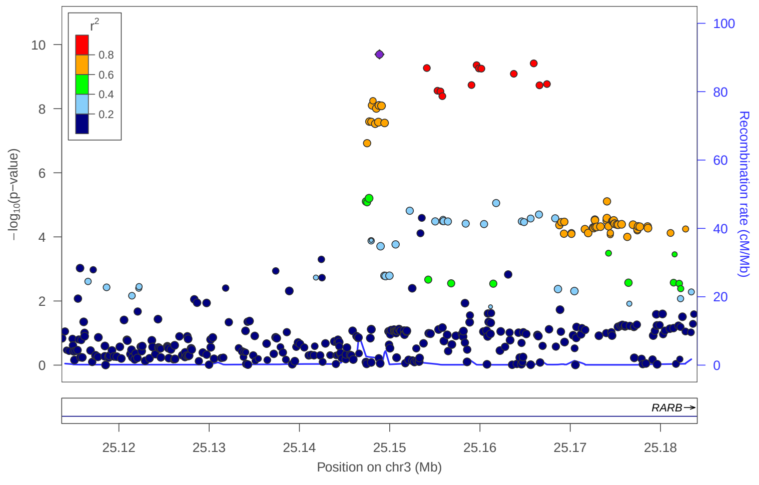


### locus 104 rs2122231

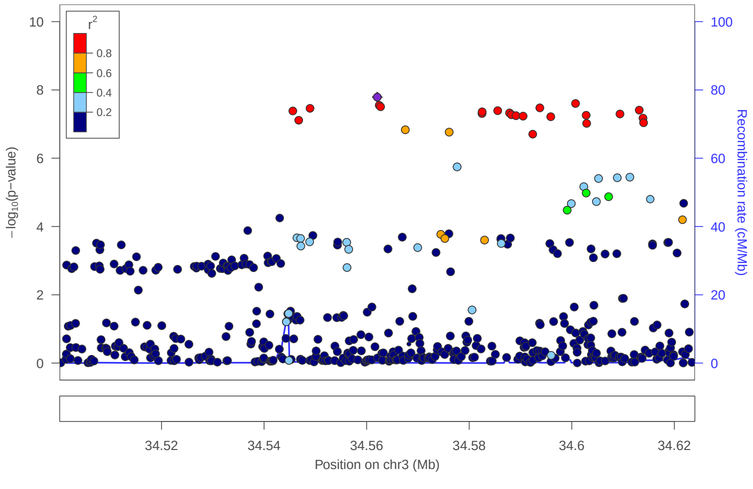


### locus 105 rs9875582

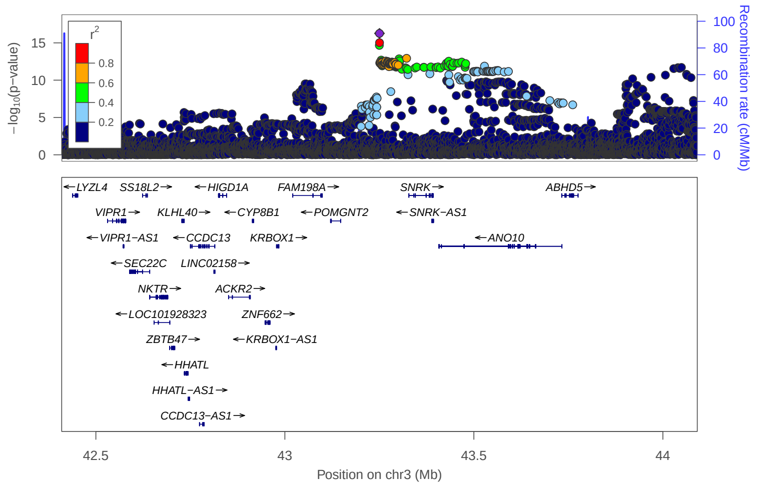

locus 106 rs3755831

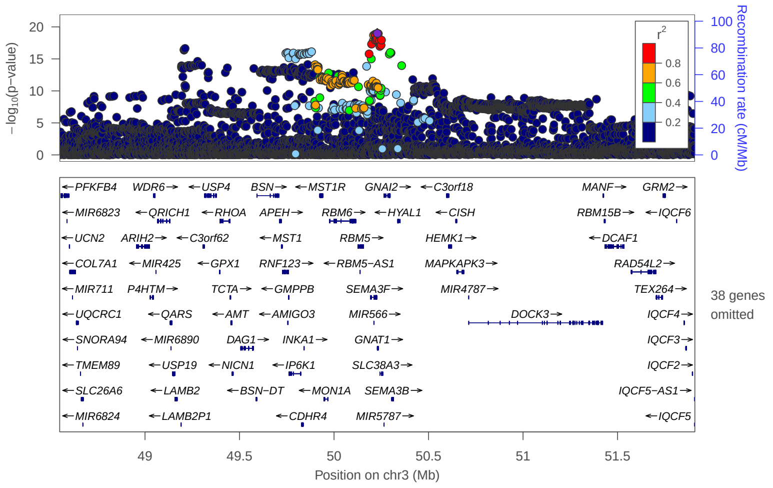


locus 107 rs62252339

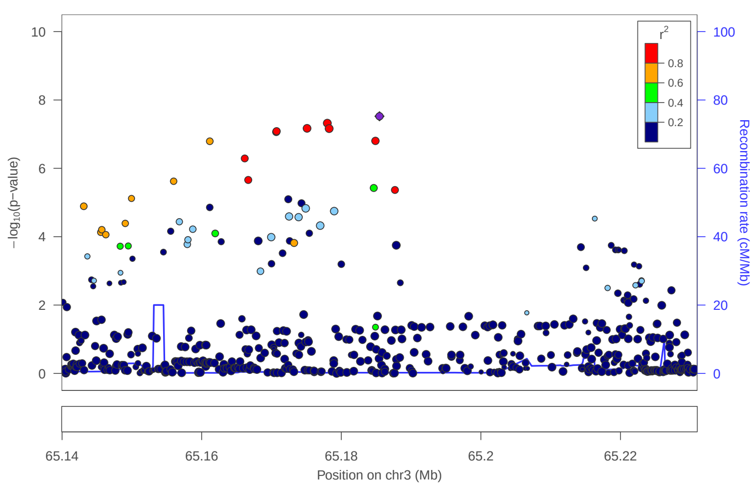


locus 108 rs6762926

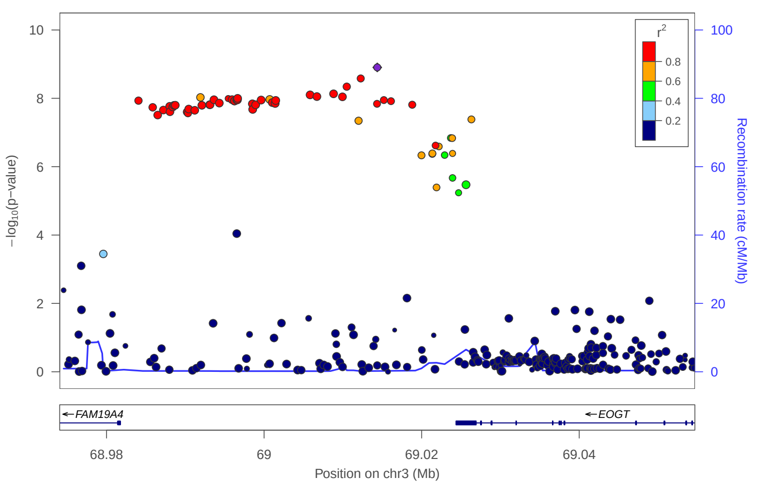


locus 109 rs67588220

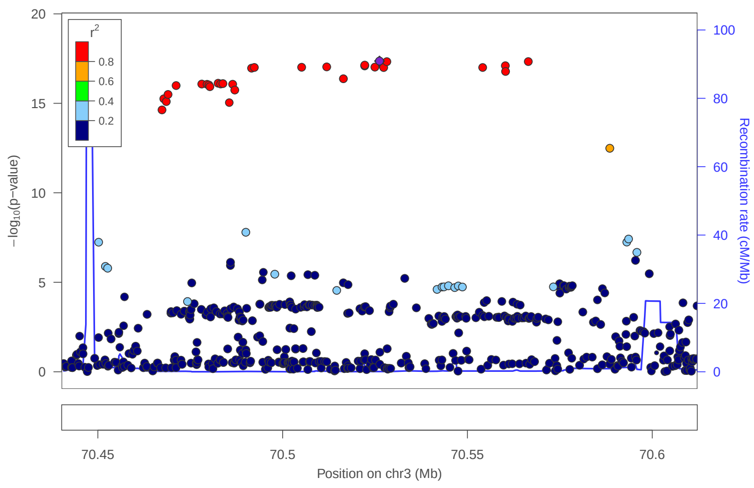


locus 110 rs115433278

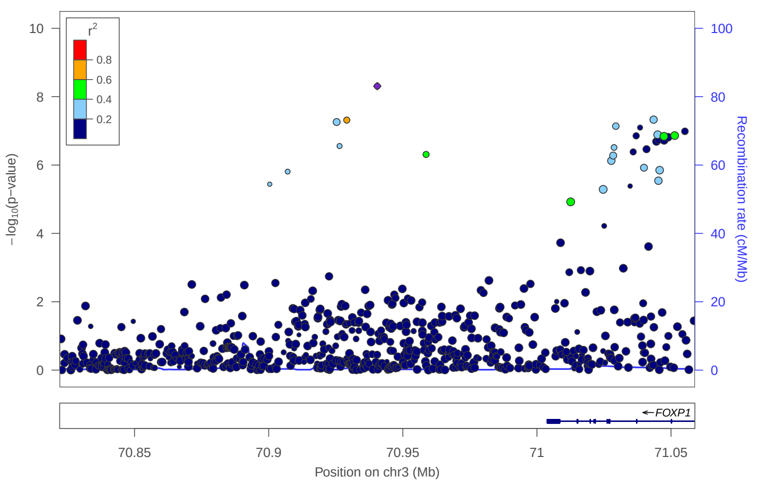


locus 111 rs1864896

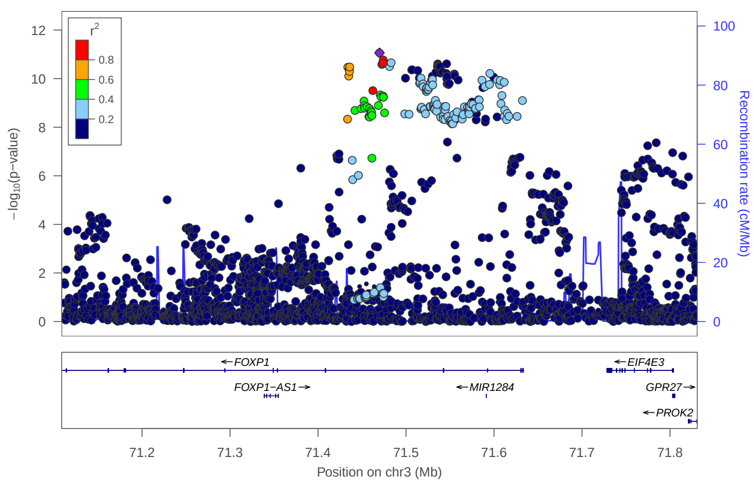


locus 112 rs113757655

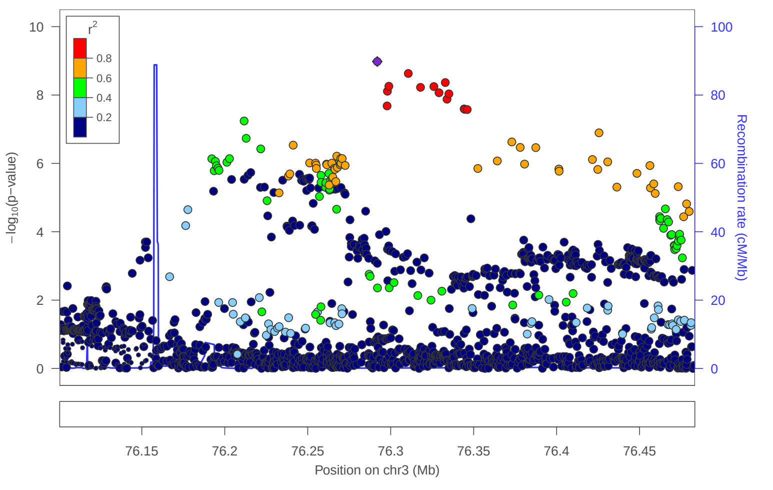


locus 113 rs11425226

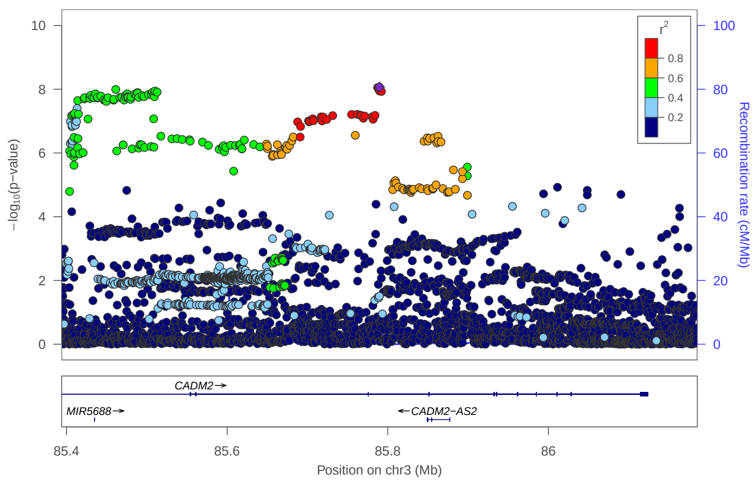

locus 114 rs13093657

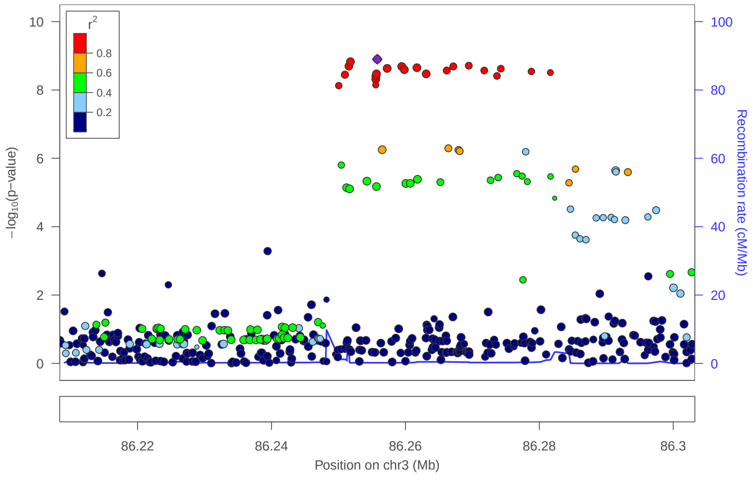


locus 115 rs147012818

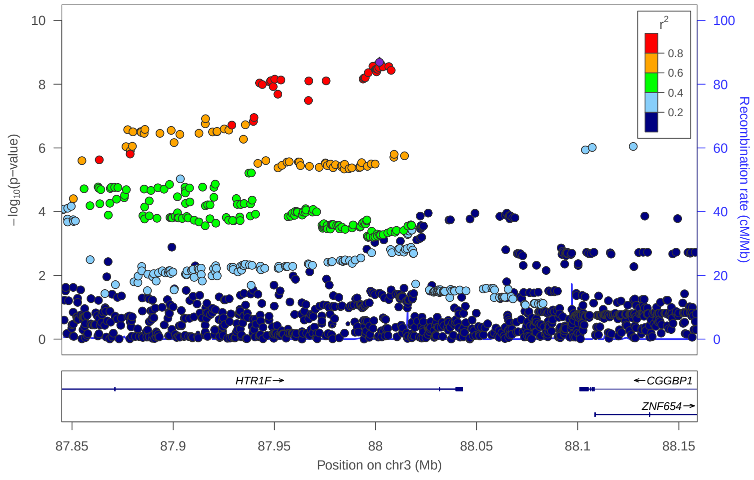


locus 116 rs78531597

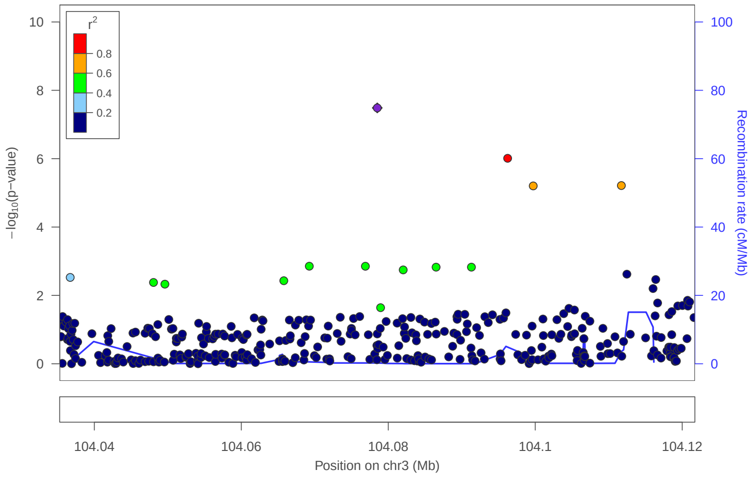


locus 117 rs2016469

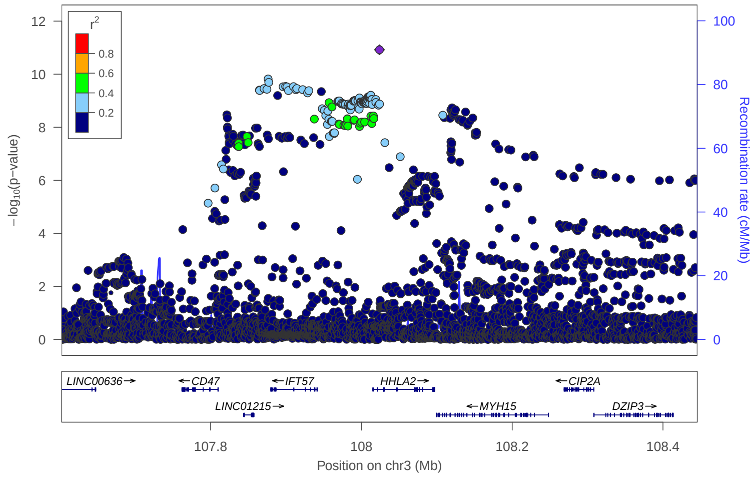


locus 118 rs7619638

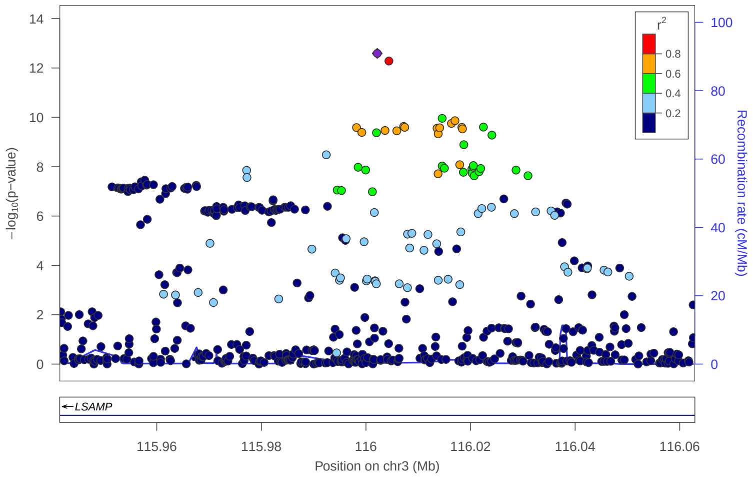


locus 119 rs9815484

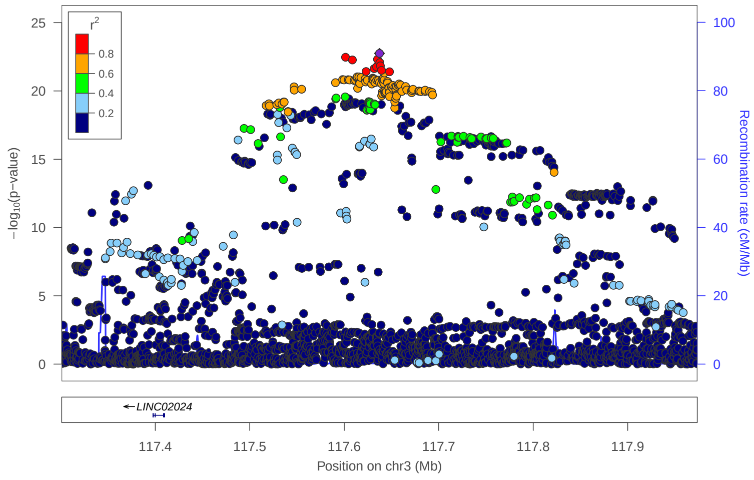


locus 120 rs75869405

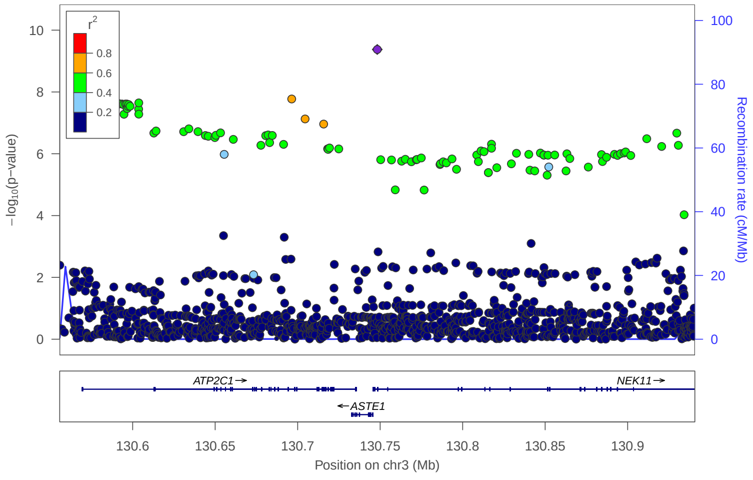


locus 121 rs67294685

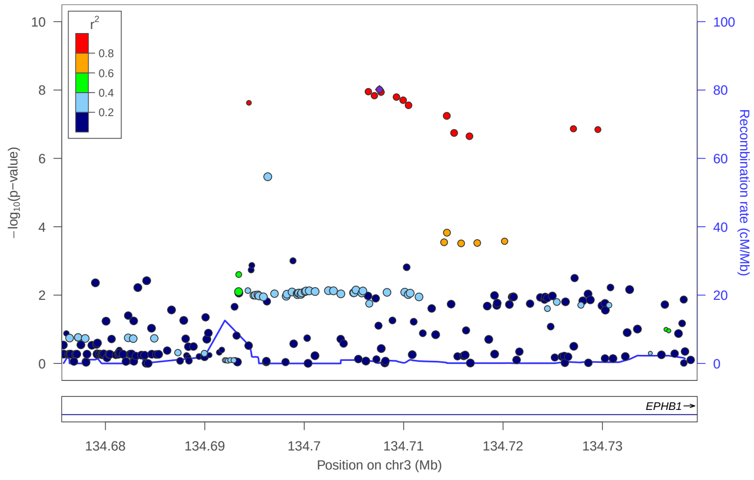

### locus 122 rs4681358

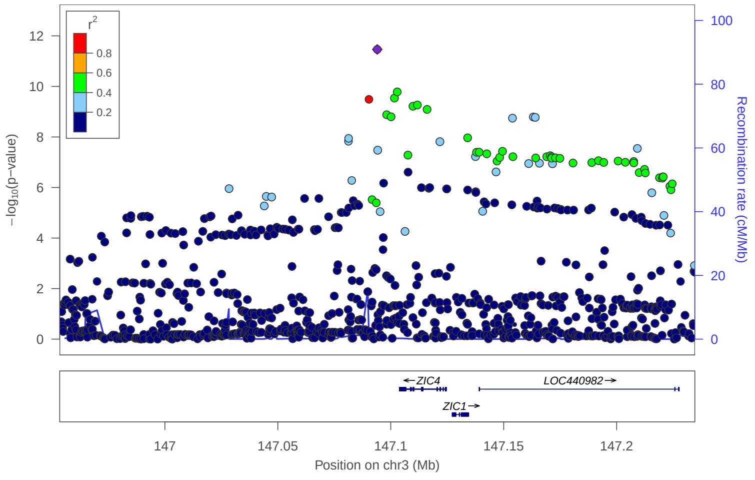


### locus 123 rs55827025

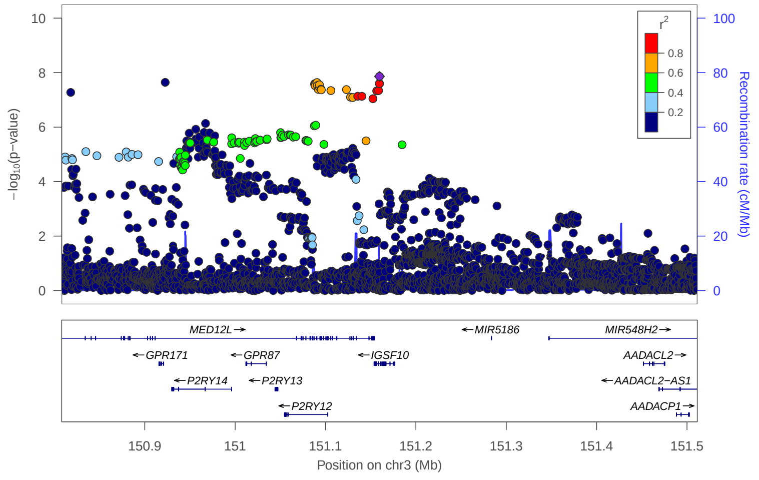


### locus 124 rs9872362

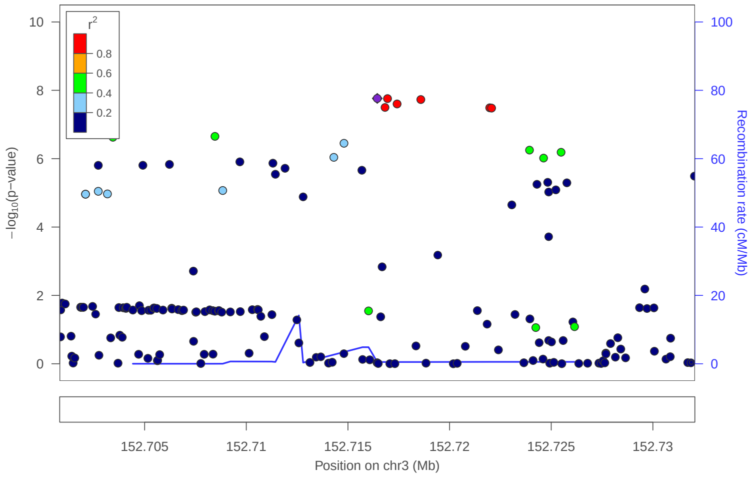


### locus 125 rs13095140

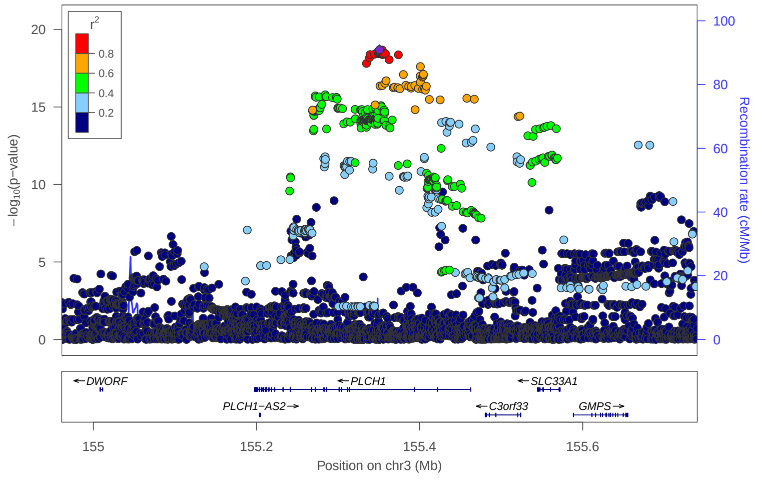


### locus 126 rs2306531

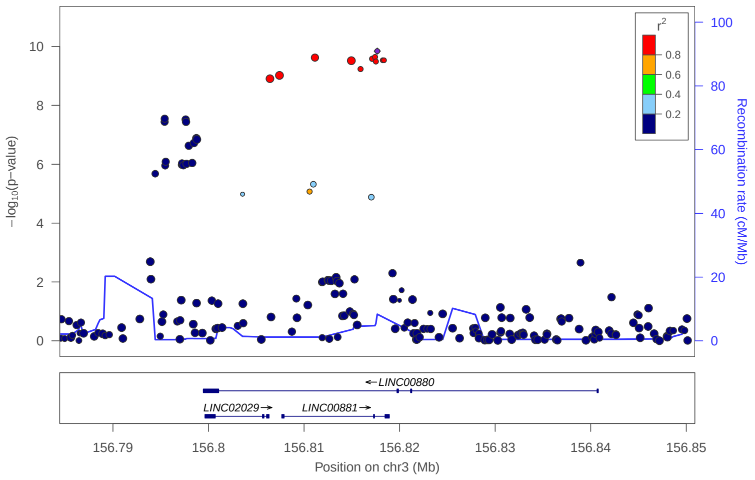


### locus 127 rs34995891

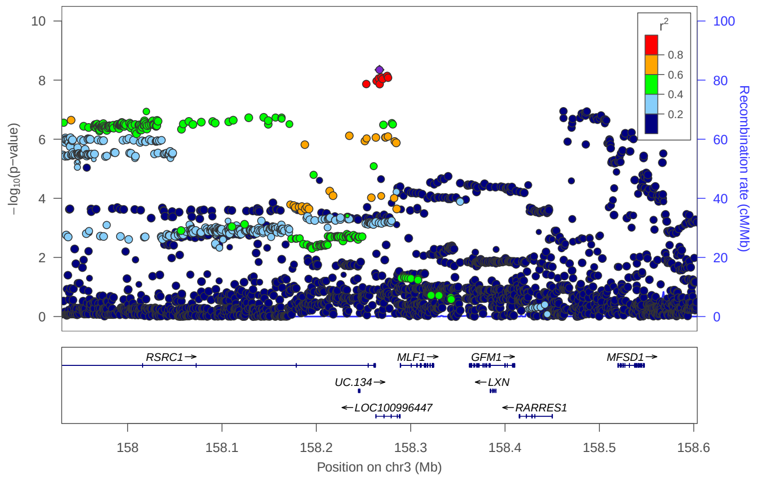


### locus 128 rs111550352

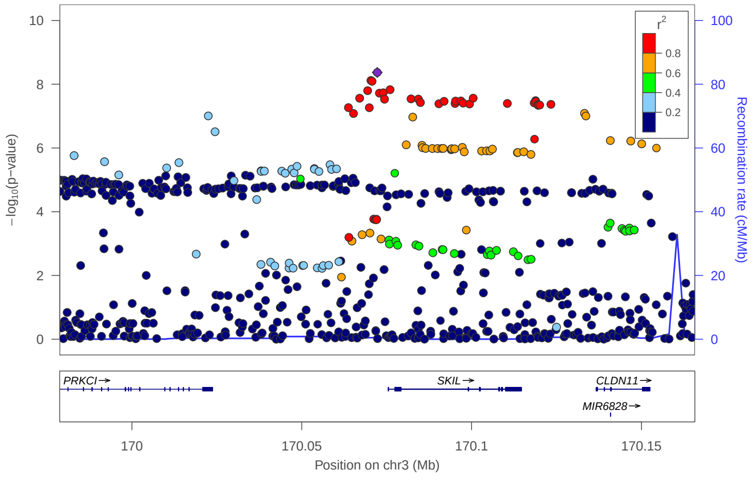


### locus 129 rs10936661

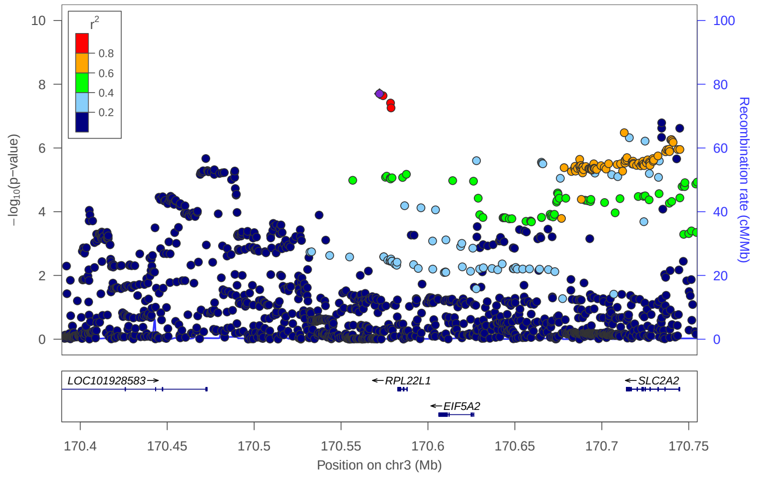

locus 130 rs694786

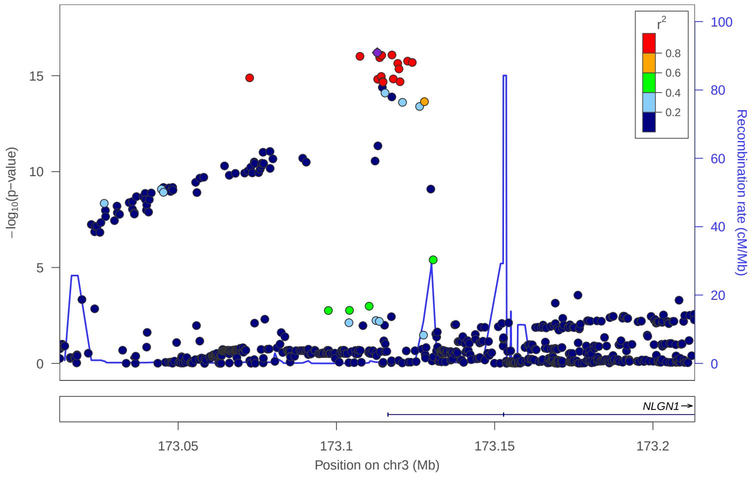


locus 131 rs60018000

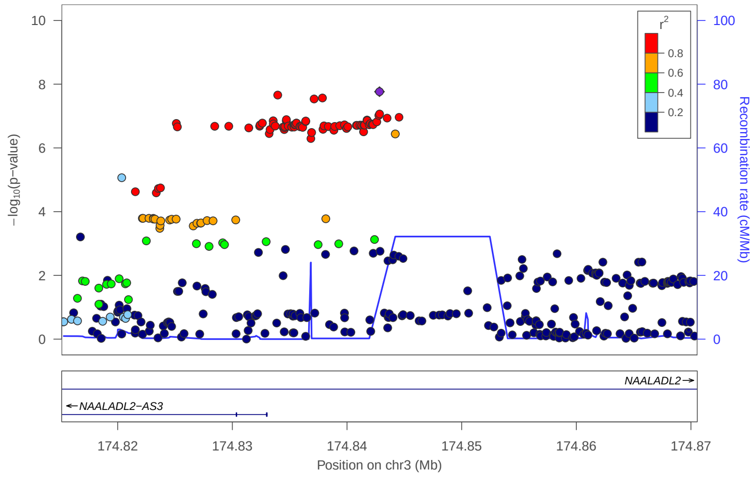


locus 132 rs4260410

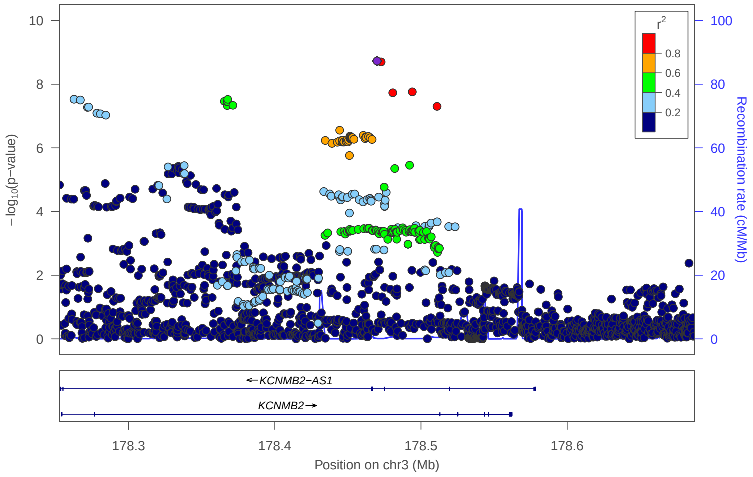


locus 133 rs2216427

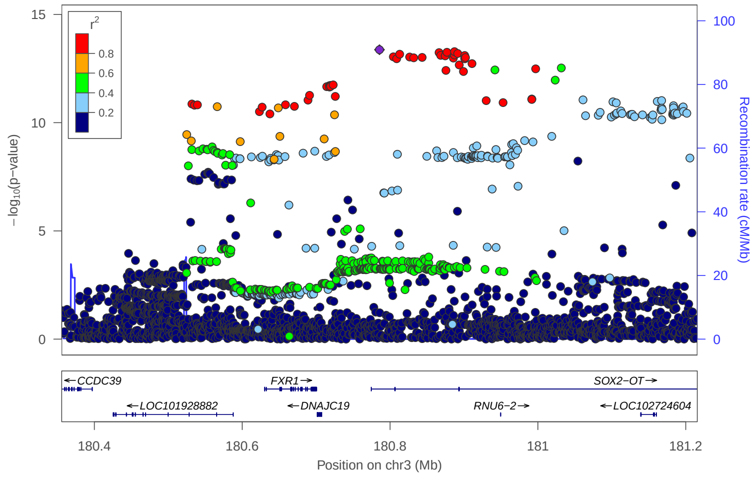


locus 134 rs6786798

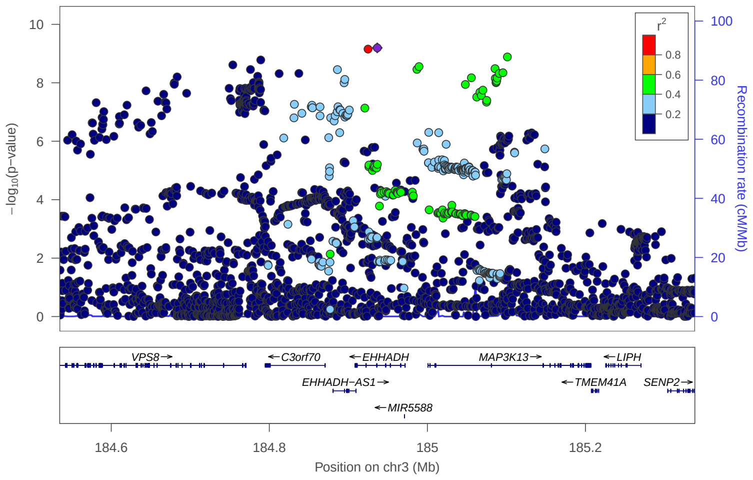


locus 135 rs745850919

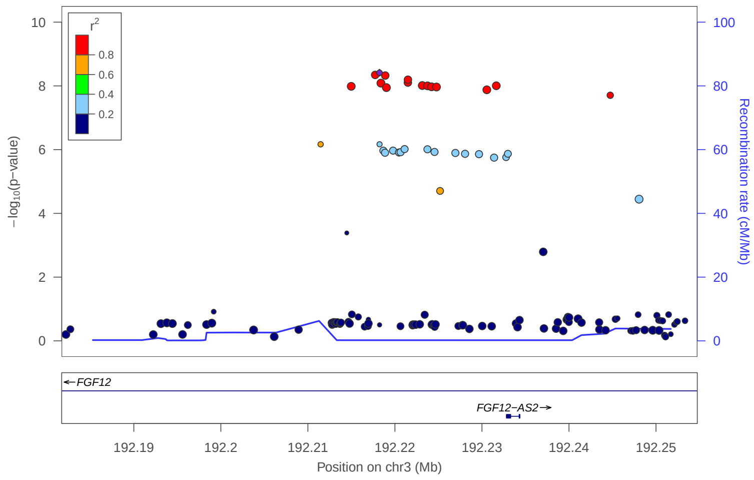


locus 136 rs12647020

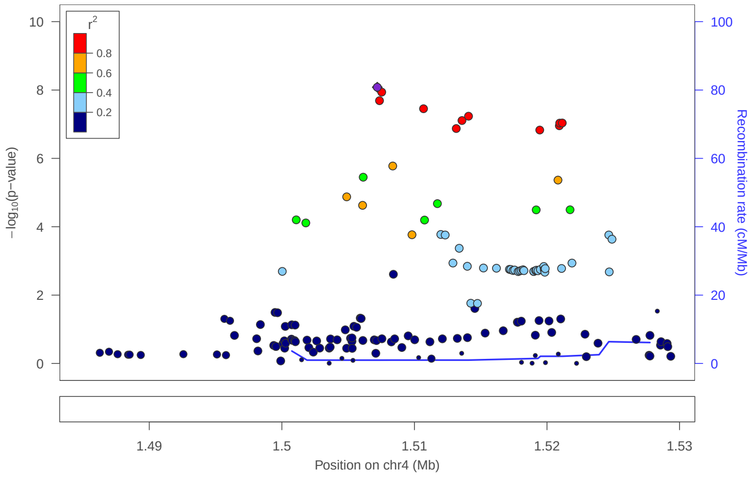


locus 137 rs6857447

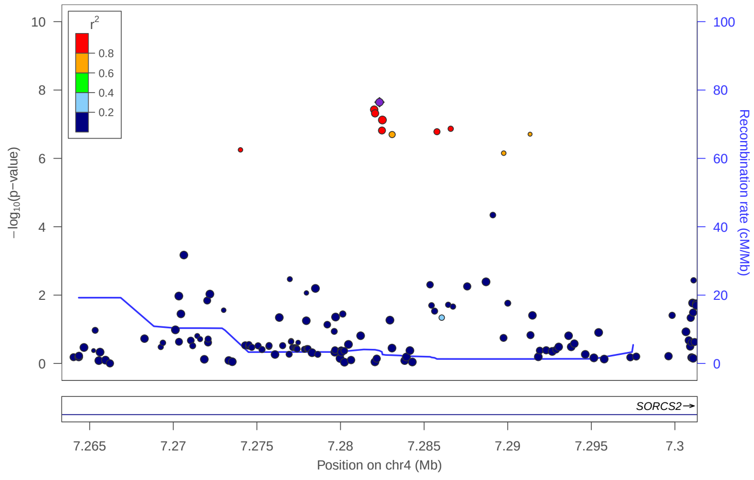

### locus 138 rs2658509

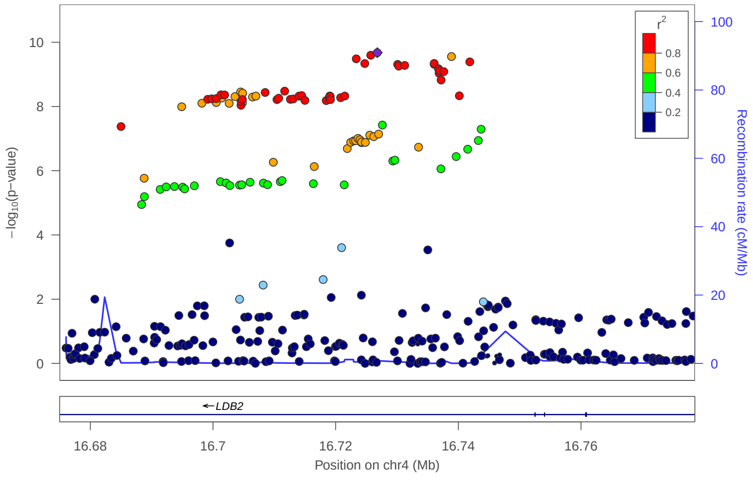


### locus 139 rs113591949

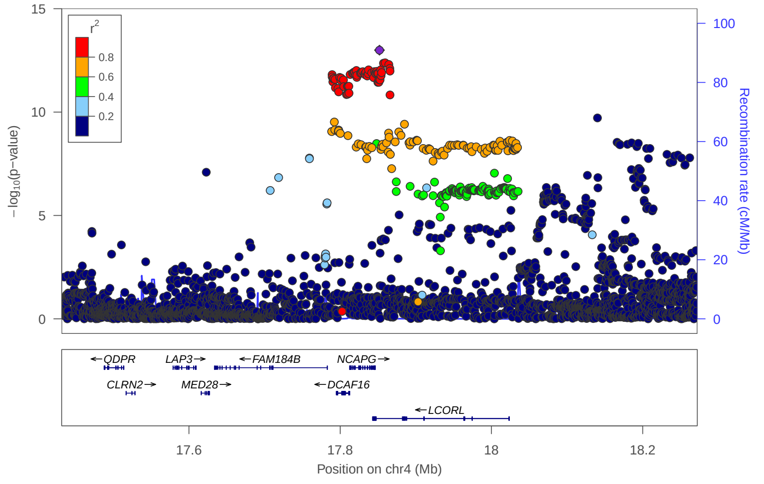


### locus 140 rs34791171

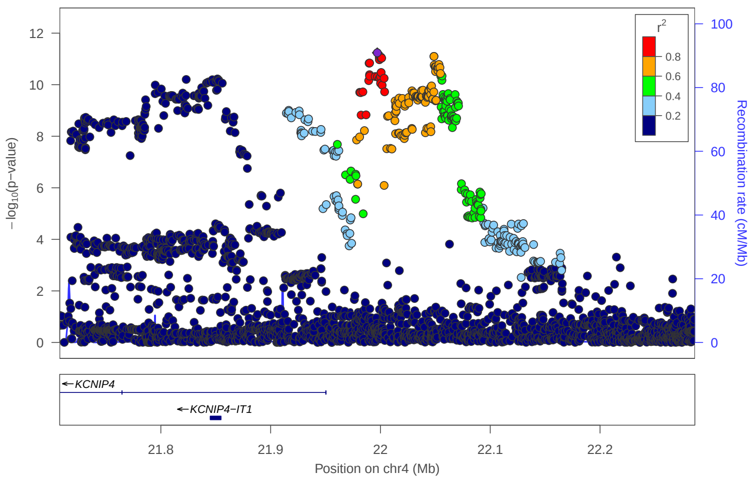


### locus 141 rs2667298

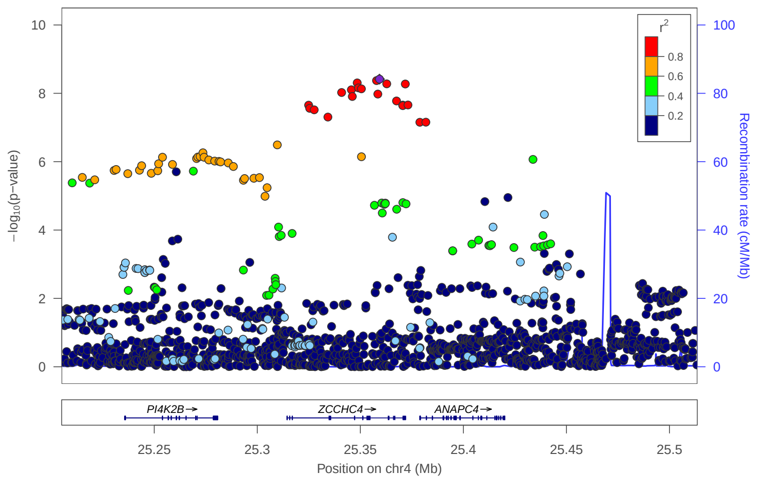


### locus 142 rs190495

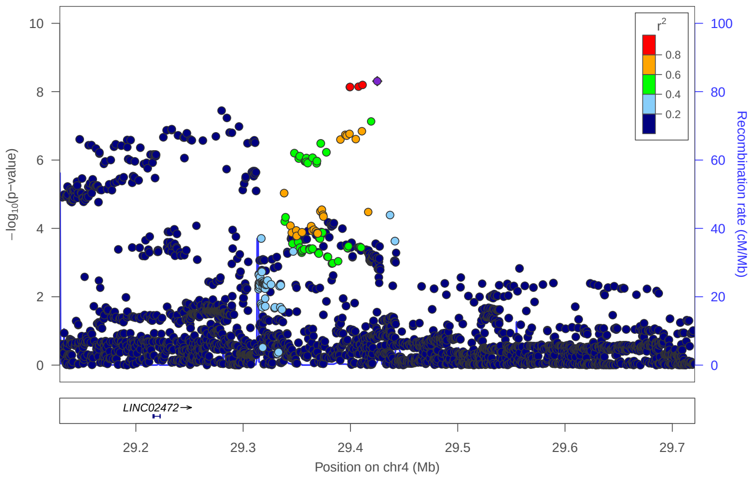


### locus 143 rs10026643

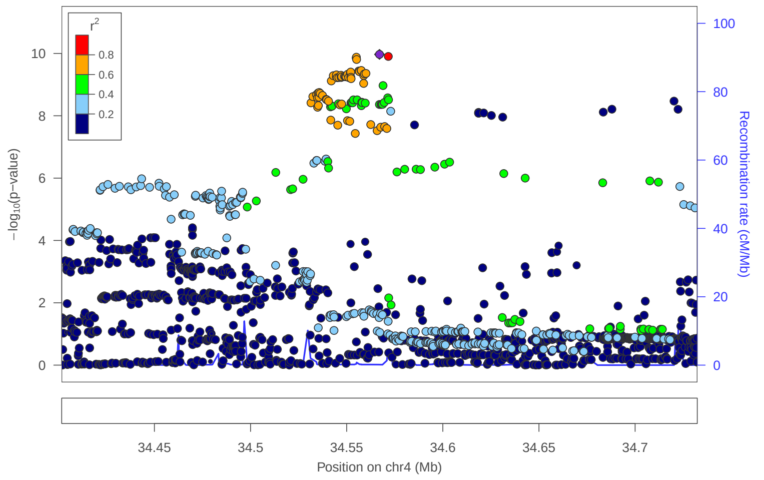


### locus 144 rs2581458

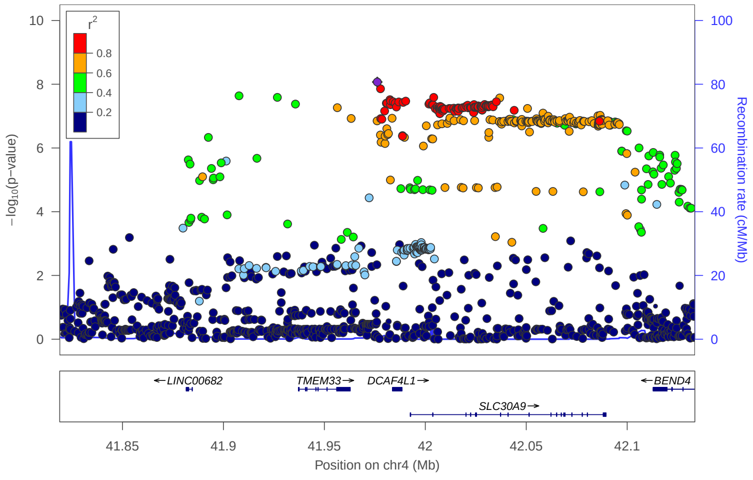


### locus 145 rs10517119

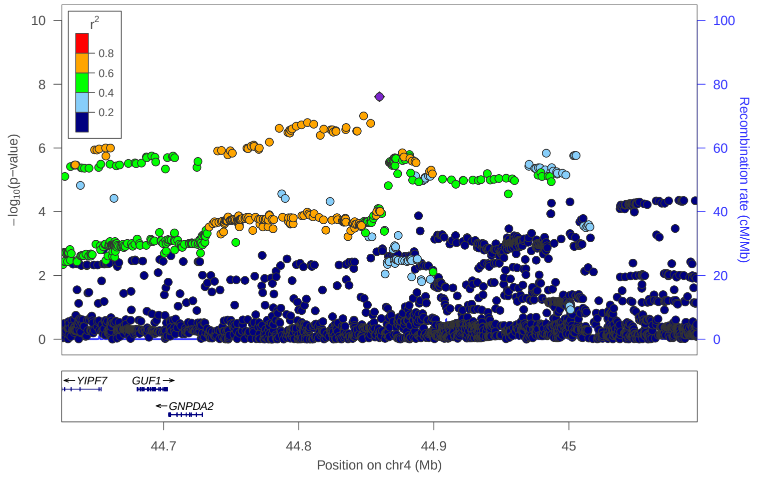

### locus 146 rs4860090

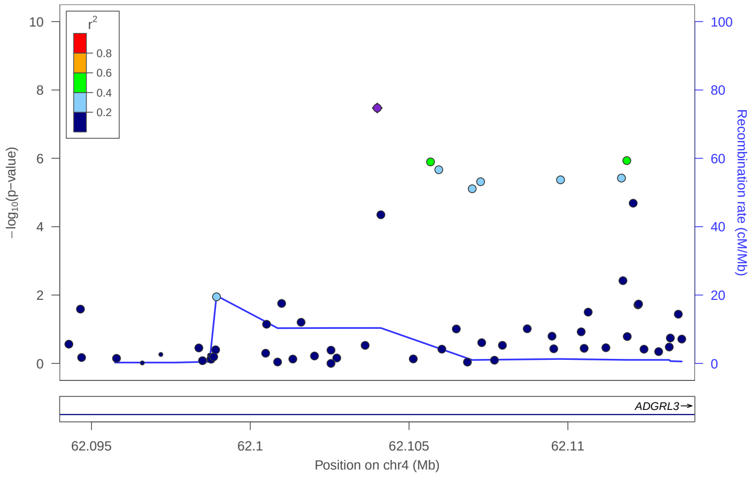


### locus 147 rs6812227

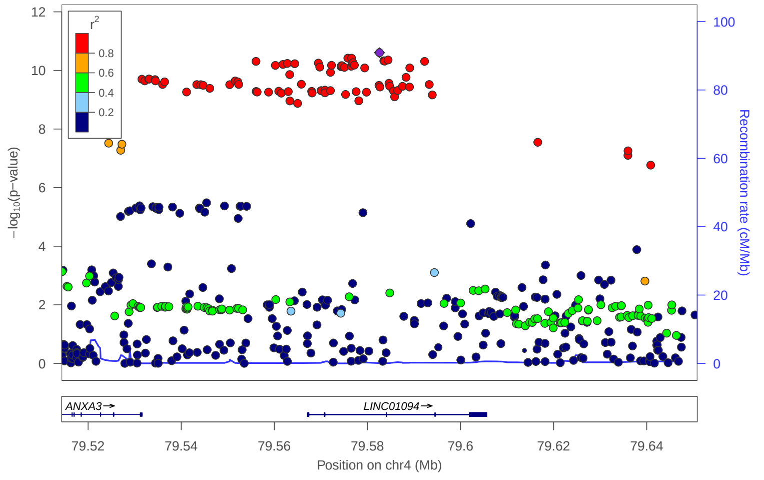


### locus 148 rs1132460

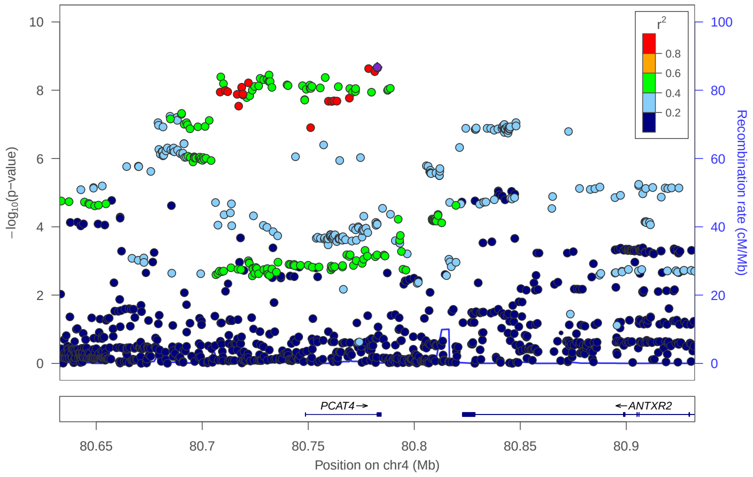


### locus 149 rs2011962

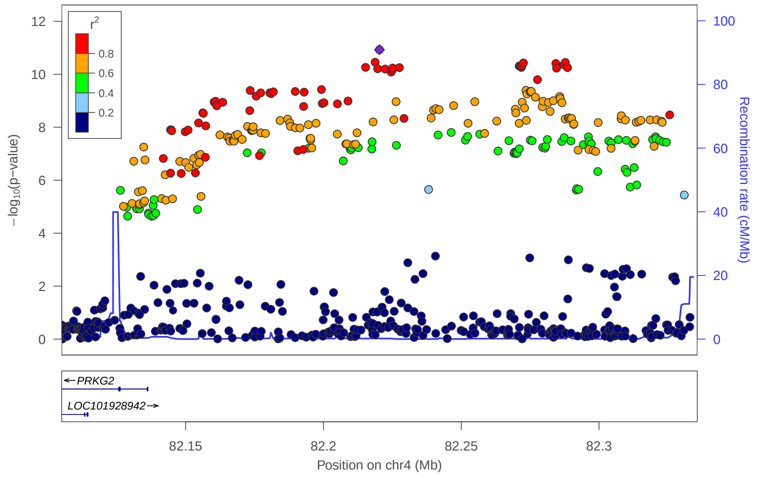


### locus 150 rs356179

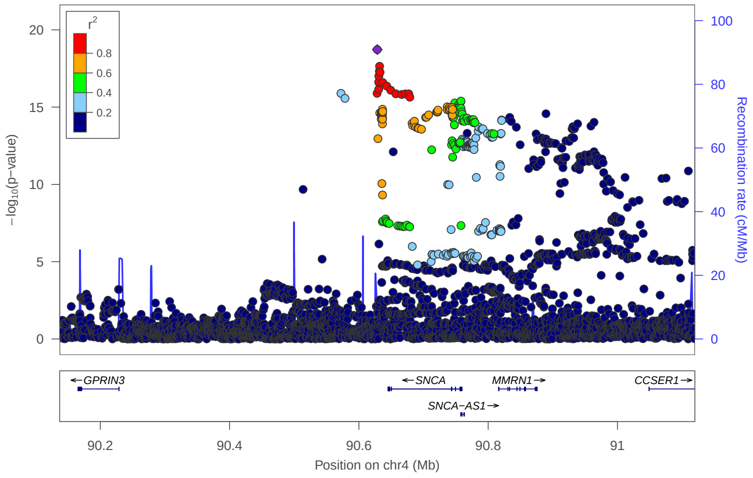


### locus 151 rs11730384

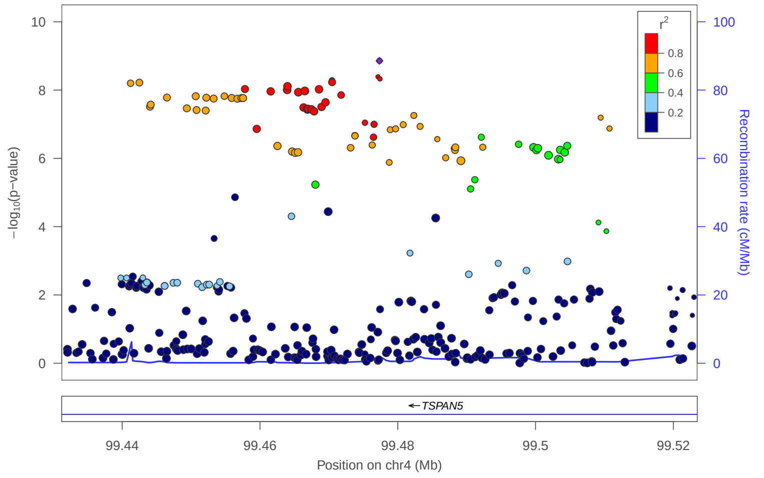


### locus 152 rs11940529

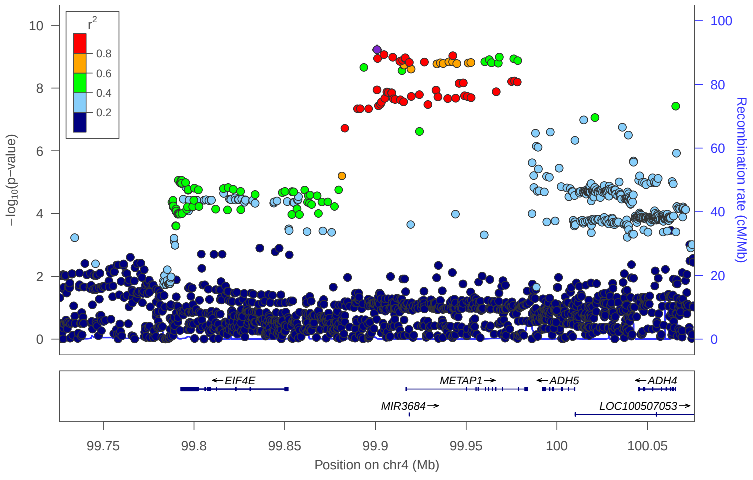


### locus 153 rs62307308

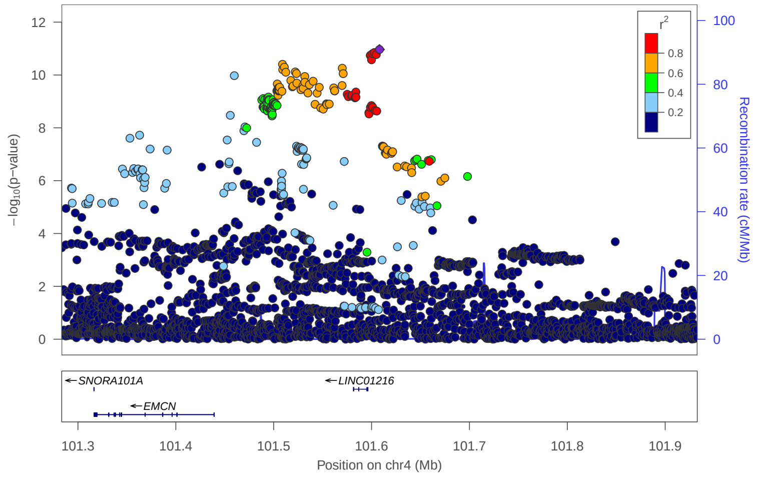

### locus 154 rs13107325

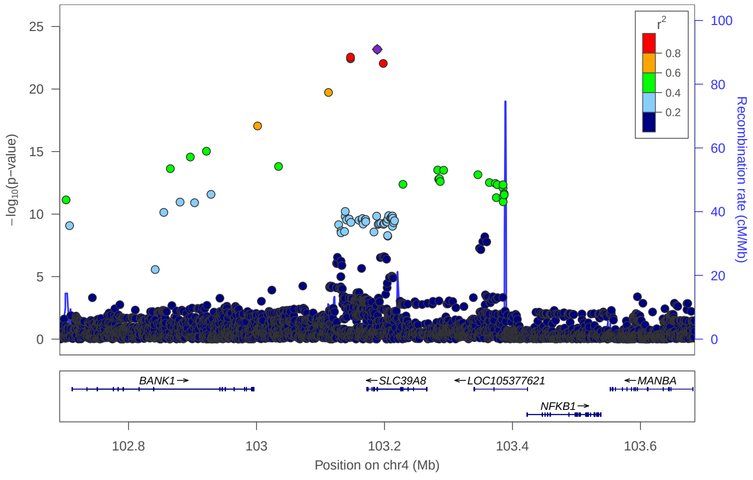


### locus 155 rs2903385

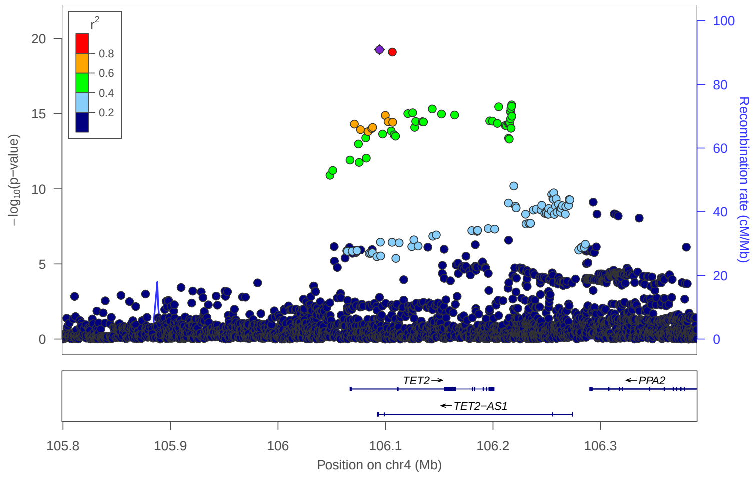


### locus 156 rs13108200

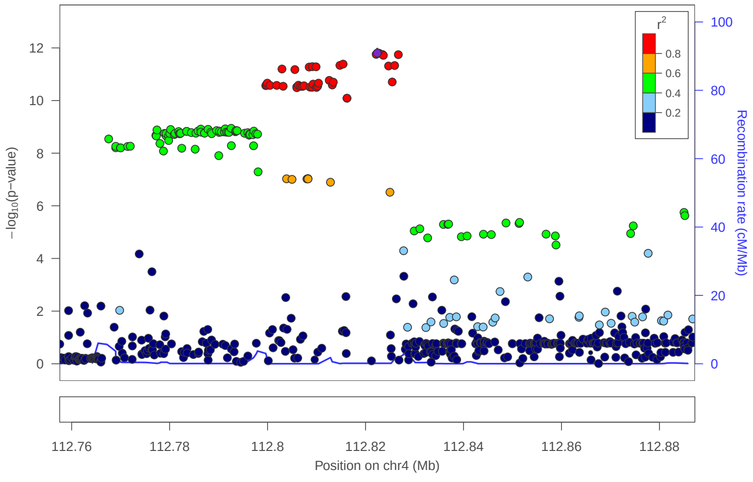


### locus 157 rs6533604

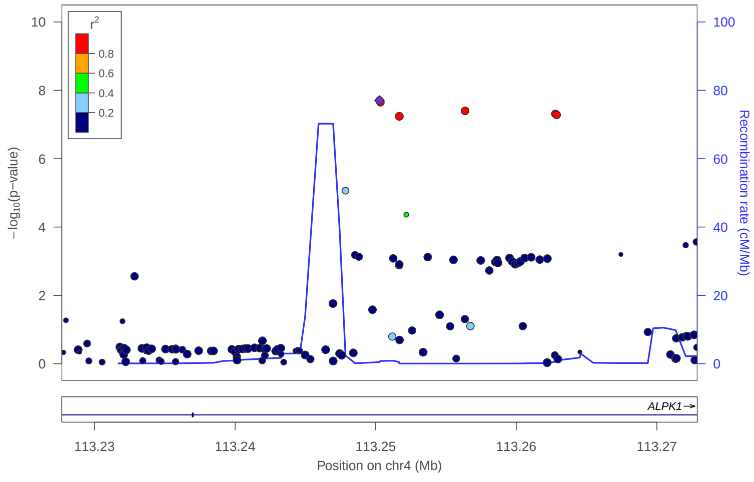


### locus 158 rs45510091

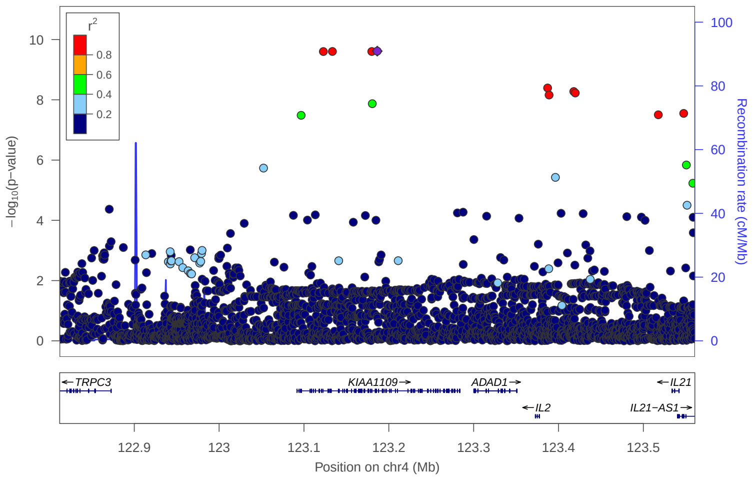


### locus 159 rs7657954

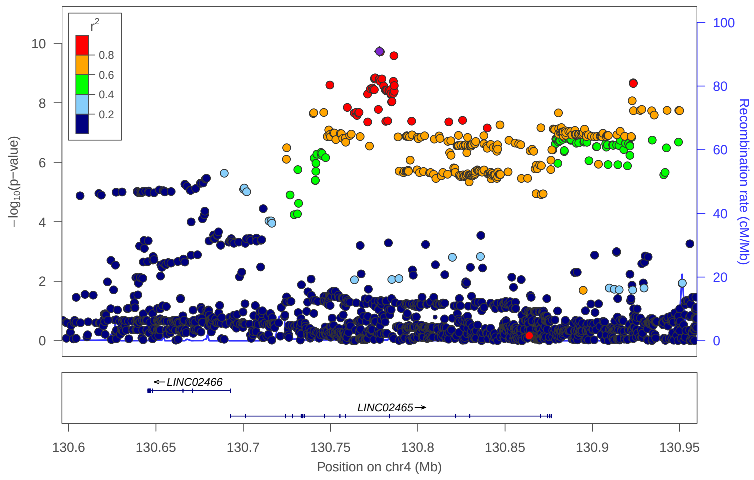


### locus 160 rs3733421

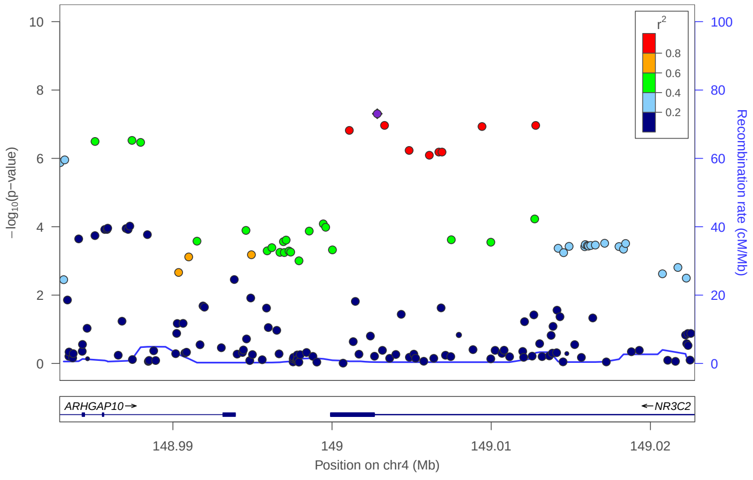


### locus 161 rs1820514

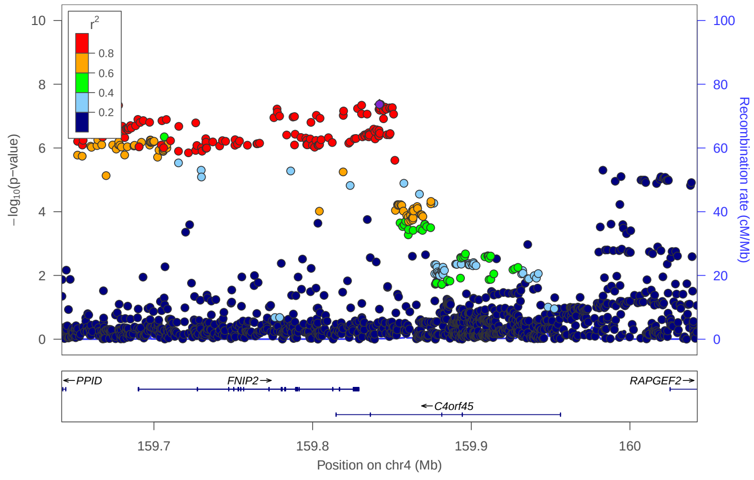

locus 162 rs11729015

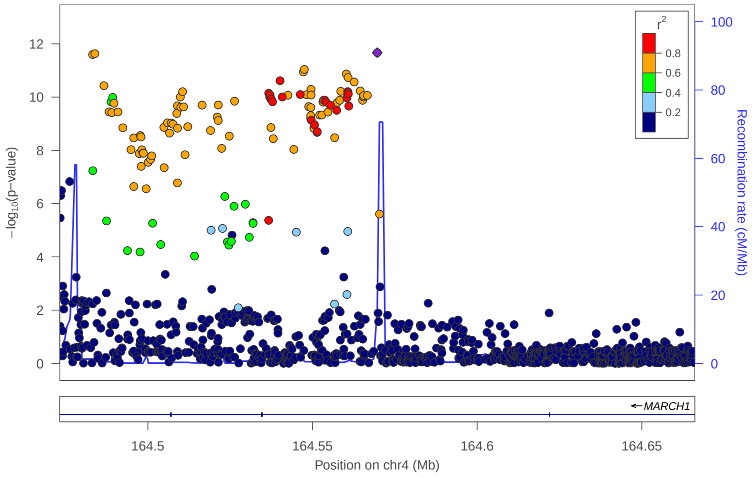


locus 163 rs11724690

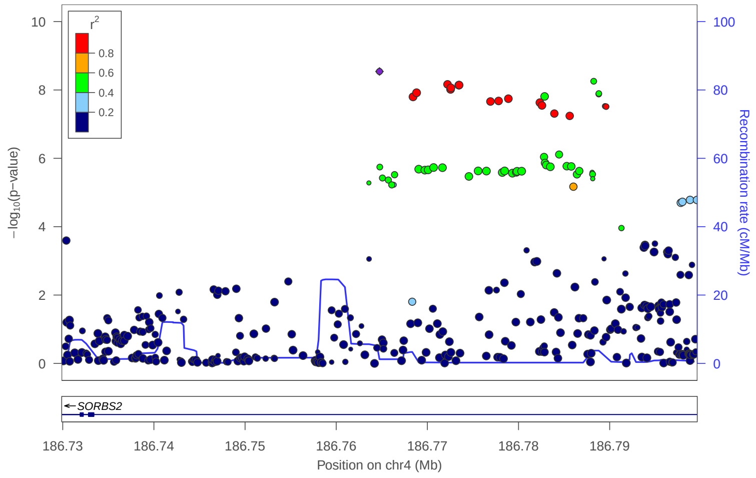


locus 164 rs147728881

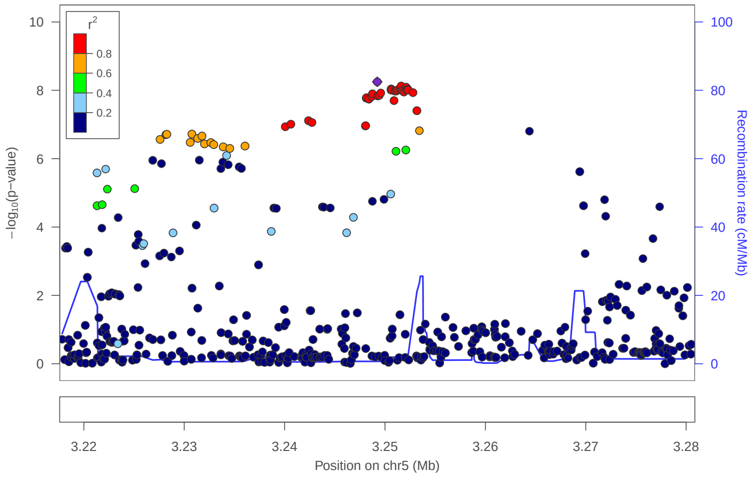


locus 165 rs4454035

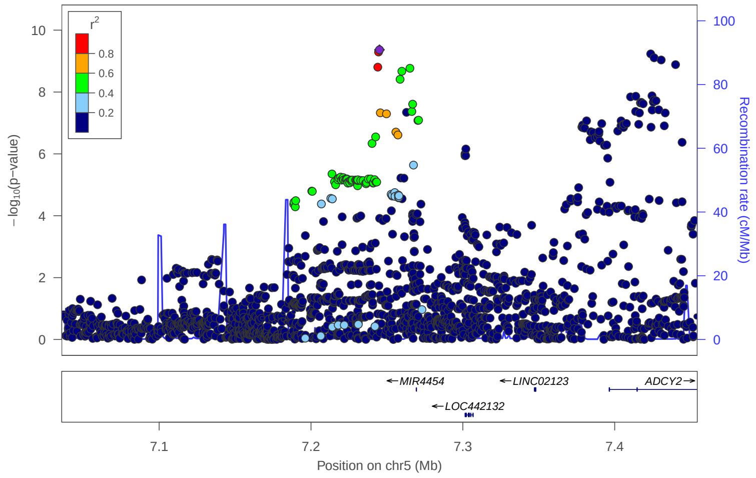


locus 166 rs6872919

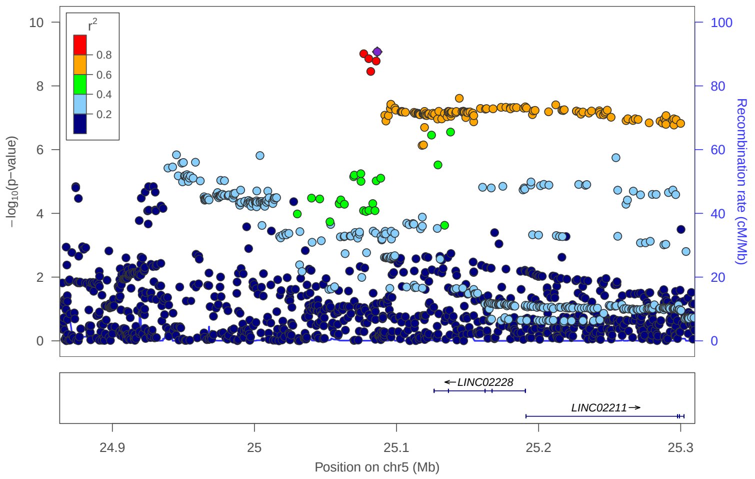


locus 167 rs34971686

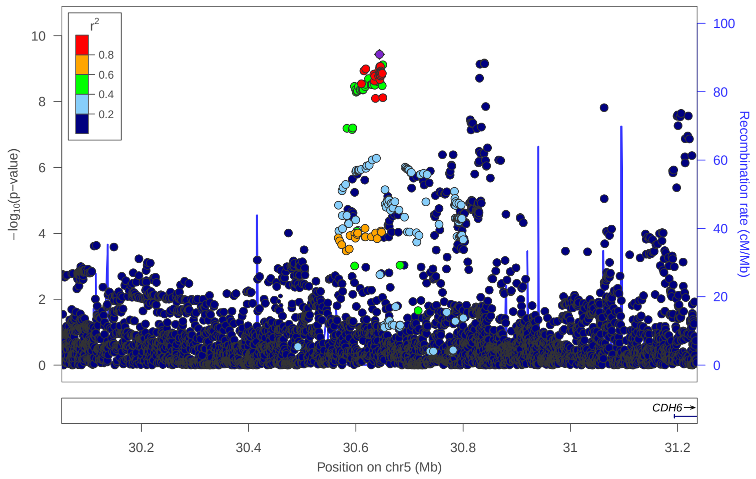


locus 168 rs77176363

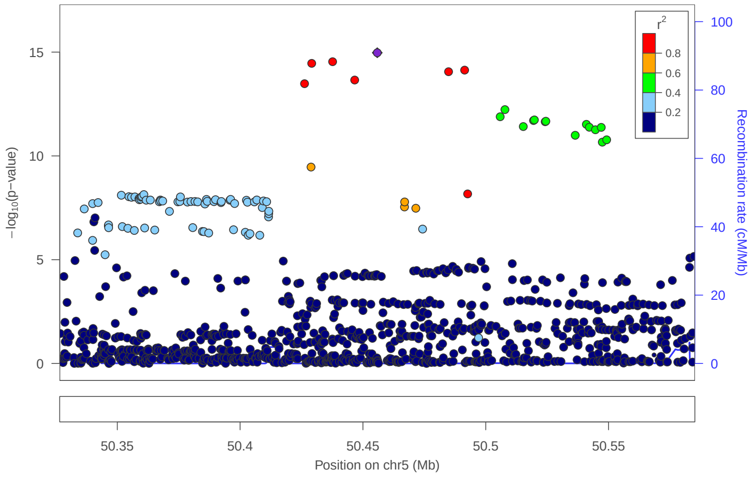


locus 169 rs6449590

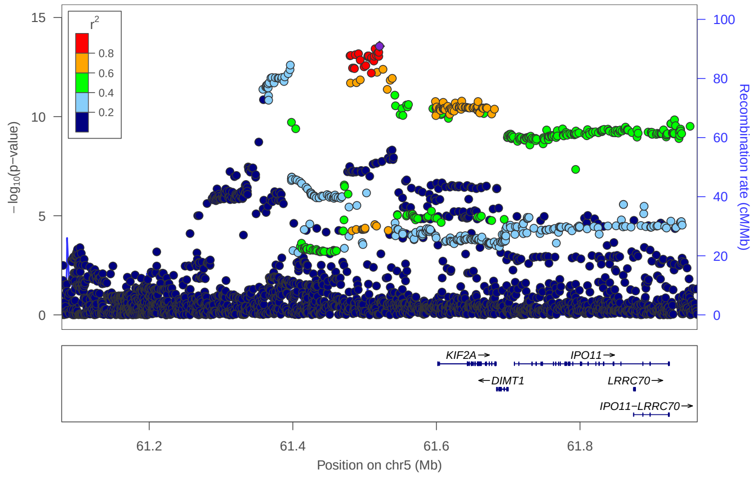

### locus 170 rs669245

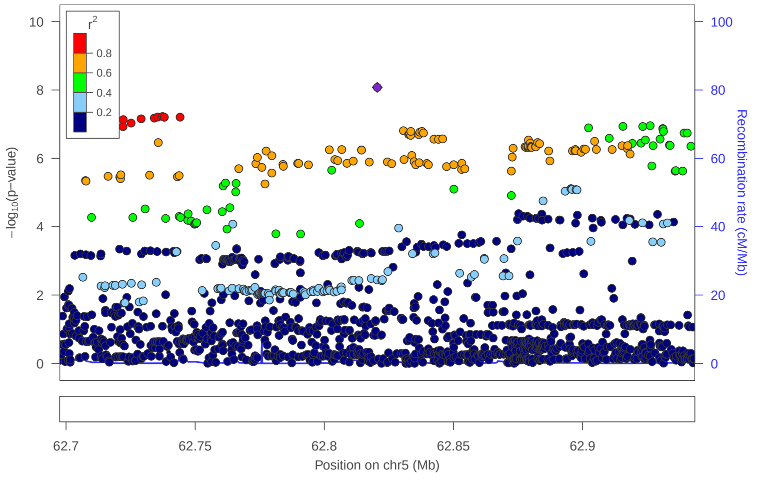


### locus 171 rs6864168

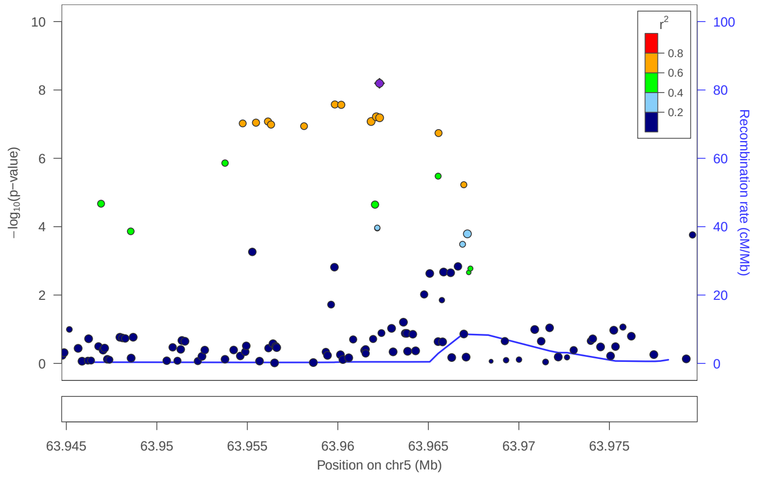


### locus 172 rs62363245

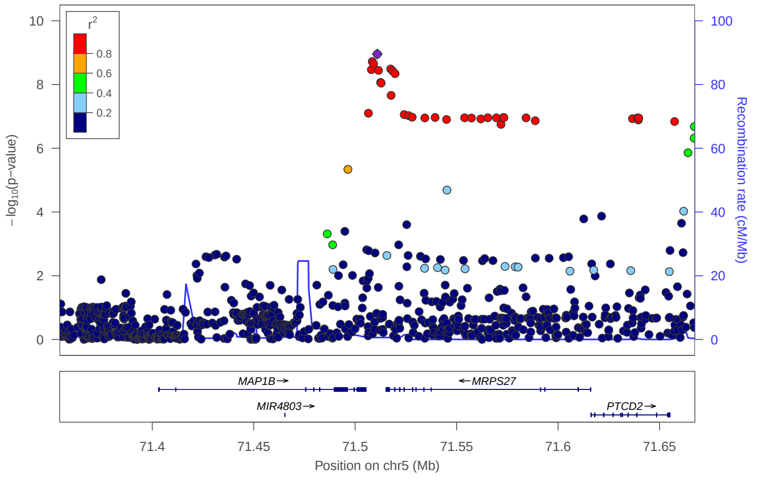


### locus 173 rs11949379

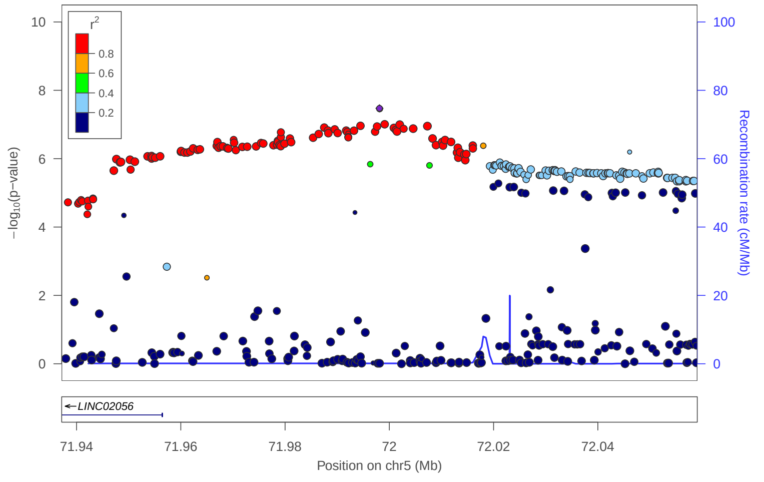


### locus 174 rs7708900

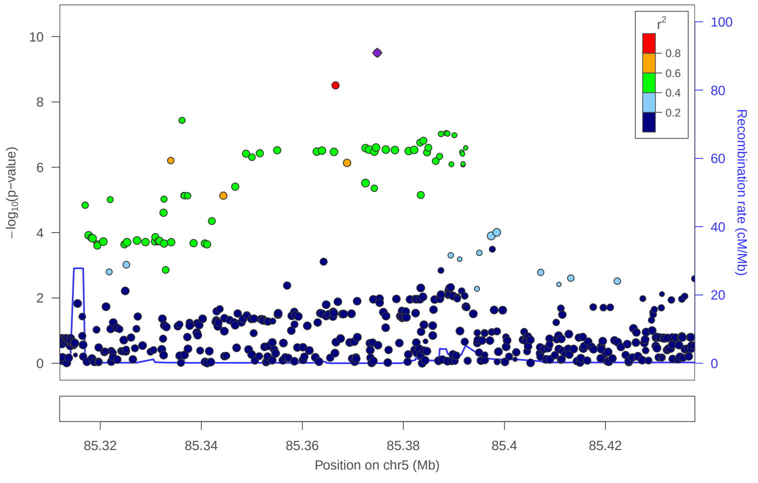


### locus 175 rs34164261

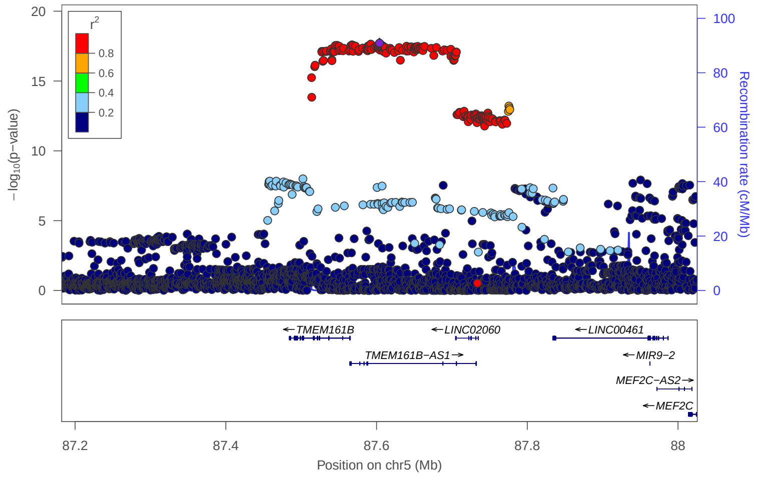


### locus 176 rs4492142

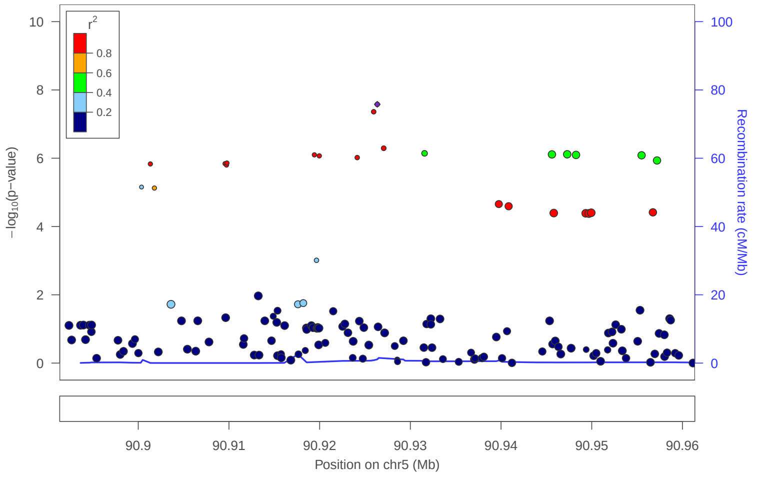


### locus 177 rs34315689

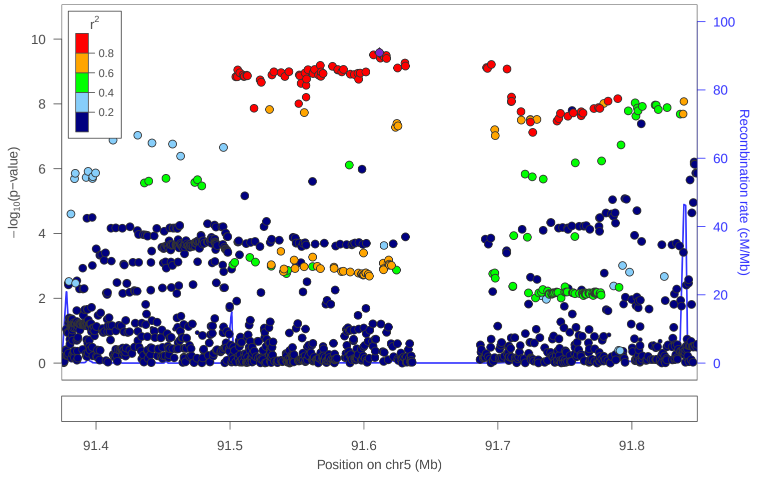

### locus 178 rs11135493

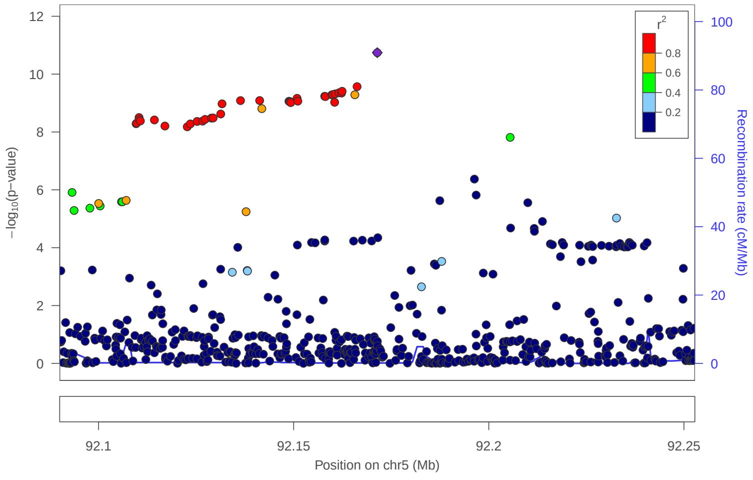


### locus 179 rs10066873

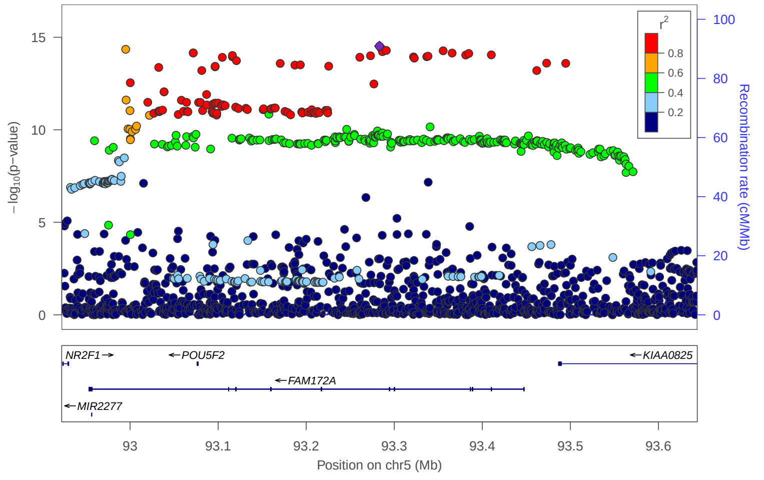


### locus 180 rs2569013

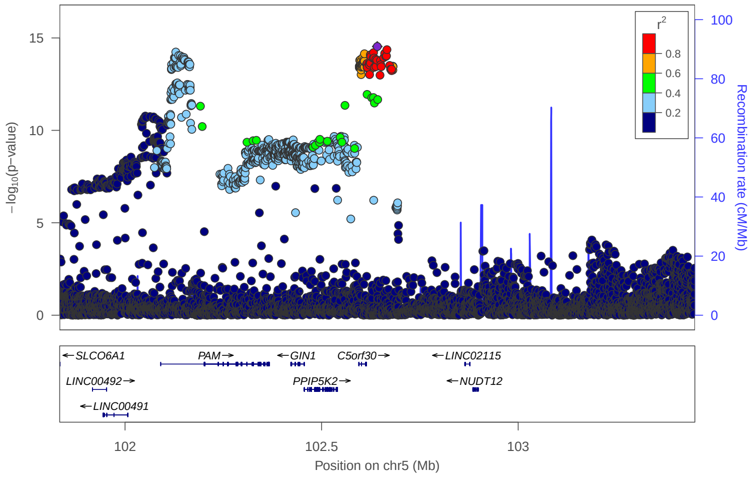


### locus 181 rs2431108

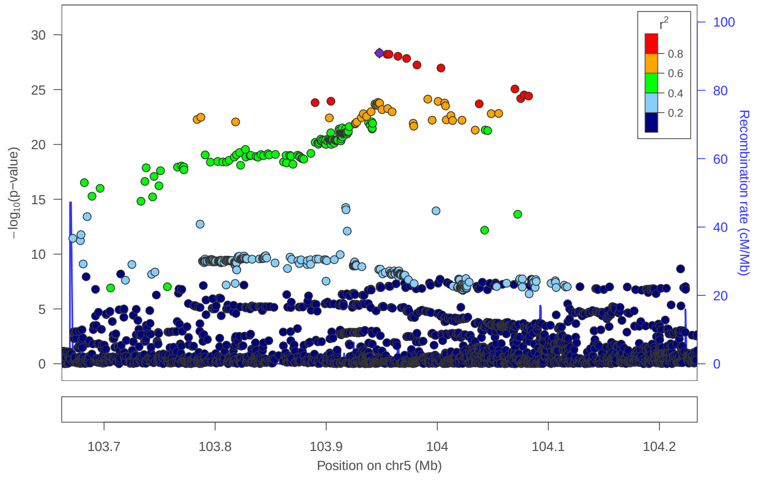


### locus 182 rs71575448

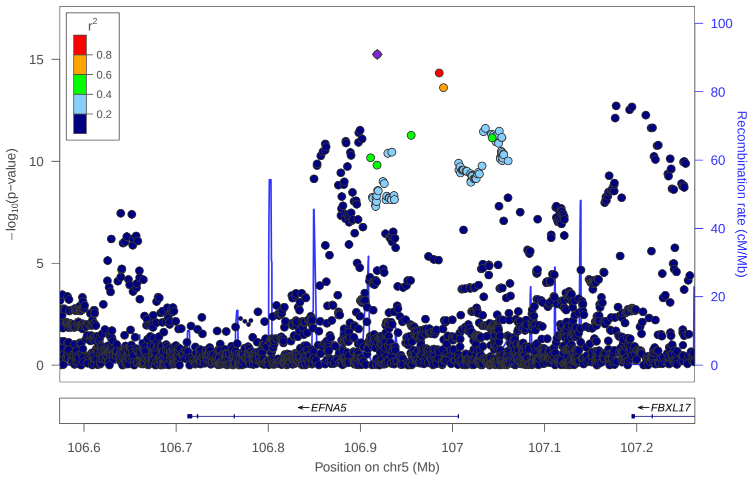


### locus 183 rs872621

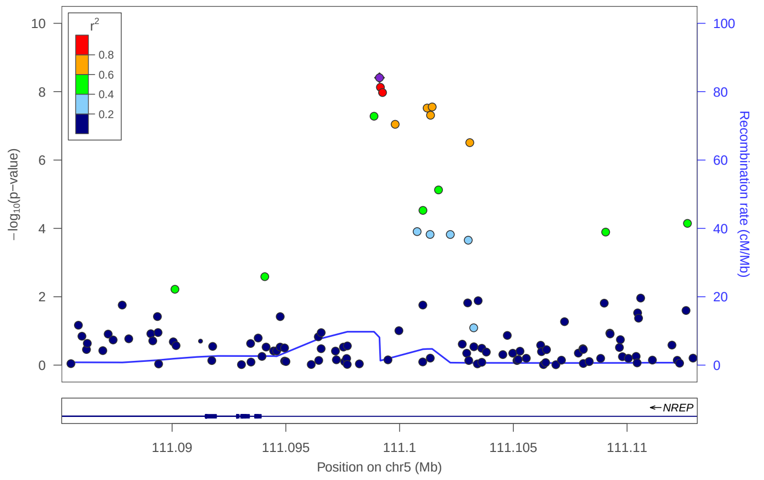


### locus 184 rs62372418

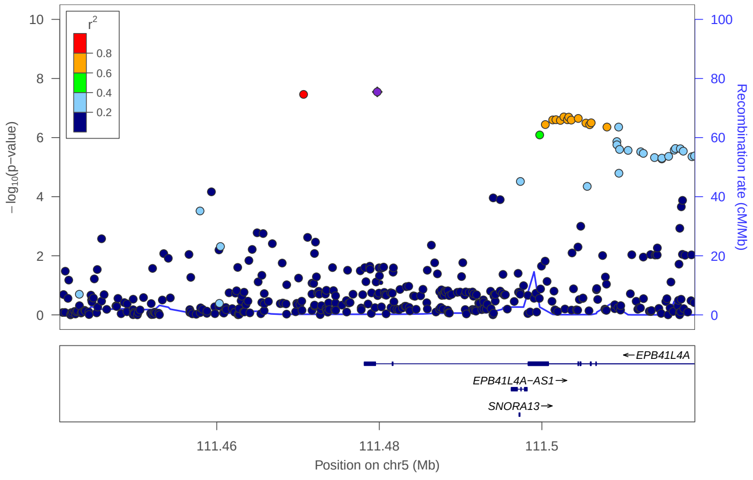


### locus 185 rs11954162

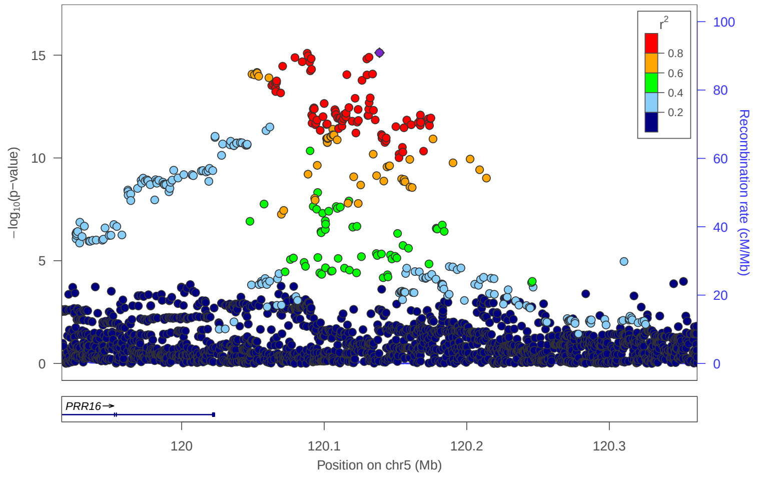

locus 186 rs7733986

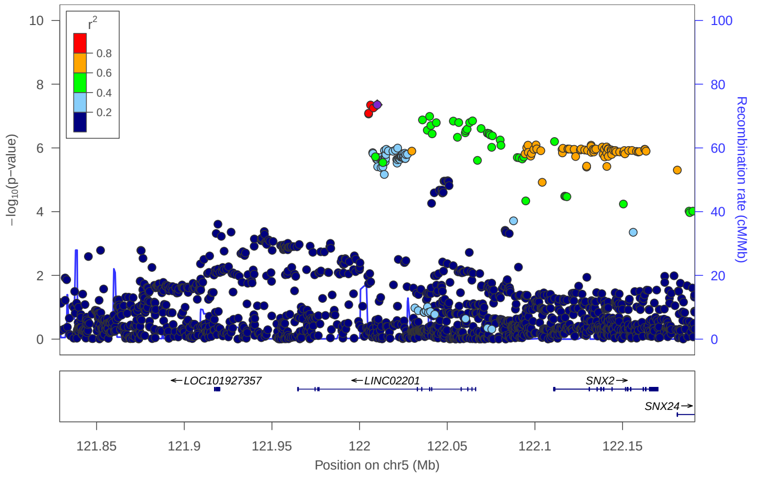


locus 187 rs35681248

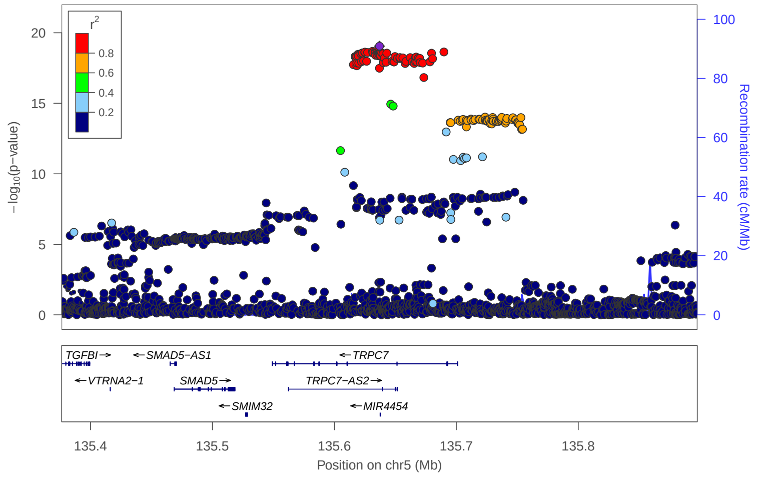


locus 188 rs10043722

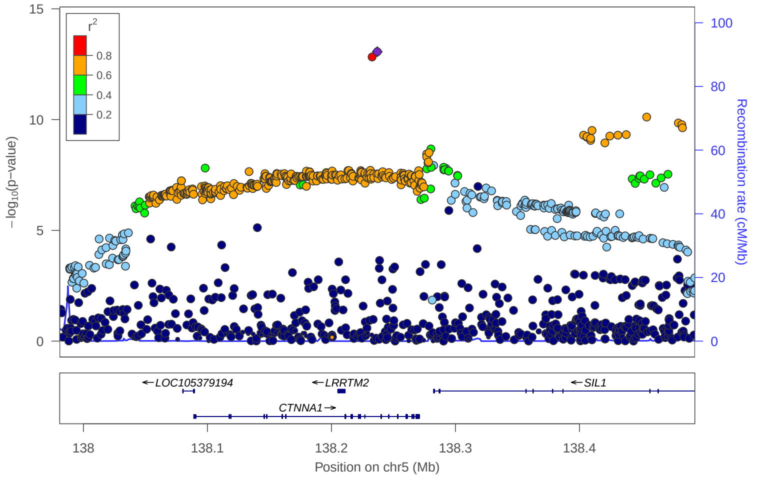


locus 189 rs251369

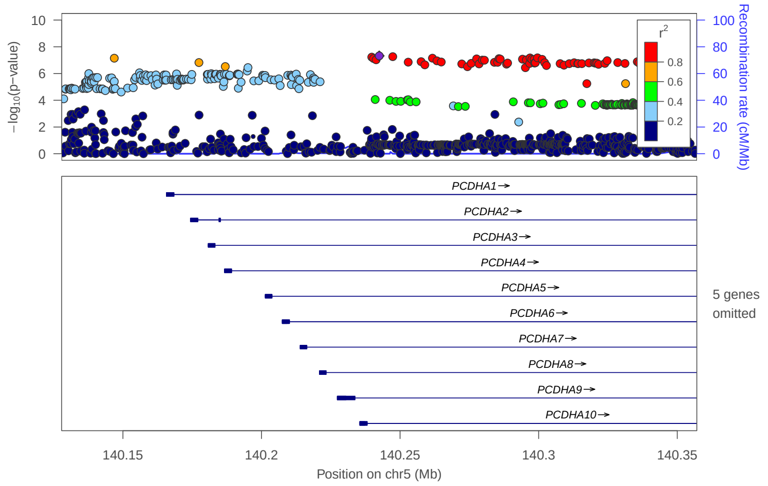


locus 190 rs6888135

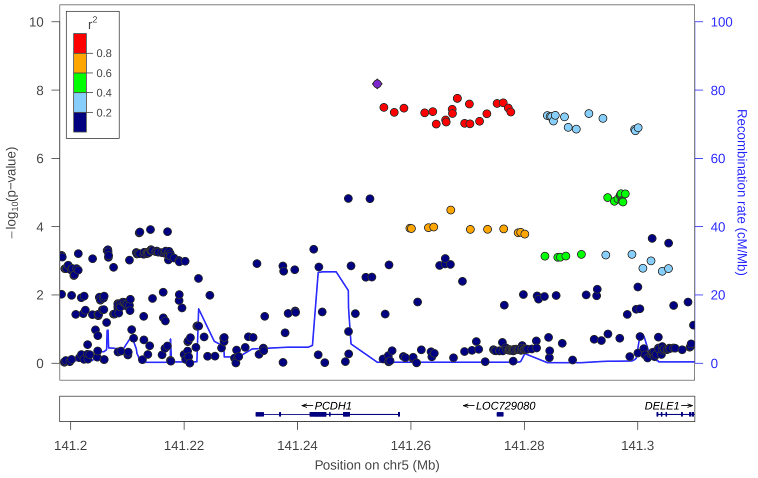


locus 191 rs4912844

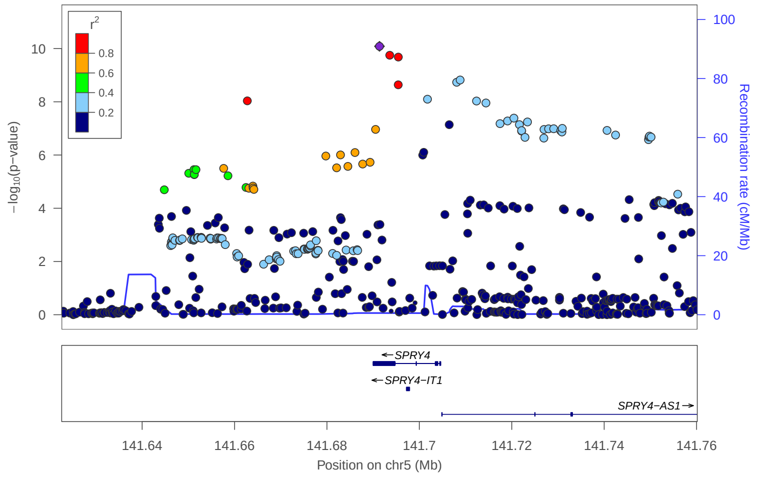


locus 192 rs463245

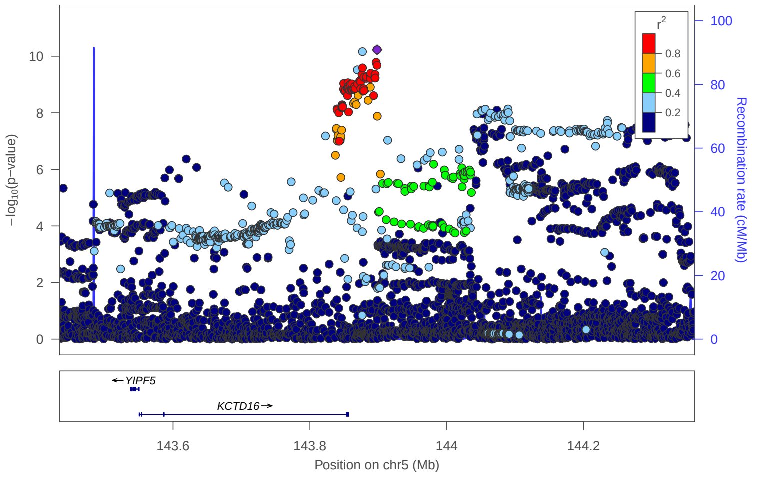


locus 193 rs6580564

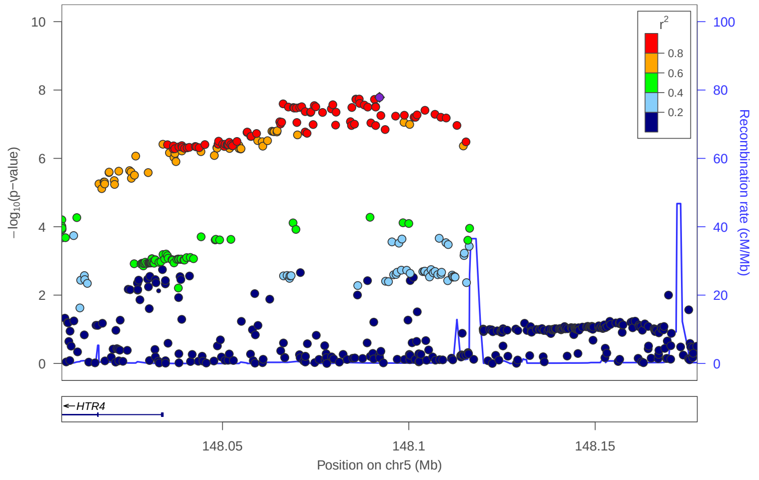

### locus 194 rs10045427

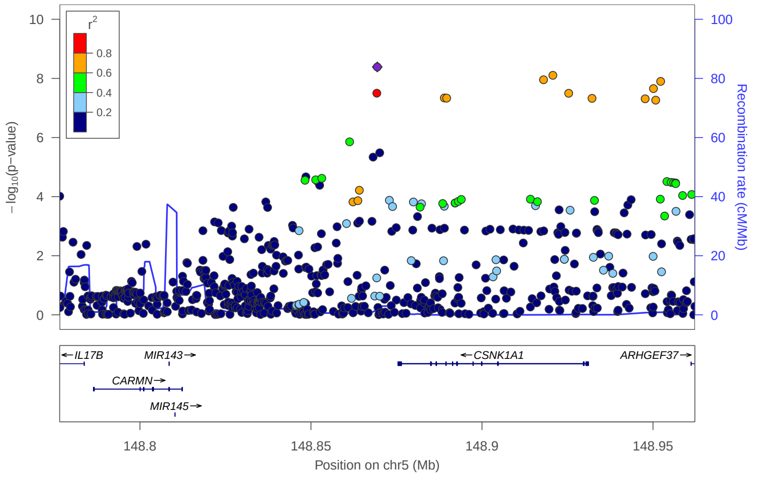


### locus 195 rs10035289

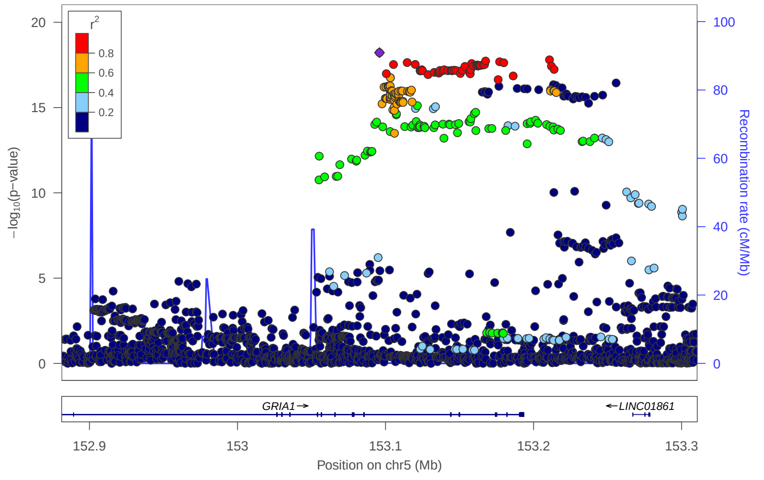


### locus 196 rs11956866

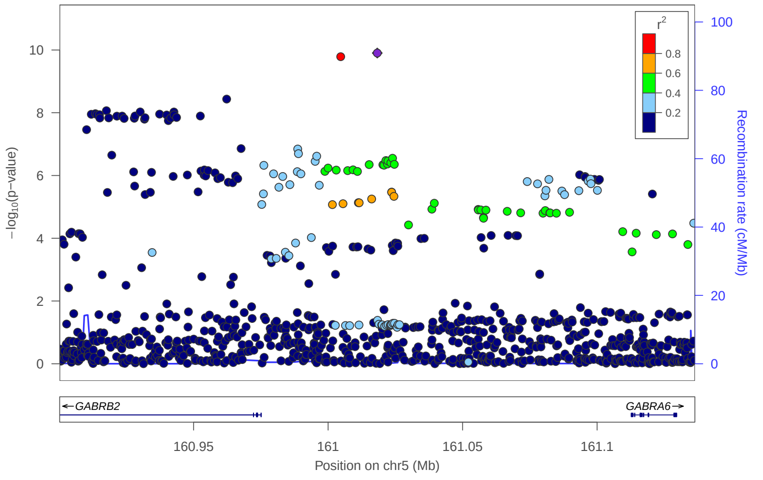


### locus 197 rs62383290

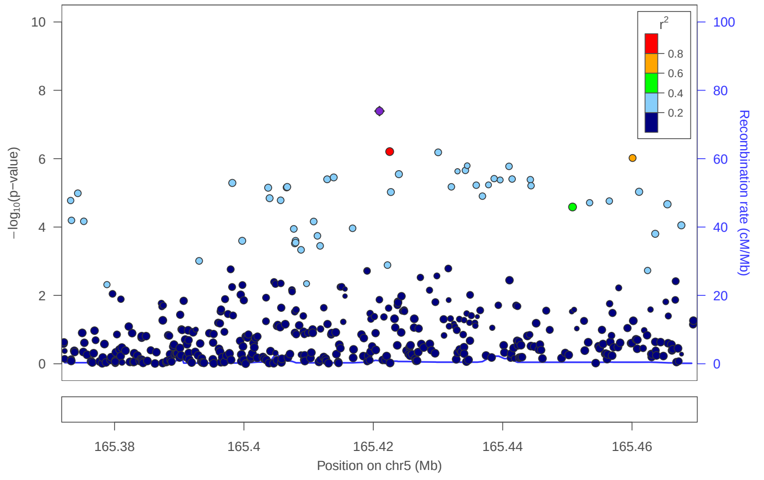


### locus 198 rs9313374

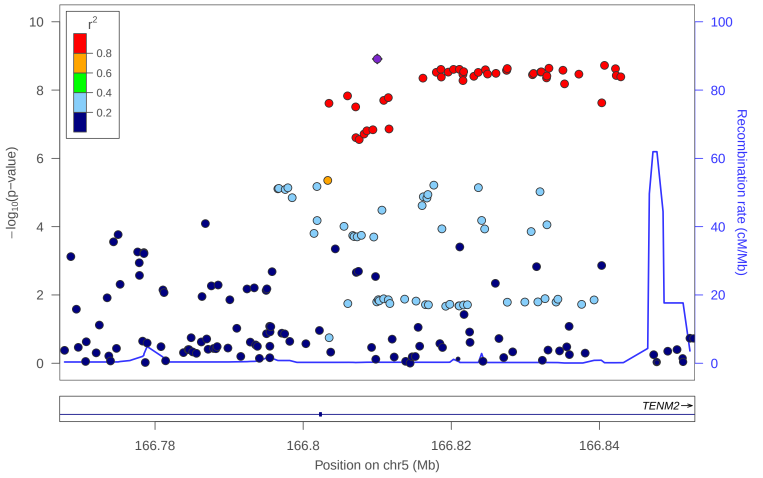


### locus 199 rs879483

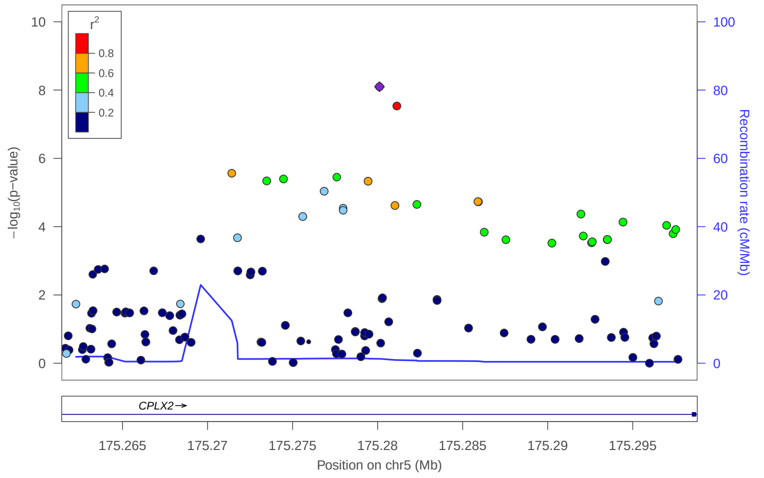


### locus 200 rs6878050

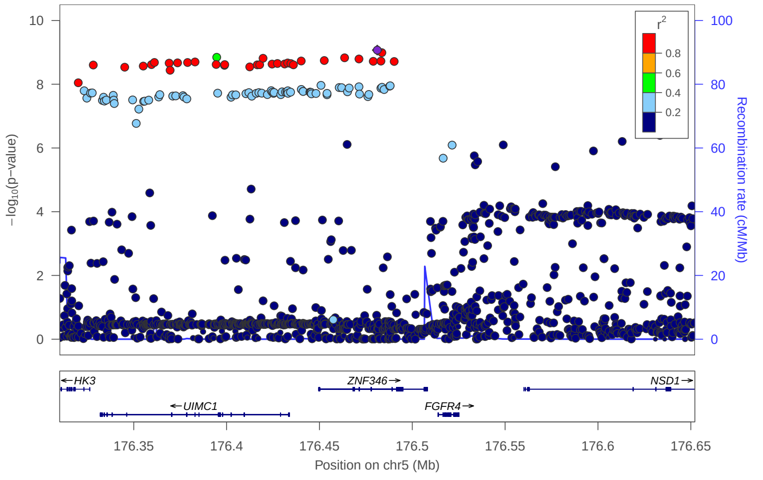


### locus 201 rs4290977

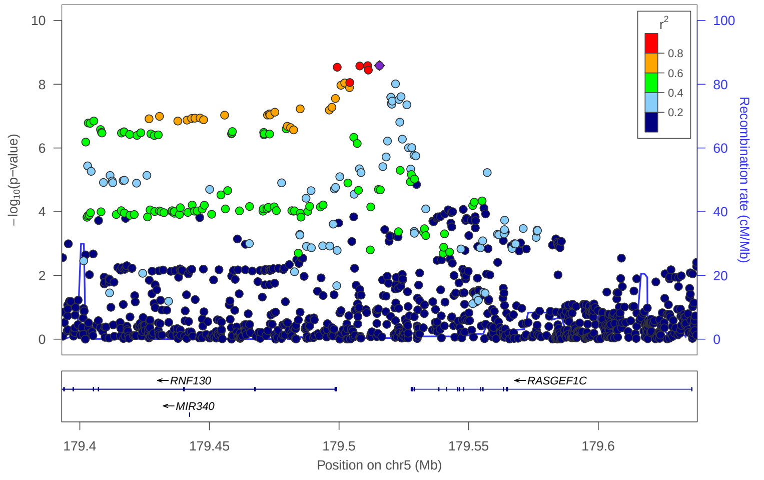

### locus 202 rs377472105

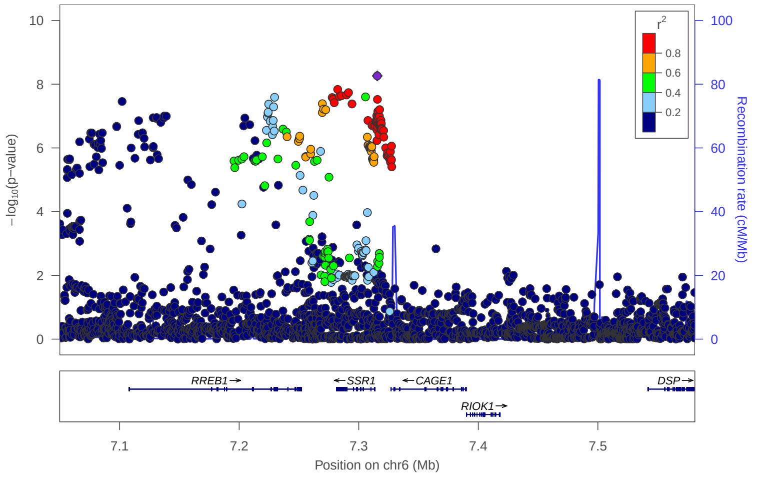


### locus 203 rs35056688

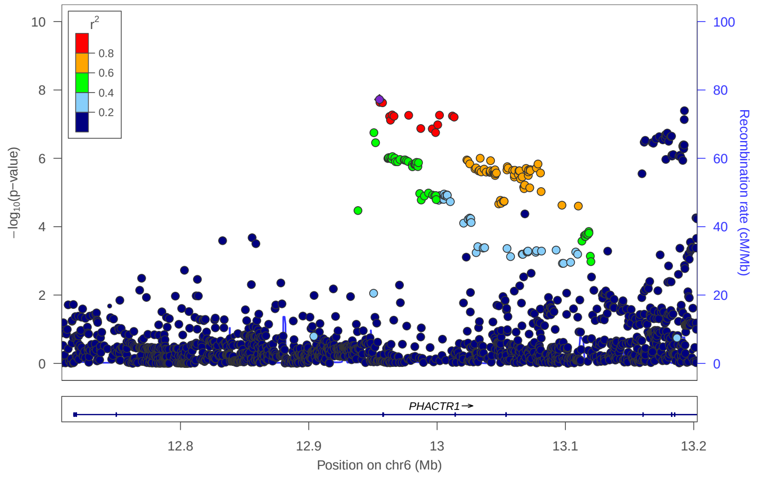


### locus 204 rs427597

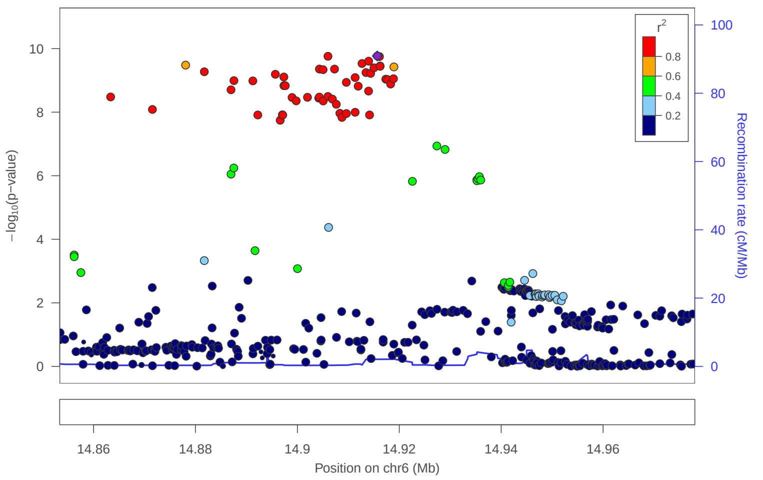


### locus 205 rs1397844

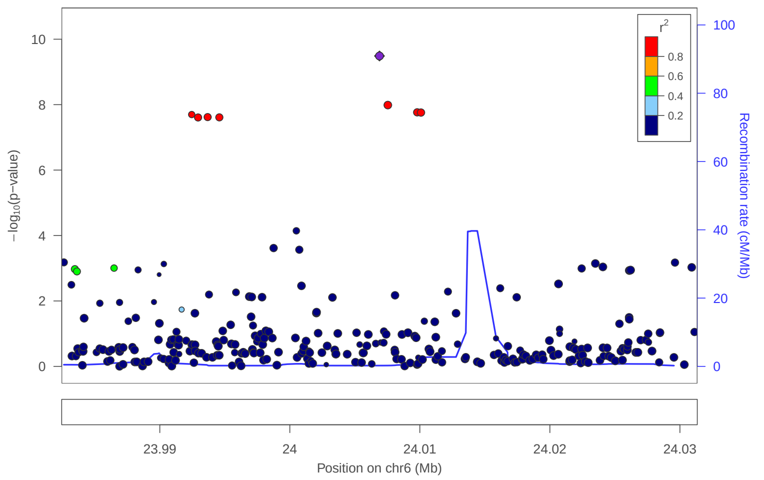


### locus 206 rs2066295

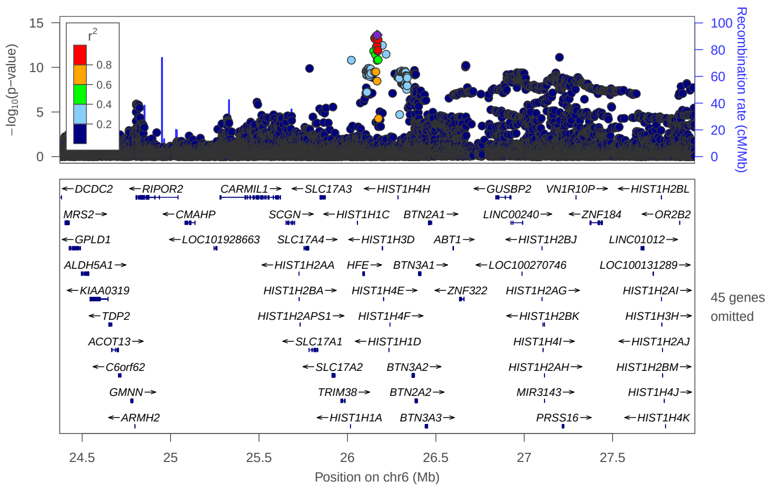


### locus 207 rs55777621

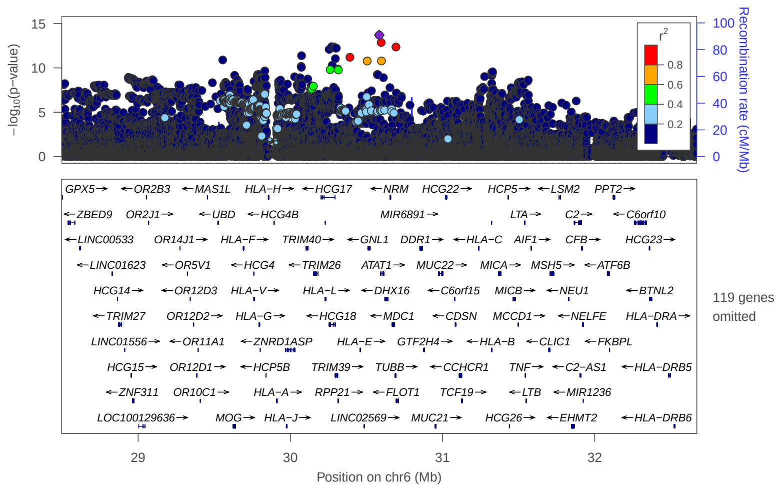


### locus 208 rs2523578

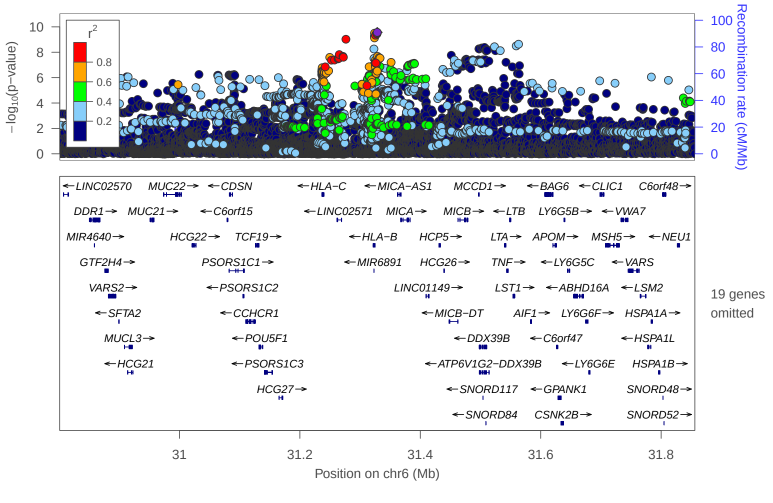


### locus 209 rs9271763

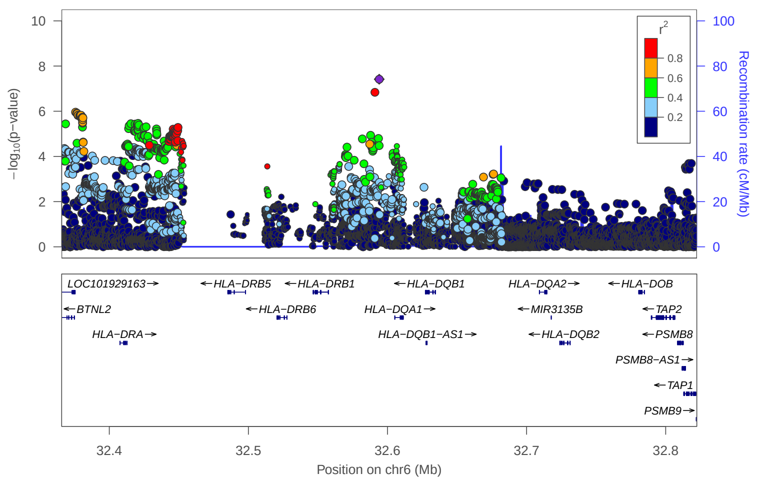

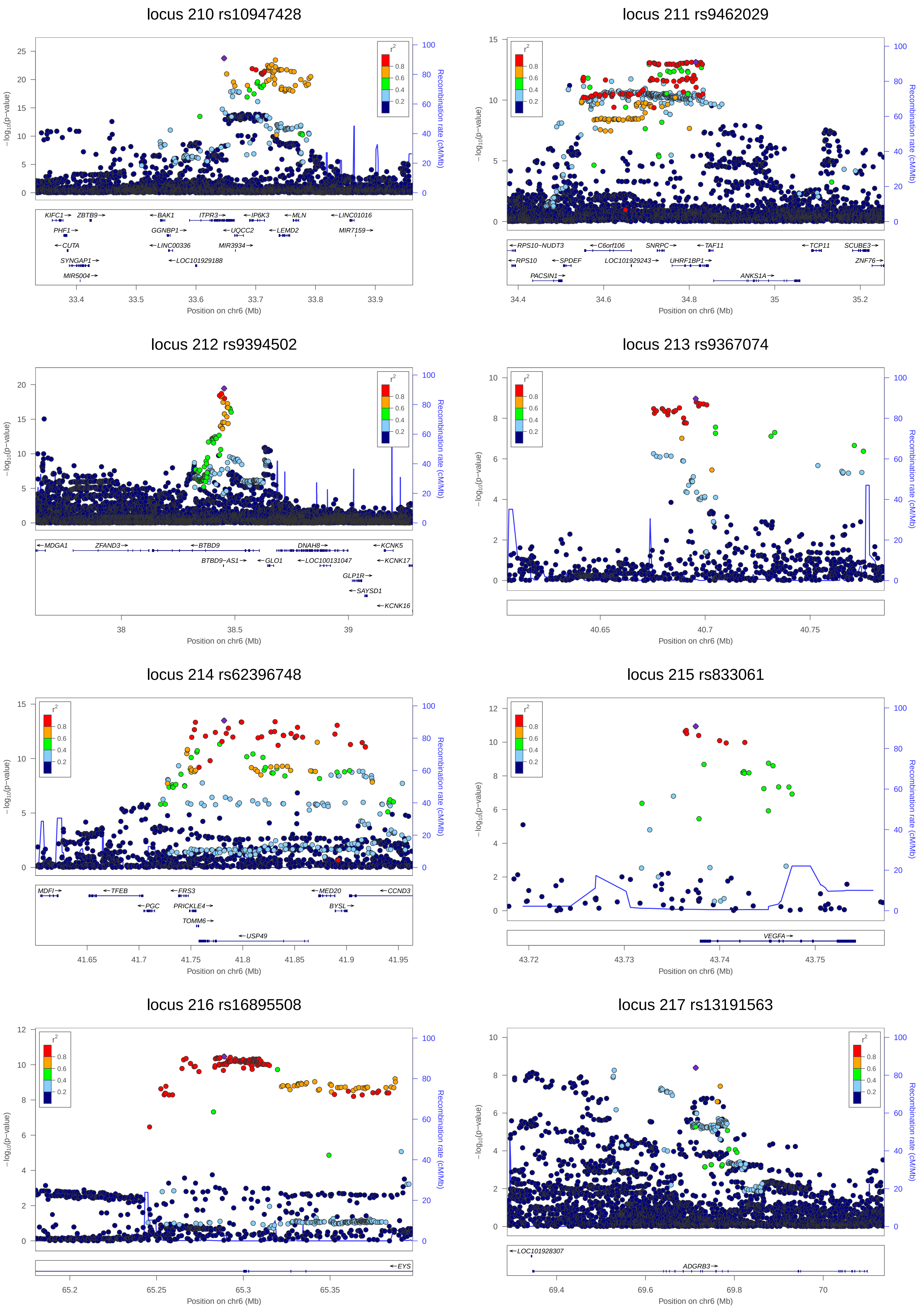

locus 210 rs10947428
KIFC1 ZBTB9 BAK1 ITPR3 IP6K3 MLN LINC01016
PHF1 GGNBP1 UQCC2 LEMD2 MIR7159
CUTA LINC00336 MIR3934
SYNGAP1 LOC101929188
MIR5004
Position on chr6 (Mb)
locus 211 rs9462029
RPS10-NUDT3 C6orf106 SNRPC TAF11 TCP11 SCUBE3
RPS10 SPDEF LOC101929243 UHRF1BP1 ZNF76
PACSIN1 ANKS1A
Position on chr6 (Mb)
locus 212 rs9394502
MDGA1 ZFAND3 BTBD9 DNAH8 KCNK5
BTBD9-AS1 GLO1 LOC100131047 KCNK17
GLP1R
SAYSD1
KCNK16
Position on chr6 (Mb)
locus 213 rs9367074
Position on chr6 (Mb)
locus 214 rs62396748
MDFI TFEB FRS3 MED20 CCND3
PGC PRICKLE4 BYSL
TOMM6
USP49
Position on chr6 (Mb)
locus 215 rs833061
VEGFA
Position on chr6 (Mb)
locus 216 rs16895508
EYS
Position on chr6 (Mb)
locus 217 rs13191563
LOC101928307
ADGRB3
Position on chr6 (Mb)
−log10(p−value)
Recombination rate (cM/Mb)
r2

locus 218 rs615526

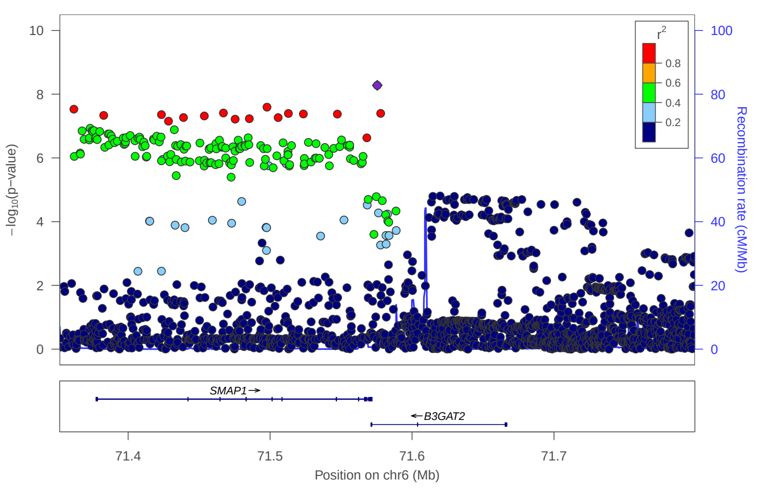


locus 219 rs1204329

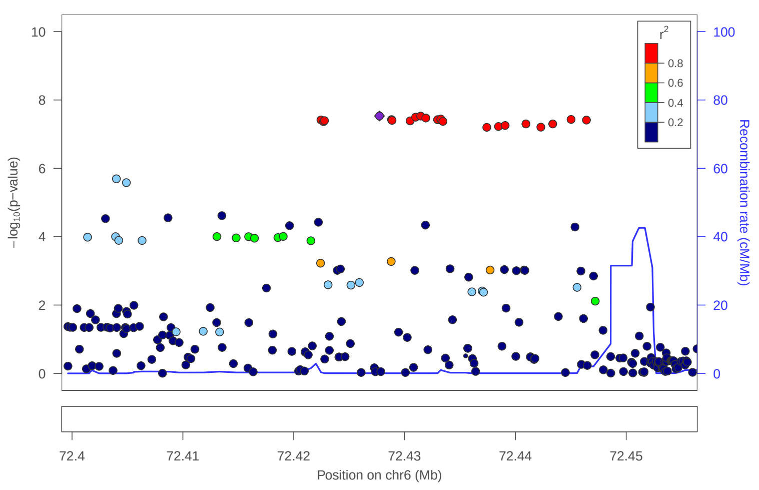


locus 220 rs9342993

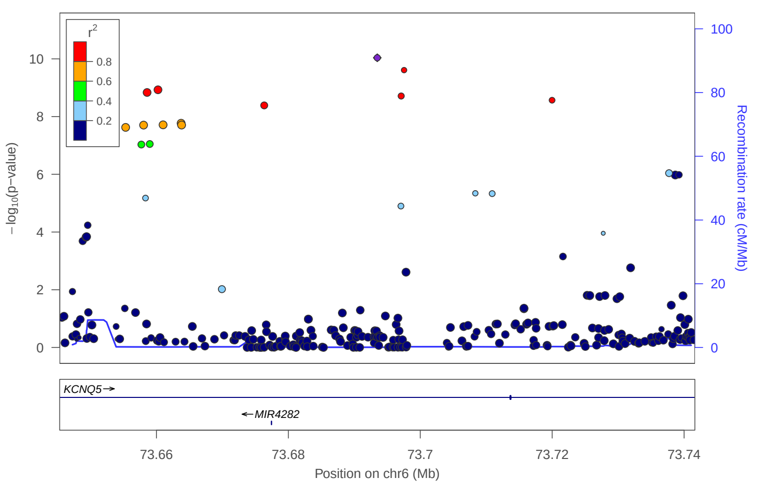


locus 221 rs16885957

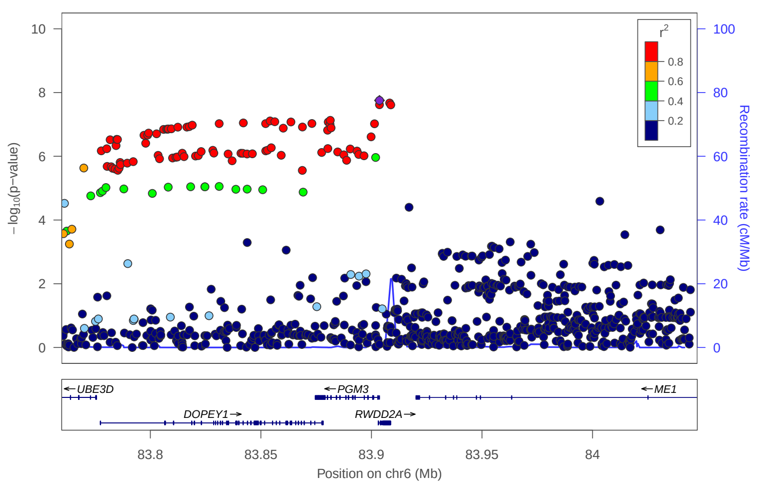


locus 222 rs62414180

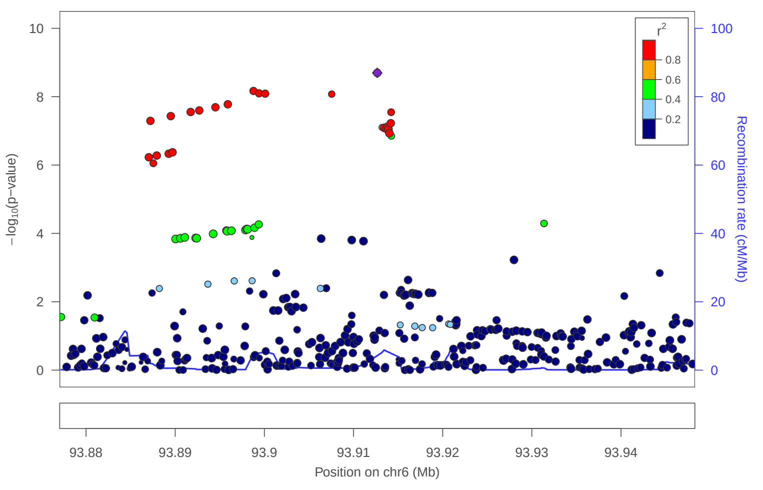


locus 223 rs9452444

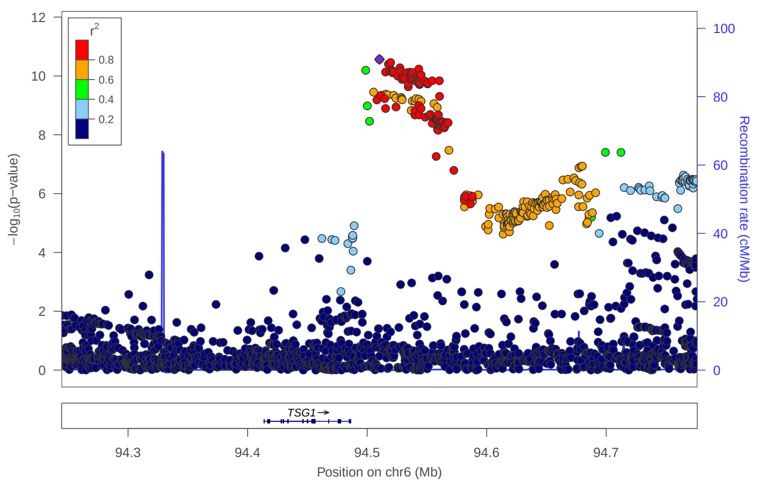


locus 224 rs9375945

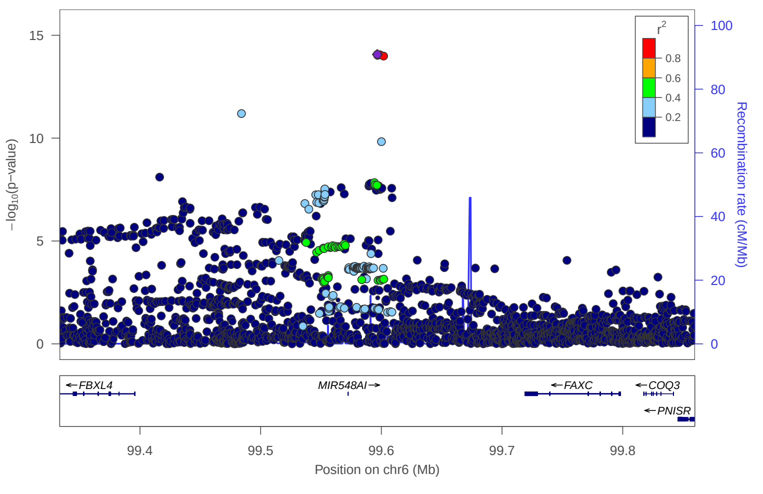


locus 225 rs240110

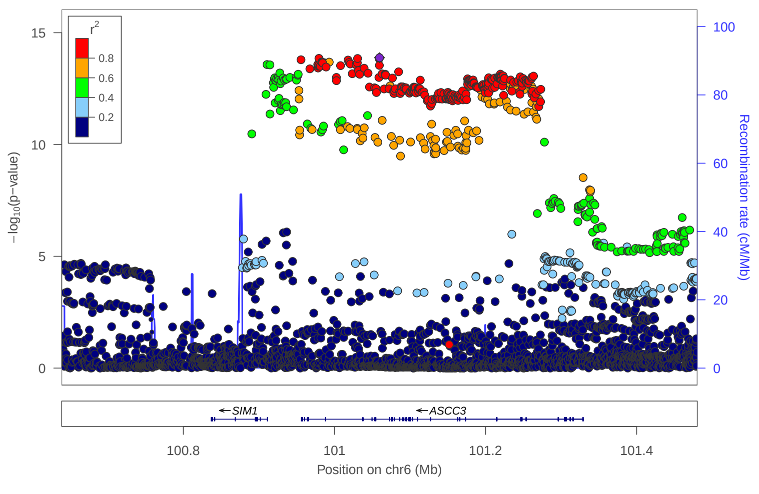

### locus 226 rs314281

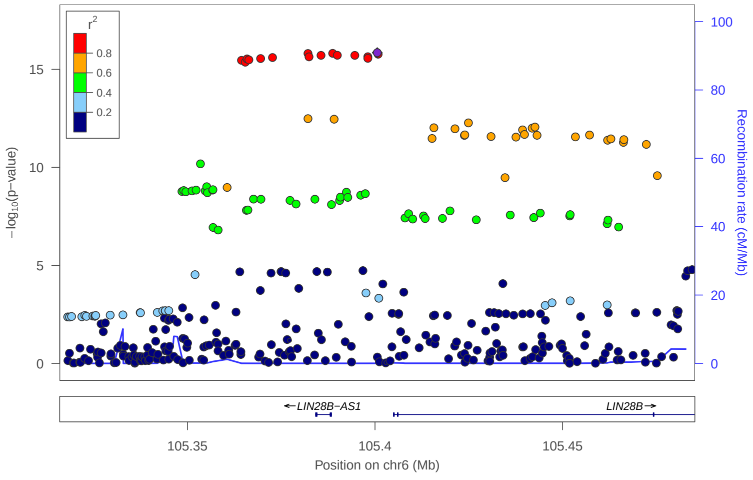


### locus 228 rs6149792

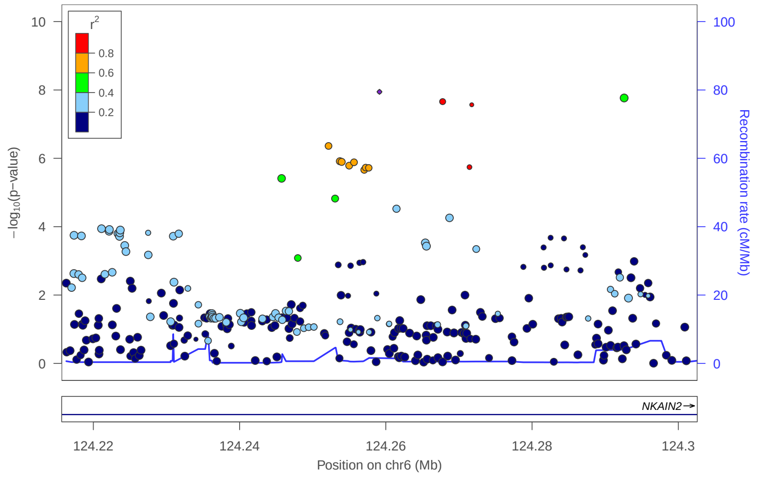


### locus 229 rs62429521

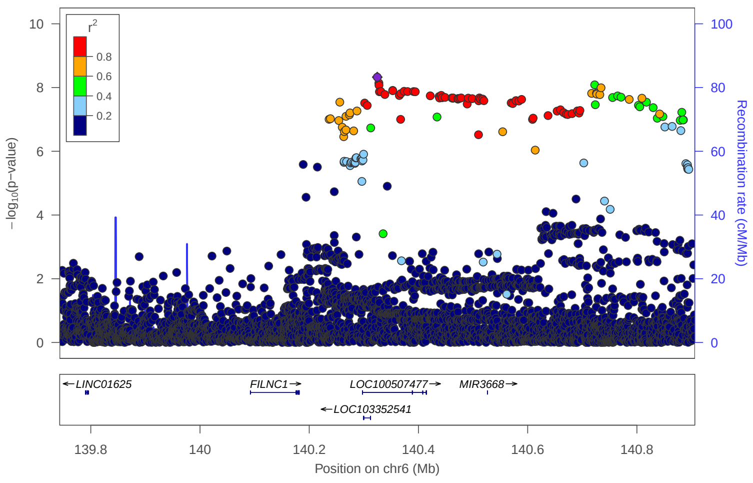


### locus 230 rs1789695

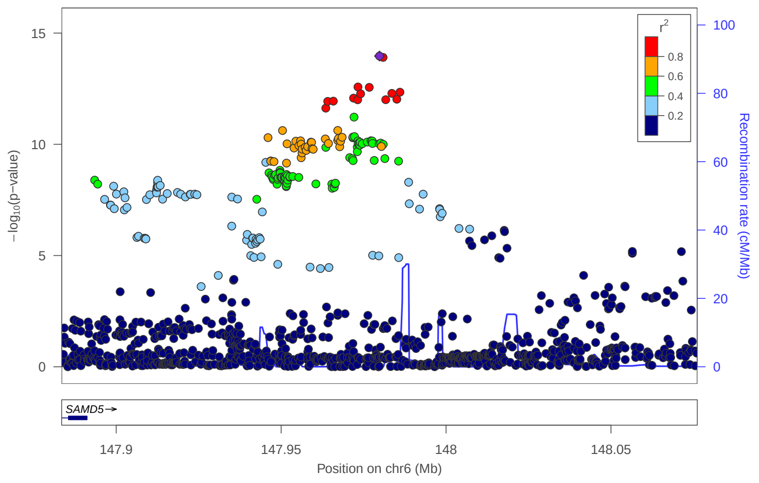


### locus 231 rs13210756

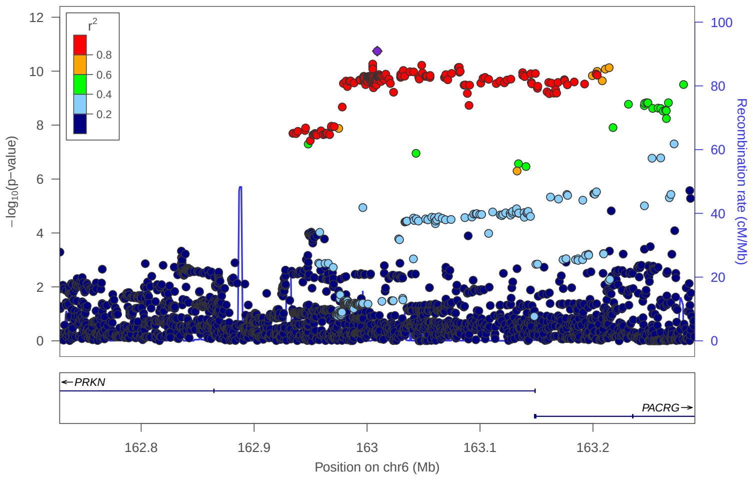


### locus 232 rs62444044

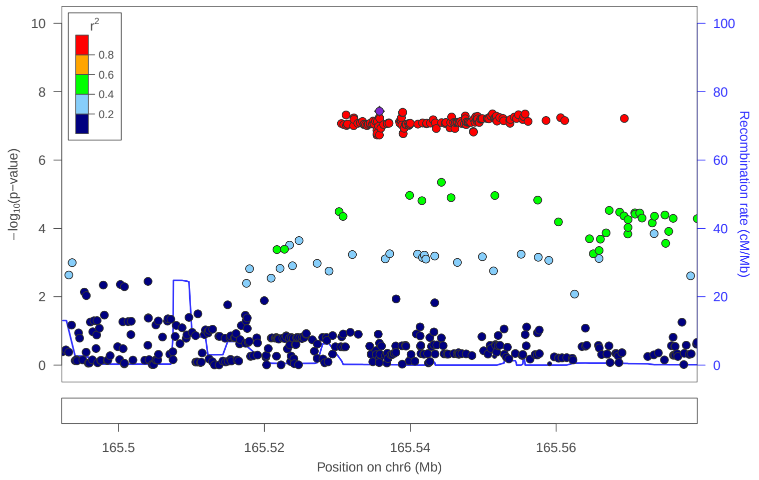


### locus 233 rs55790766

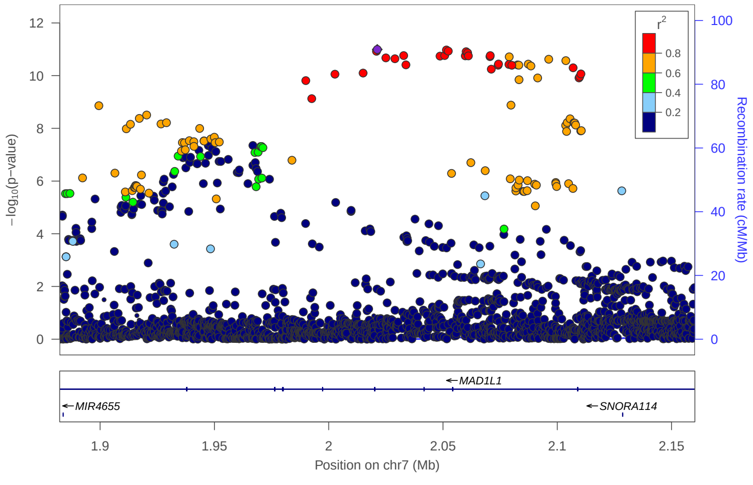


### locus 234 rs73033403

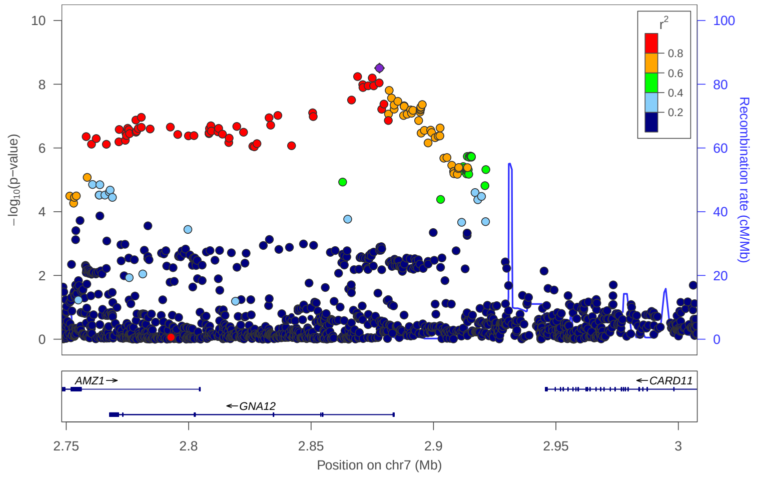

### locus 235 rs75218827

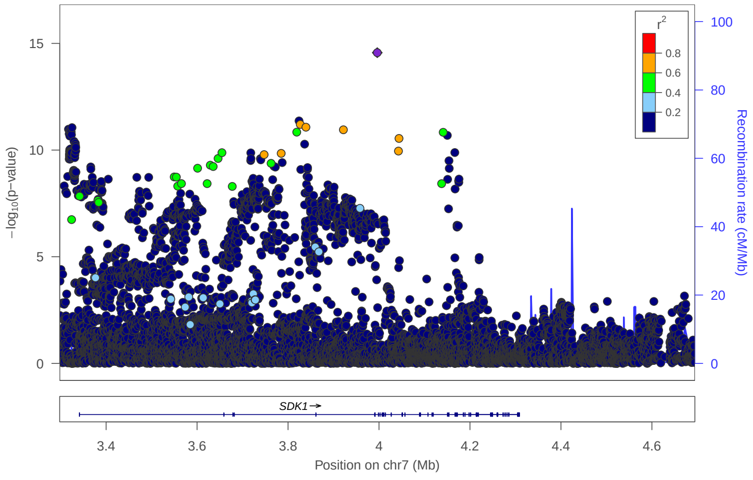


### locus 236 rs138713861

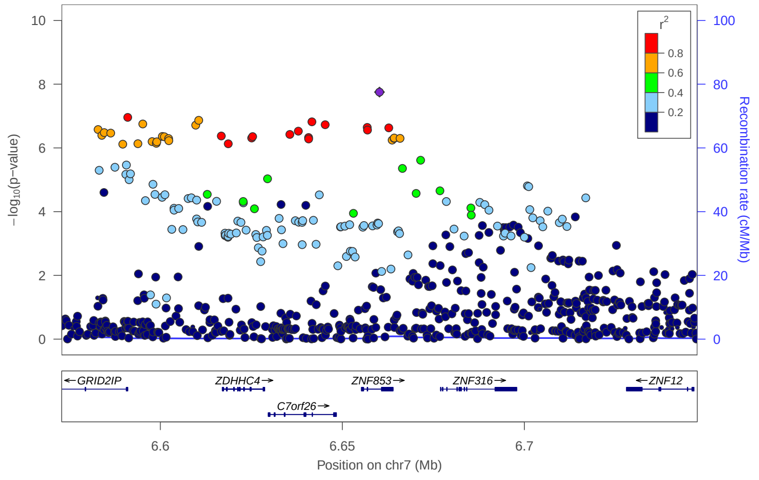


### locus 237 rs5010183

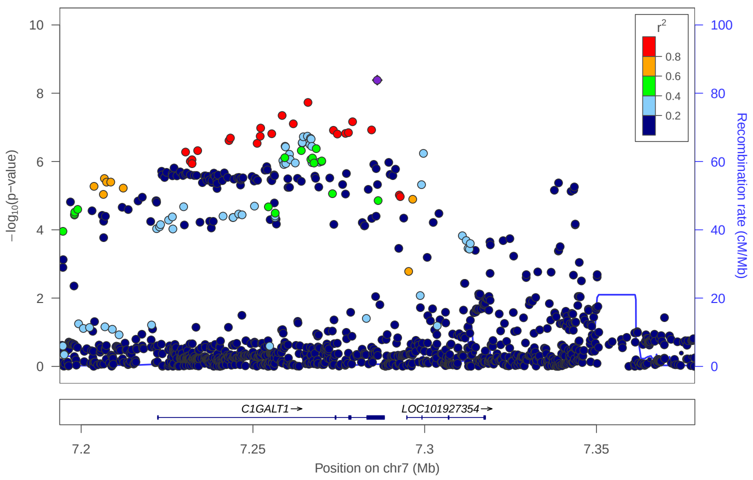


### locus 238 rs4721061

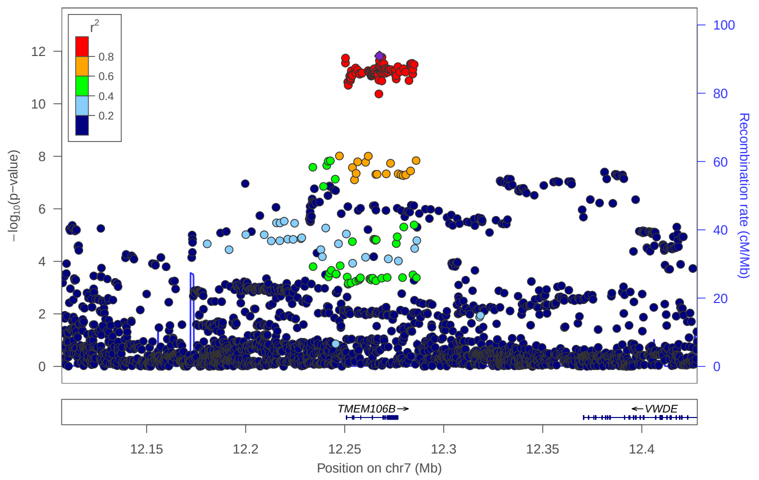


### locus 239 rs12333379

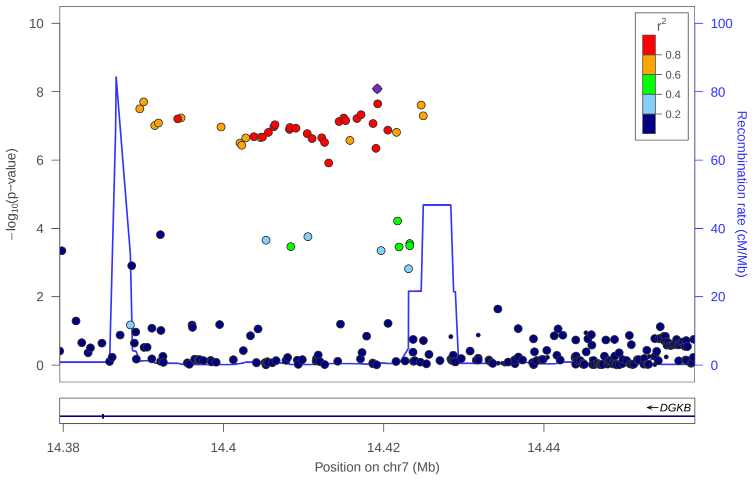


### locus 240 rs2158414

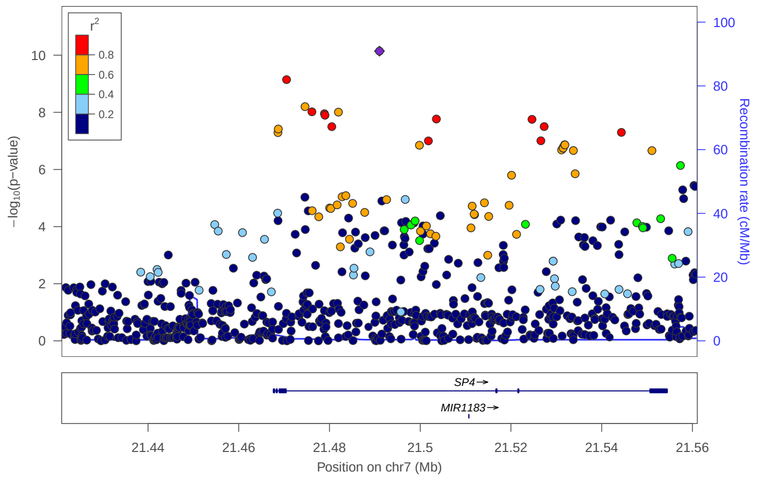


### locus 241 rs7805564

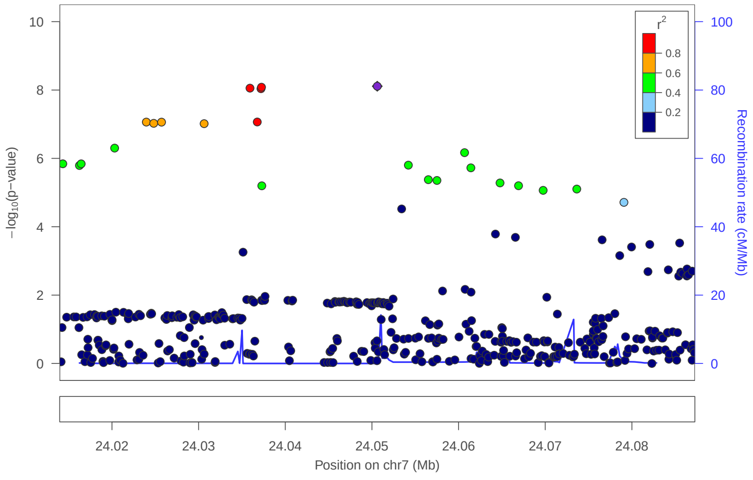


### locus 242 rs5883355

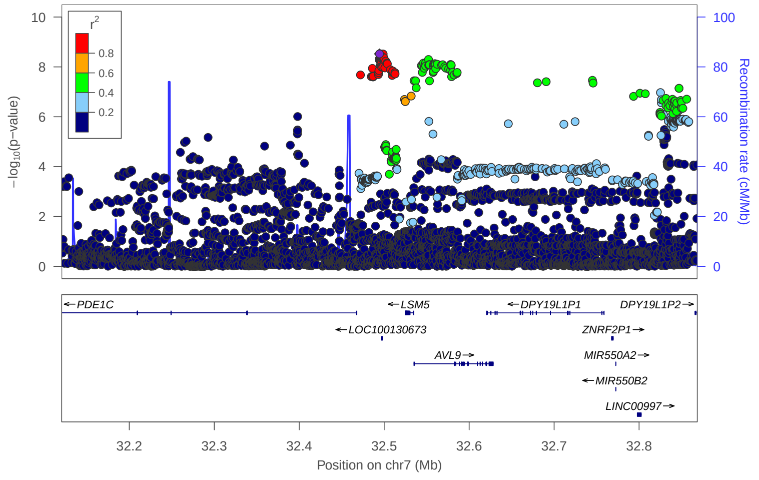

locus 243 rs62442231

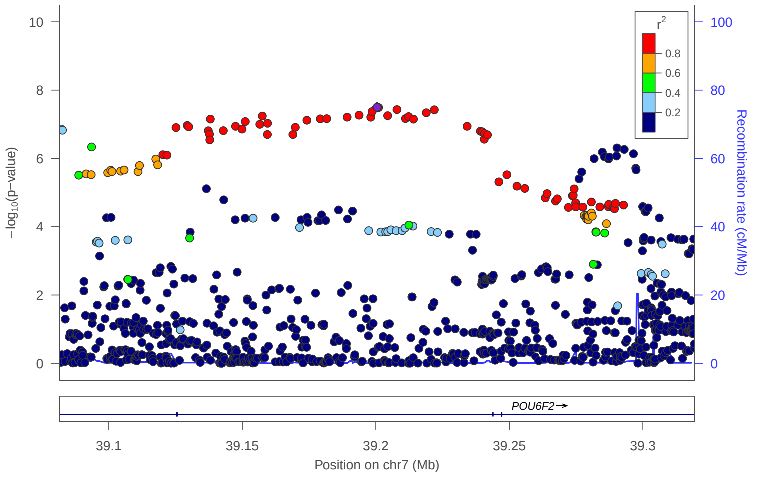


locus 244 rs10234070

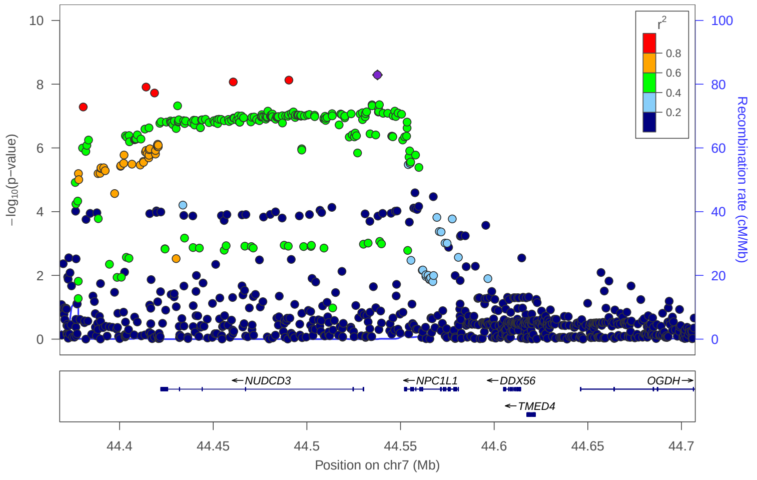


locus 245 rs73696927

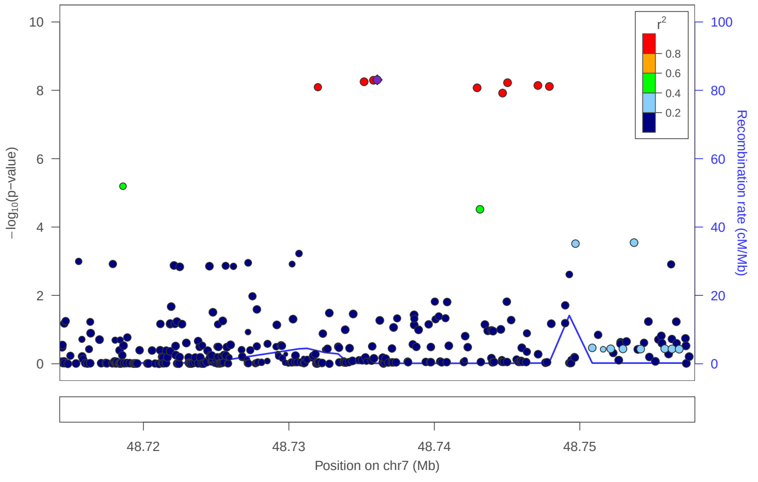


locus 246 rs11765062

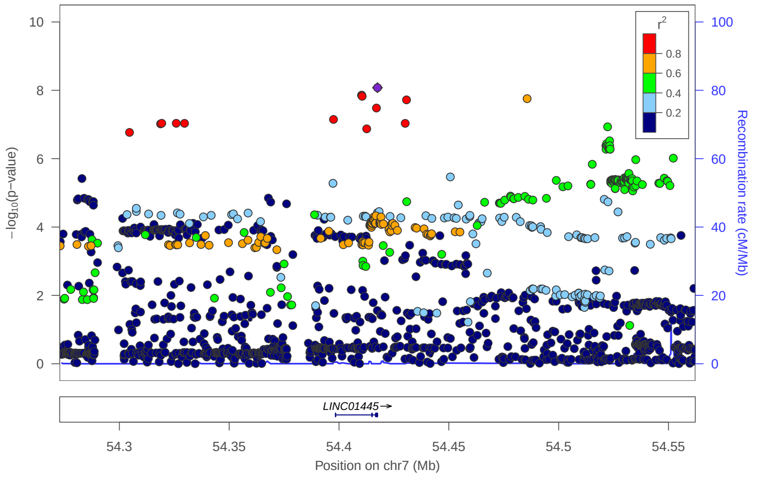


locus 247 rs60510343

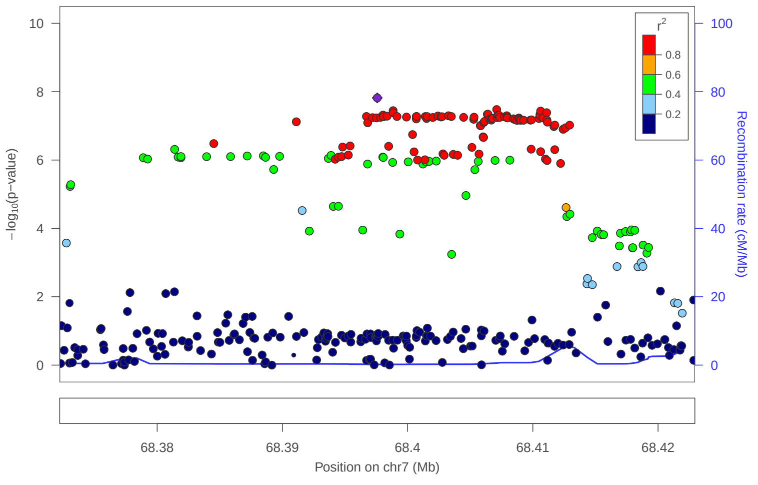


locus 248 rs11984380

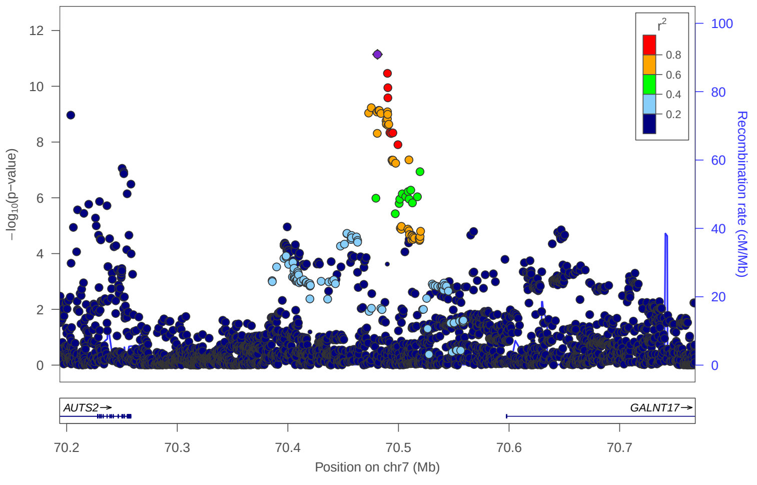


locus 249 rs35659126

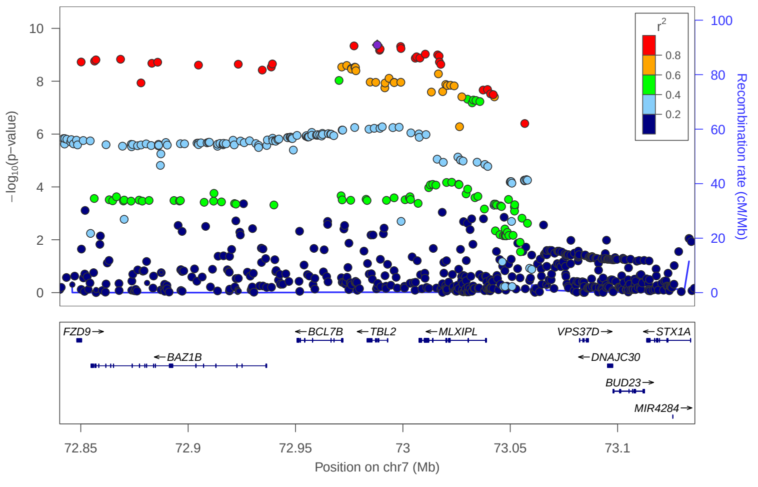


locus 250 rs1468774

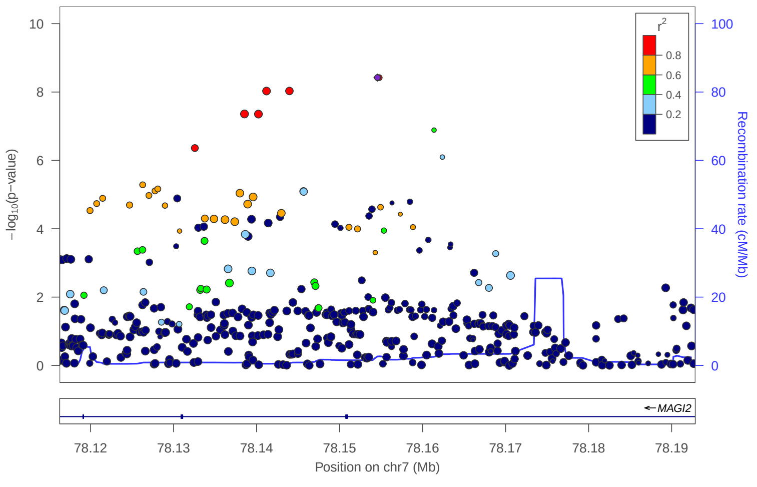

### locus 251 rs10269321

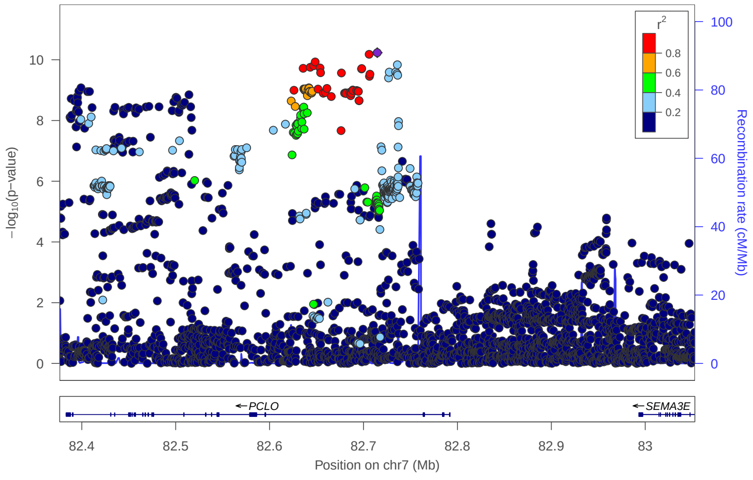


### locus 252 rs11980428

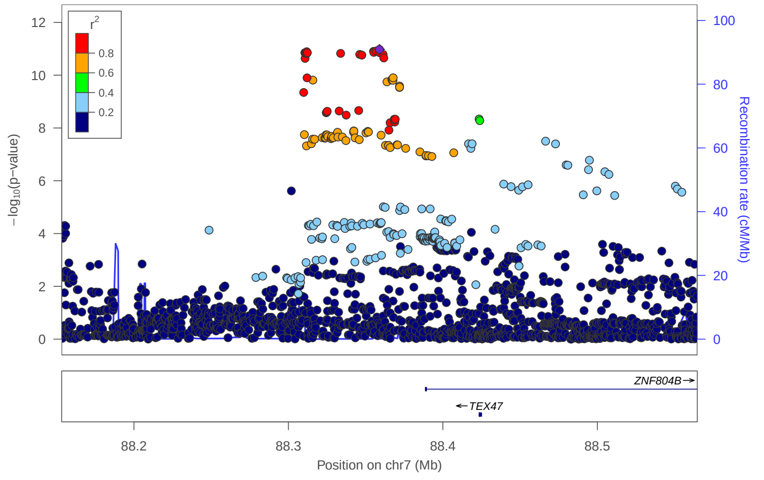


### locus 253 rs6965799

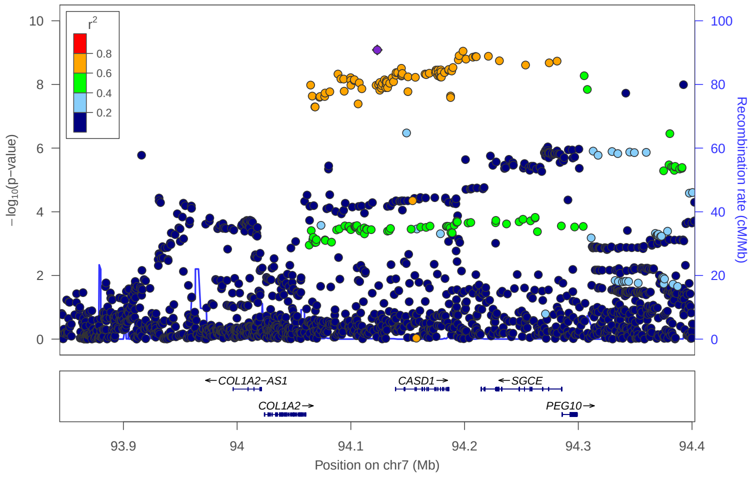


### locus 254 rs66526712

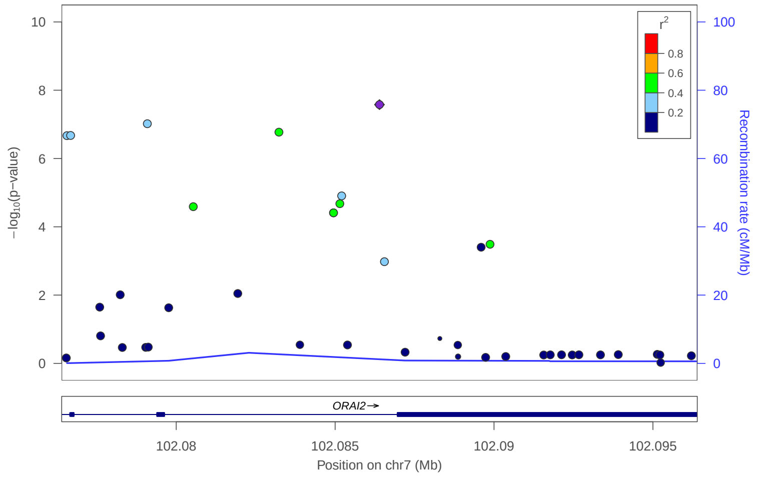


### locus 255 rs6949391

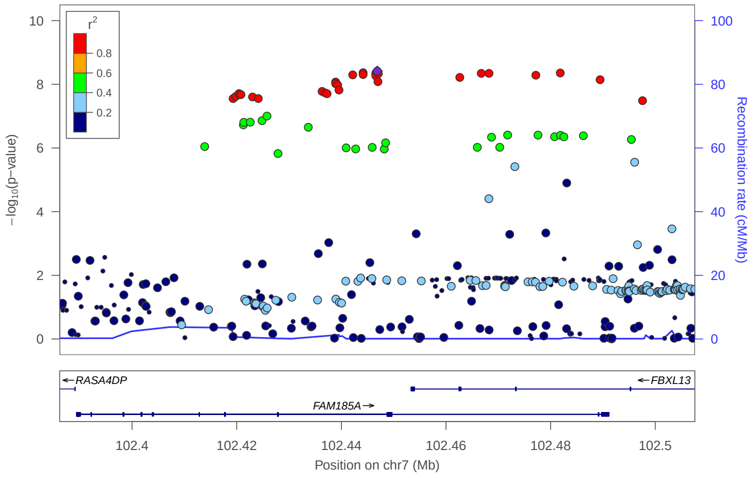


### locus 256 rs112583482

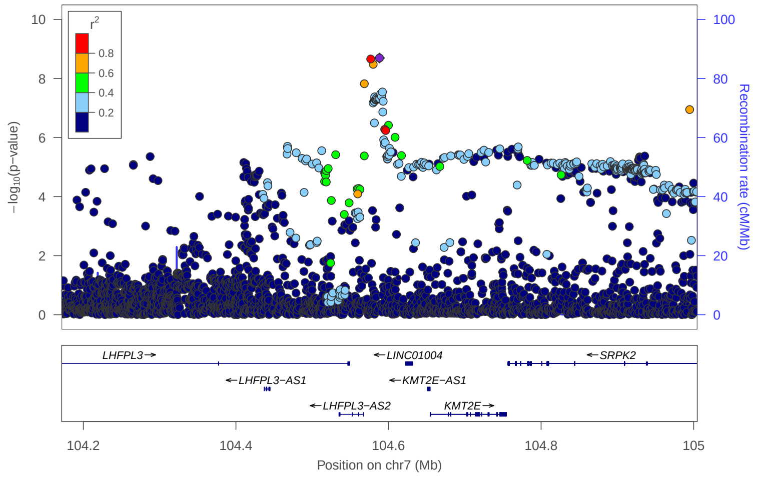


### locus 257 rs76924513

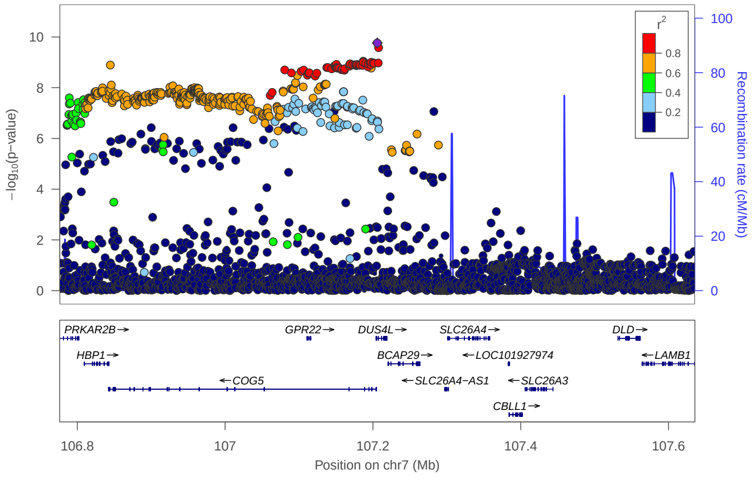


### locus 258 rs1513922

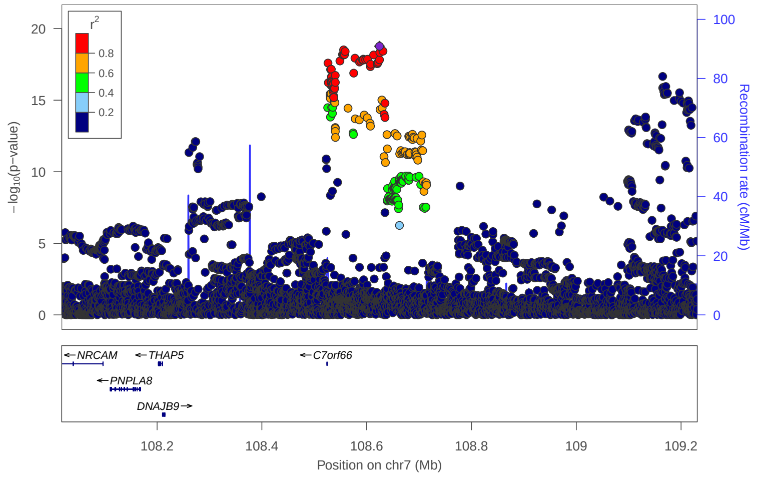

locus 259 rs2528691

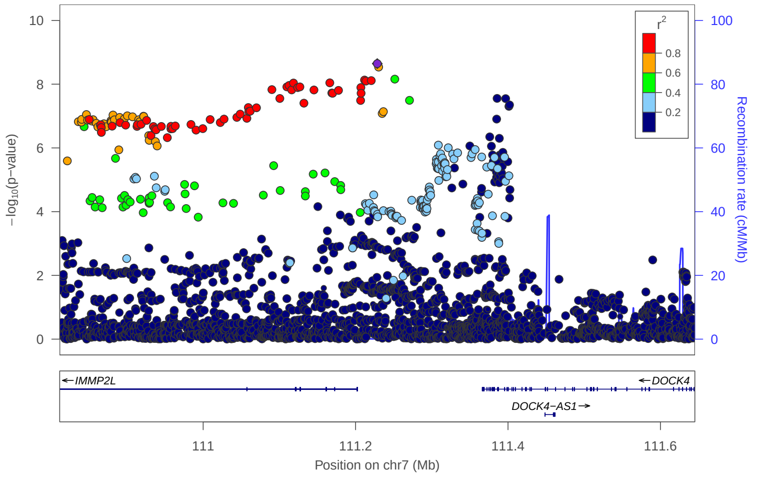


locus 260 rs8180817

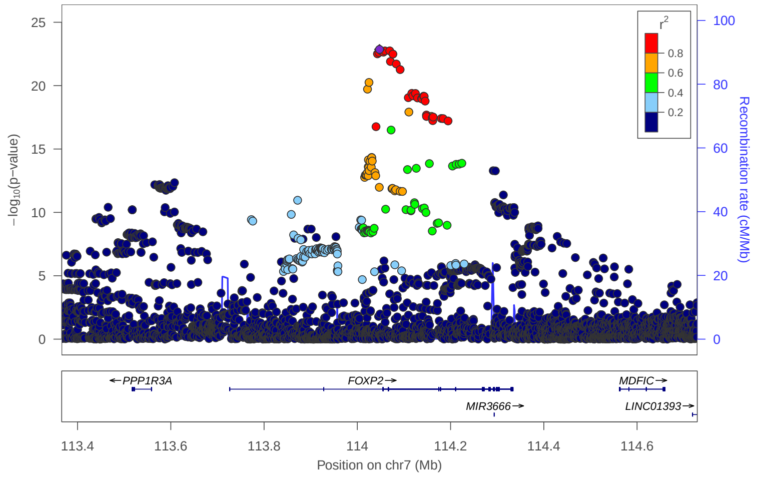


locus 261 rs1358393

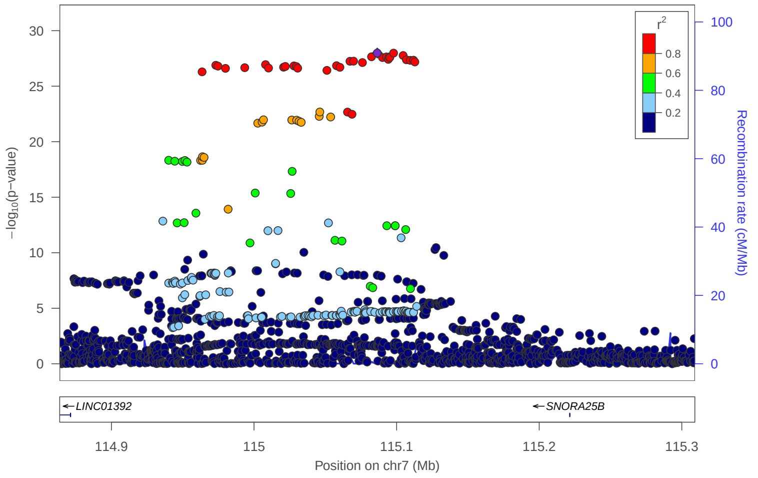


locus 262 rs35590186

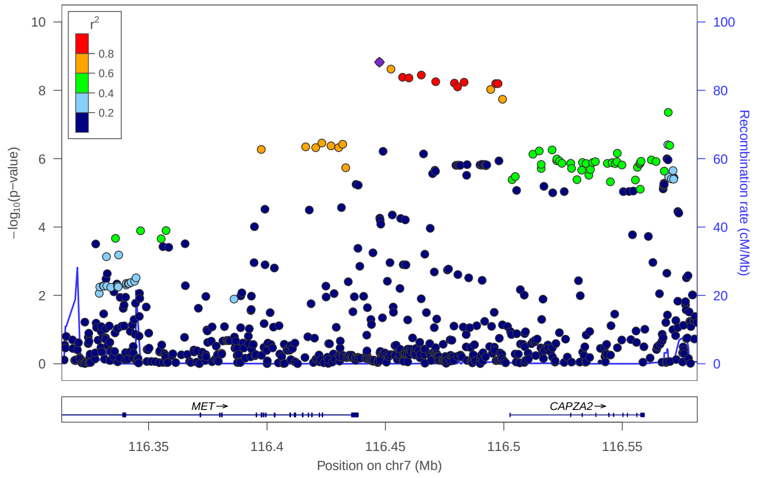


locus 263 rs1156954

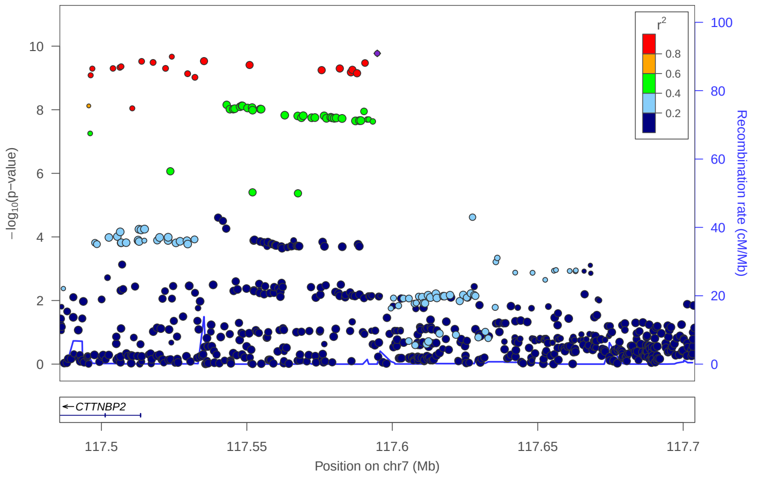


locus 264 rs62472321

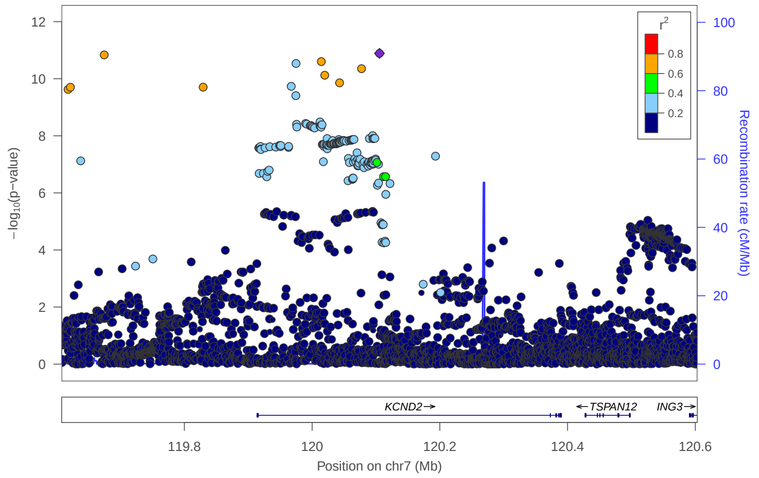


locus 265 rs7778413

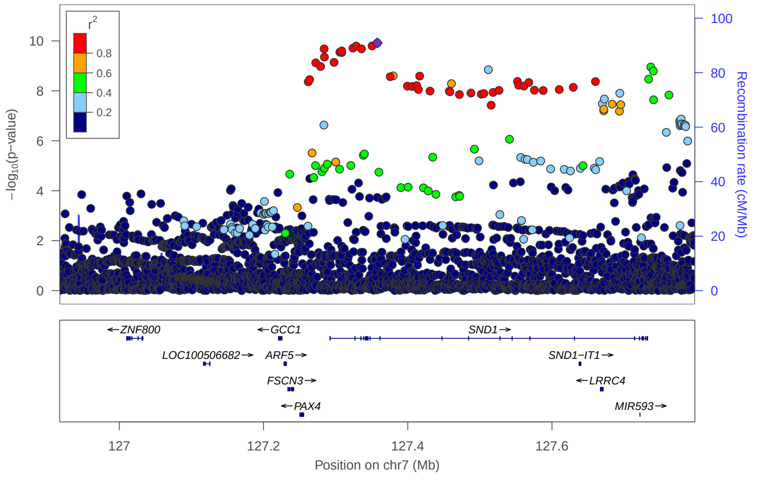


locus 266 rs56182580

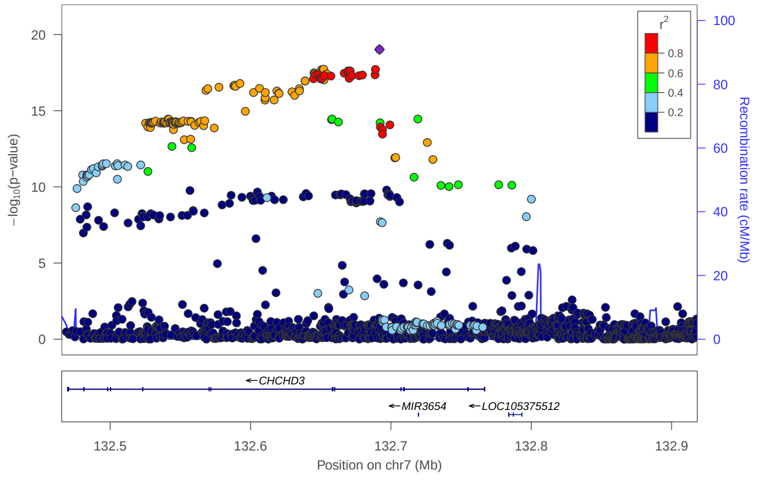

### locus 267 rs2544211

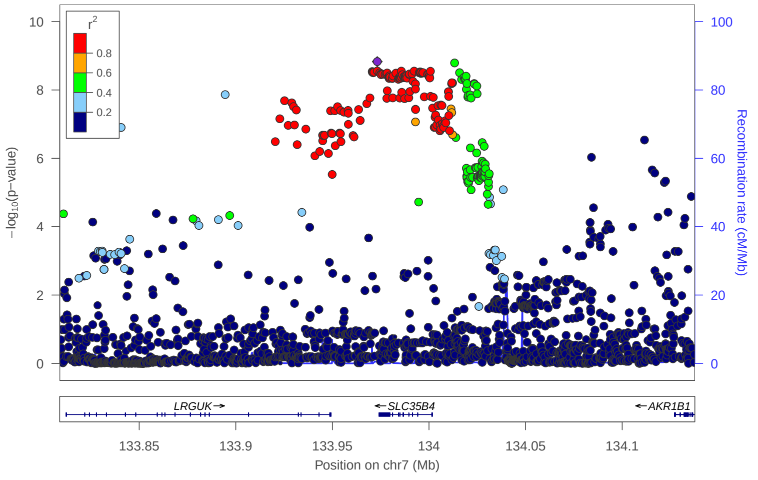


### locus 268 rs1731951

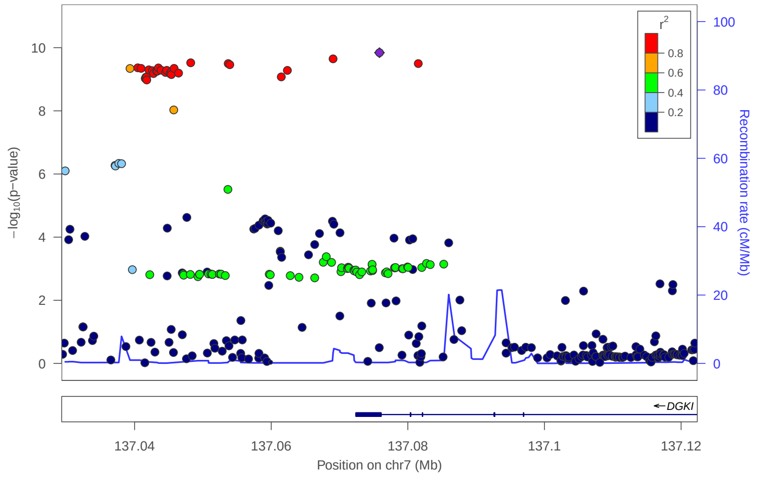


### locus 269 rs13269117

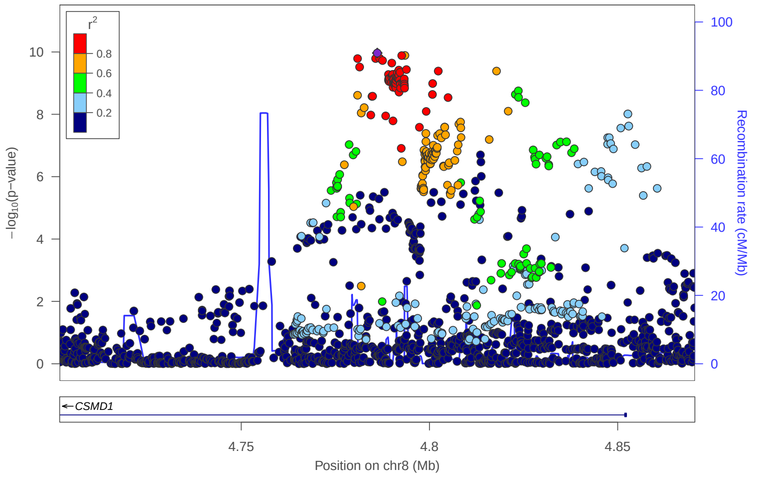


### locus 270 rs6601426

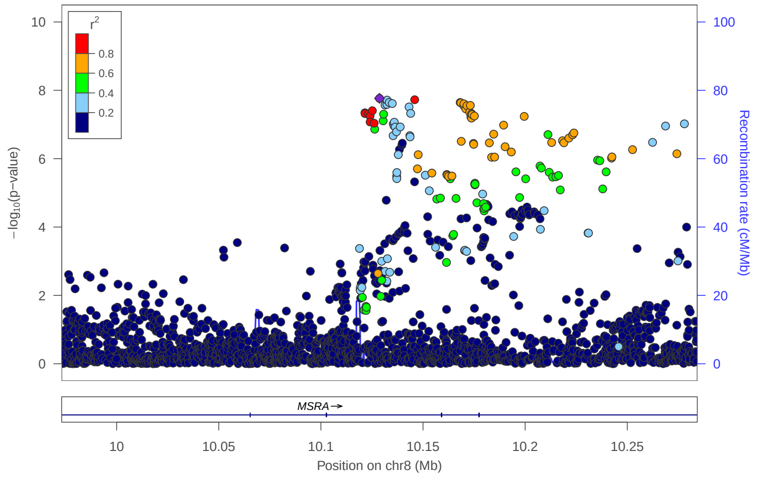


### locus 271 rs4831647

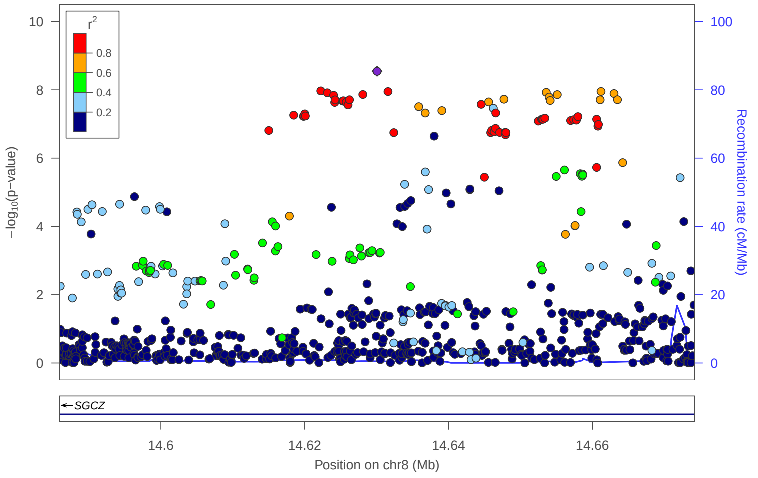


### locus 272 rs2616192

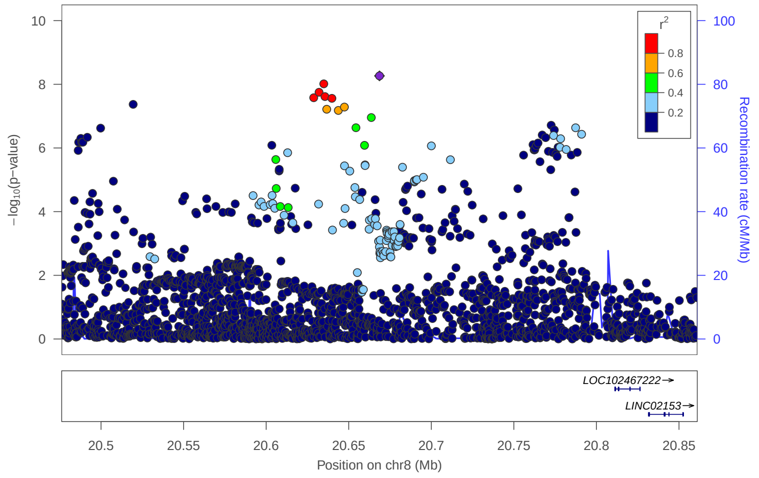


### locus 273 rs751680

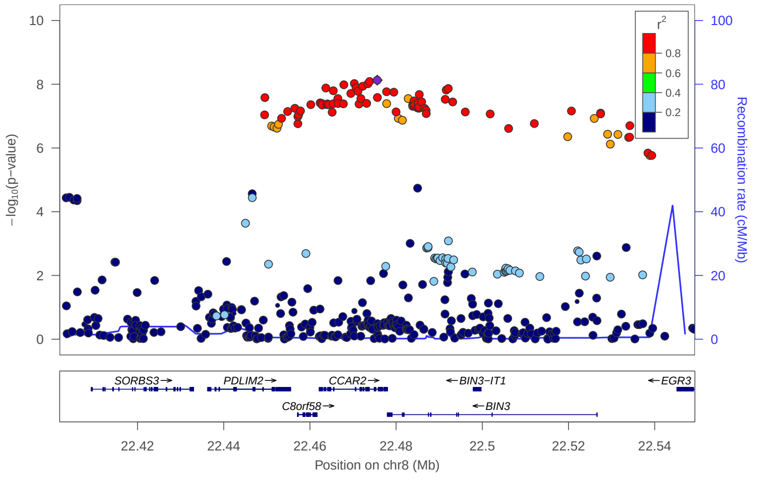


### locus 274 rs62501777

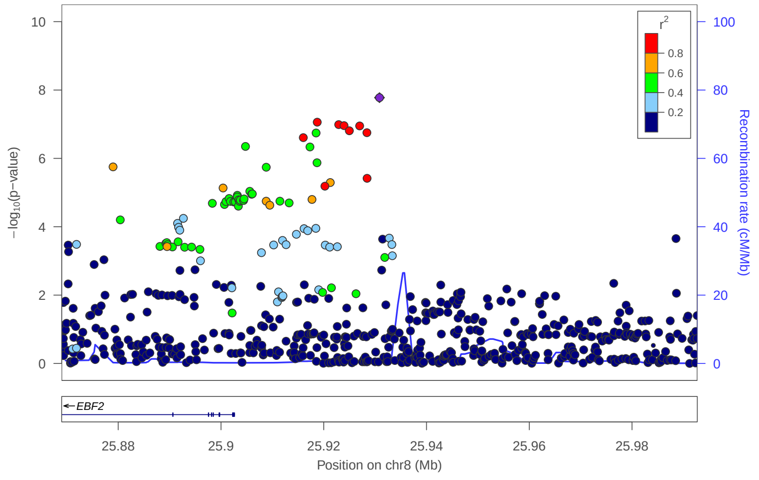

locus 275 rs10110224

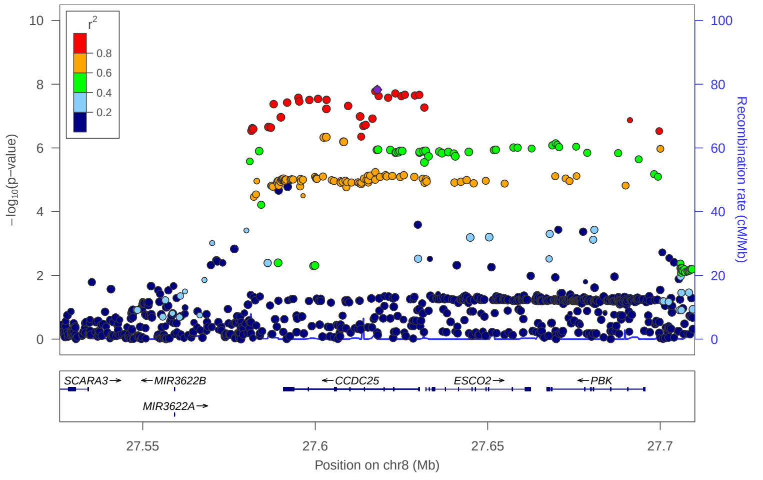


locus 276 rs2344121

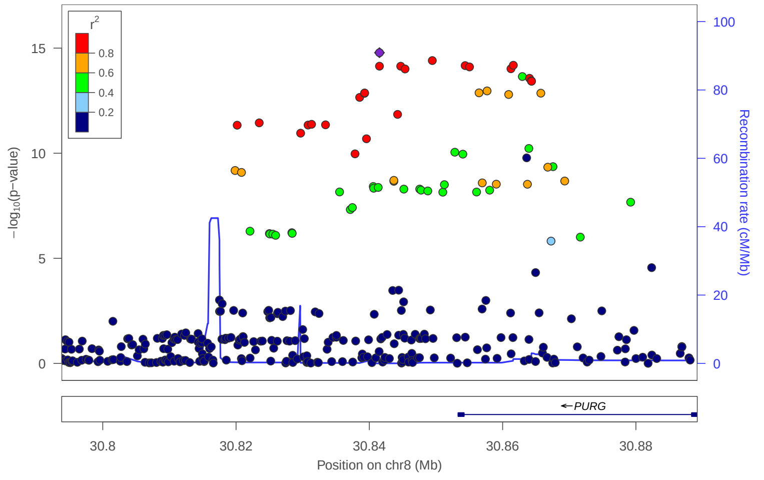


locus 277 rs1383890

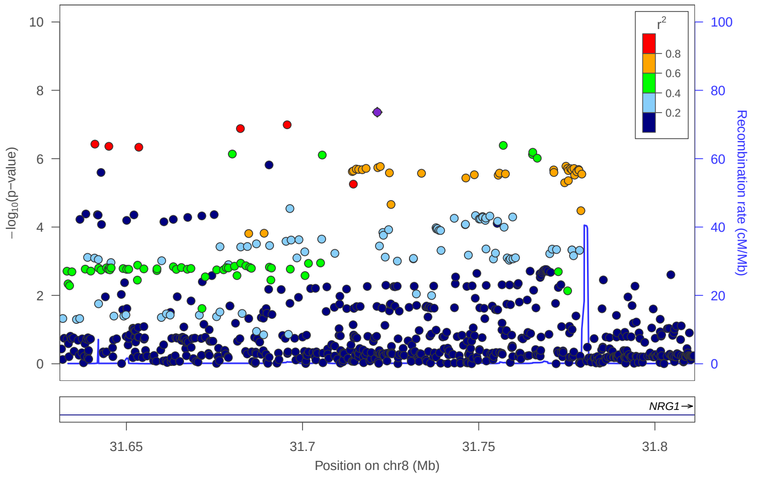


locus 278 rs79050075

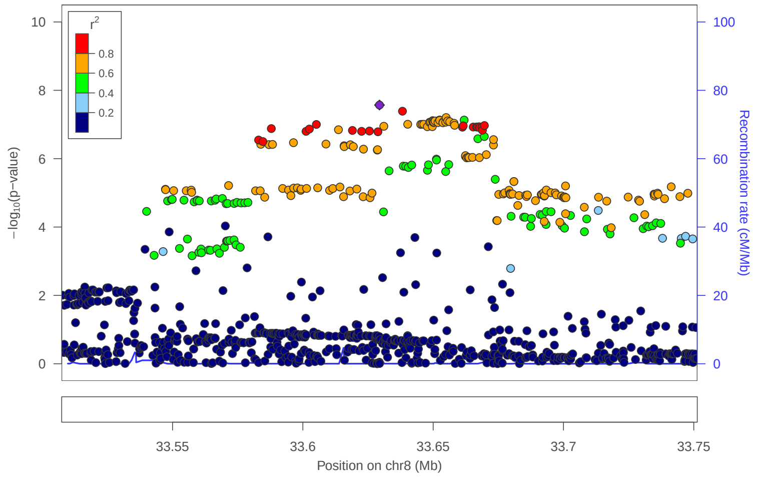


locus 279 rs573937698

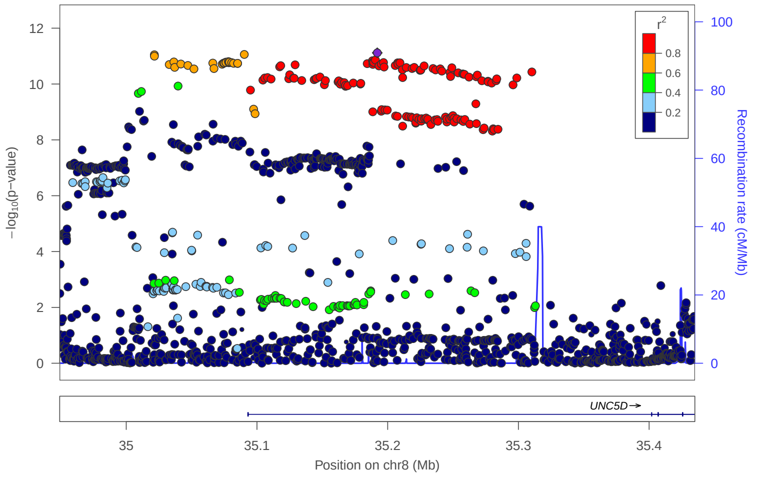


locus 280 rs5891160

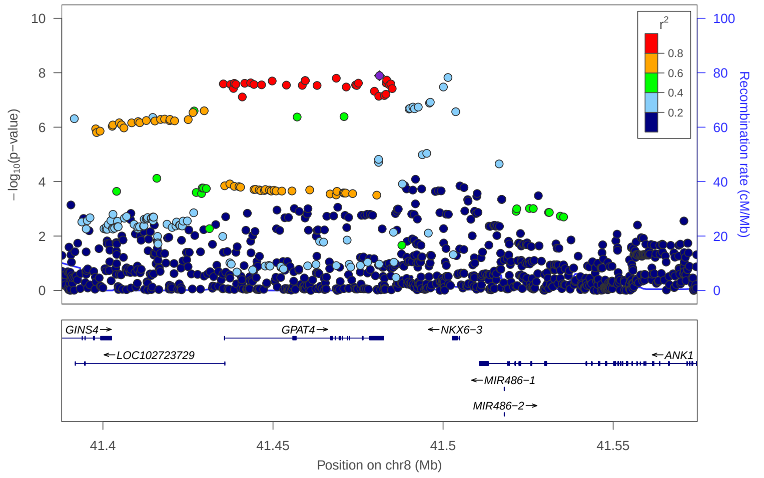


locus 281 rs12677105

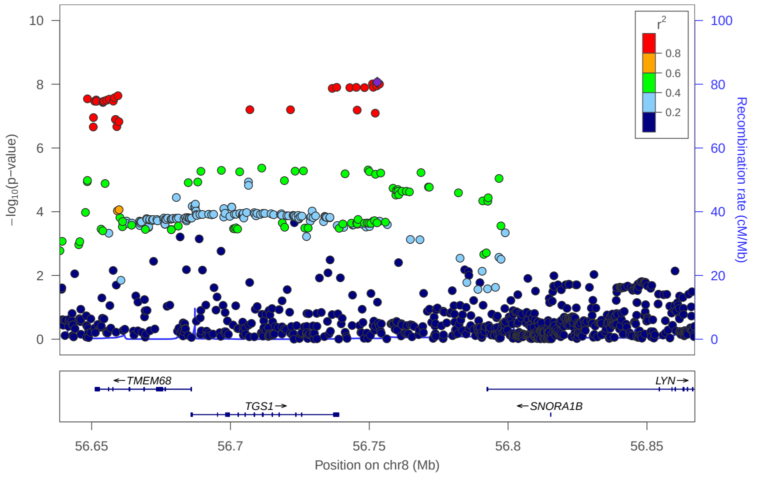


locus 282 rs10092649

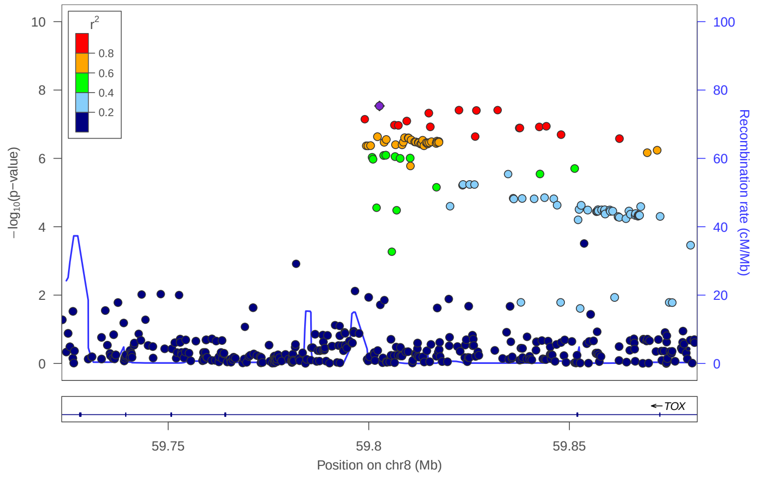

### locus 283 rs643730

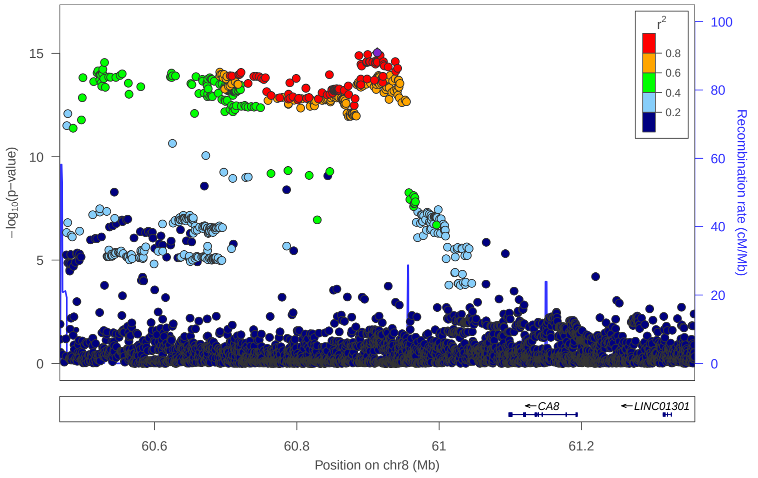


### locus 284 rs77753524

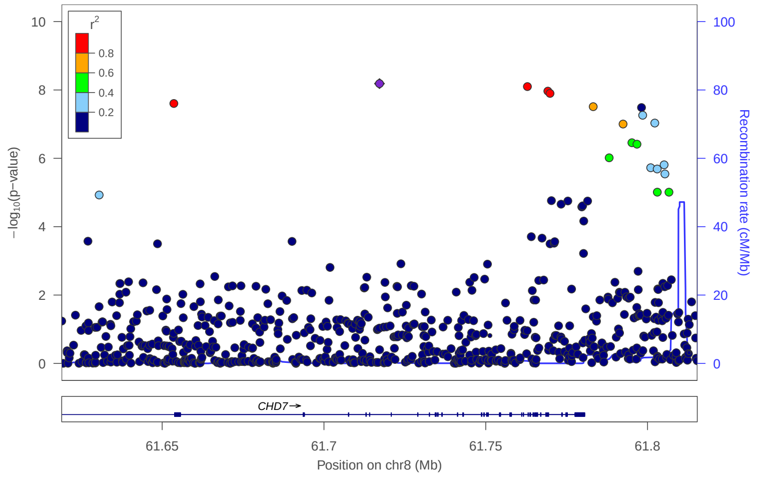


### locus 285 rs10957292

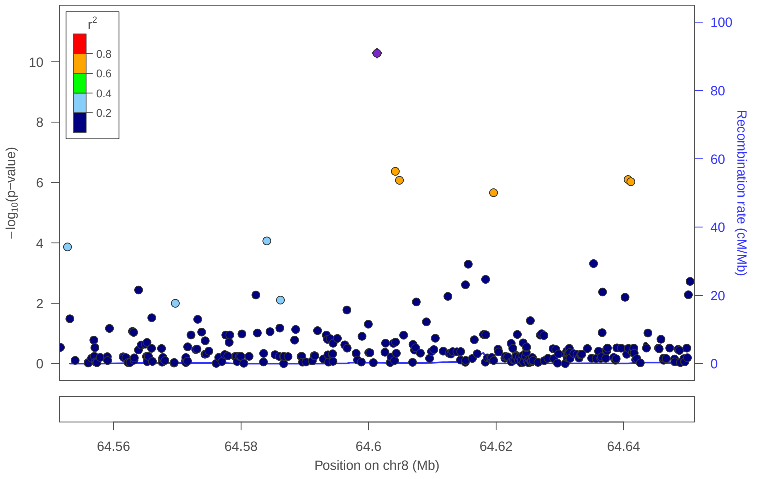


### locus 286 rs298195

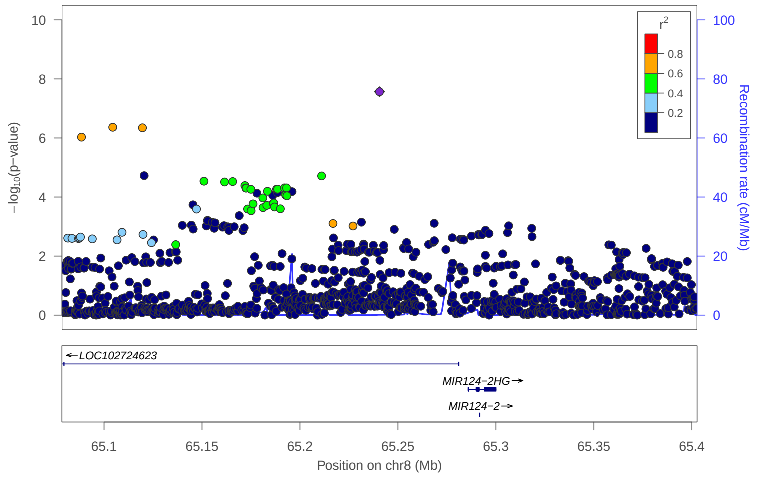


### locus 287 rs10104523

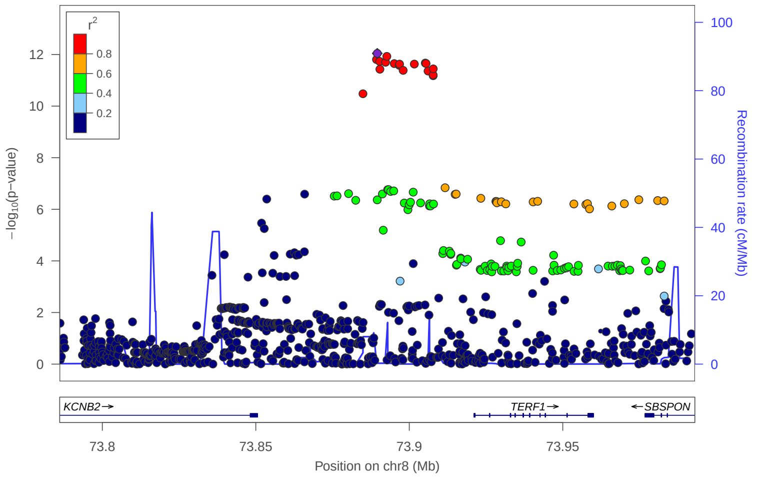


### locus 288 rs35862289

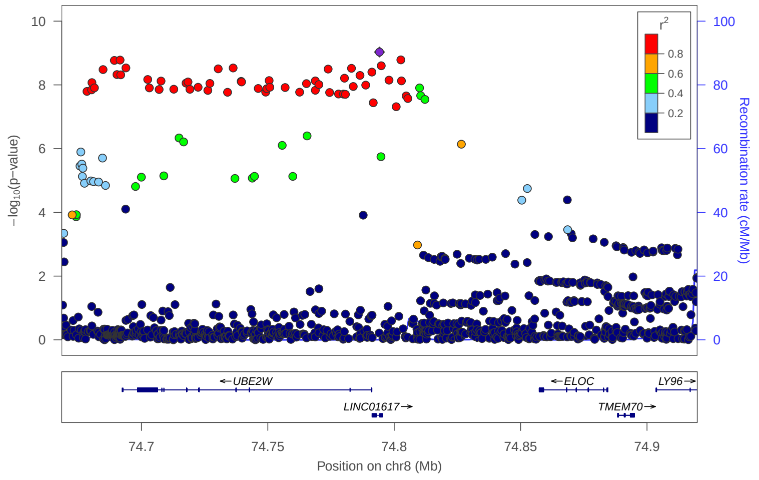


### locus 289 rs1070036

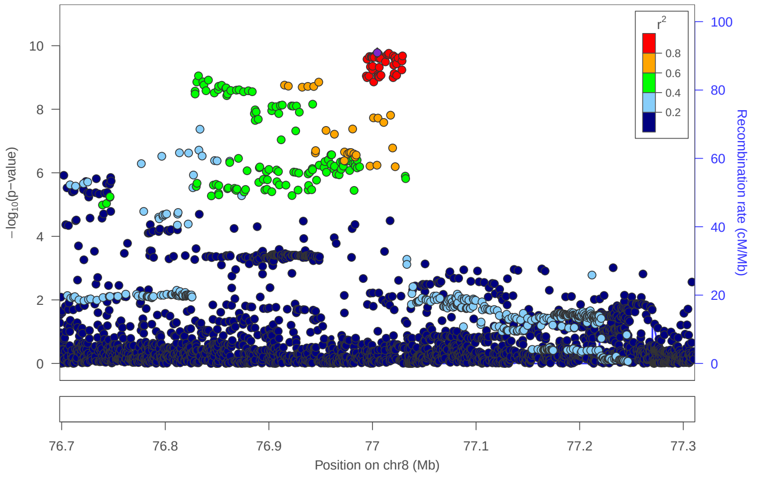


### locus 290 rs4735738

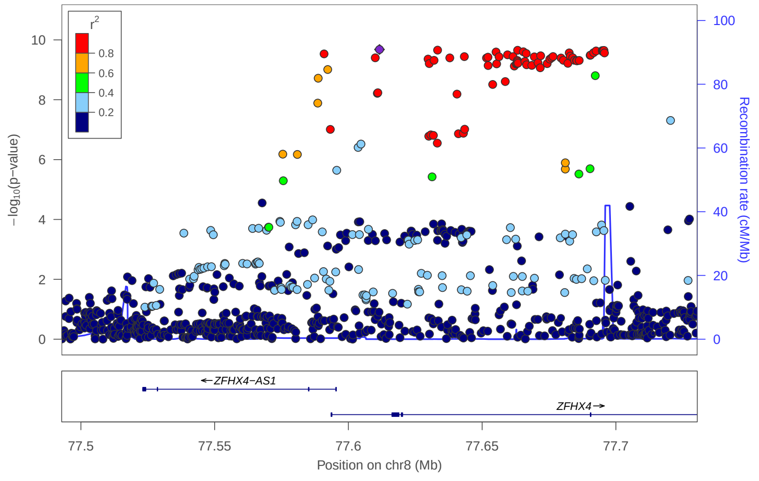

locus 291 rs4409363

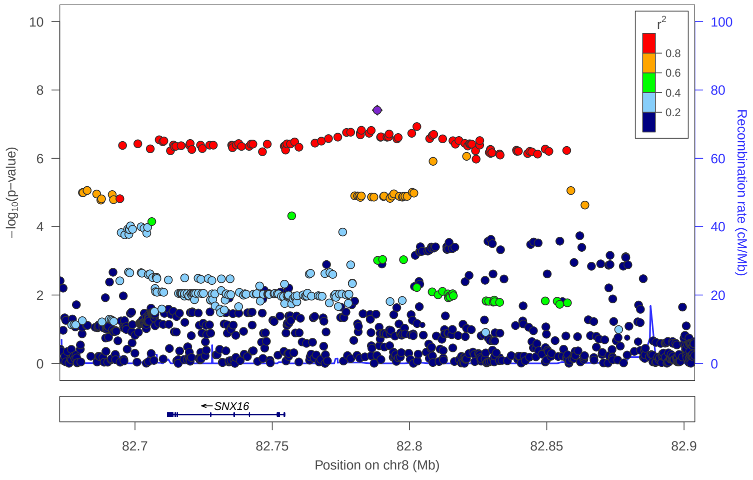


locus 292 rs62530187

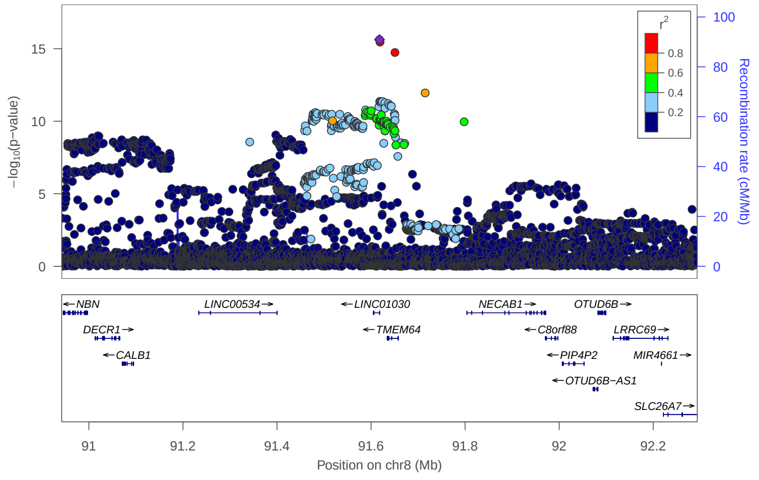


locus 293 rs7844069

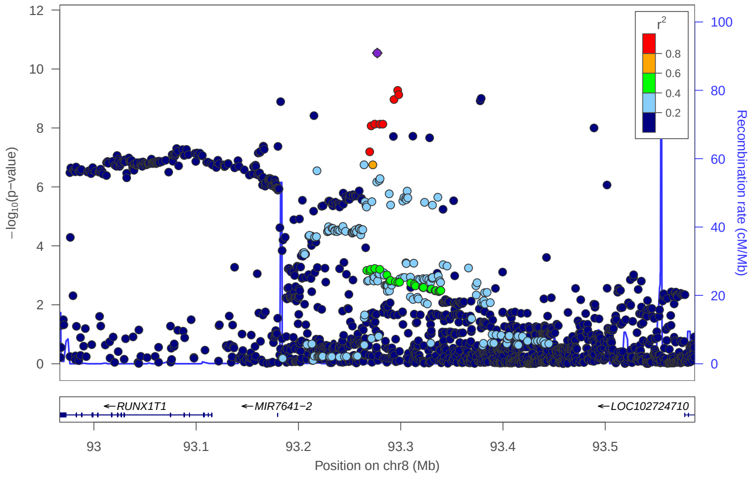


locus 294 rs56402193

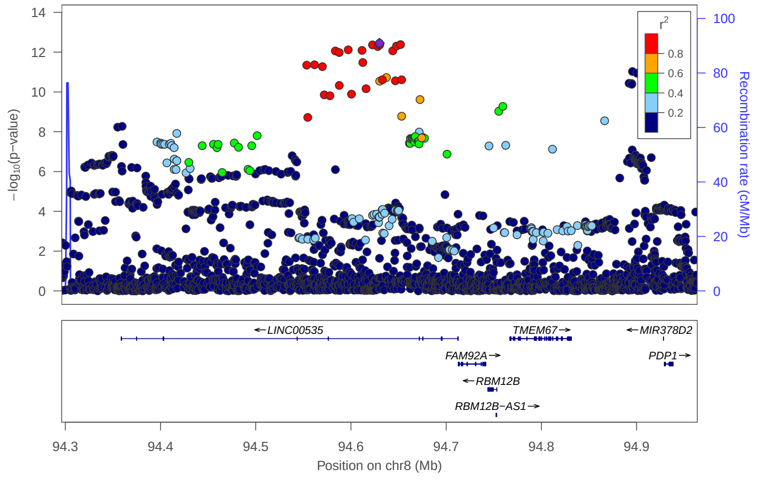


locus 295 rs11412375

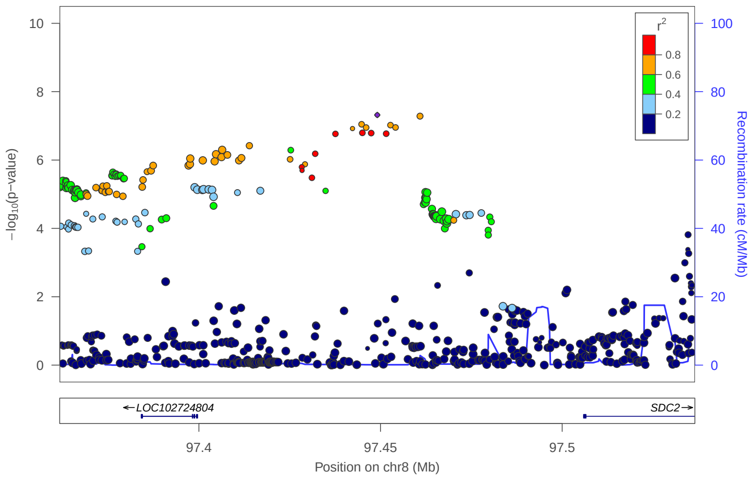


locus 296 rs28552587

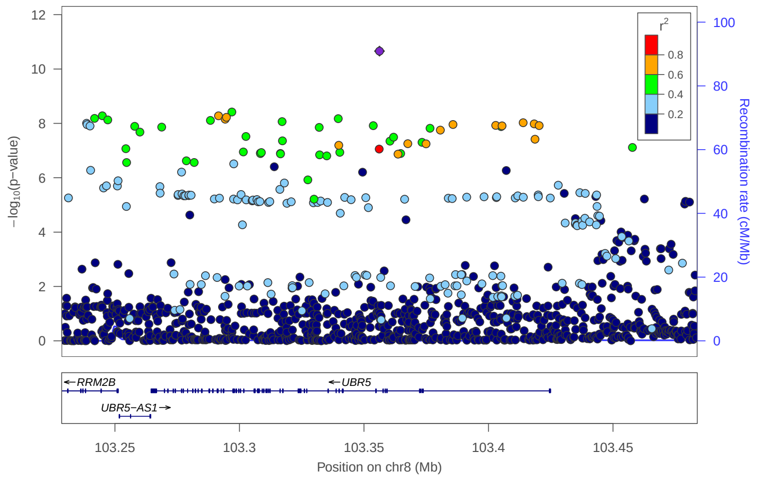


locus 297 8:104648730_CCCTGGGA_C

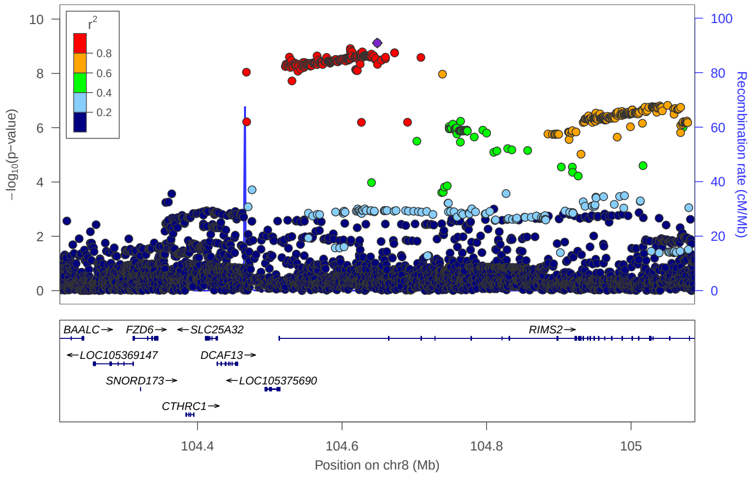


locus 298 rs776935

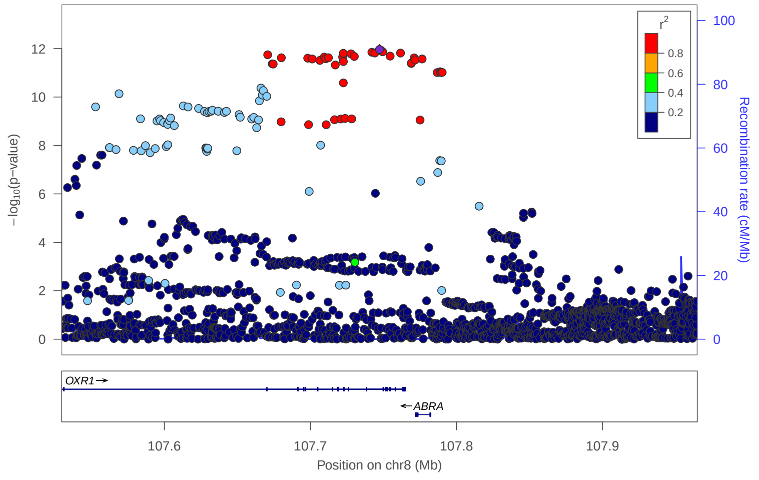

### locus 299 rs1532797

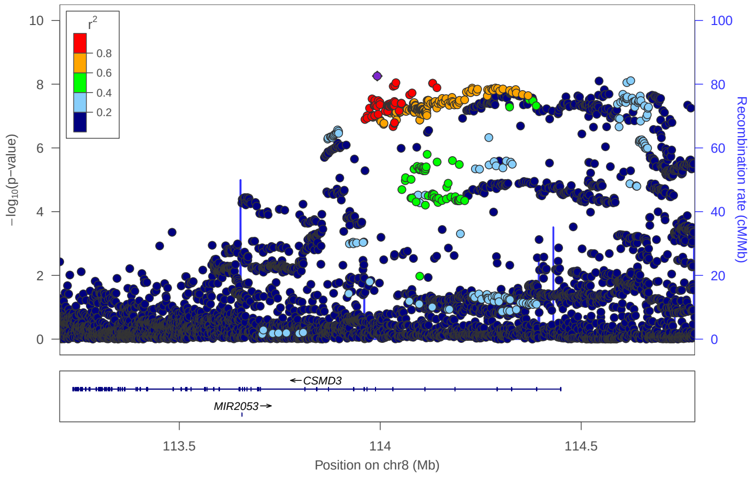


### locus 300 rs62511348

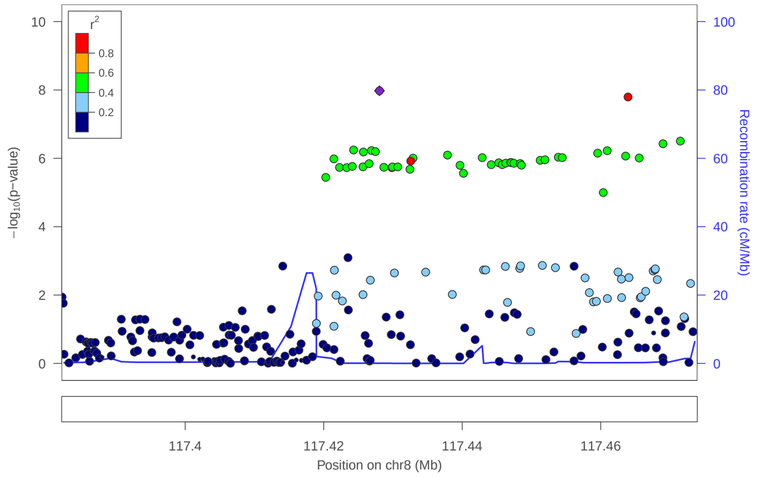


### locus 301 rs6471065

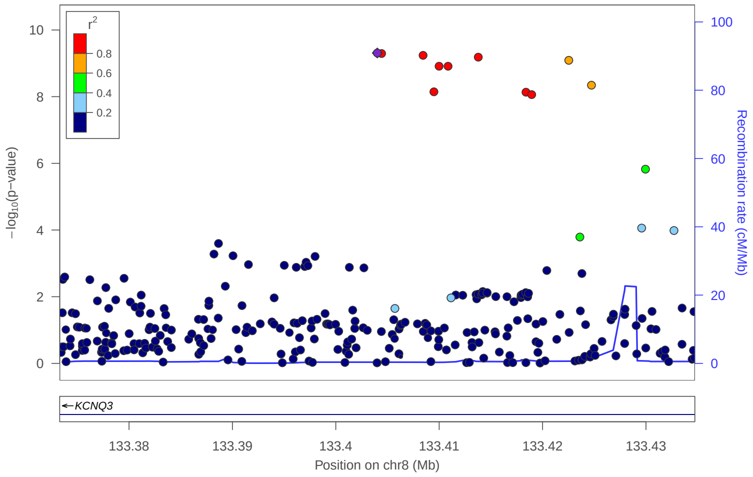


### locus 302 rs2242090

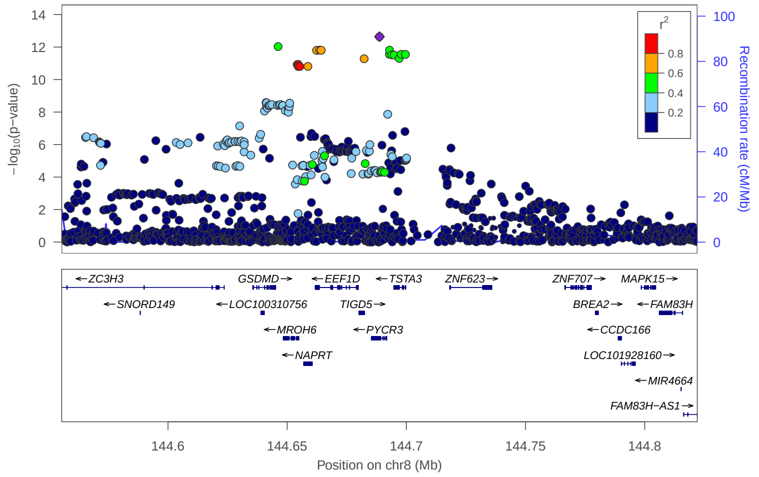


### locus 303 rs17727688

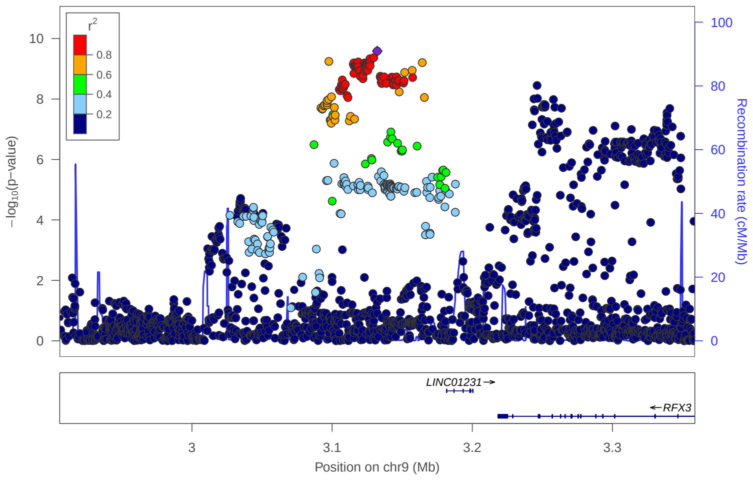


### locus 304 rs10758594

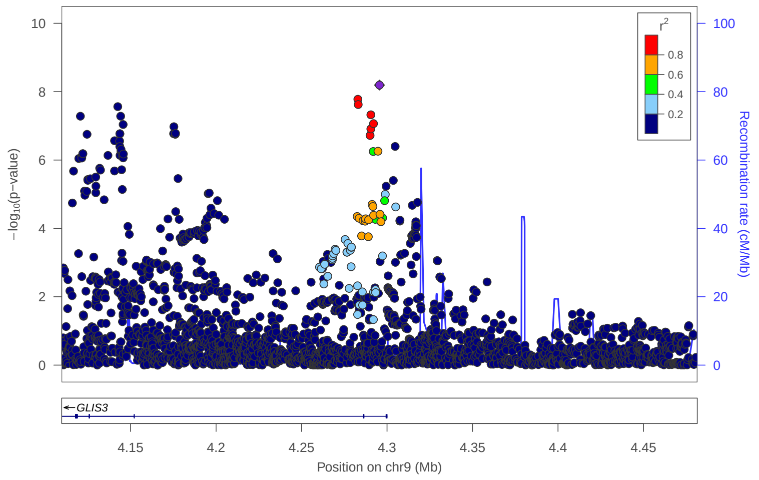


### locus 305 rs7042696

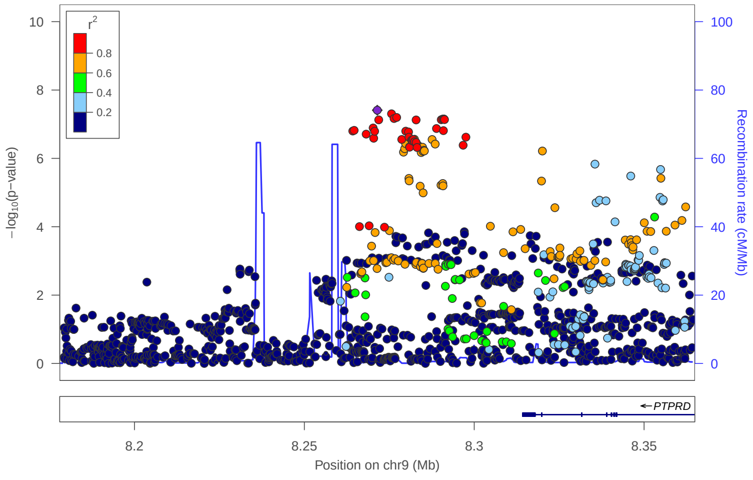


### locus 306 rs118166957

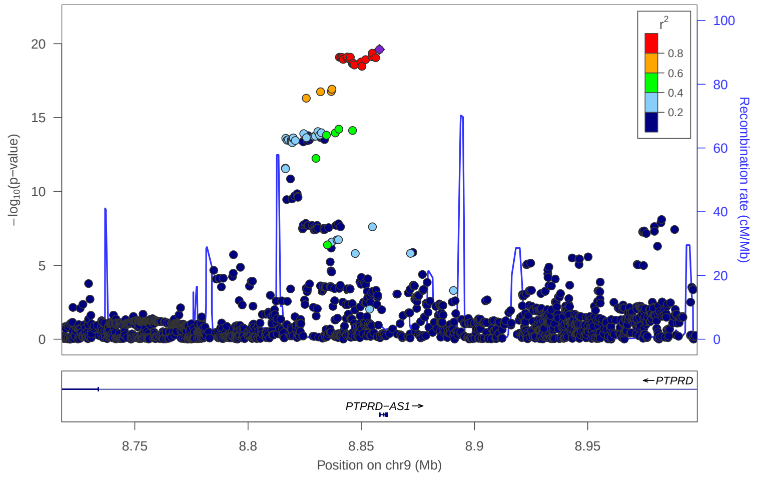

locus 307 rs1322304

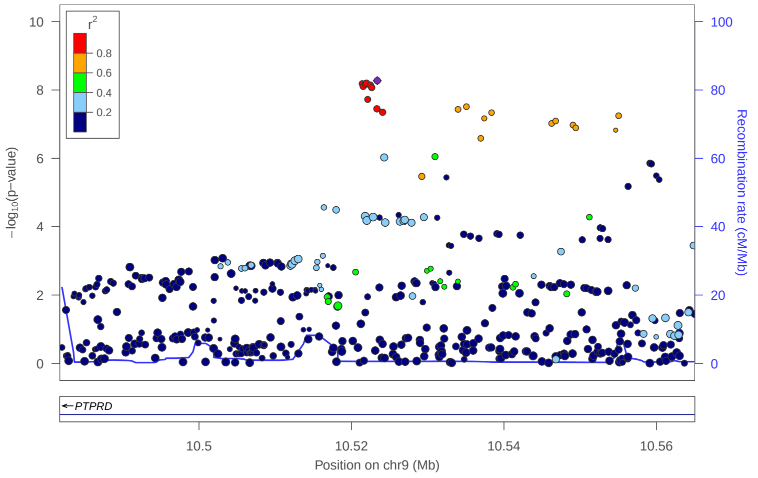


locus 308 rs2130121

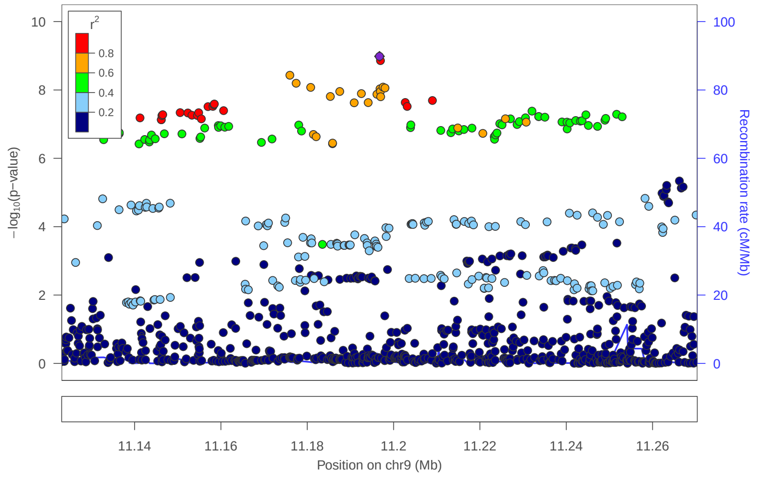


locus 309 rs10756553

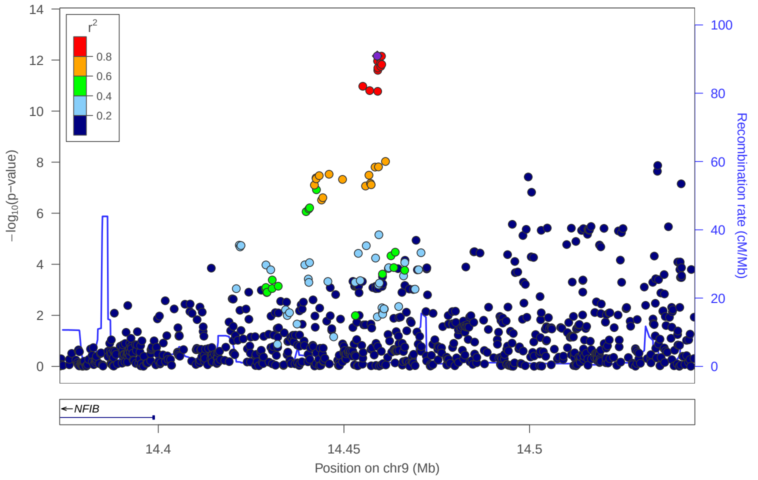


locus 310 rs393488

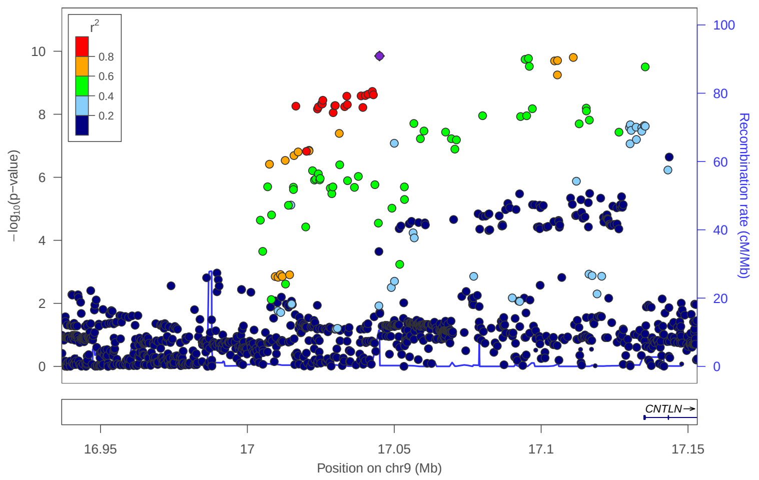


locus 311 rs7032599

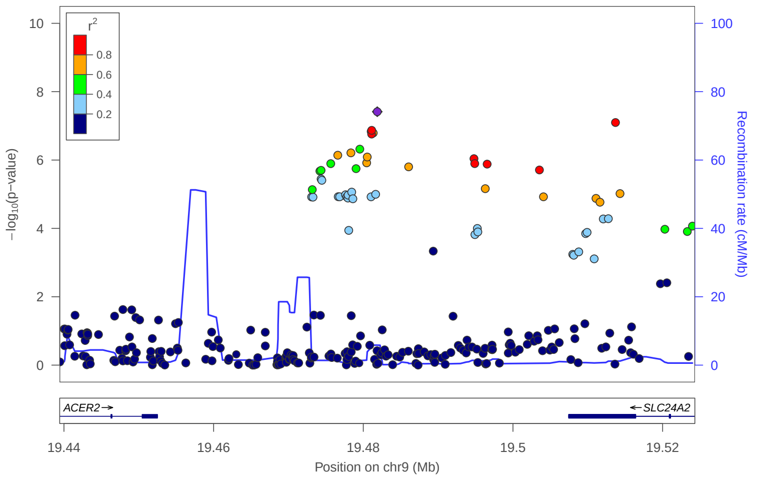


locus 312 rs55863203

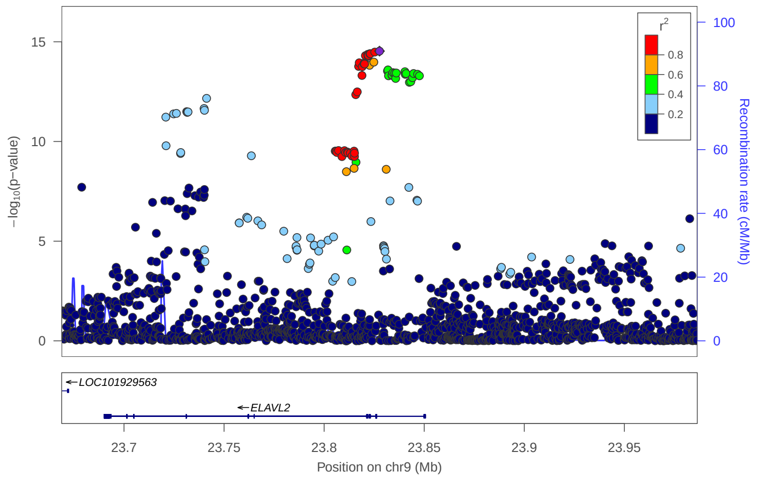


locus 313 rs3824345

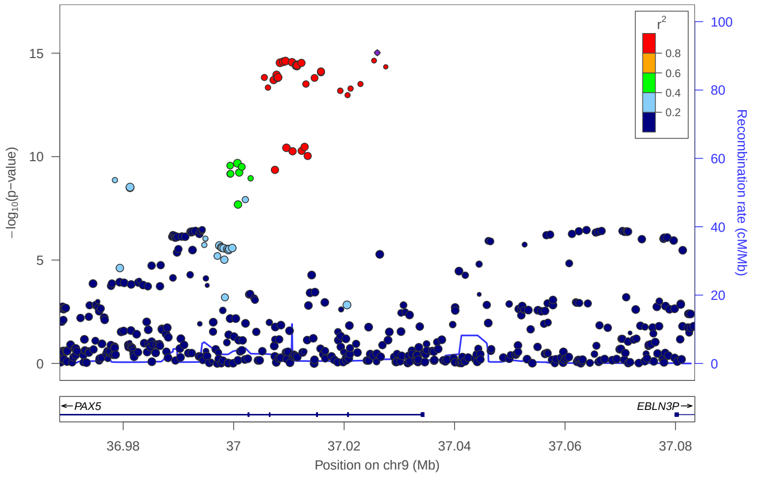


locus 314 rs9777373

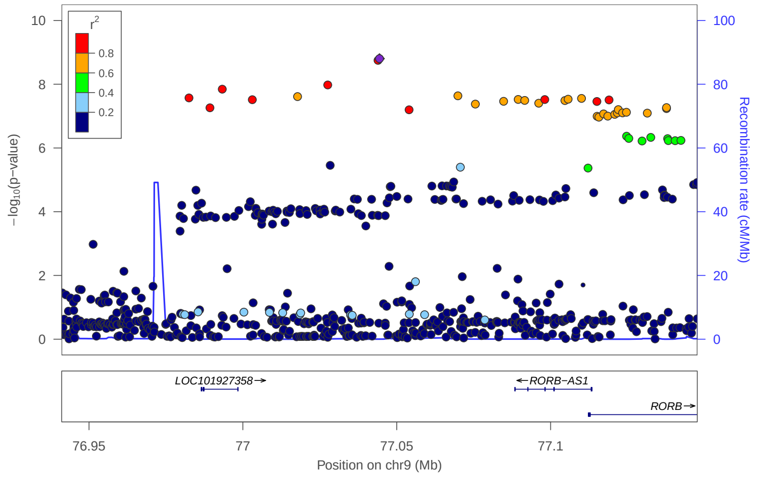

locus 315 rs990559

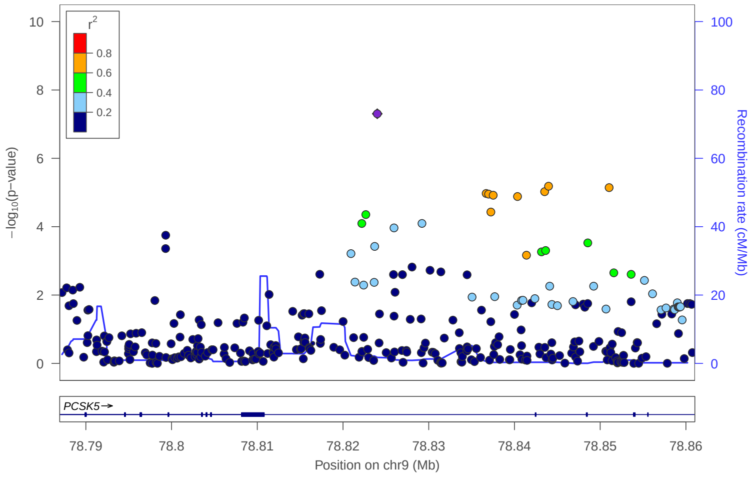


locus 316 rs35820496

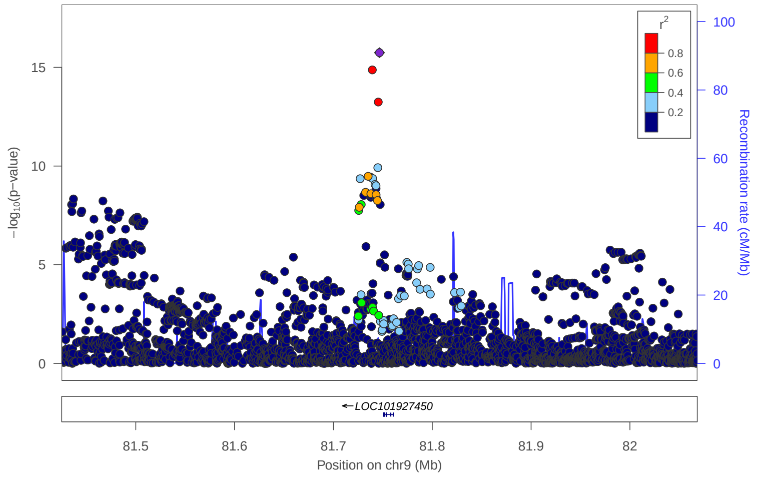


locus 317 rs1328435

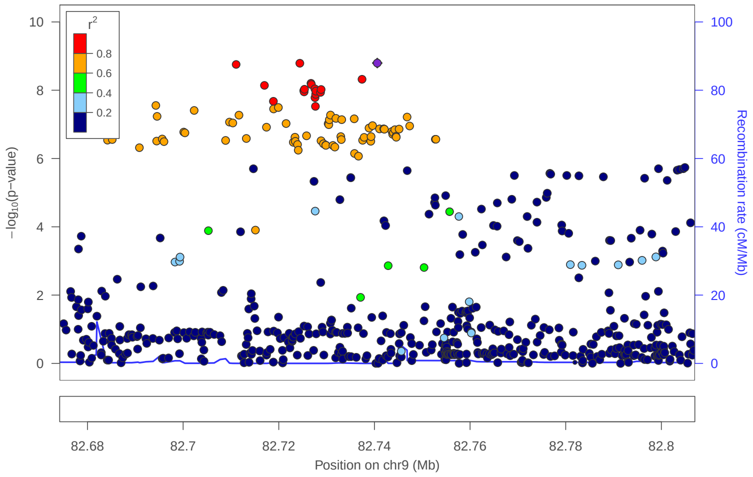


locus 318 rs10992756

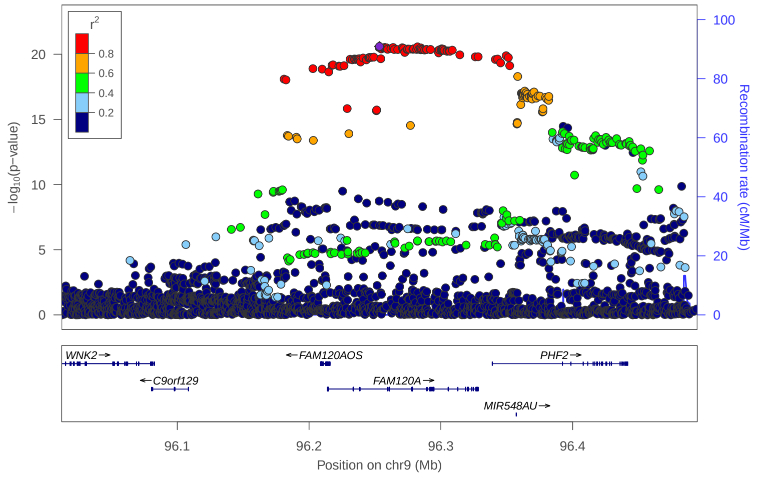


locus 319 rs7853585

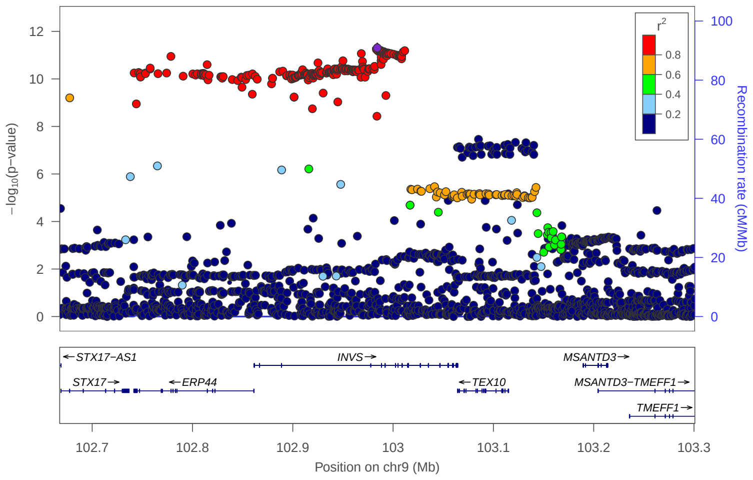


locus 320 rs10817718

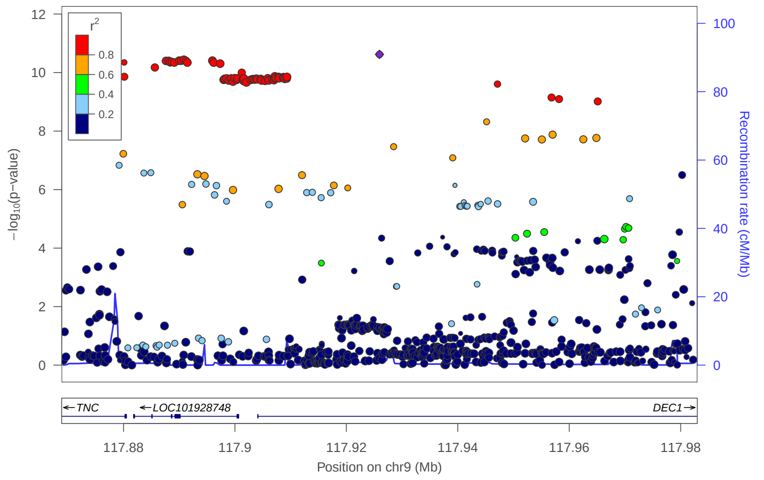


locus 321 rs4595203

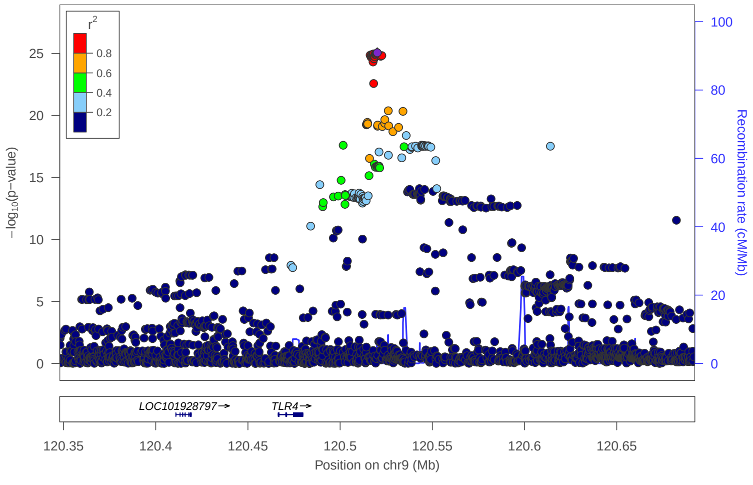


locus 322 rs10818399

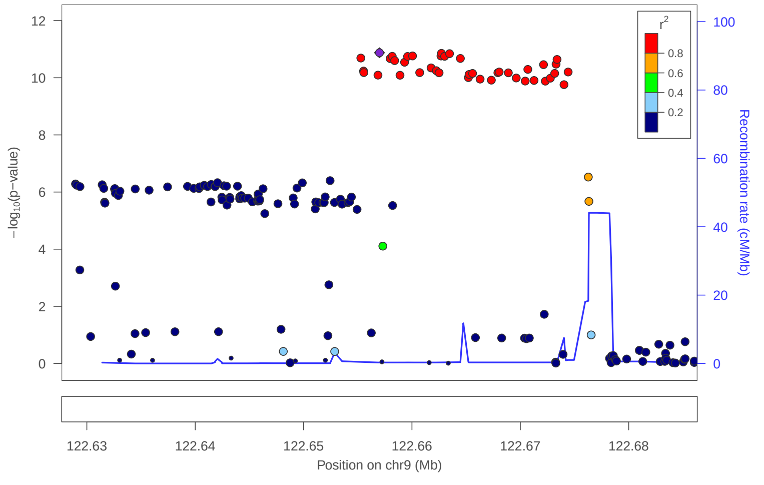

### locus 323 rs2792990

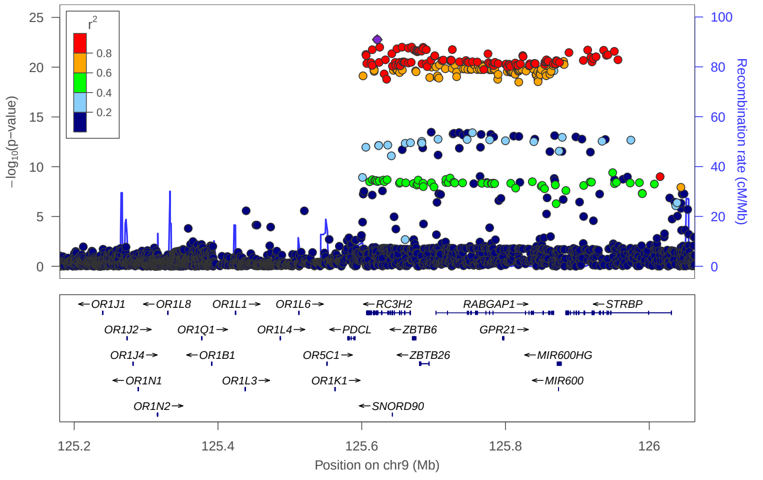


### locus 324 rs2502817

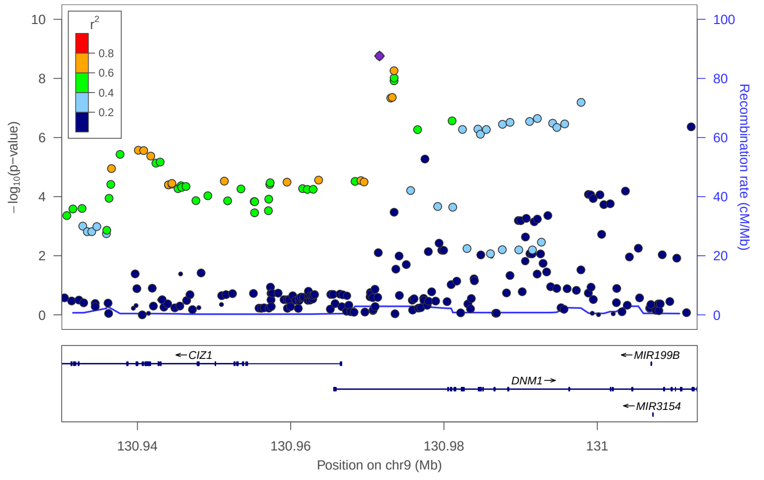


### locus 325 rs10819613

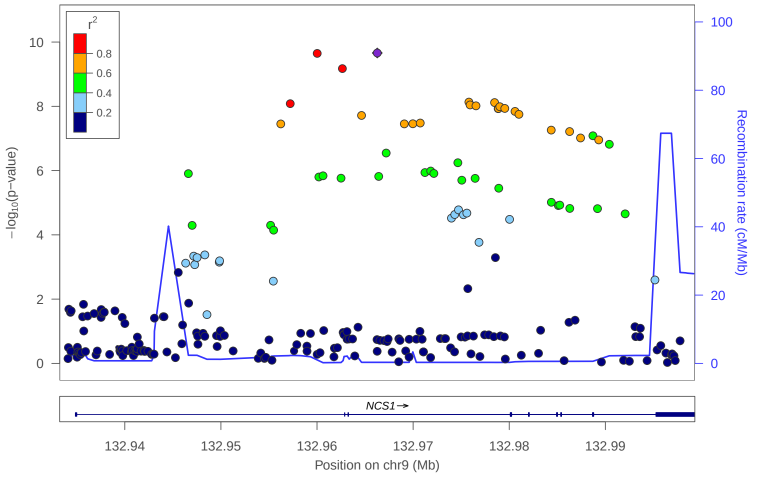


### locus 326 rs10125388

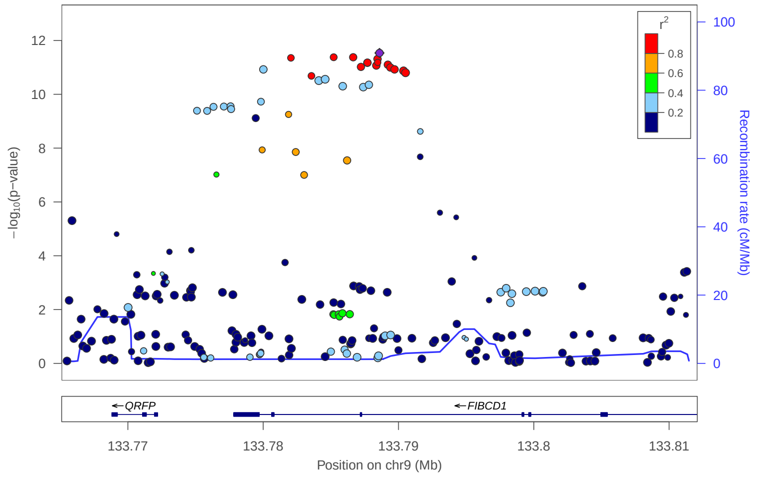


### locus 327 rs9411339

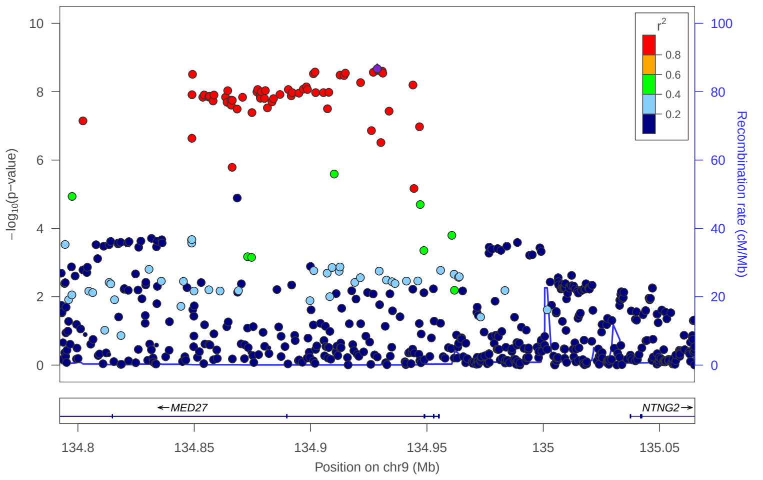


### locus 328 rs72766657

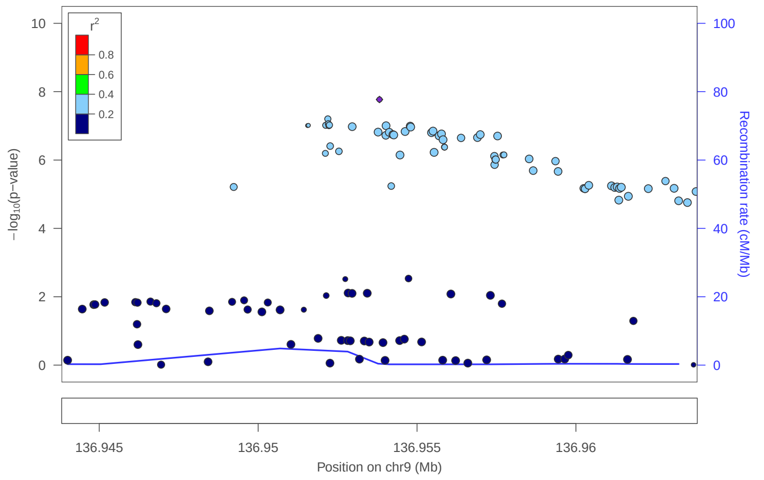


### locus 329 rs1105175

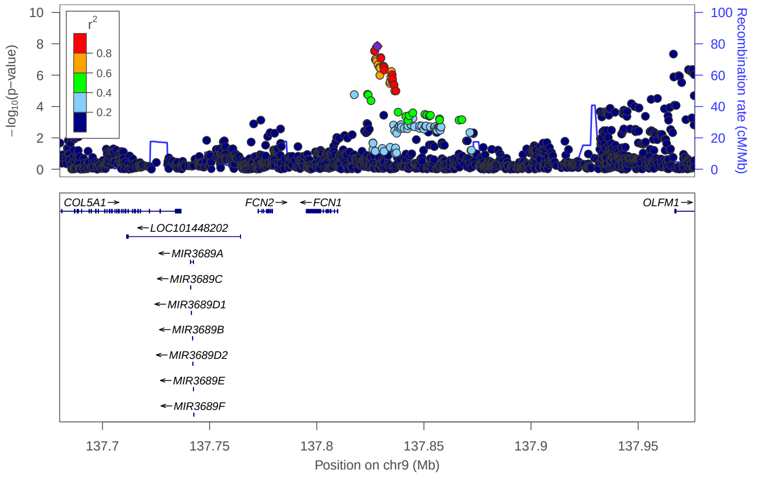


### locus 330 rs12684650

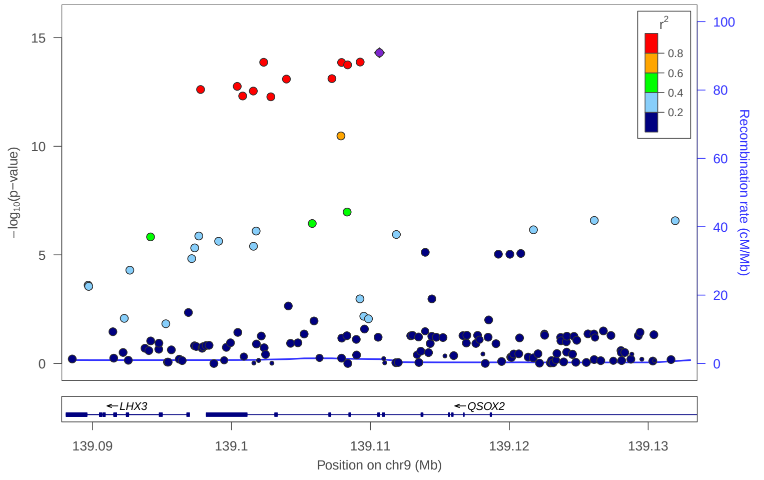

locus 331 9:140265782_C_T

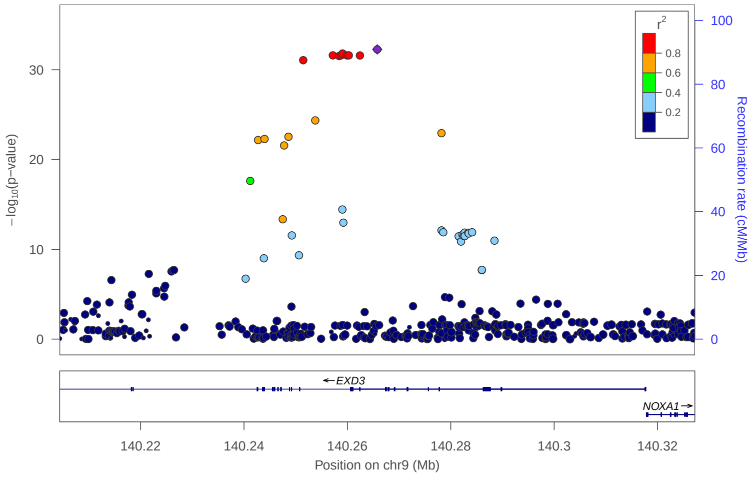


locus 332 rs12775090

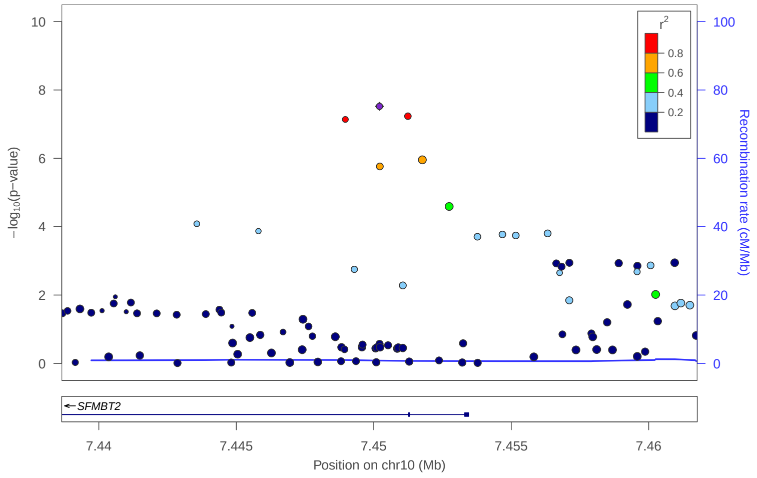


locus 333 rs10906391

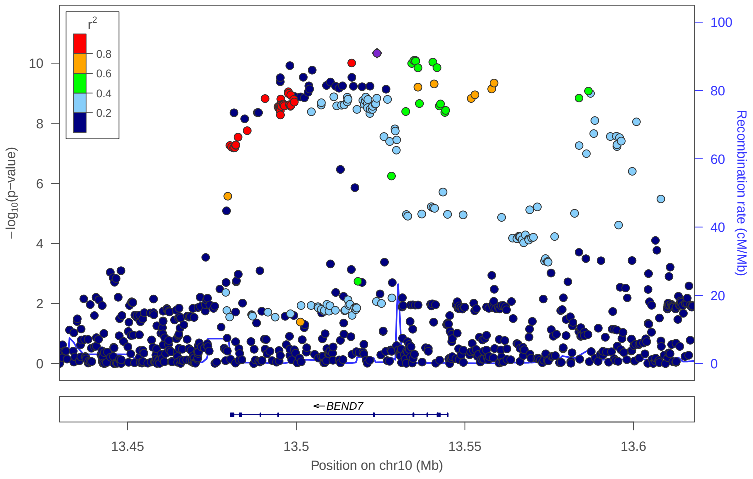


locus 334 rs1416901

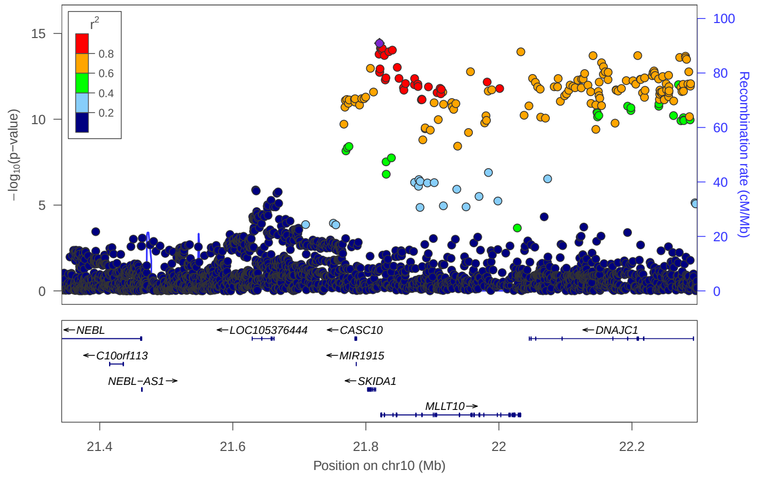


locus 335 rs9943377

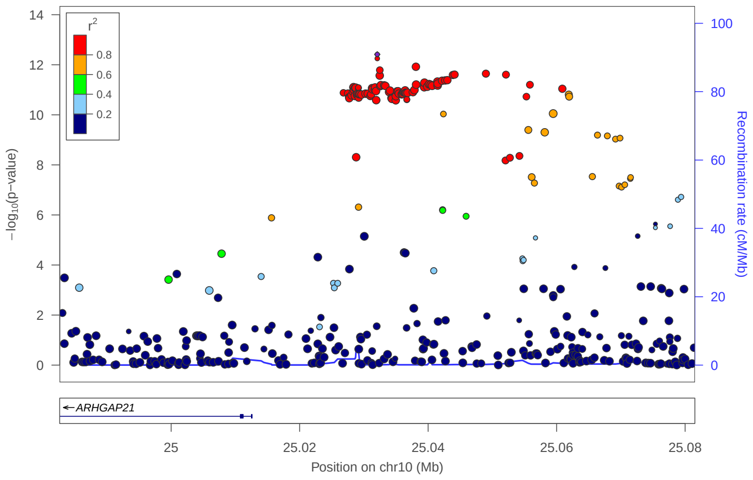


locus 336 rs117404983

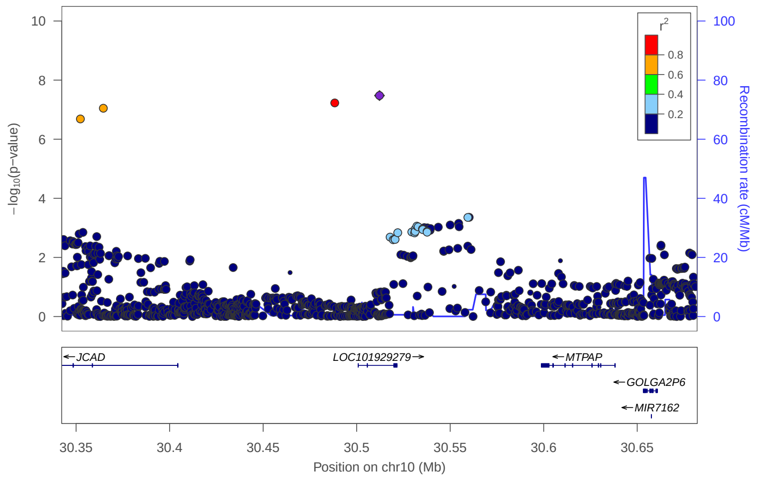


locus 337 rs12356643

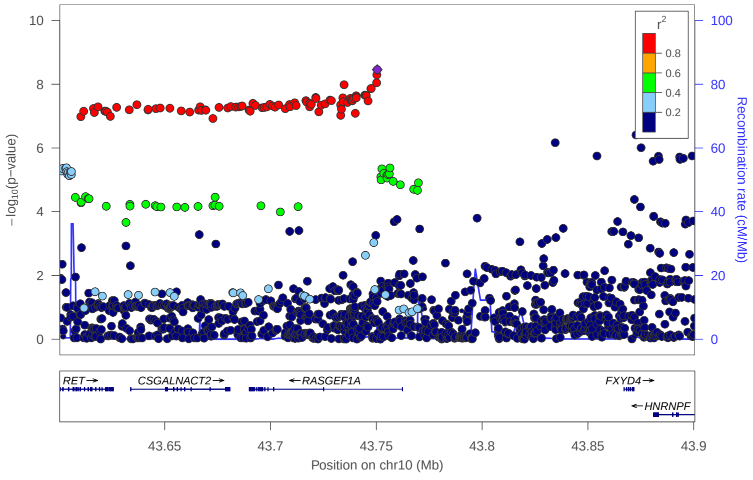


locus 338 rs11004838

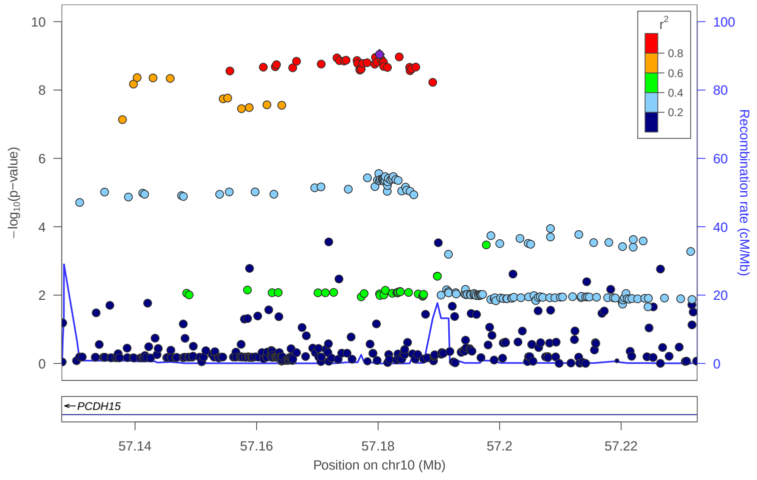

locus 339 rs224139

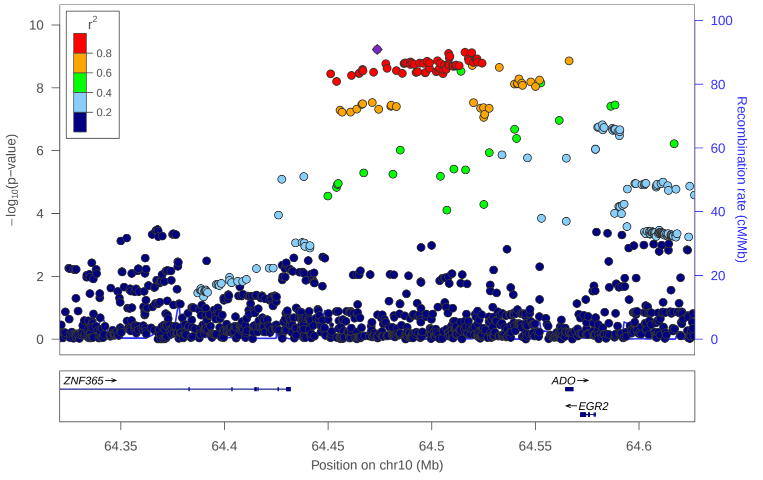


locus 340 rs7924036

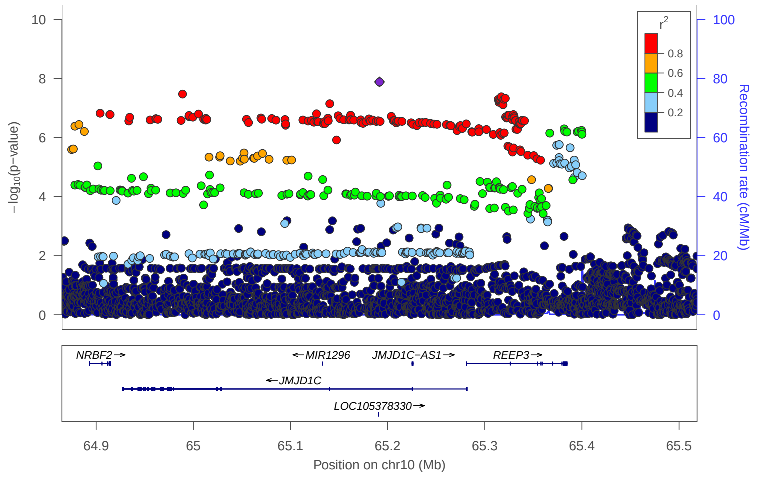


locus 341 rs7072222

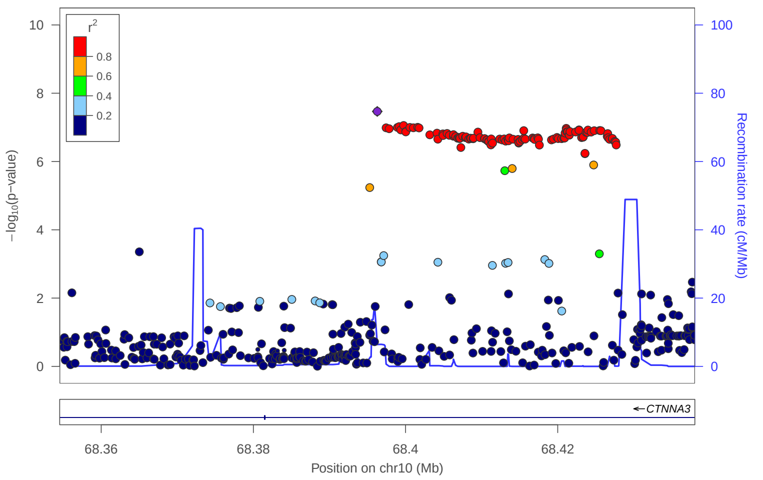


locus 342 rs111296206

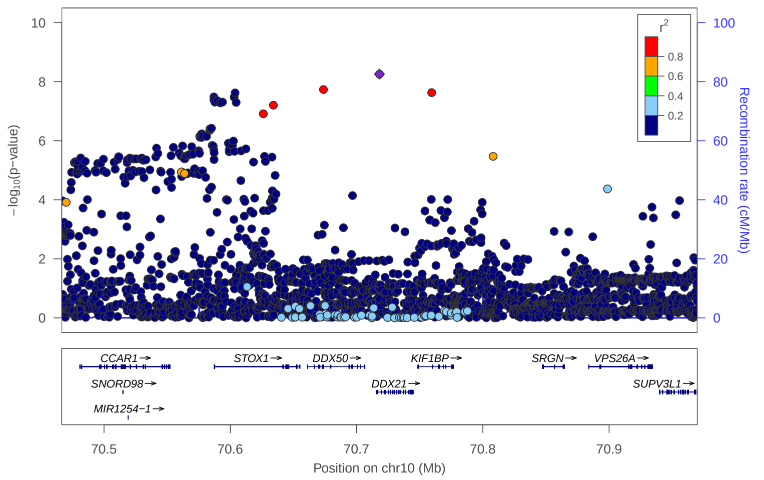


locus 343 rs2395137

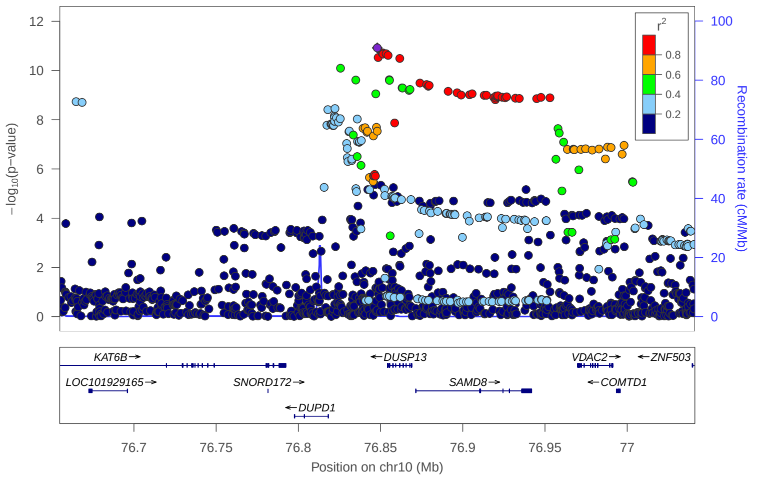


locus 344 rs1407736

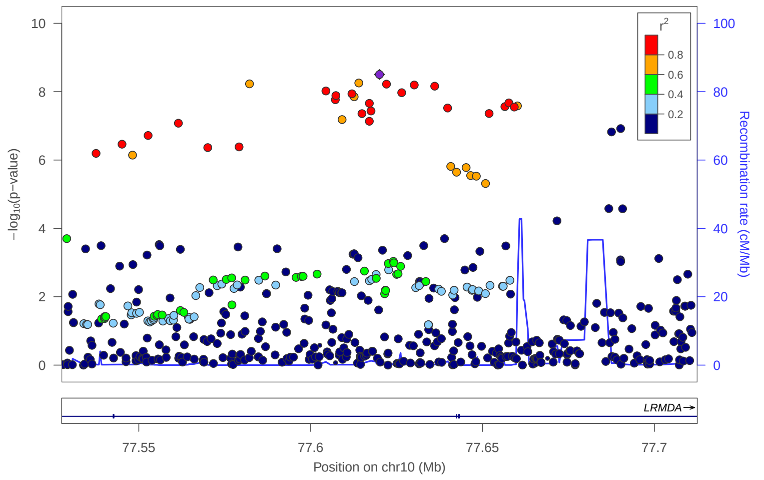


locus 345 rs566903

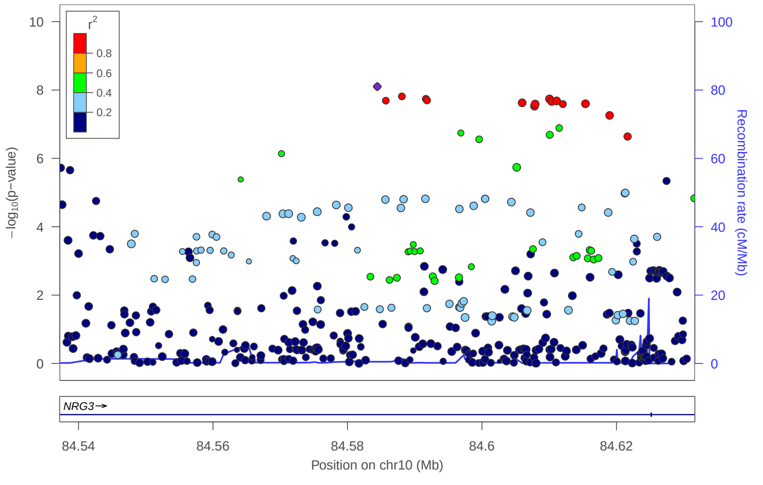


locus 346 rs188189626

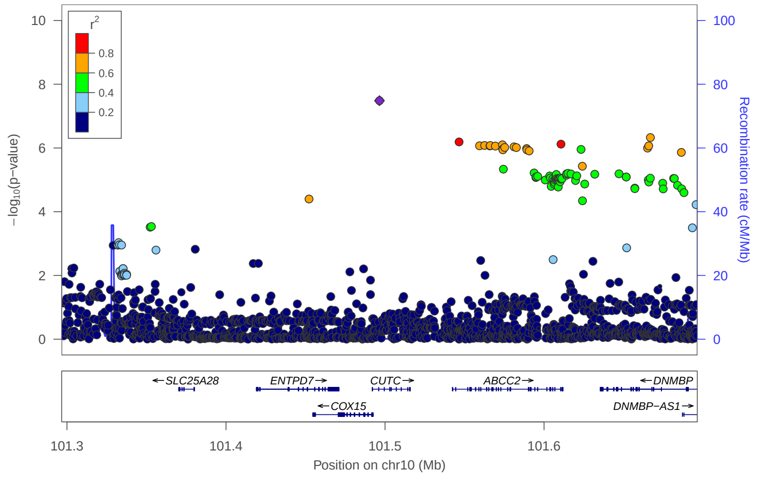

locus 347 rs111374164

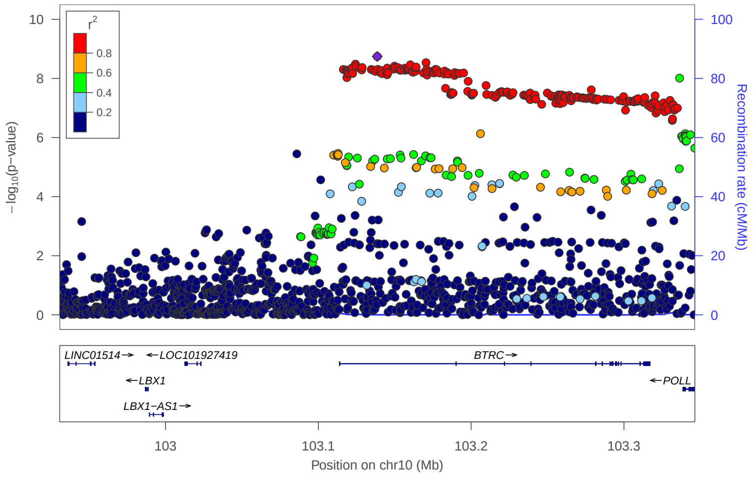


locus 348 rs11191434

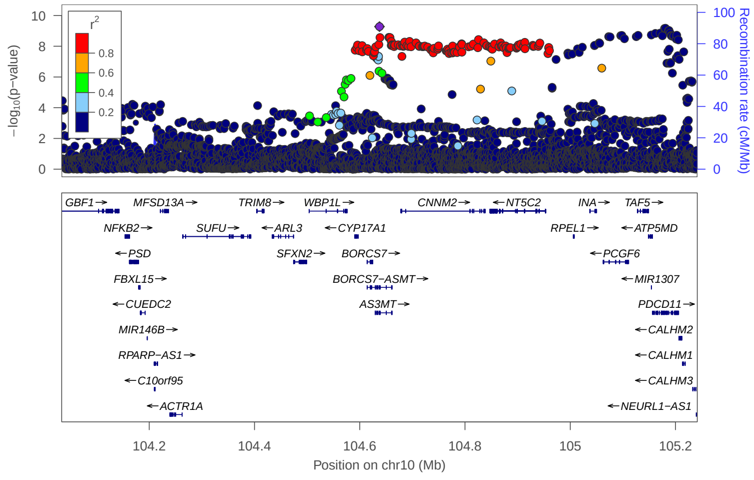


locus 349 rs56093138

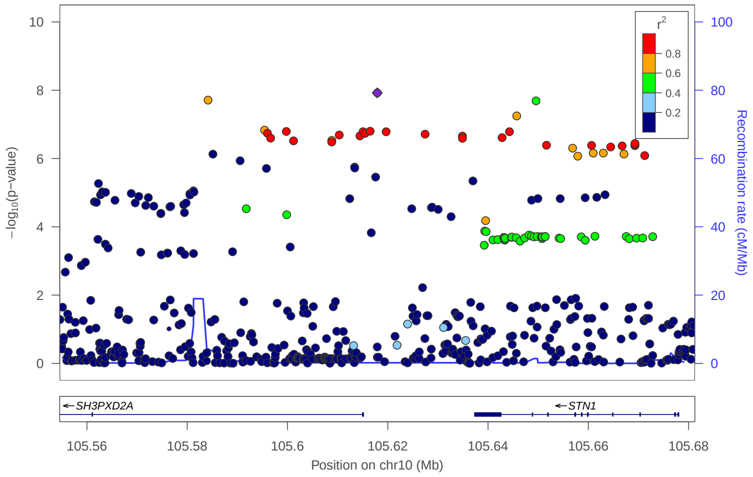


locus 350 rs35723963

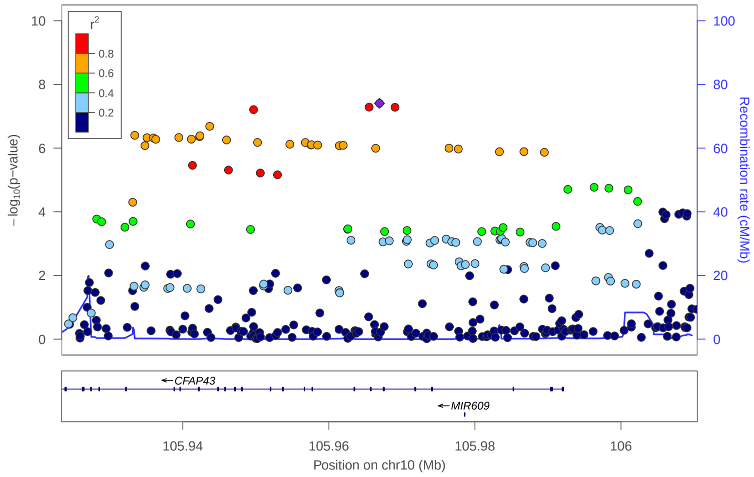


locus 351 rs9787523

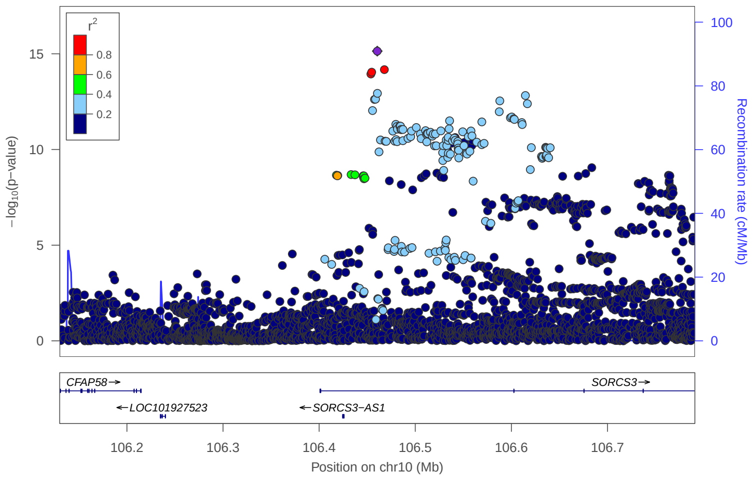


locus 352 rs1331156

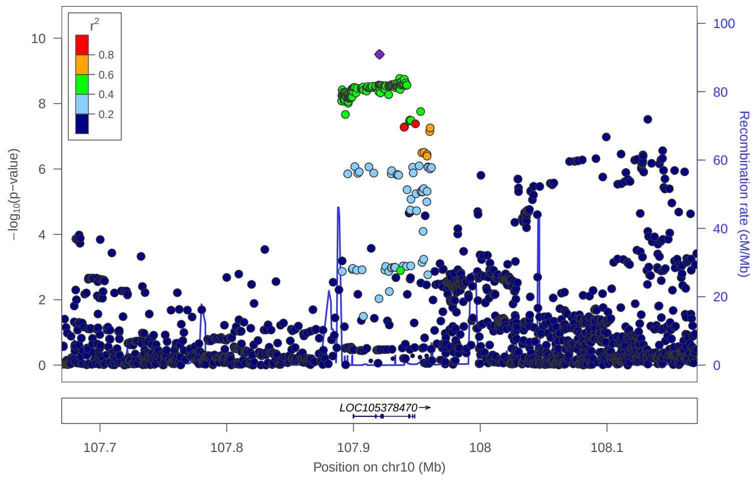


locus 353 rs2184796

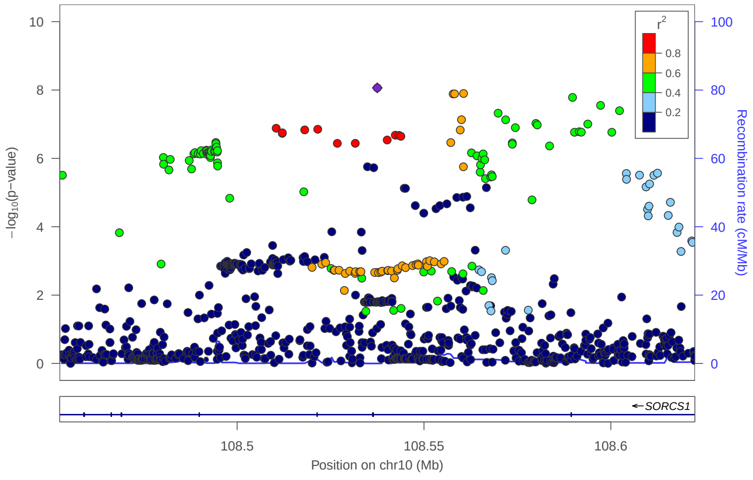


locus 354 rs72814942

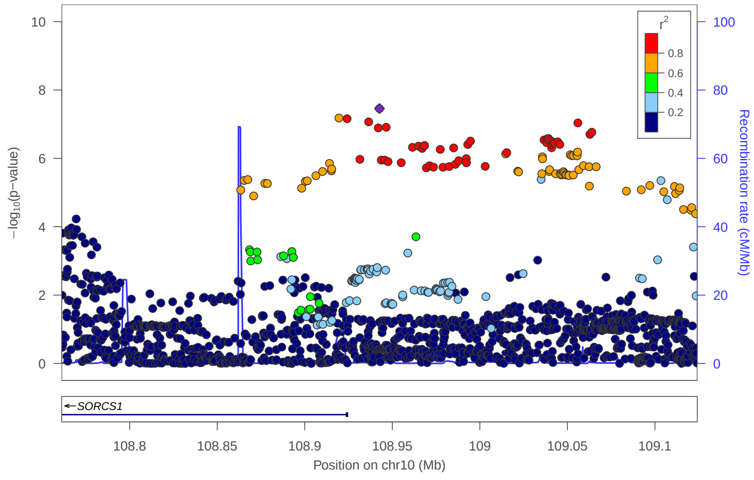

### locus 355 rs17130183

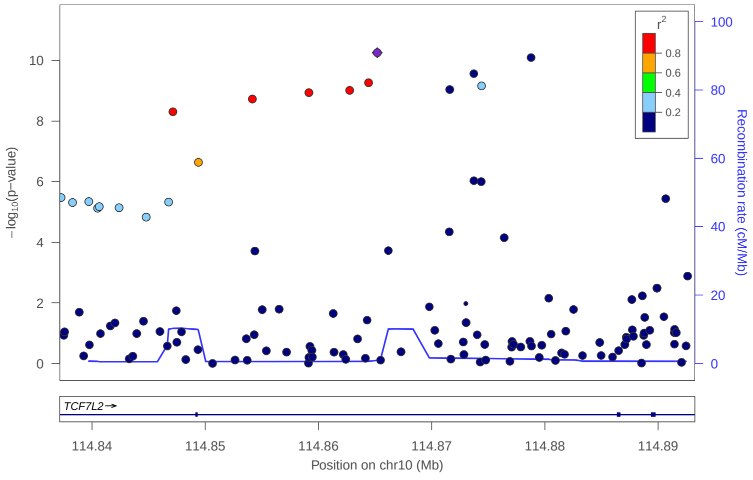


### locus 356 rs7915425

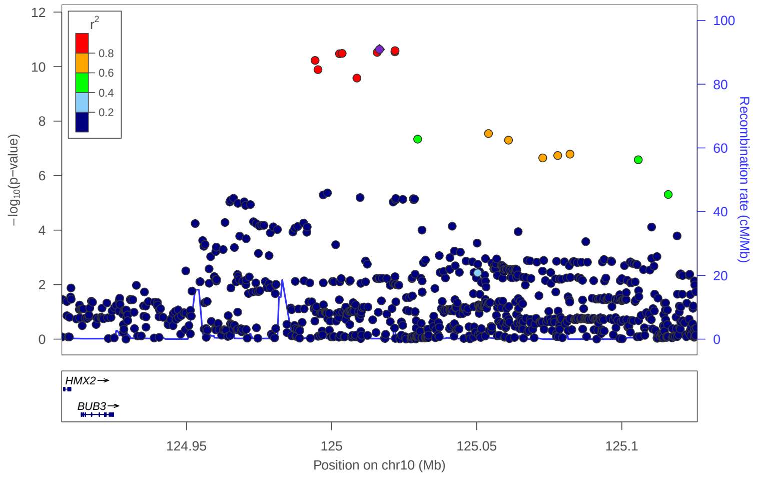


### locus 357 rs2065677

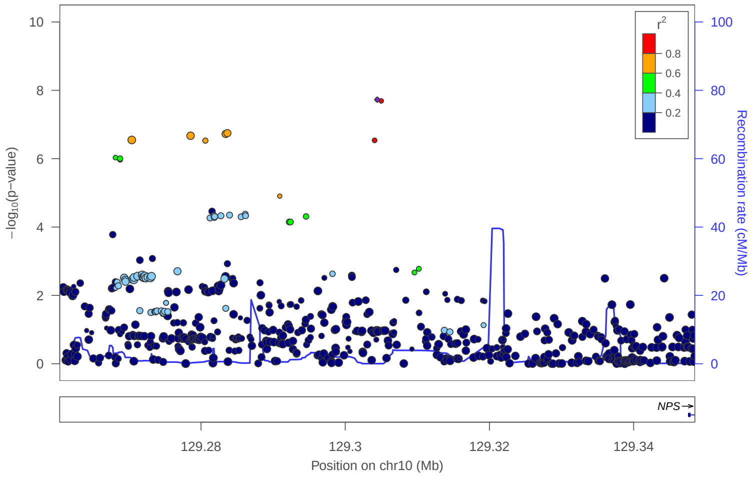


### locus 358 rs9971199

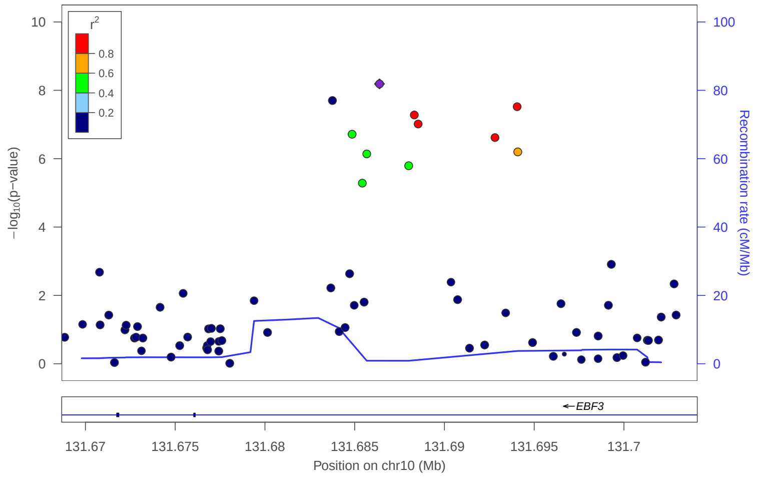


### locus 359 rs2748432

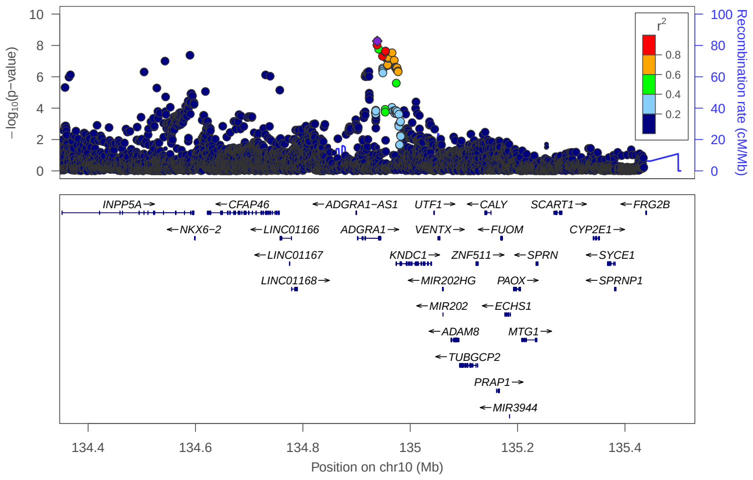


### locus 360 rs1552514

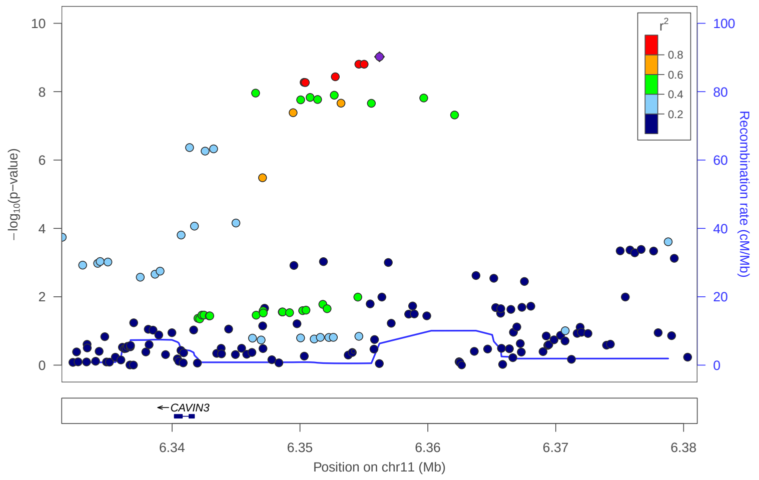


### locus 361 rs7928823

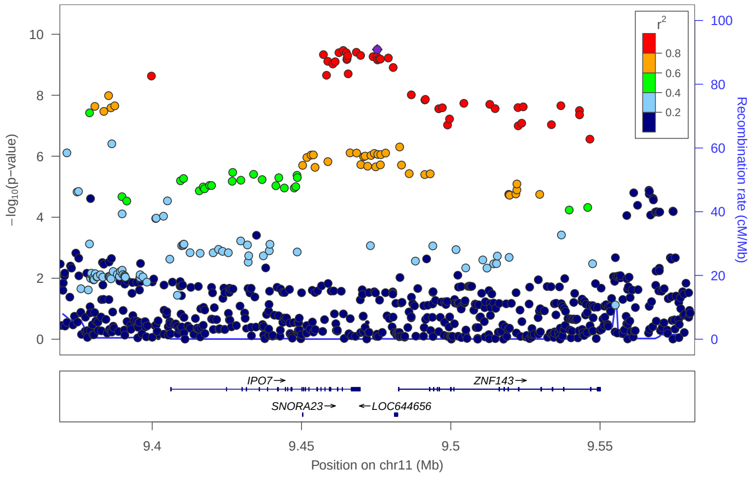


### locus 362 rs75742406

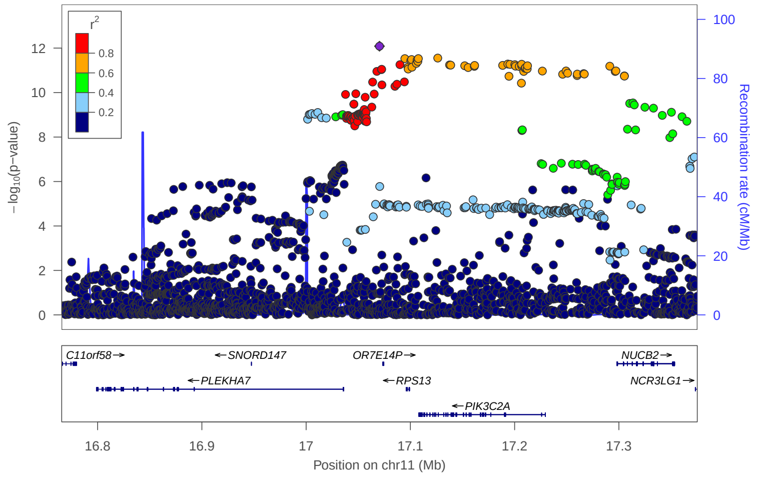

locus 363 rs593839

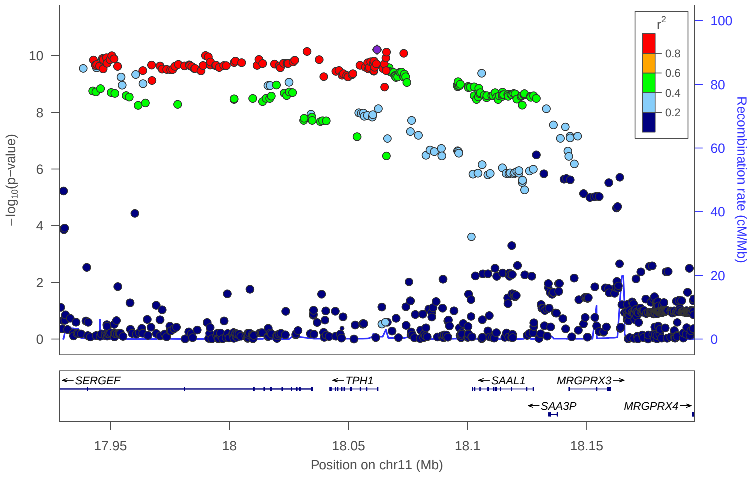


locus 364 rs12284708

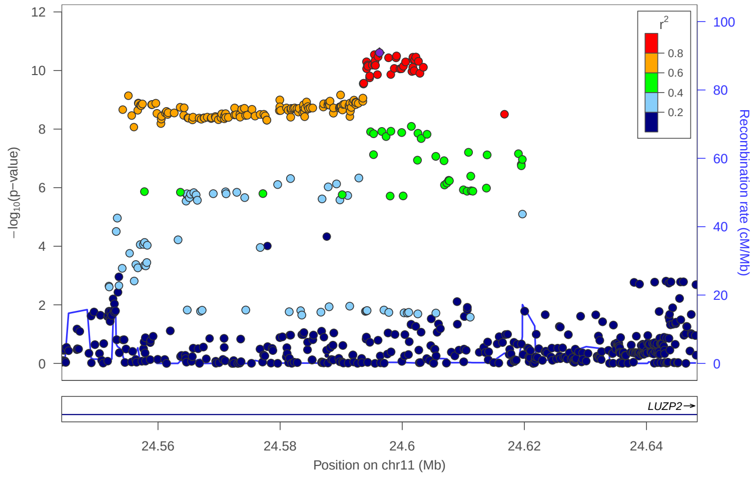


locus 365 rs1806152

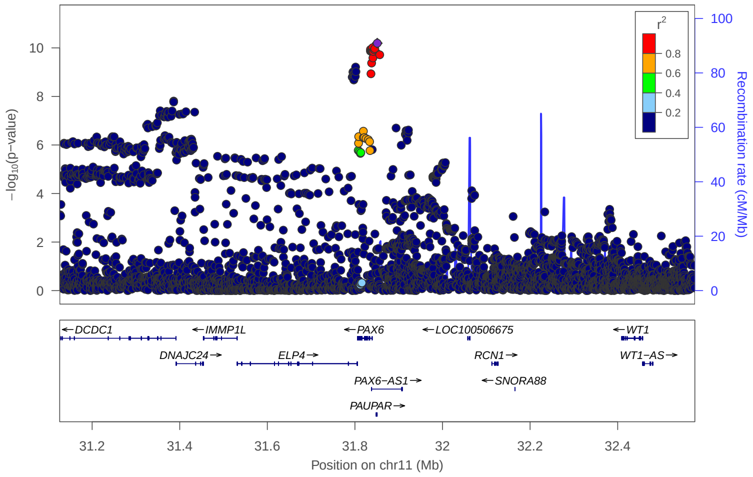


locus 366 rs10837042

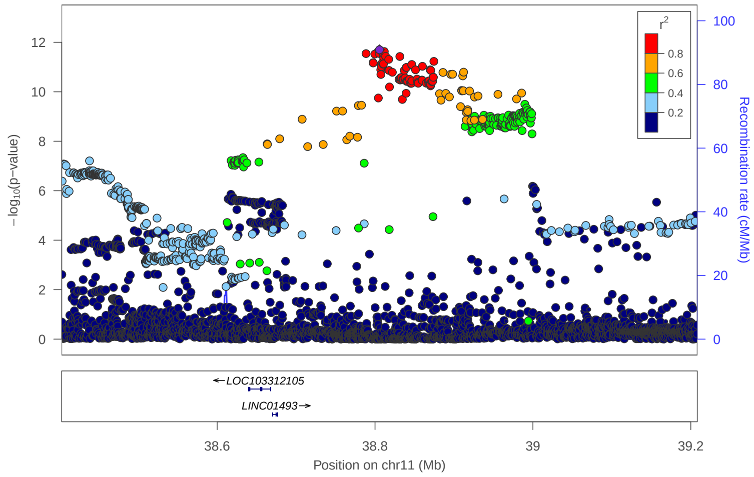


locus 367 rs72899452

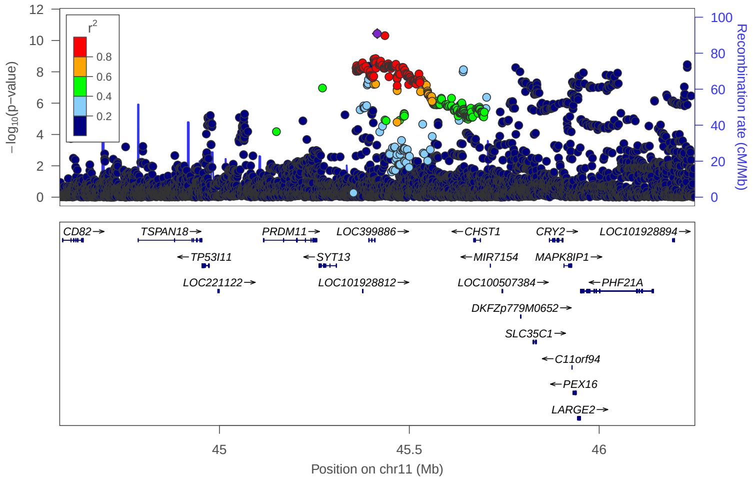


locus 368 rs150870784

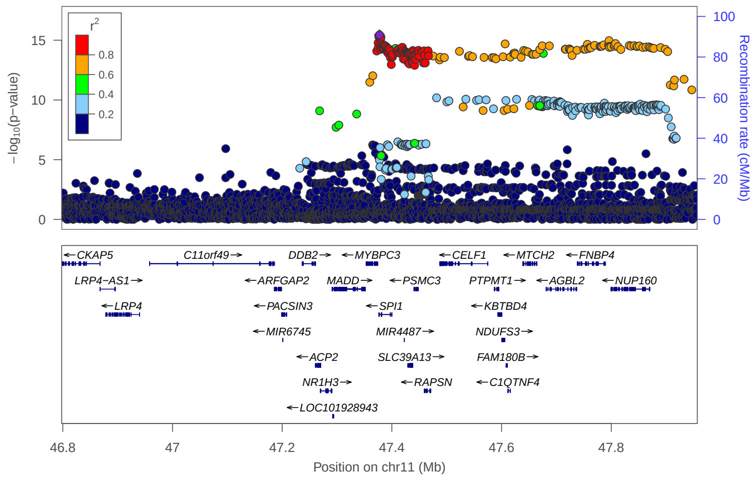


locus 369 rs7934304

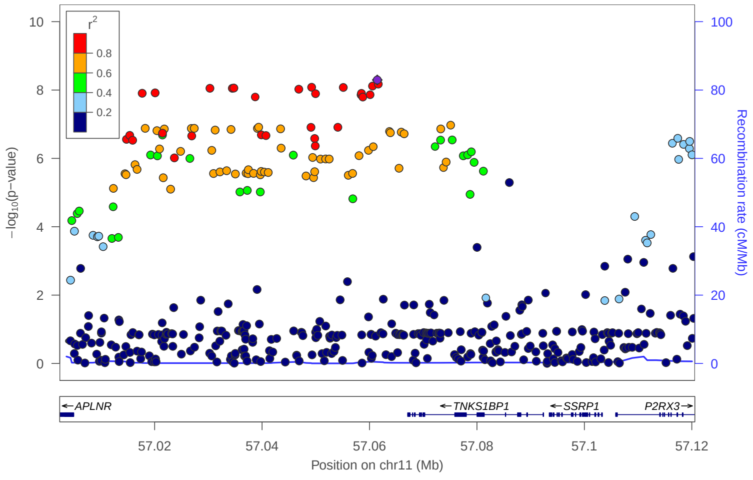


locus 370 rs12790660

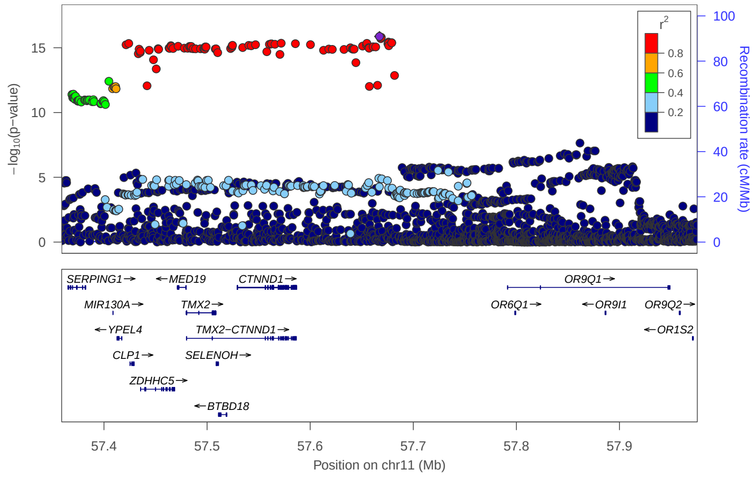

### locus 371 rs7940070

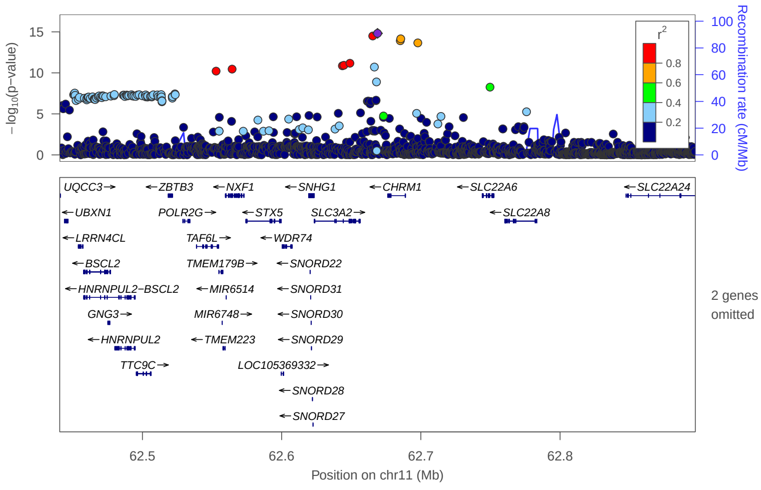


### locus 372 rs12272545

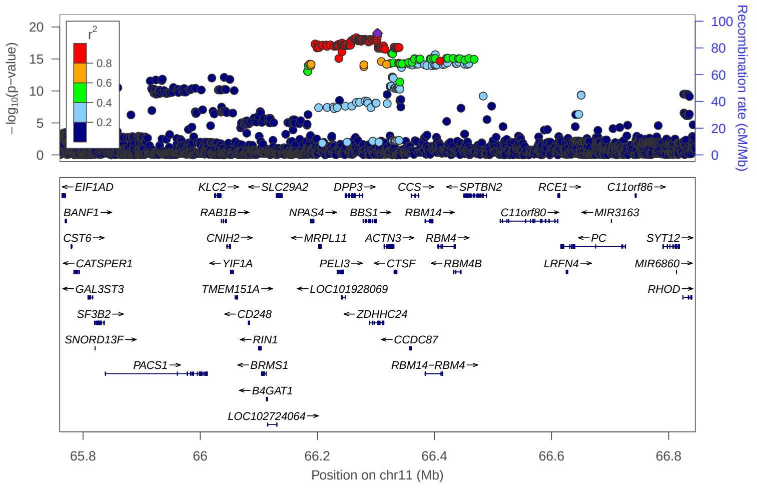


### locus 373 rs79693059

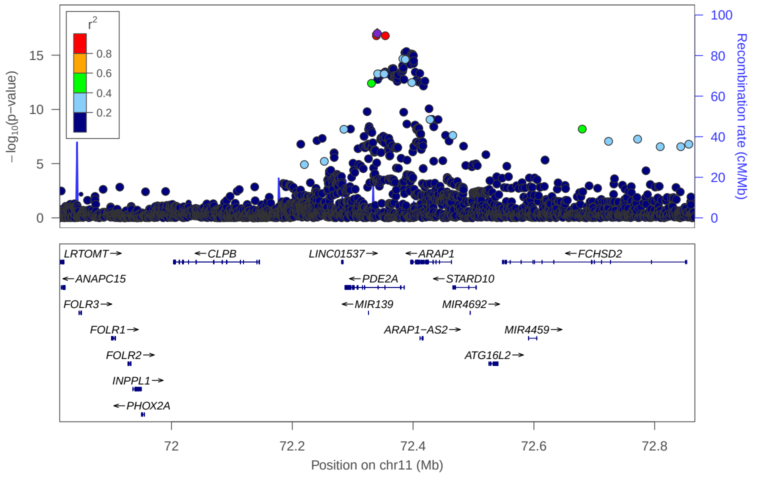


### locus 374 rs505621

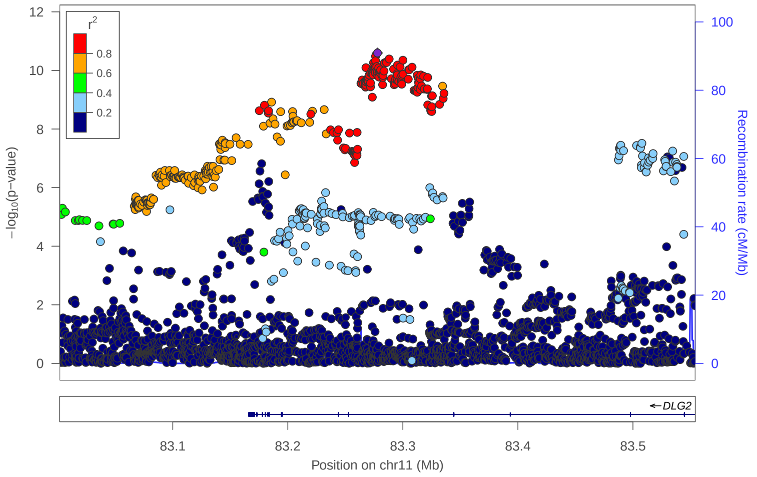


### locus 375 rs519778

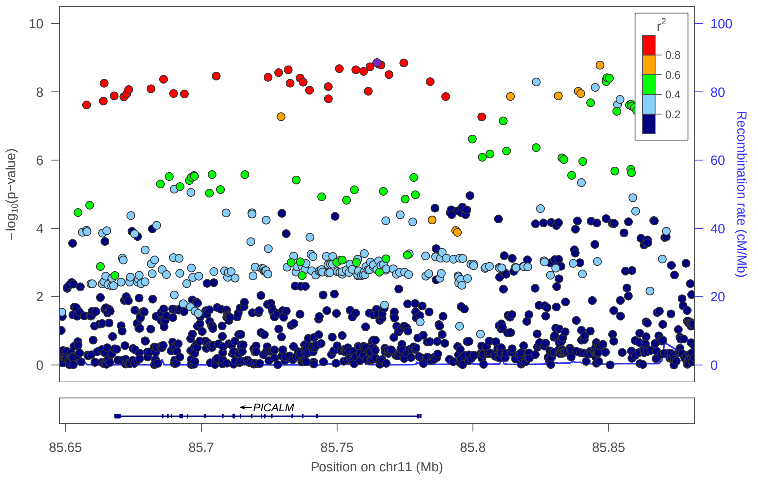


### locus 376 rs1939252

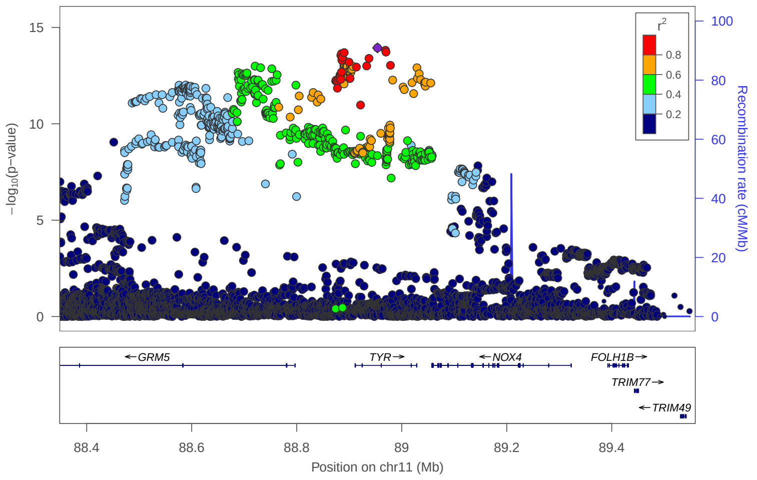


### locus 377 rs10893052

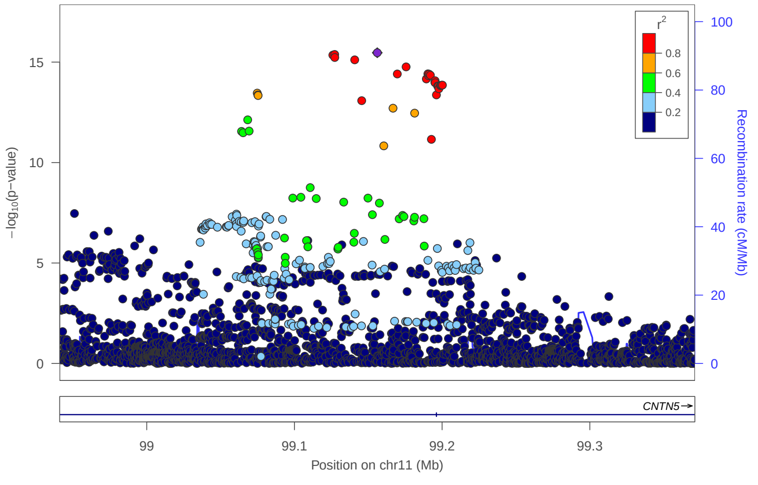


### locus 378 rs7110574

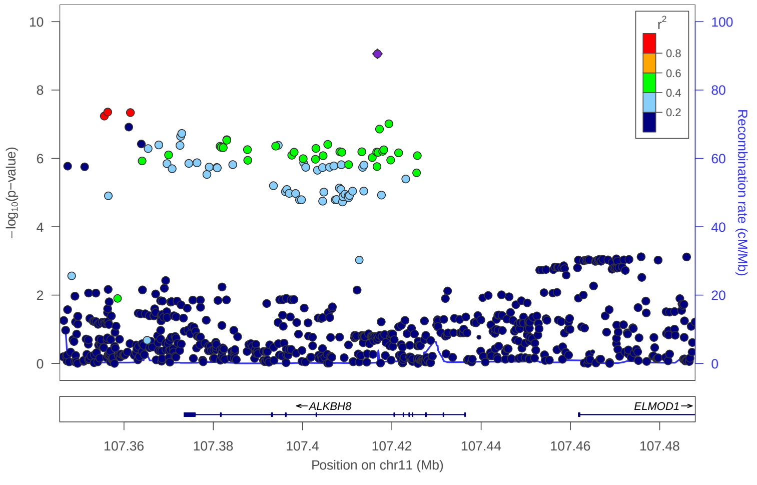

locus 379 rs3802847

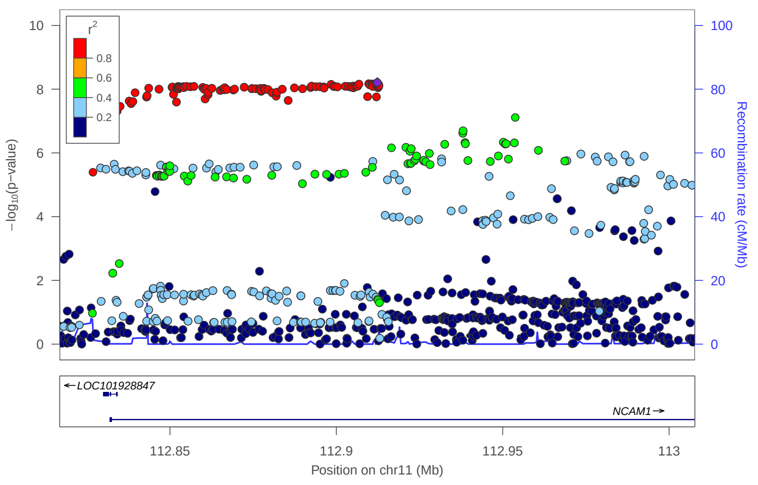


locus 380 rs1064939

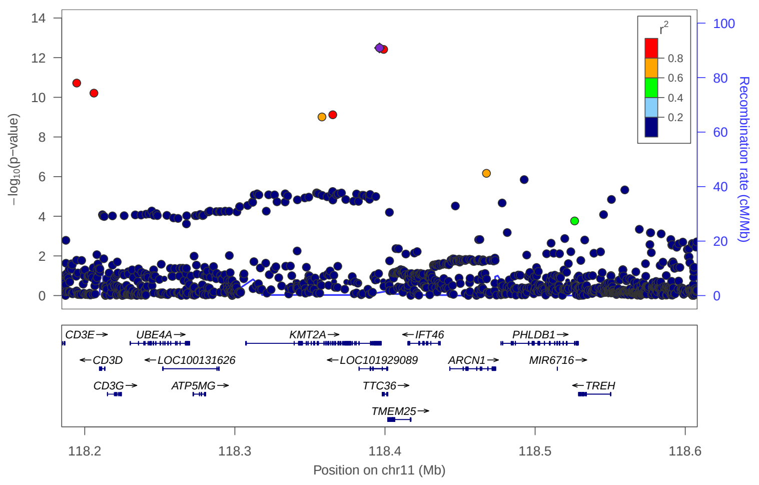


locus 381 rs142192979

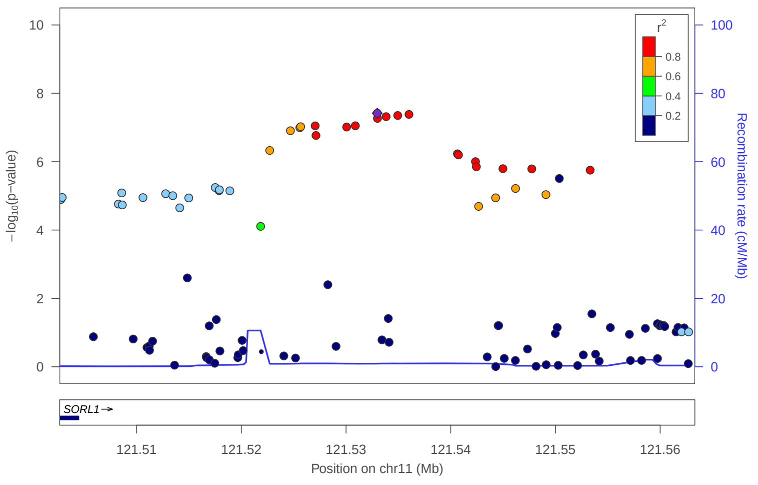


locus 382 rs10894504

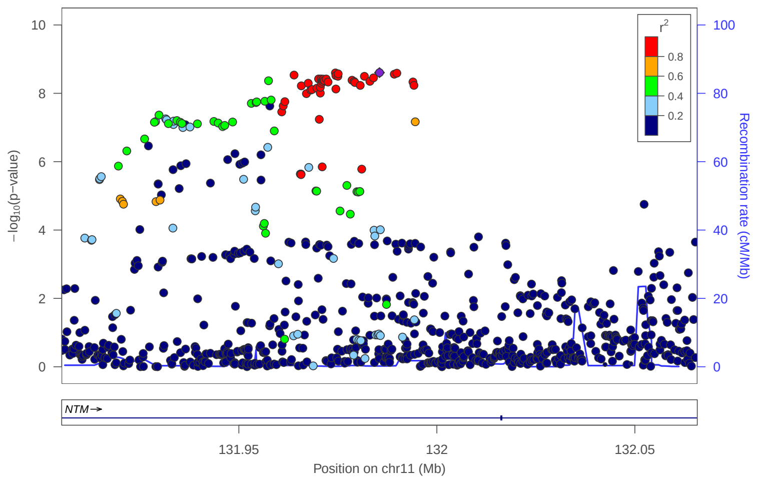


locus 383 rs4937860

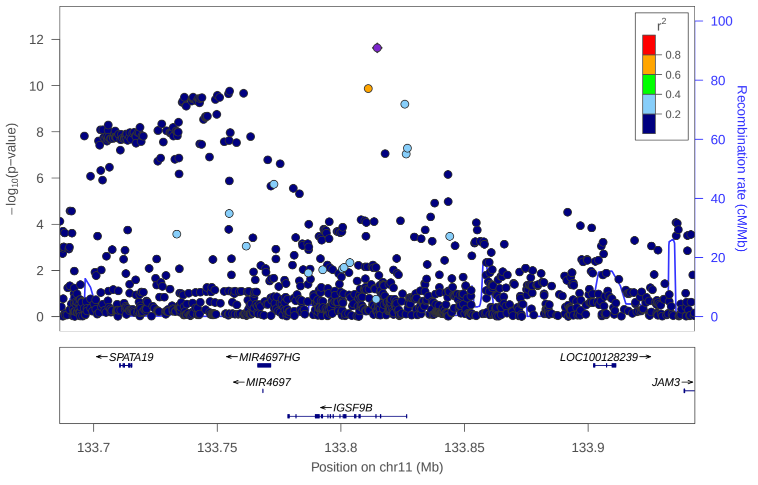


locus 384 rs58475265

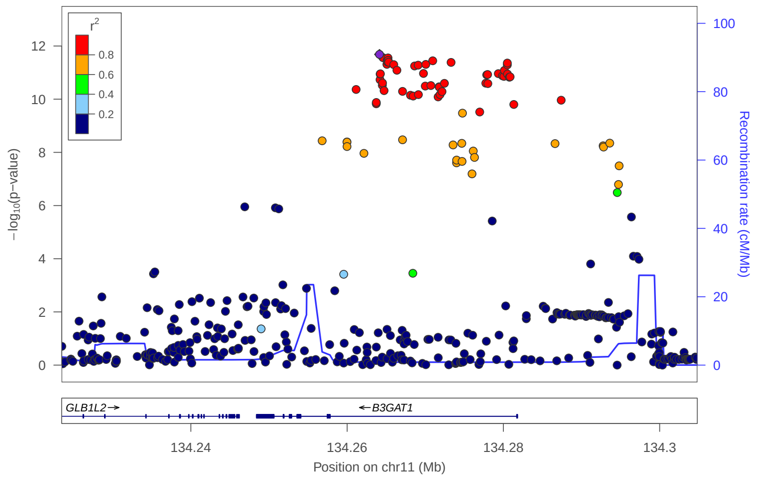


locus 385 rs35745815

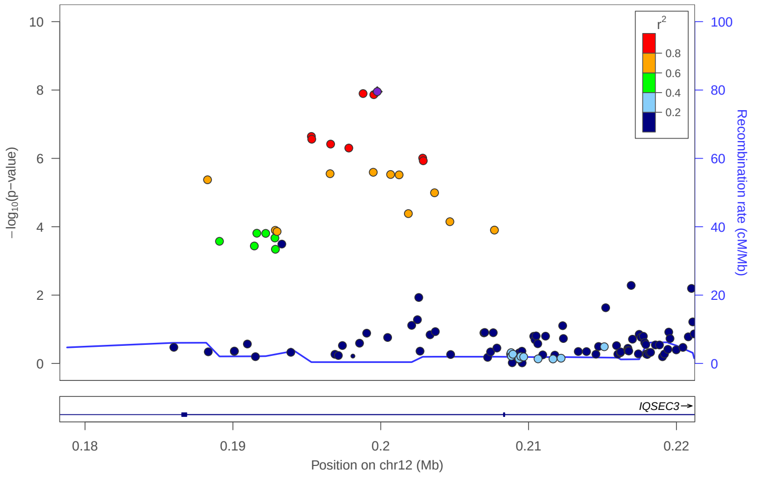


locus 386 rs2286729

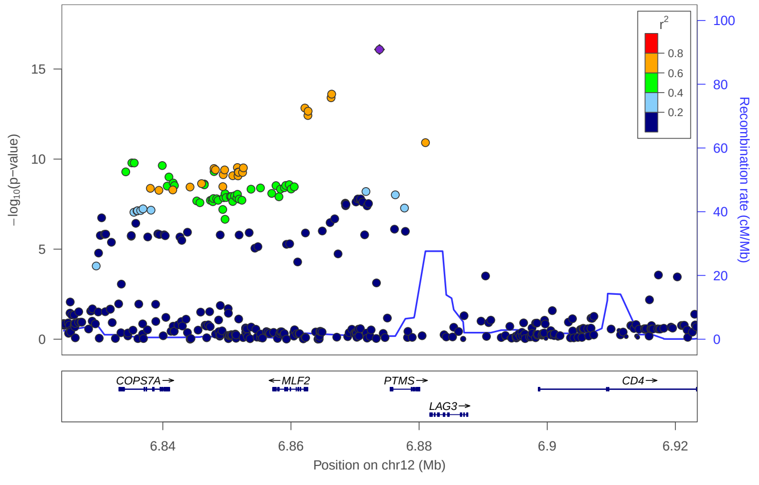

locus 387 rs547204129

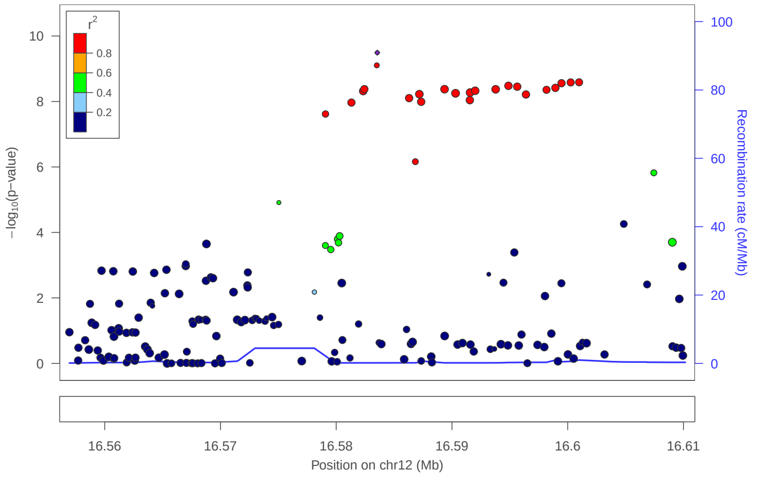


locus 388 rs10842059

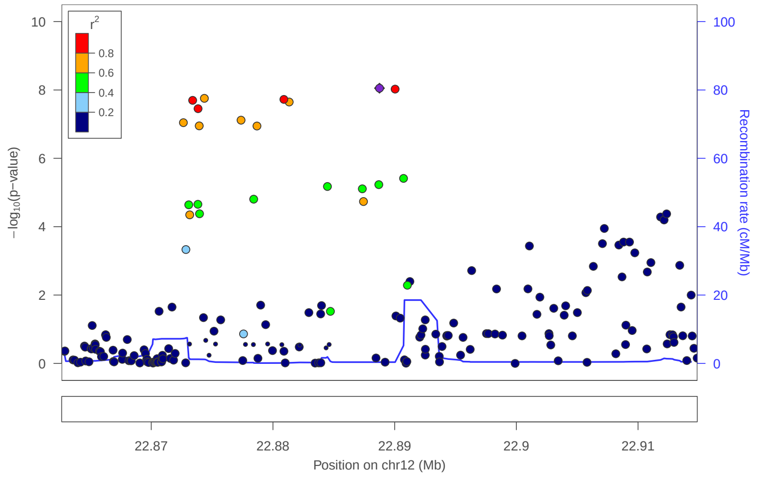


locus 389 rs11047436

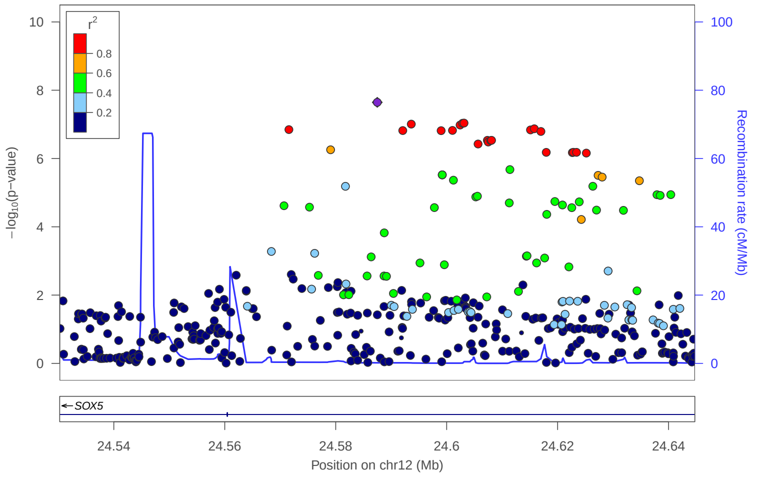


locus 390 rs12311881

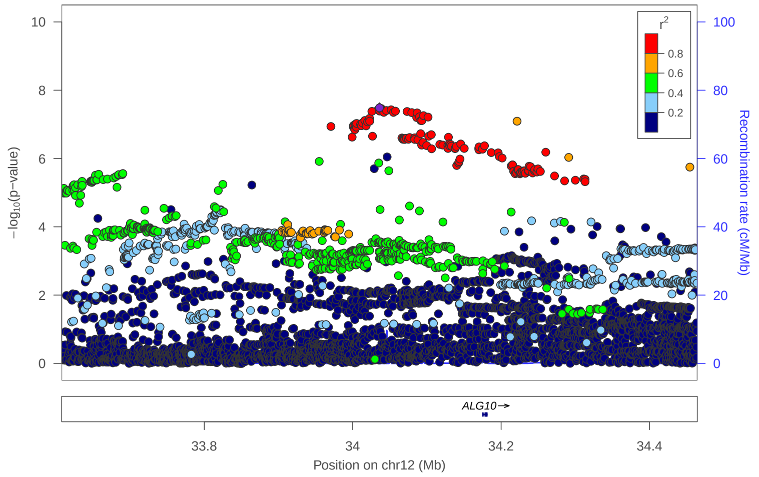


locus 391 rs10880104

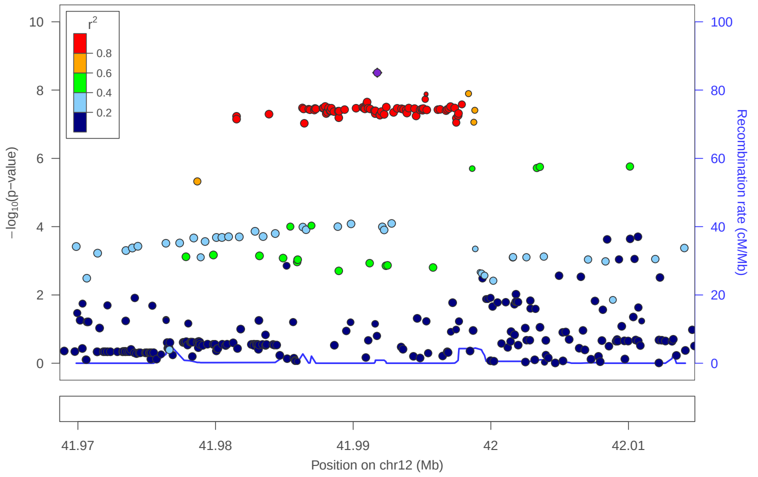


locus 392 rs12814145

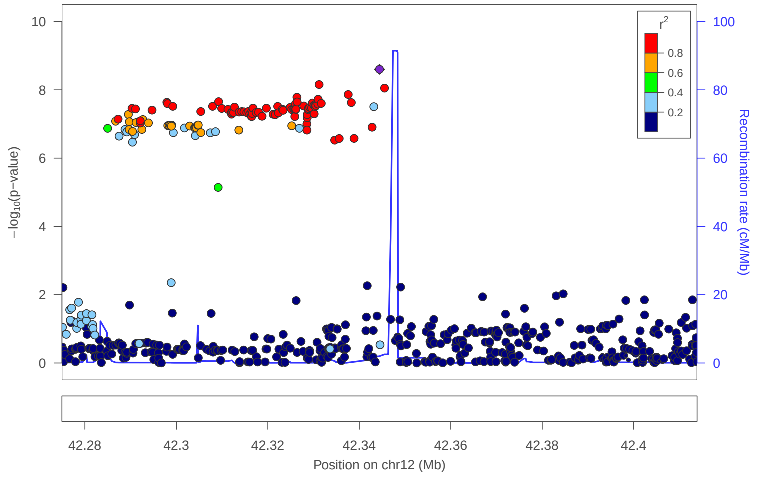


locus 393 rs1344829

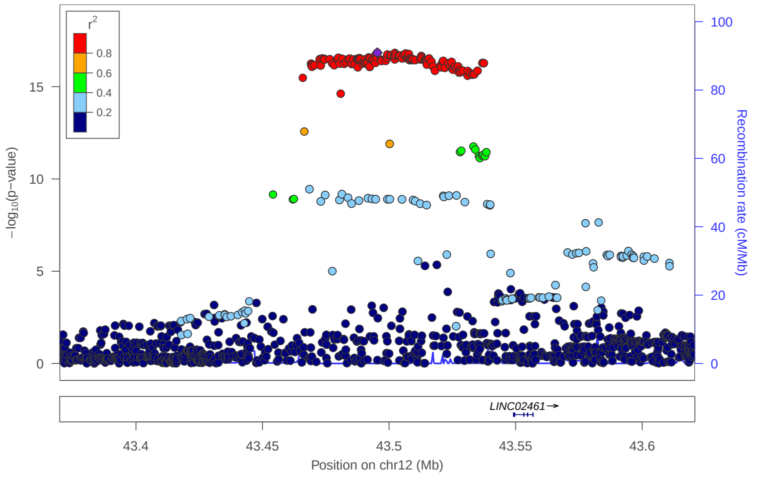


locus 394 rs12228775

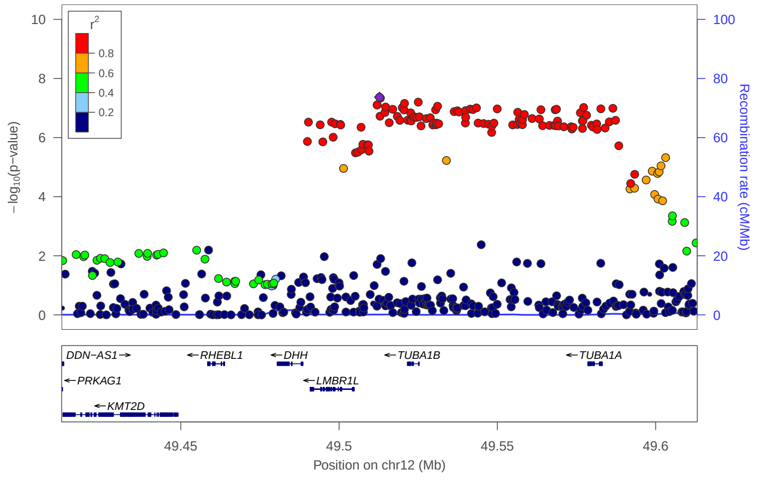

### locus 395 rs7308217

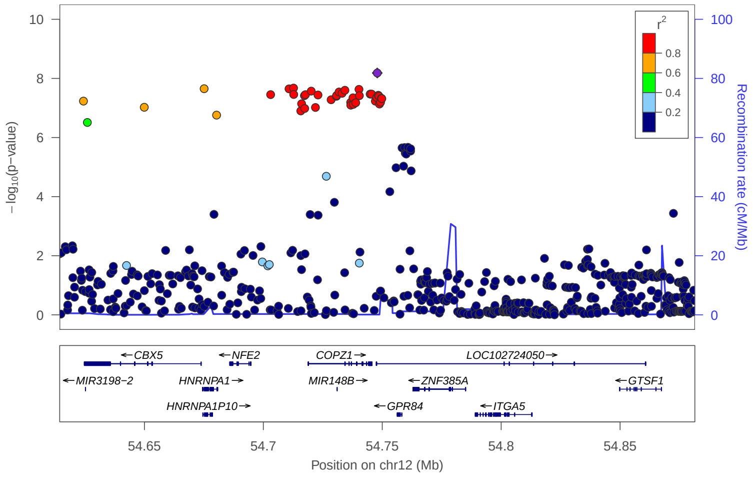


### locus 396 rs703848

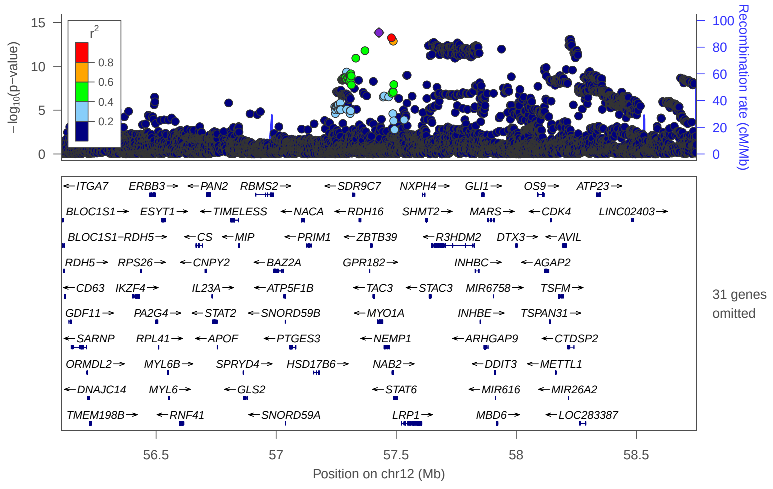


### locus 397 rs11173201

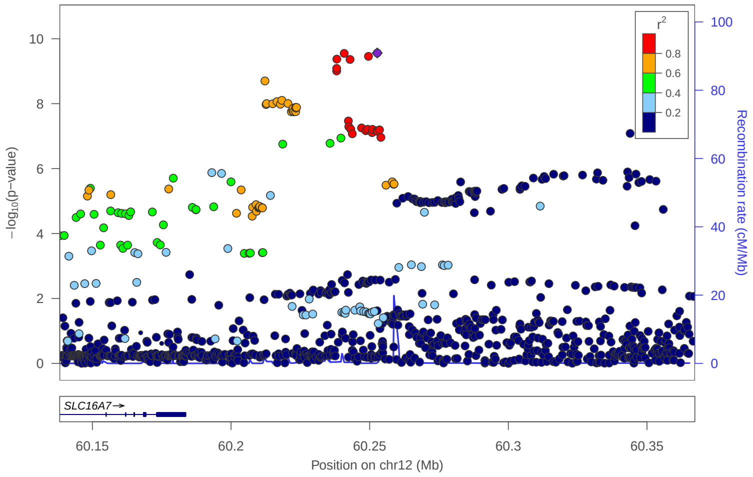


### locus 398 rs542132735

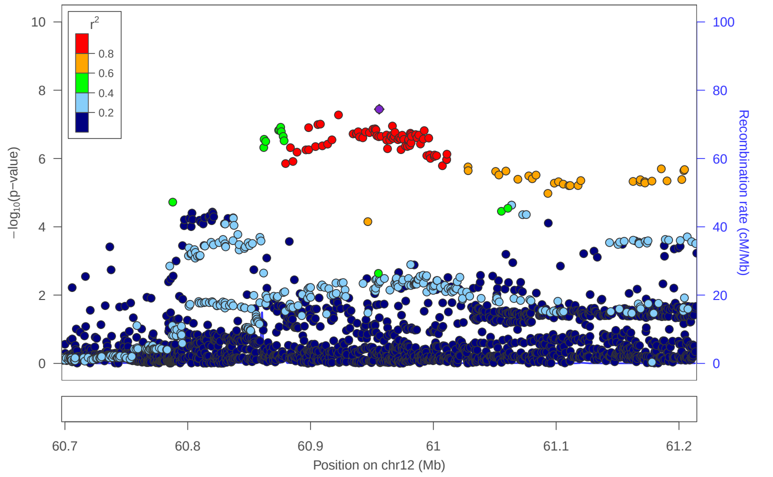


### locus 399 rs61921611

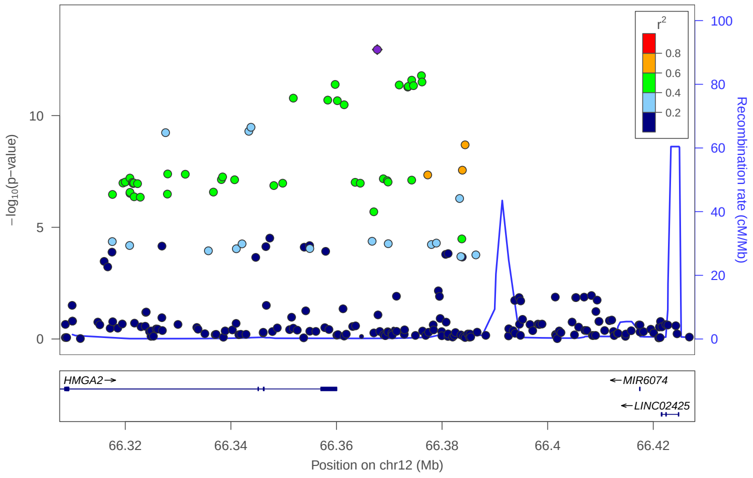


### locus 400 rs10400508

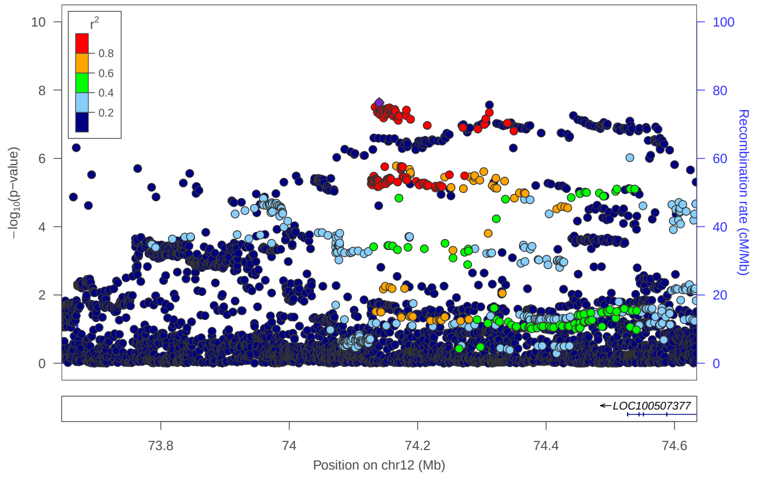


### locus 401 rs7980124

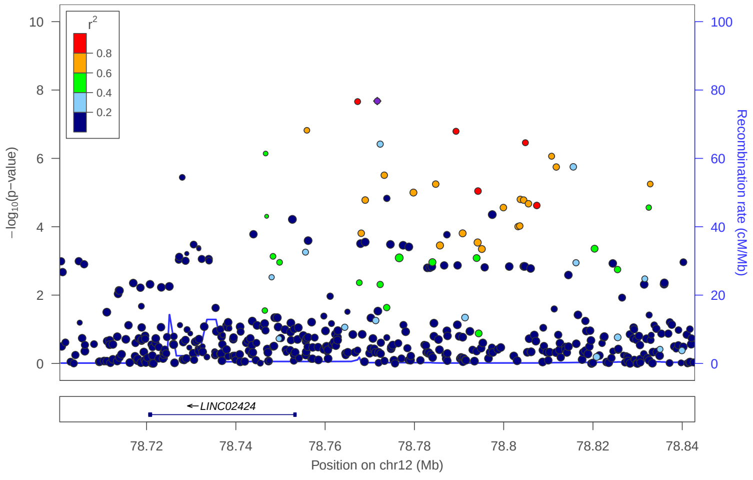


### locus 402 rs12580149

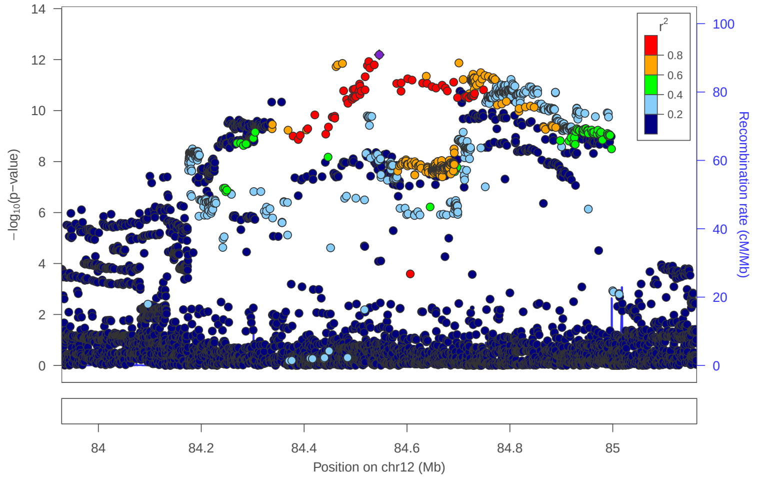

locus 403 rs797091

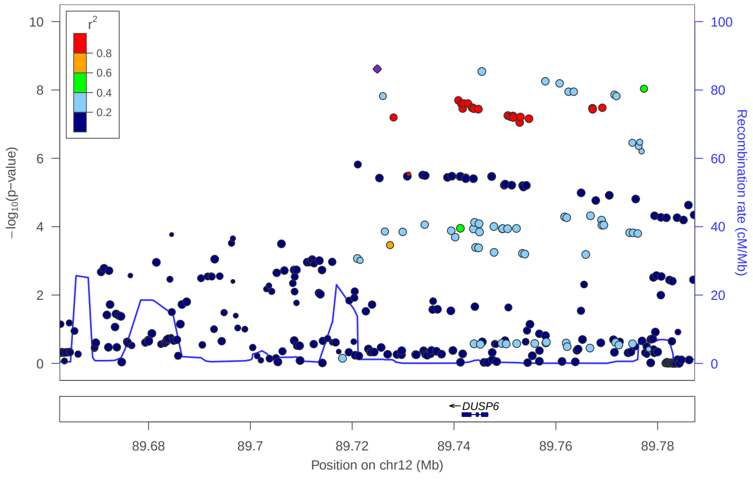


locus 404 rs9943753

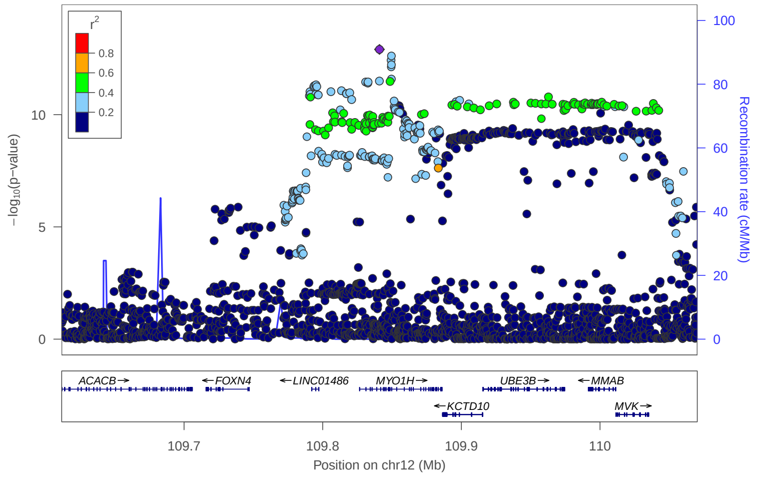


locus 405 rs7974266

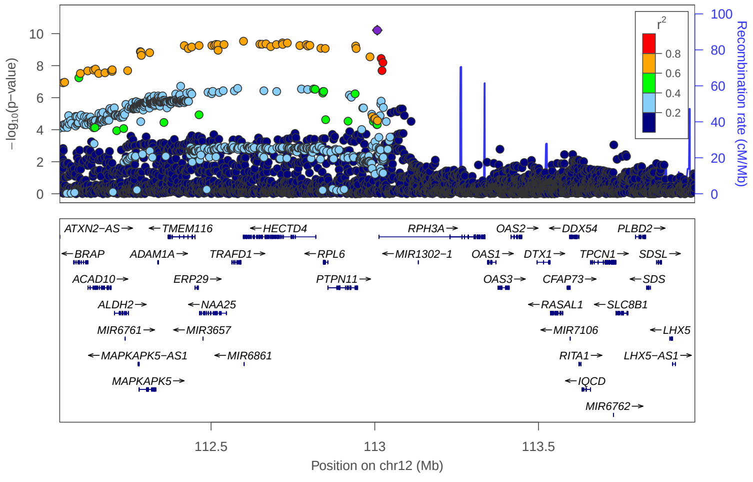


locus 406 rs1846644

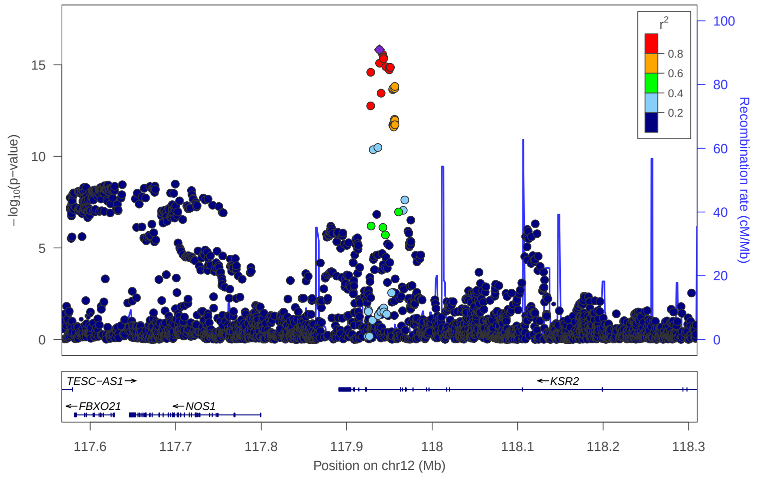


locus 407 rs2393330

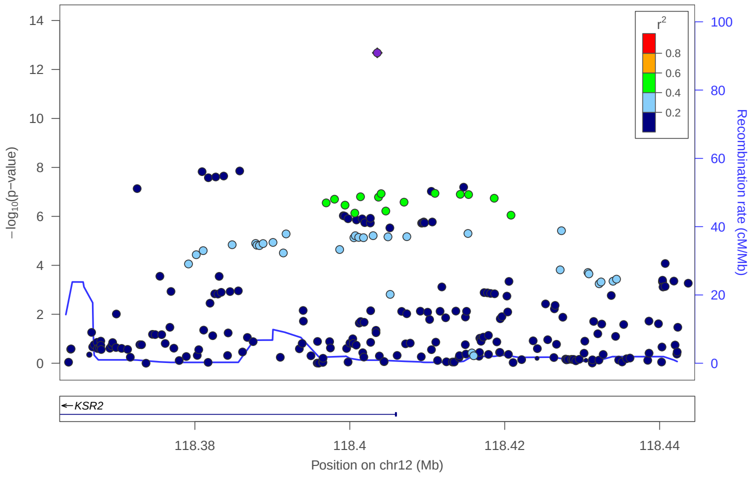


locus 408 rs585522

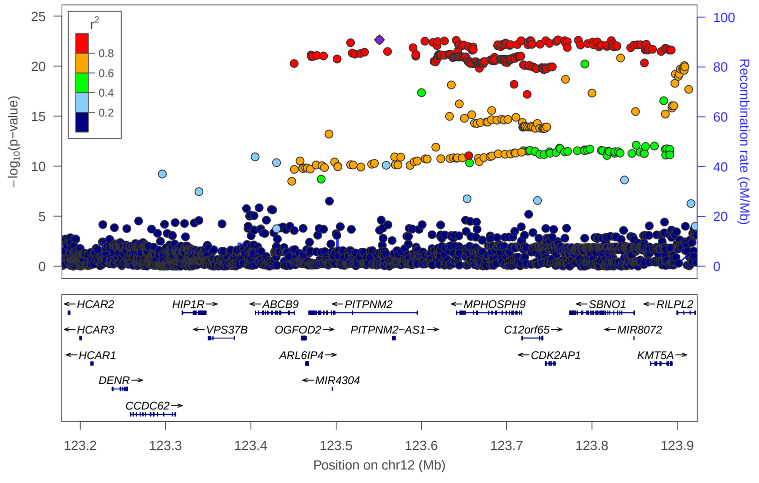


locus 409 rs7133378

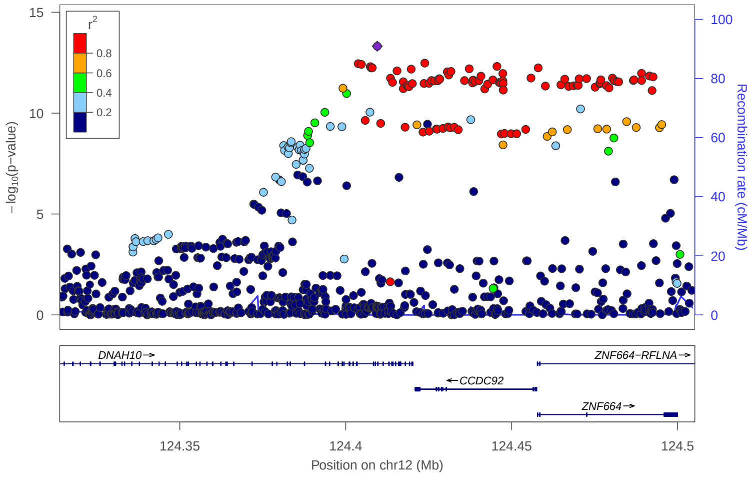


locus 410 rs9509284

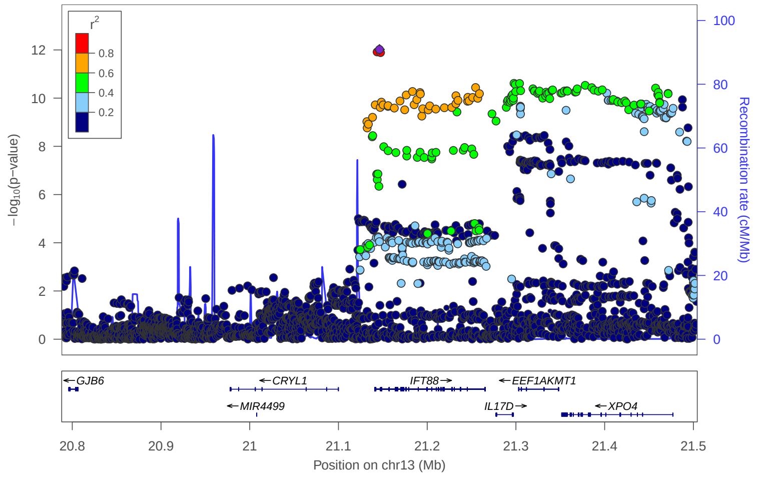

### locus 411 rs9543859

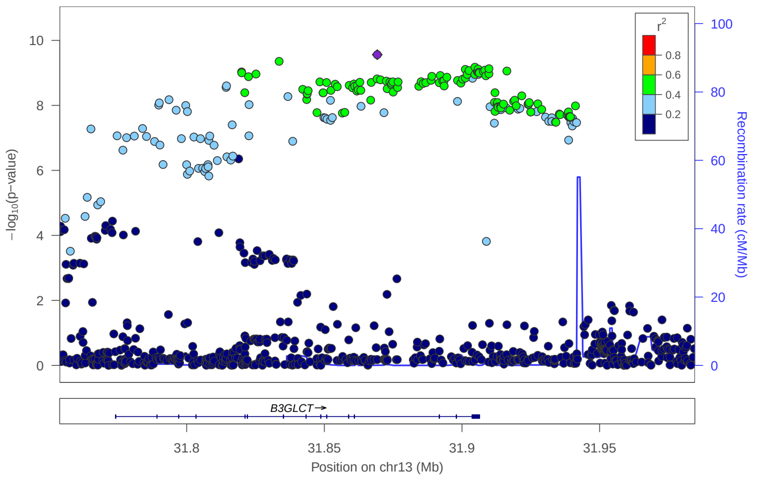


### locus 412 rs407004

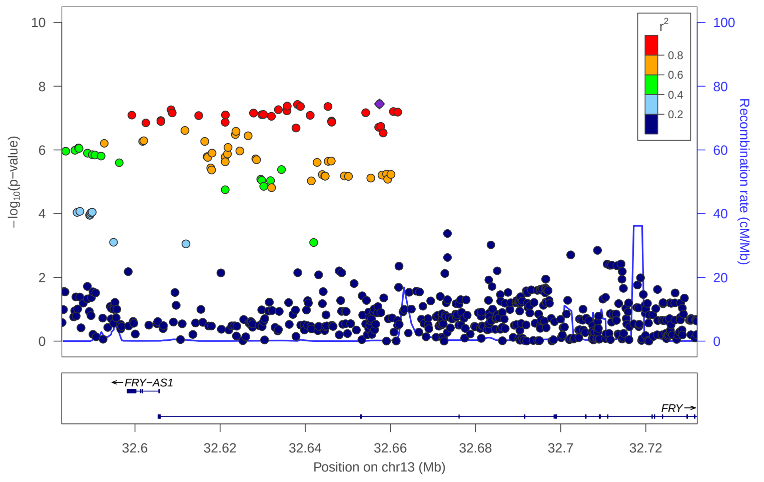


### locus 413 rs9315688

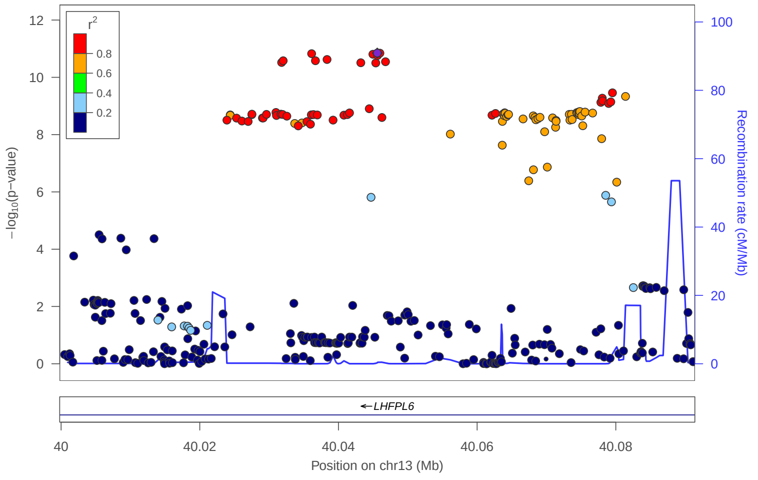


### locus 414 rs4304930

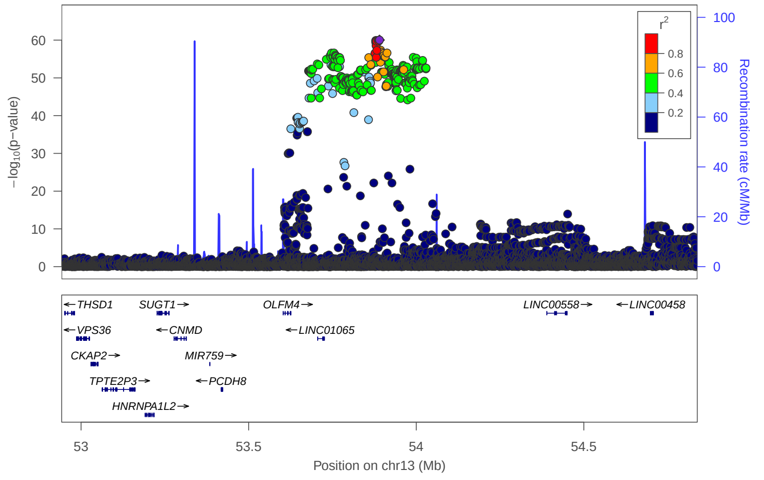


### locus 415 rs139963101

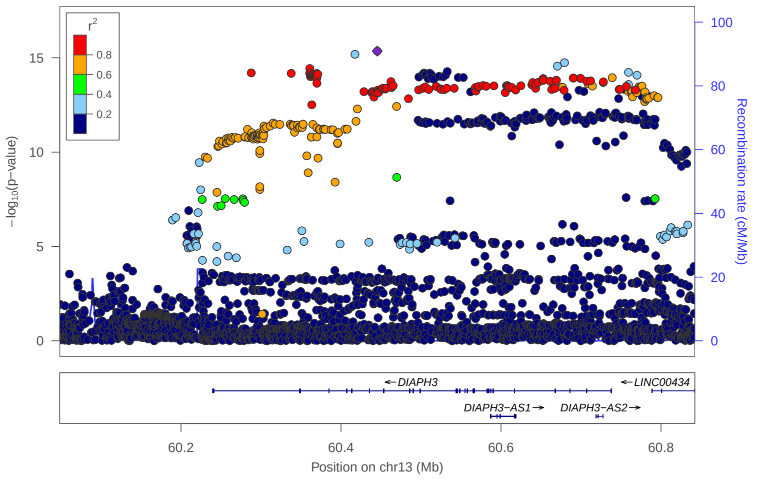


### locus 416 rs9563886

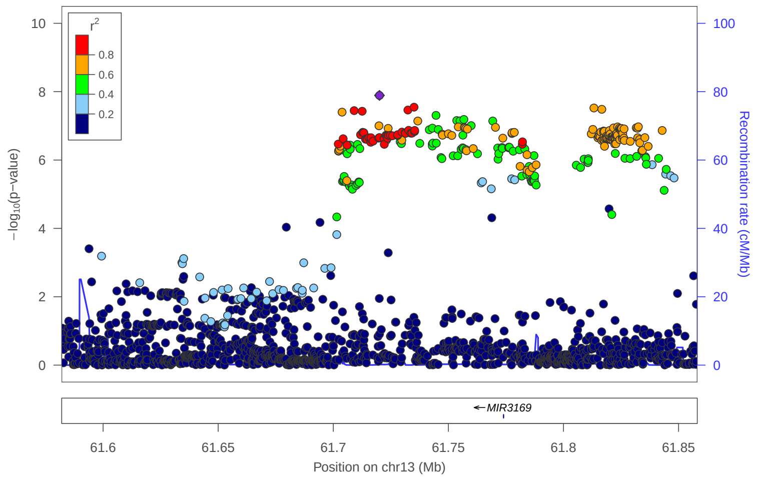


### locus 417 rs9317586

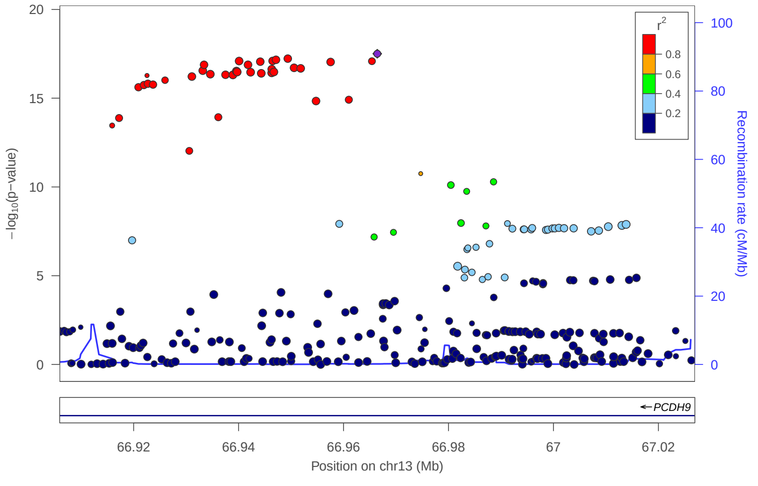


### locus 418 rs1925750

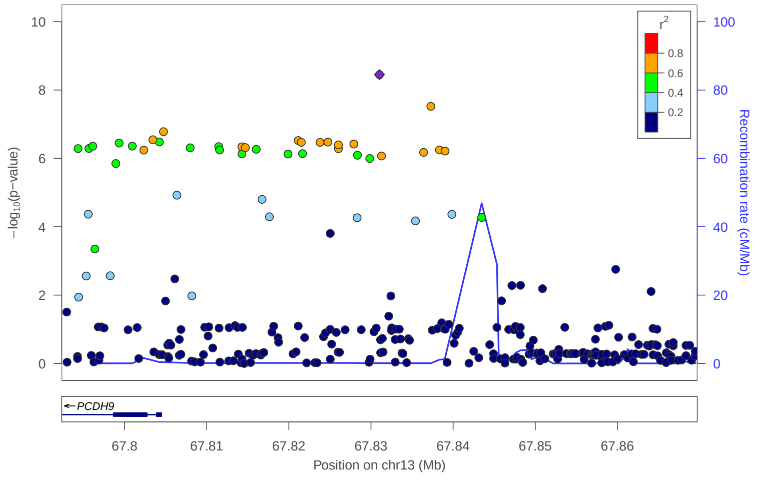

### locus 419 rs1876400

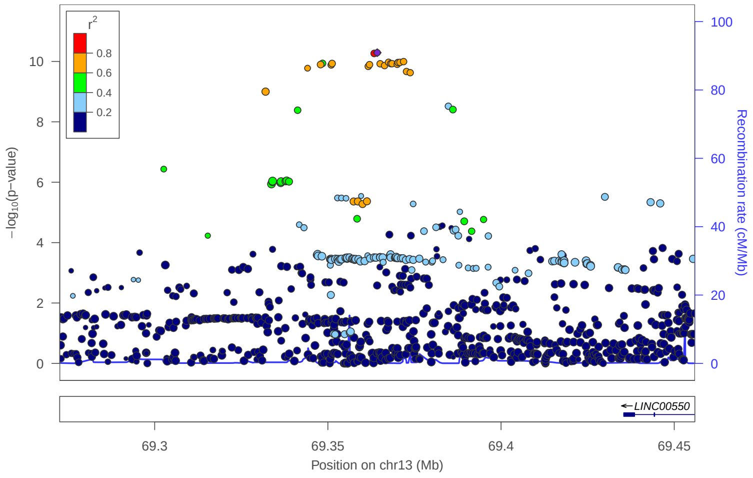


### locus 420 rs2782446

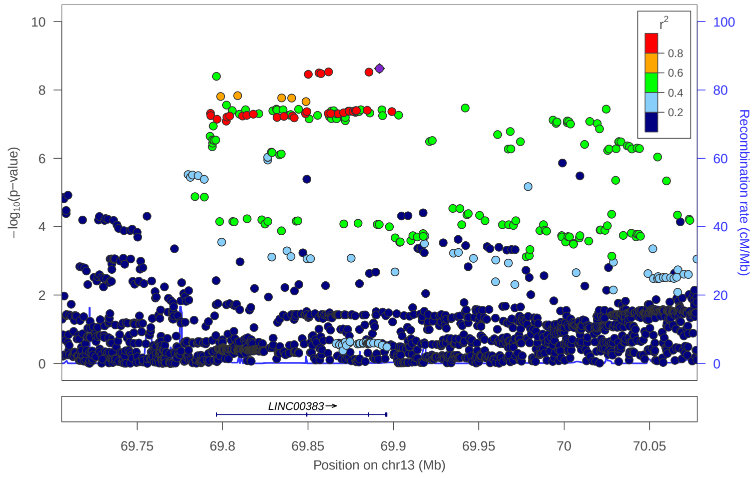


### locus 421 rs7338765

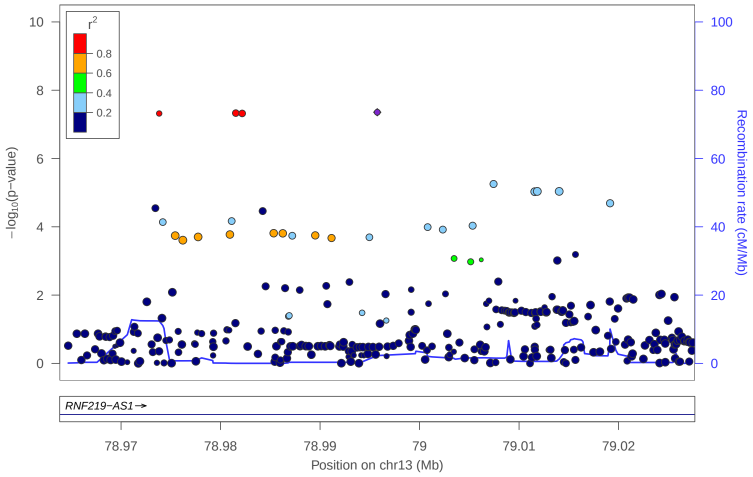


### locus 422 rs73532287

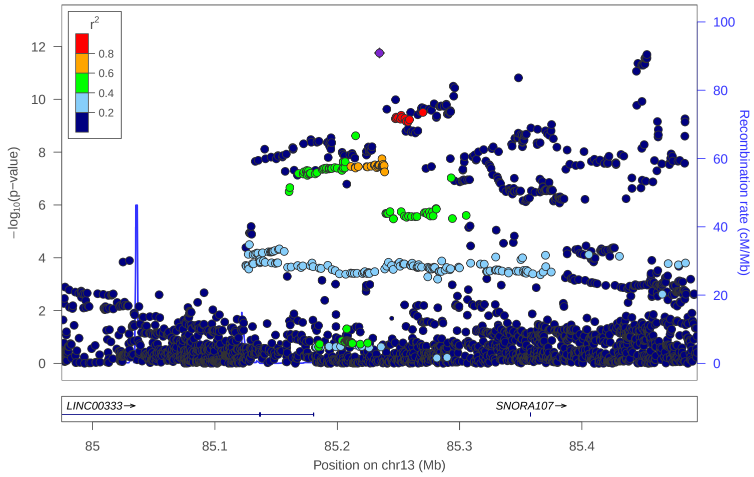


### locus 423 rs1411751

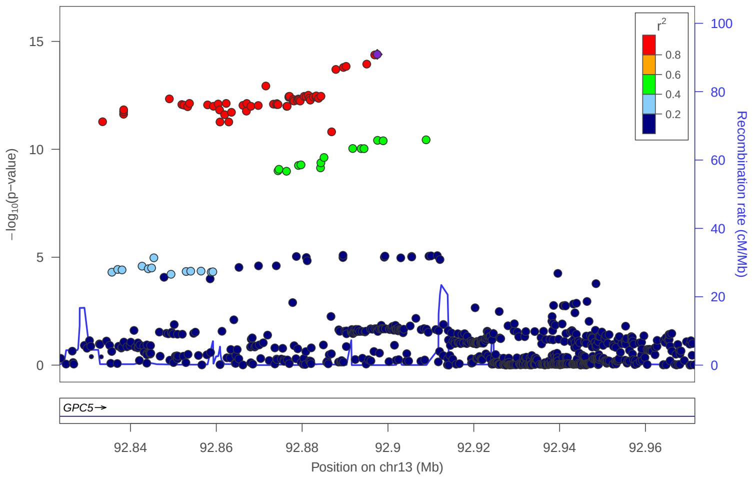


### locus 424 rs9561317

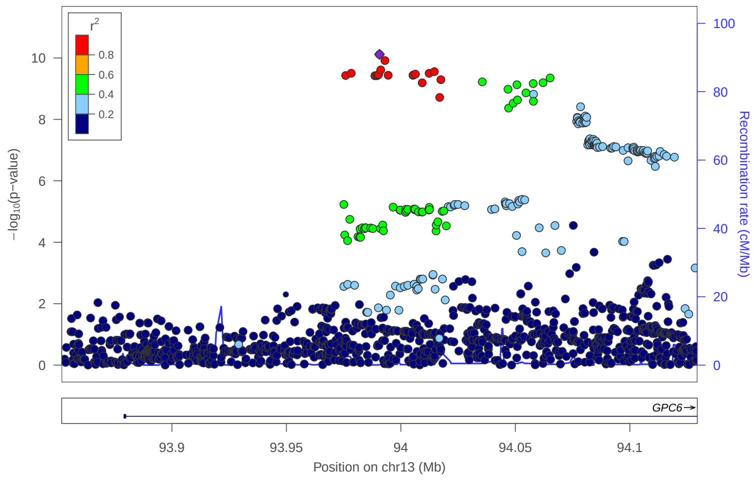


### locus 425 rs1925107

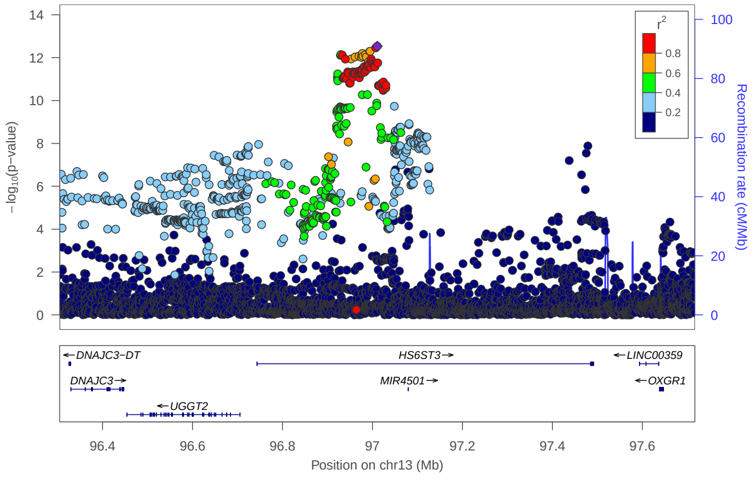


### locus 426 rs143612171

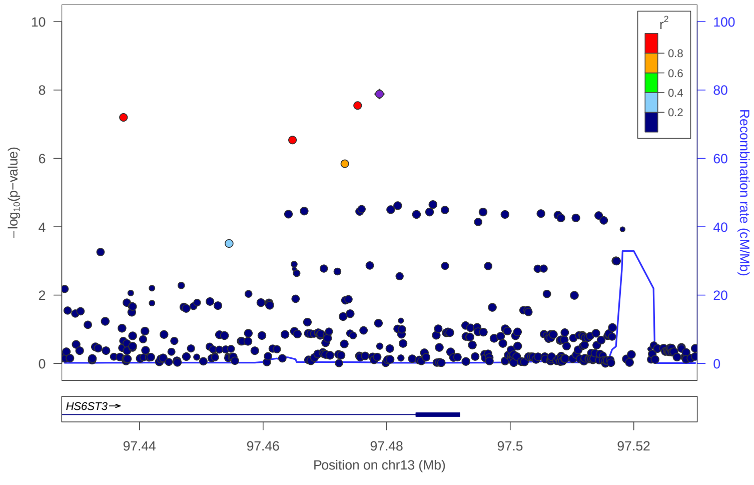

locus 427 rs4772087

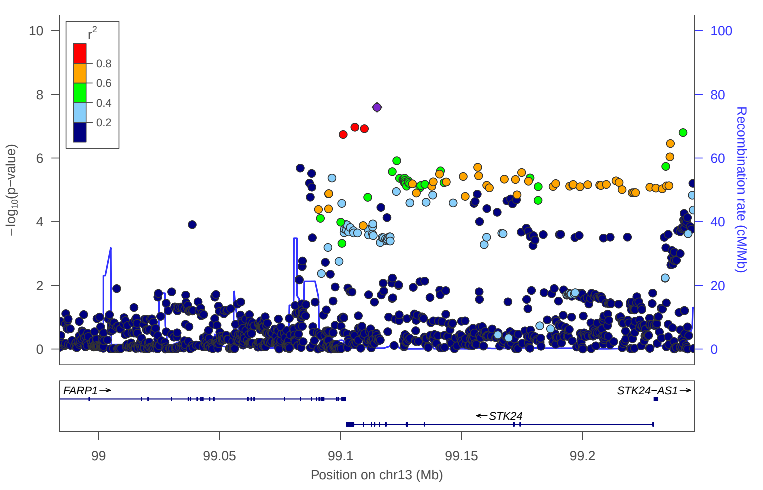


locus 428 rs61965119

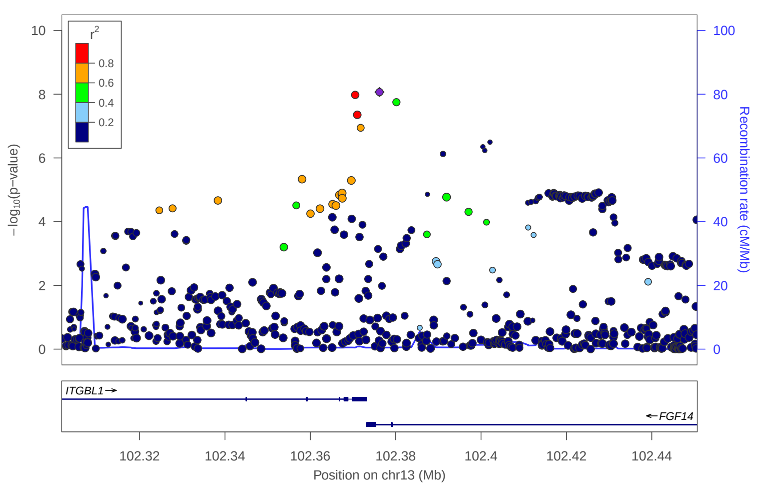


locus 429 rs17416209

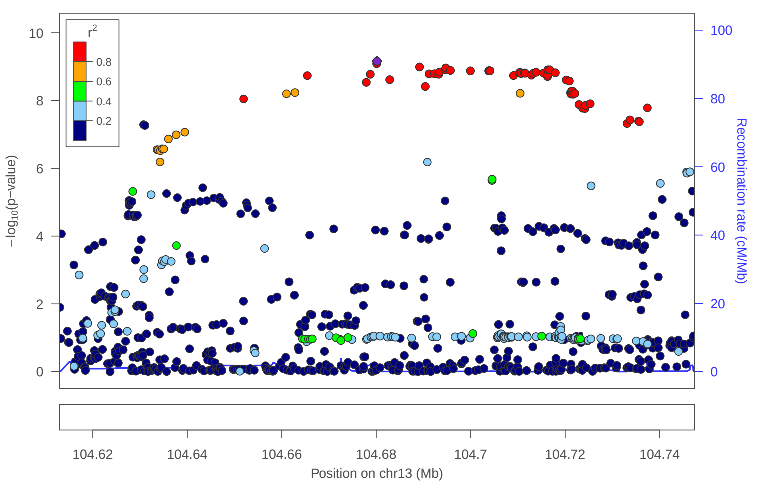


locus 430 rs275950

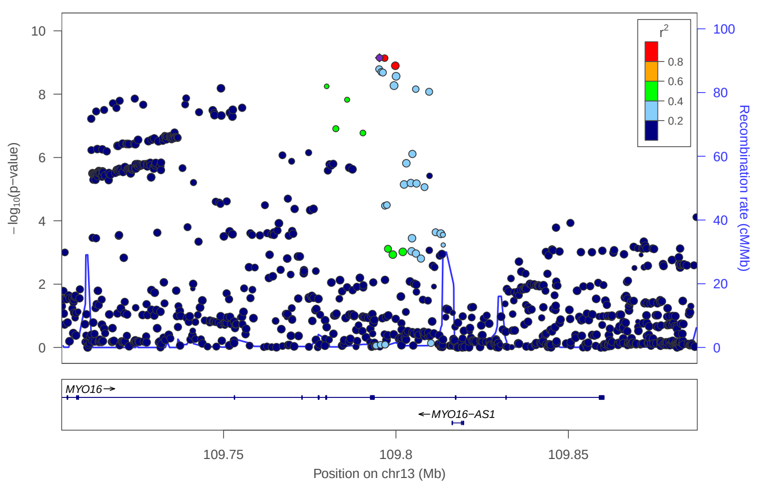


locus 431 rs1536053

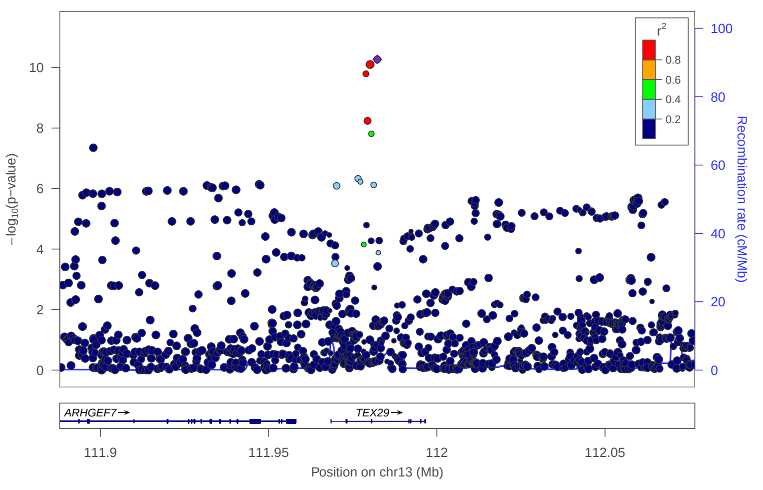


locus 432 rs61979497

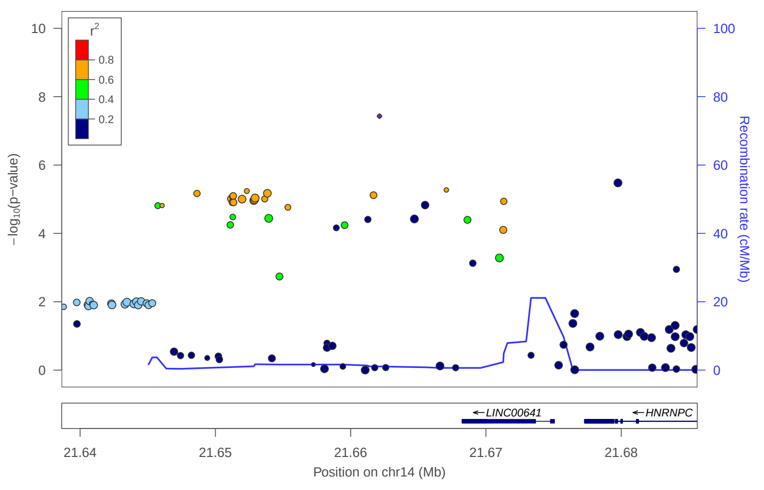


locus 433 rs4981170

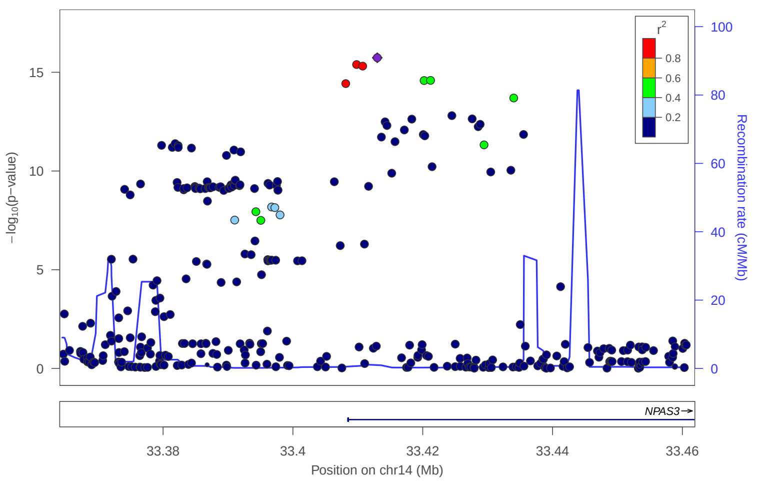


locus 434 rs10148610

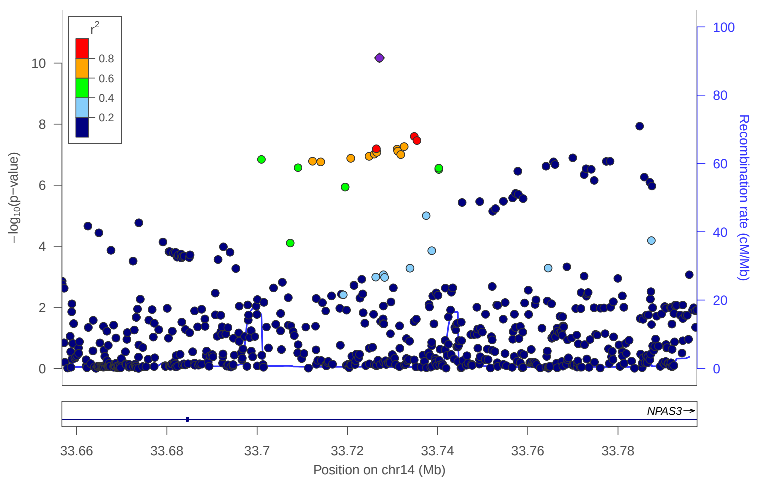

### locus 435 rs12893801

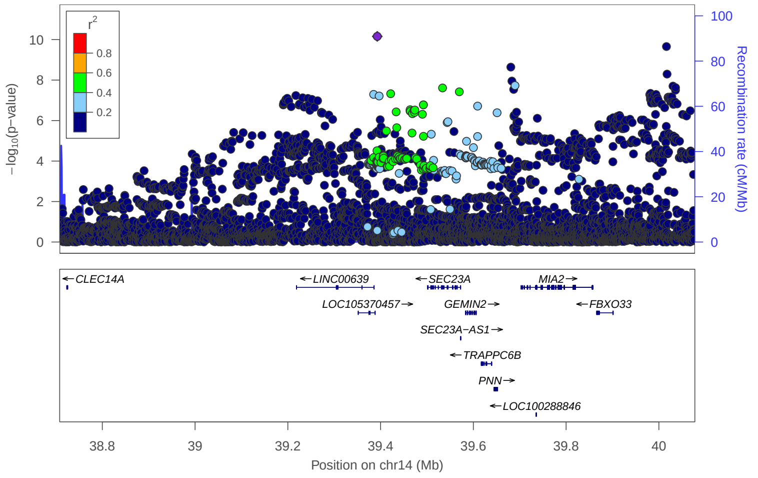


### locus 436 rs1584317

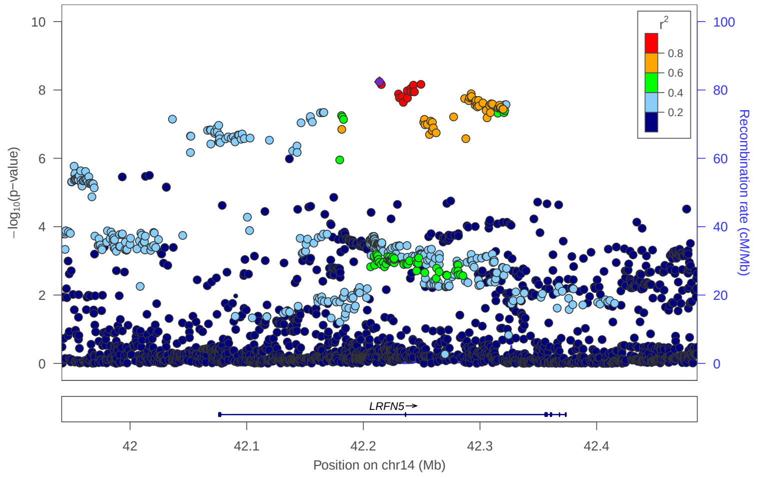


### locus 437 rs34772727

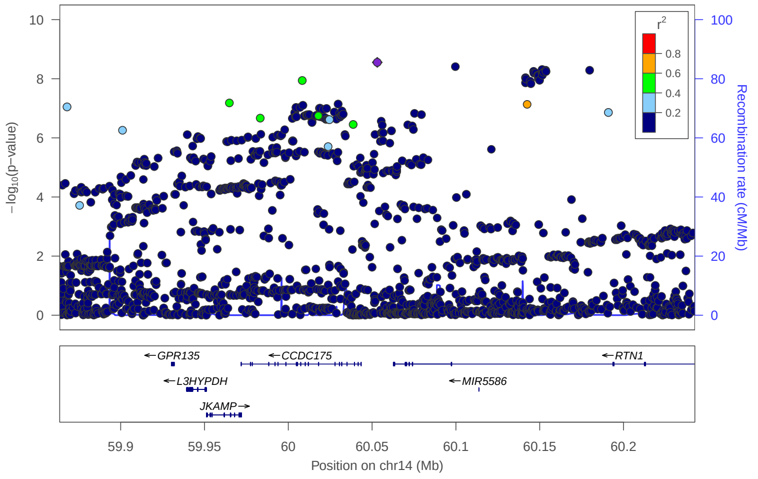


### locus 438 rs1007841

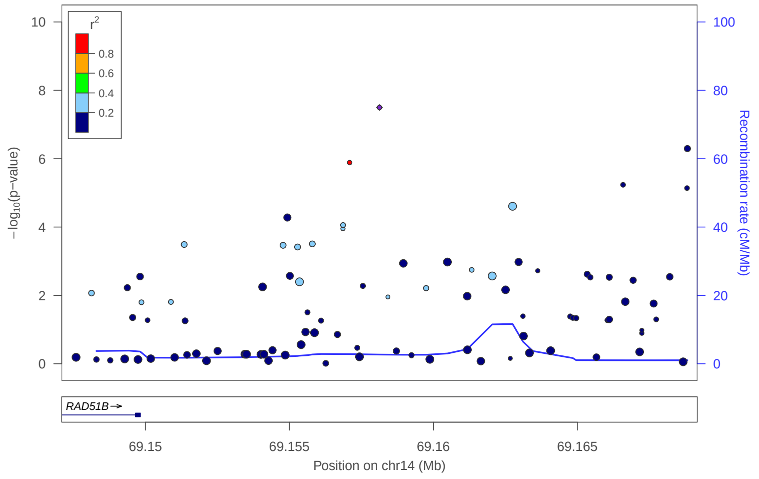


### locus 439 rs201404034

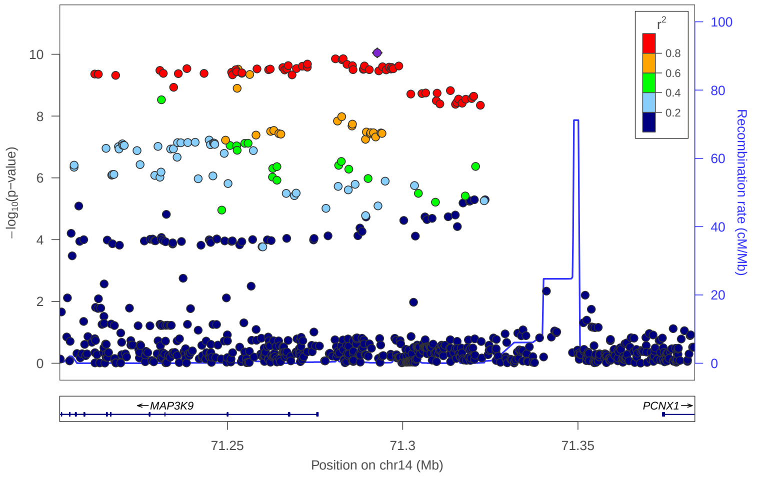


### locus 440 rs3783955

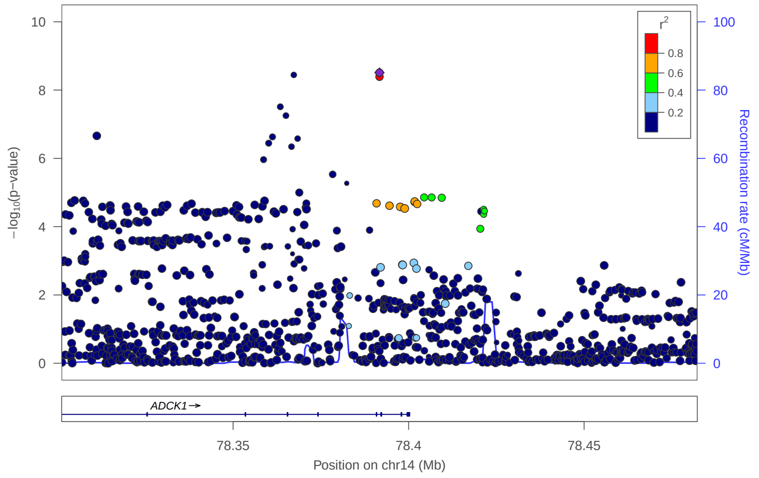


### locus 441 rs17836224

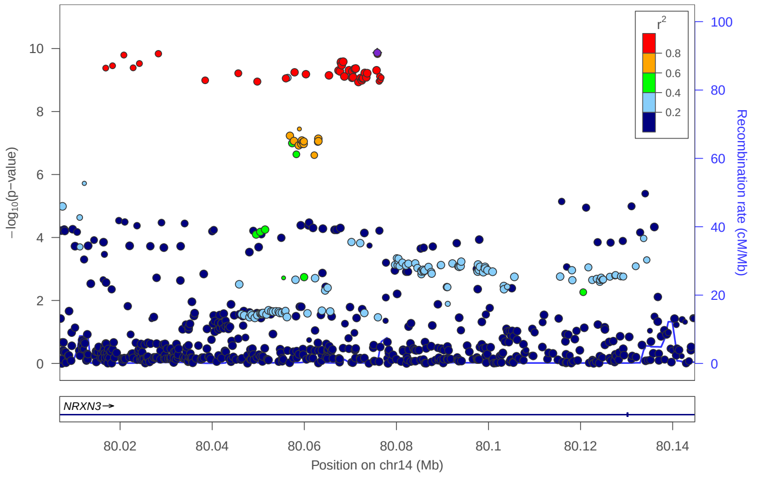


### locus 442 rs12431506

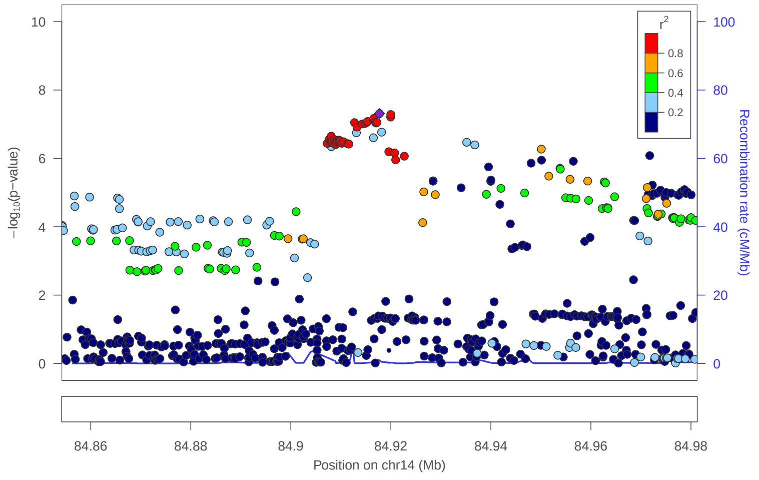

### locus 443 rs61981985

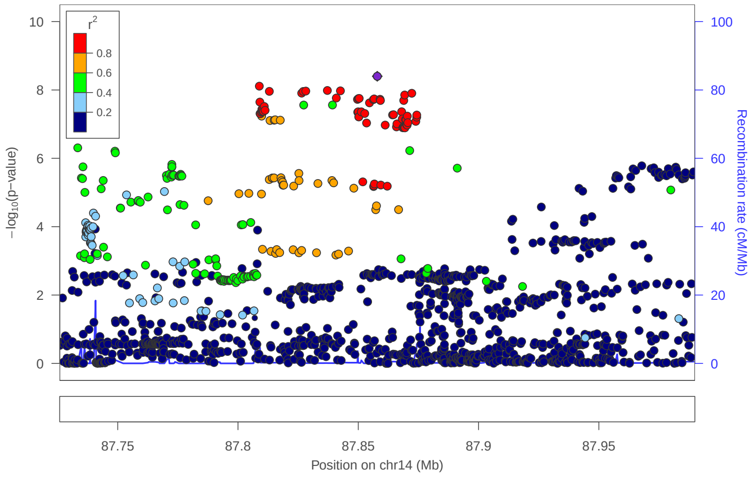


### locus 444 rs10137365

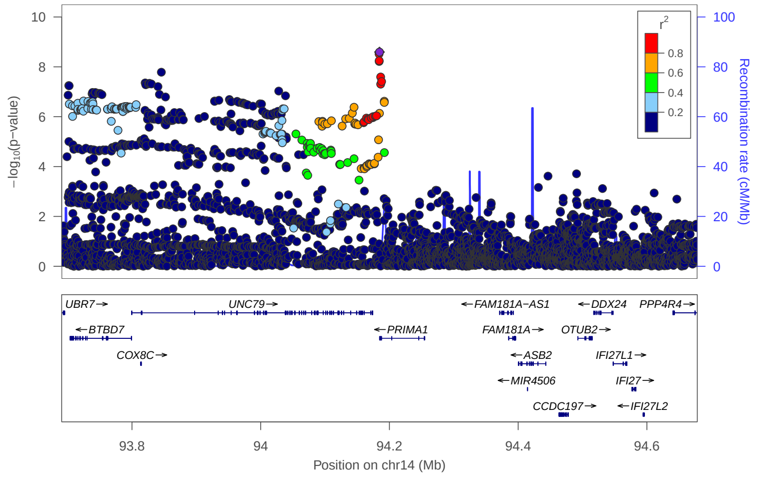


### locus 445 rs736929

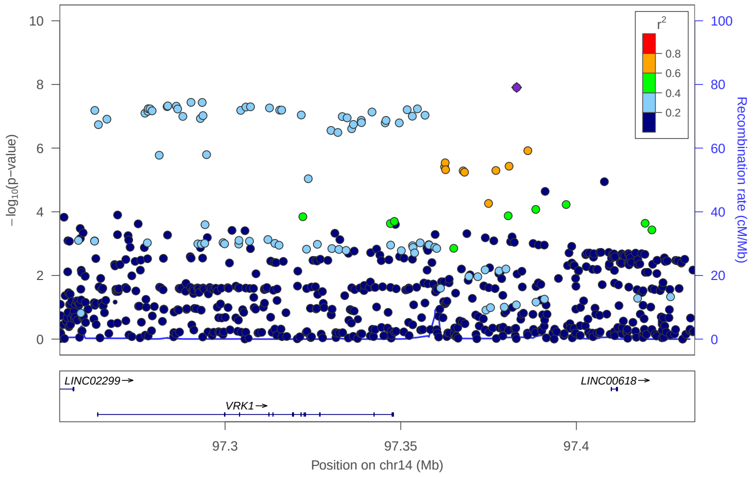


### locus 446 rs10143433

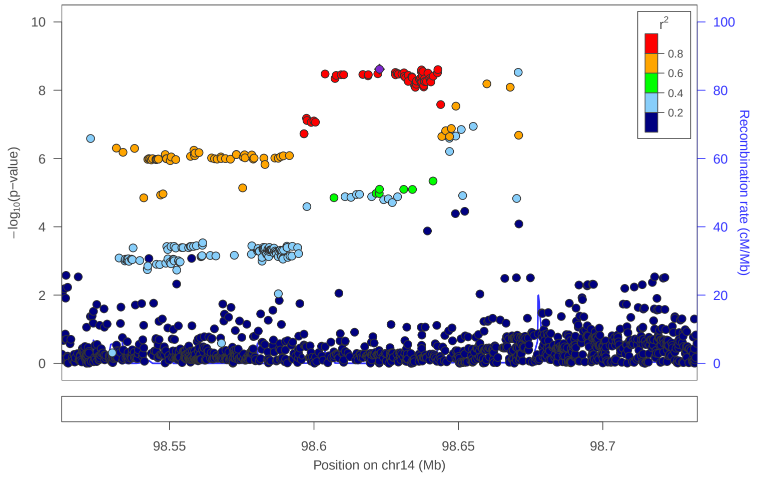


### locus 447 rs2664299

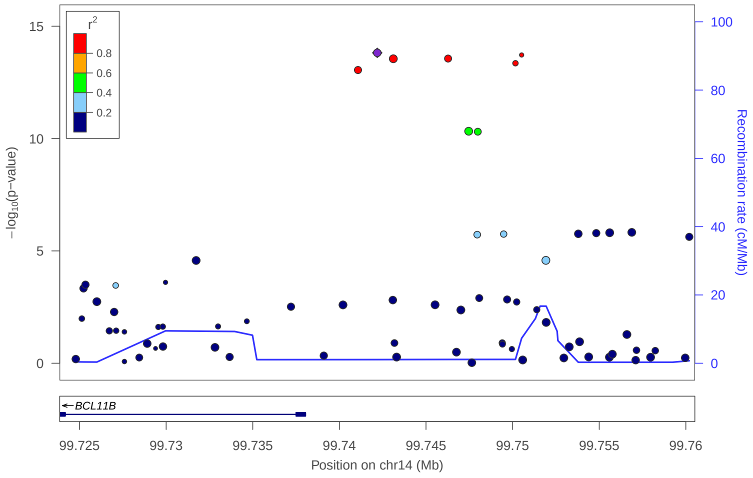


### locus 448 rs911324

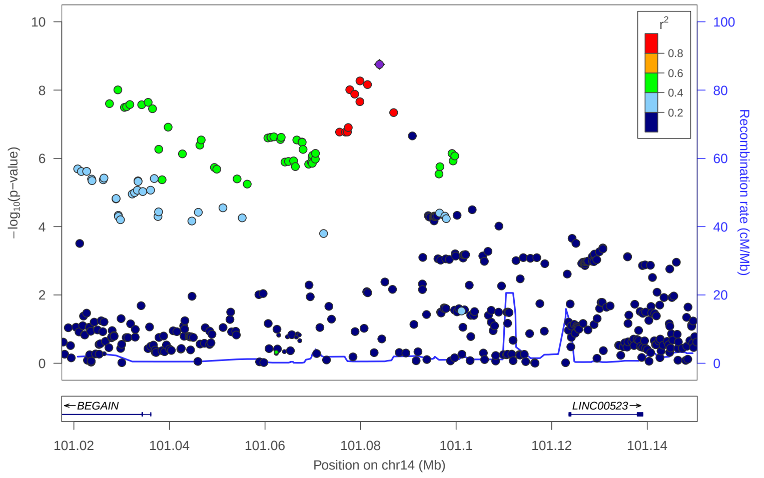


### locus 449 rs4906347

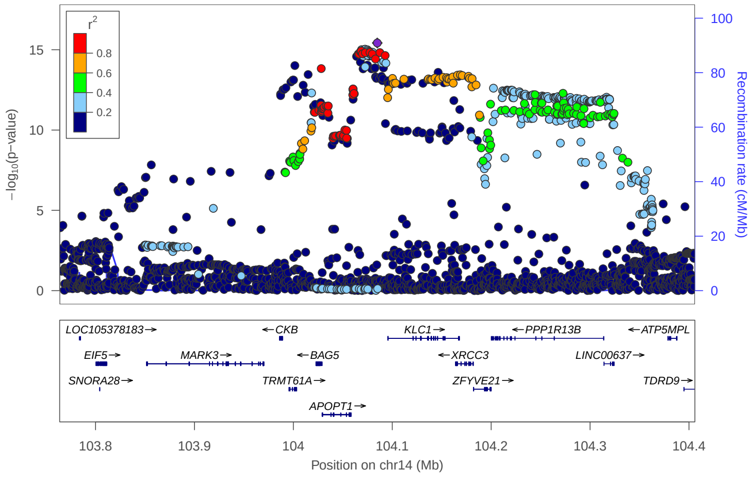


### locus 450 rs12324269

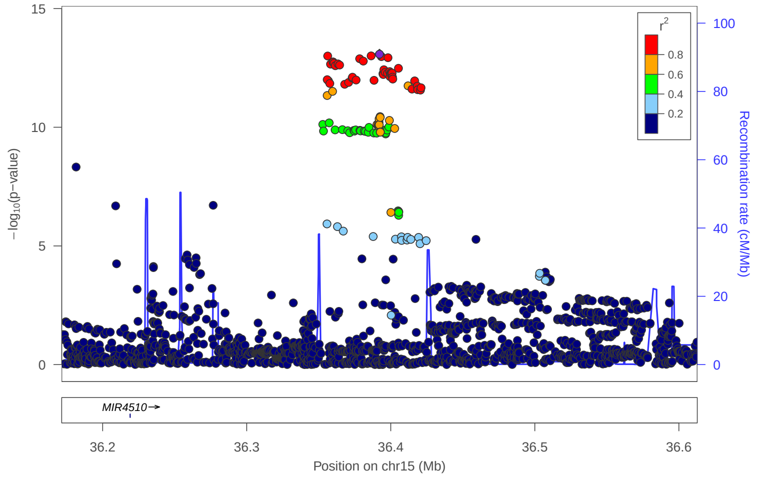

### locus 451 rs8036386

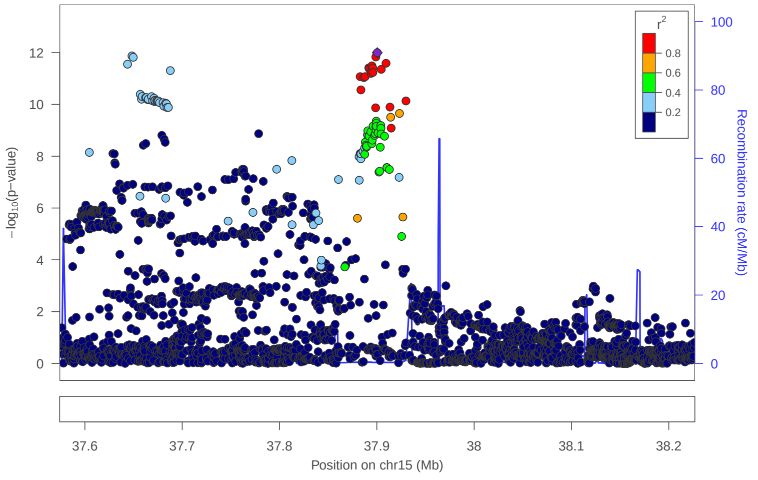


### locus 452 rs7173565

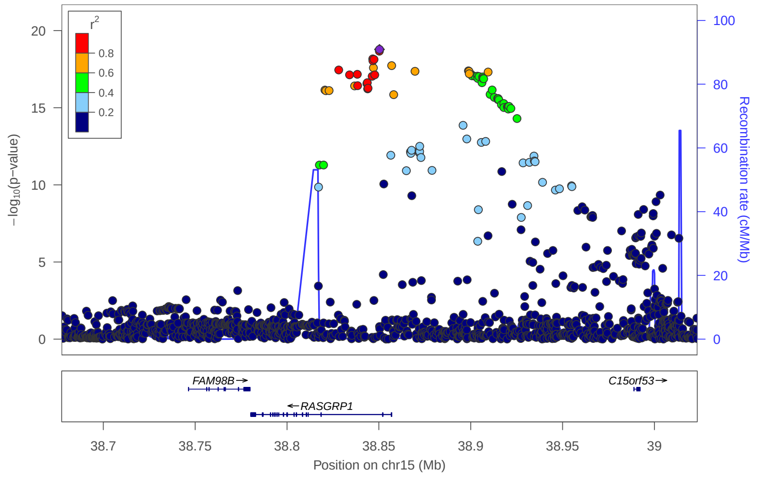


### locus 453 rs12594473

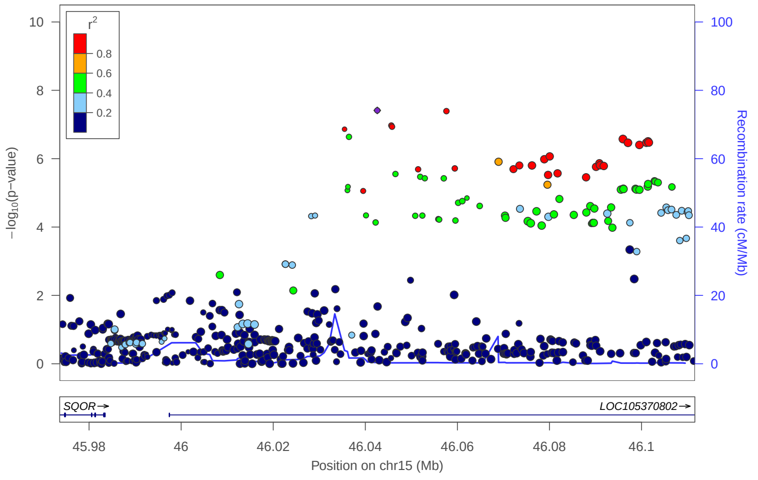


### locus 454 rs281222

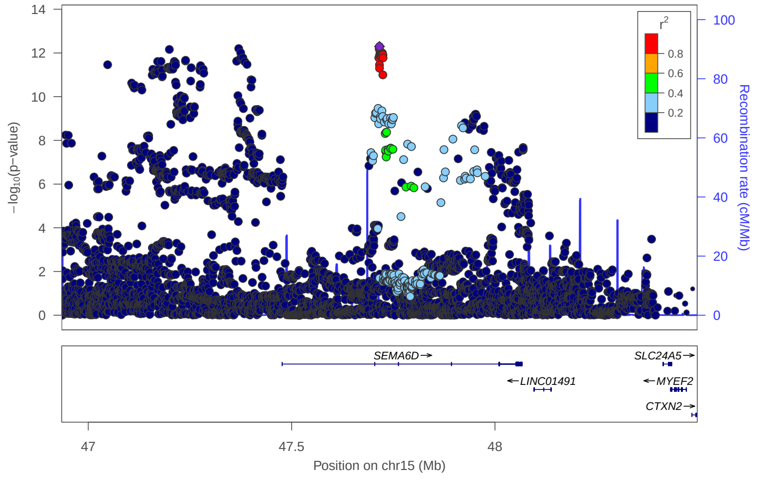


### locus 455 rs7183479

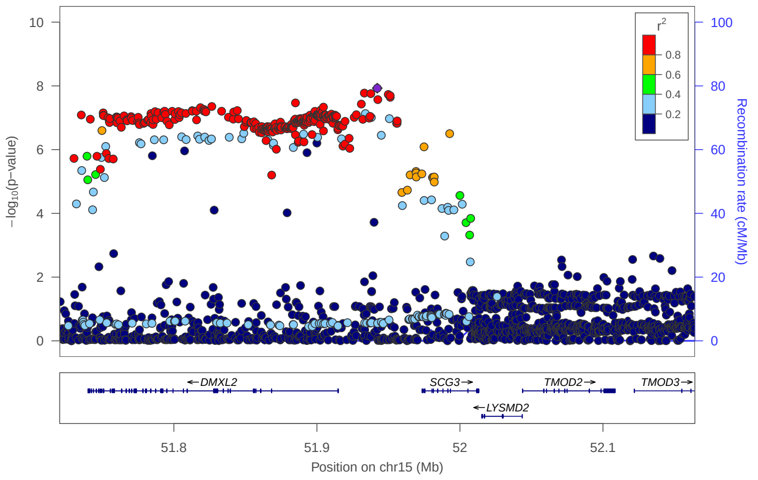


### locus 456 rs2456523

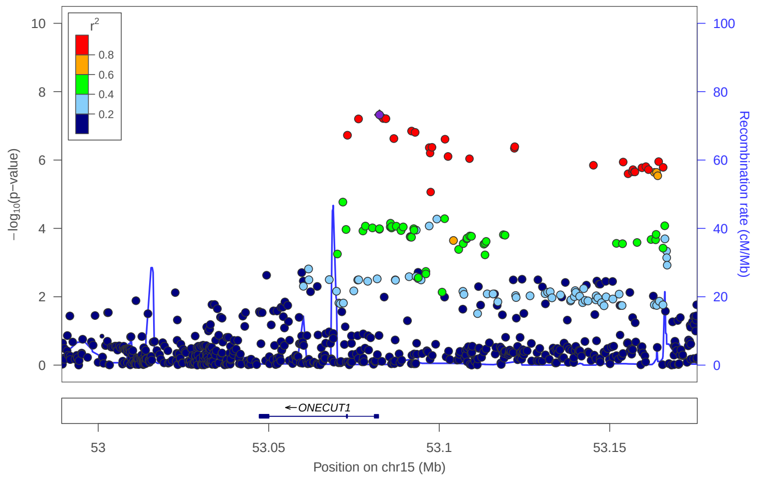


### locus 457 rs2615252

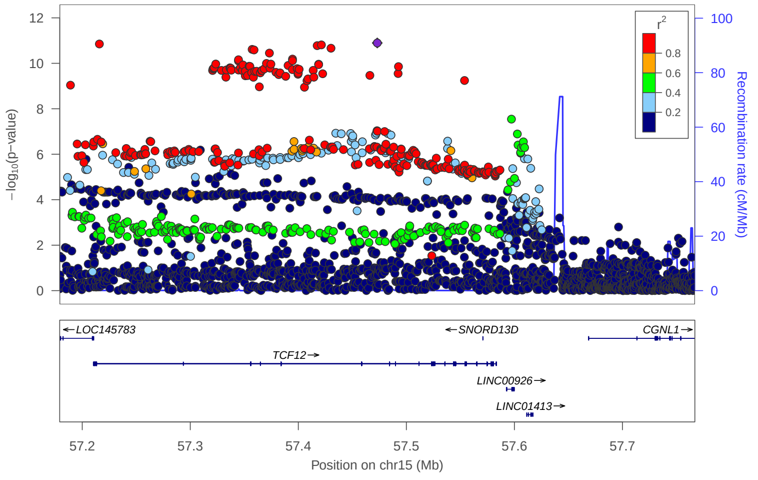


### locus 458 rs11856510

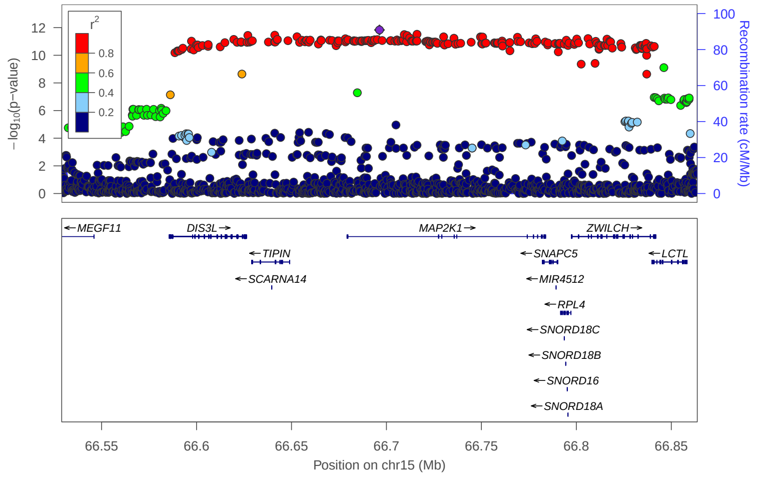

### locus 459 rs8025163

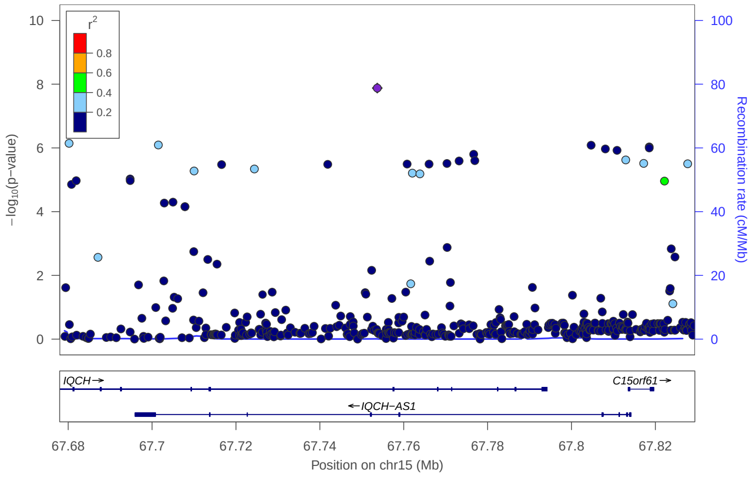


### locus 460 rs7162423

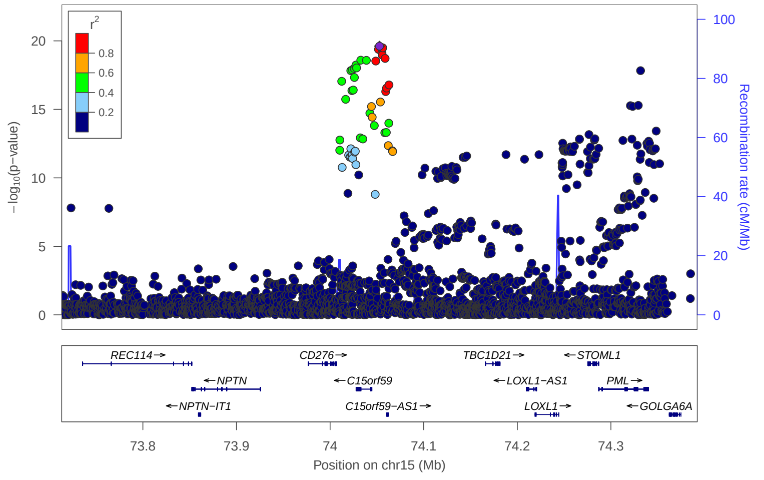


### locus 461 rs35095870

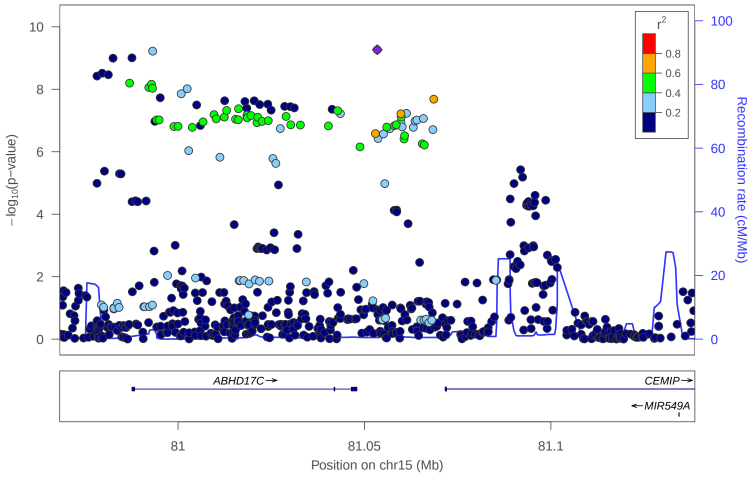


### locus 462 rs16976763

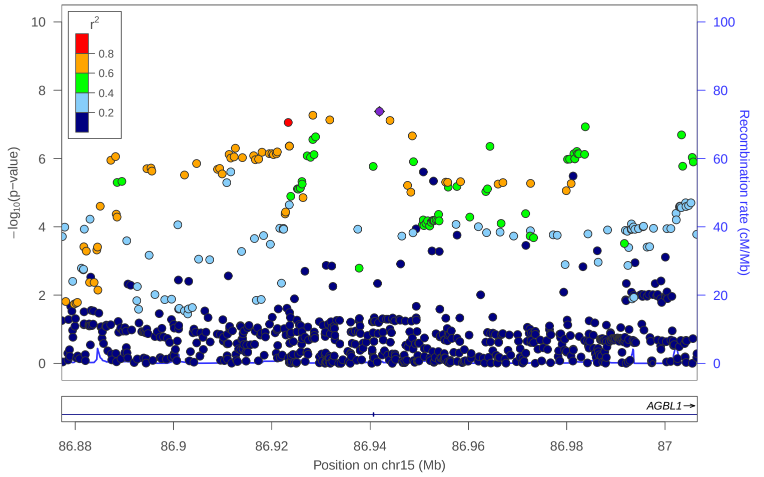


### locus 463 rs176644

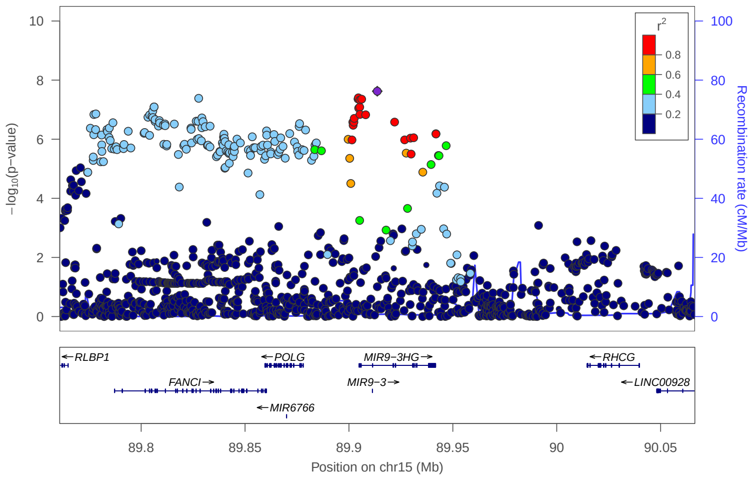


### locus 464 rs4702

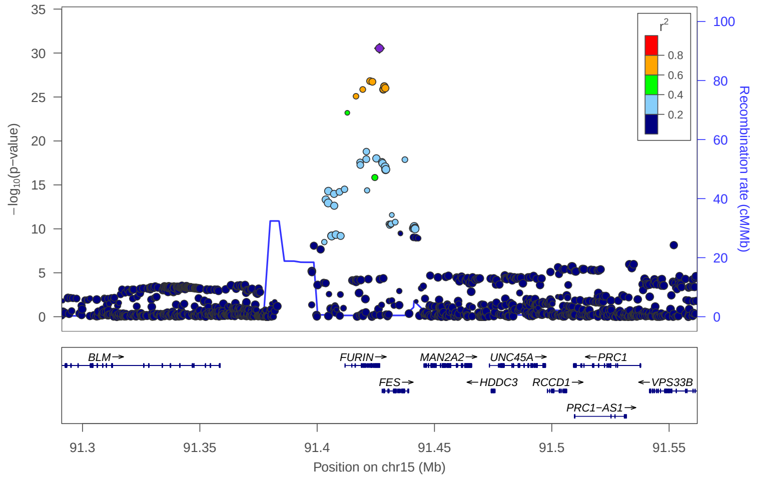


### locus 465 rs11858157

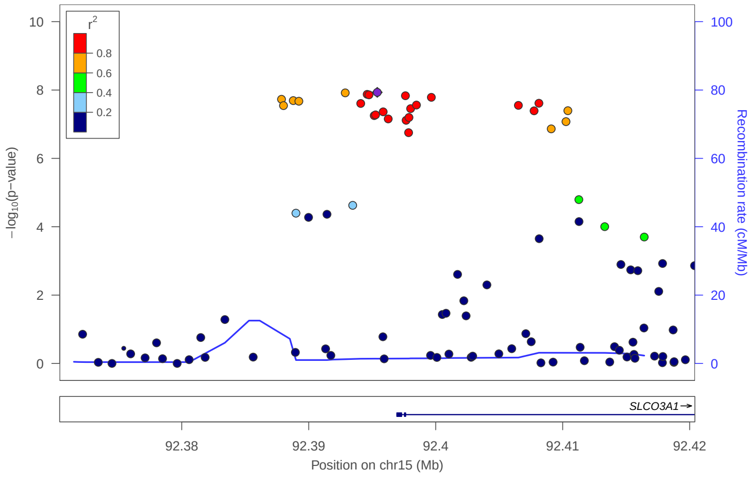


### locus 466 rs2575431

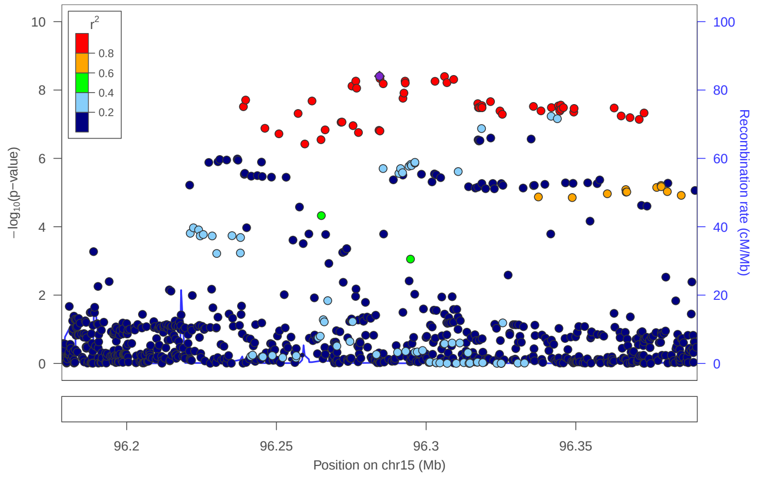

### locus 467 rs8026716

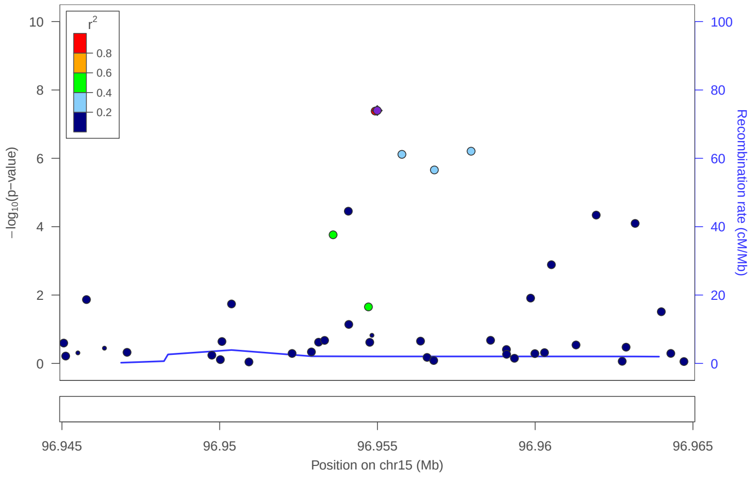


### locus 468 rs8028620

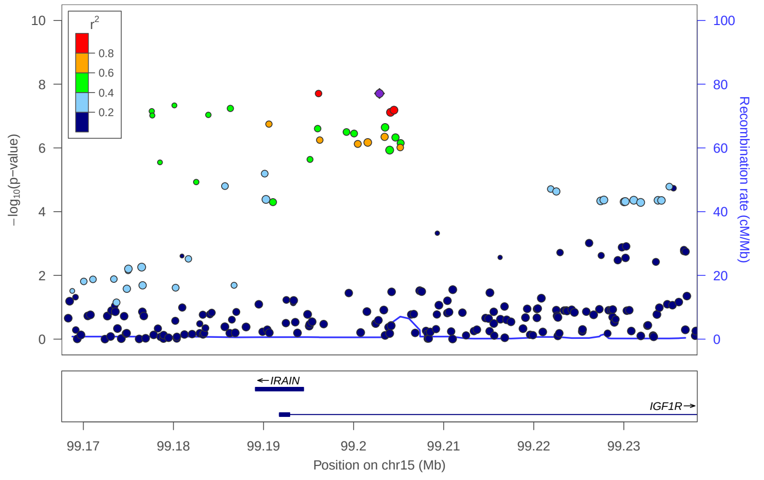


### locus 469 rs9936879

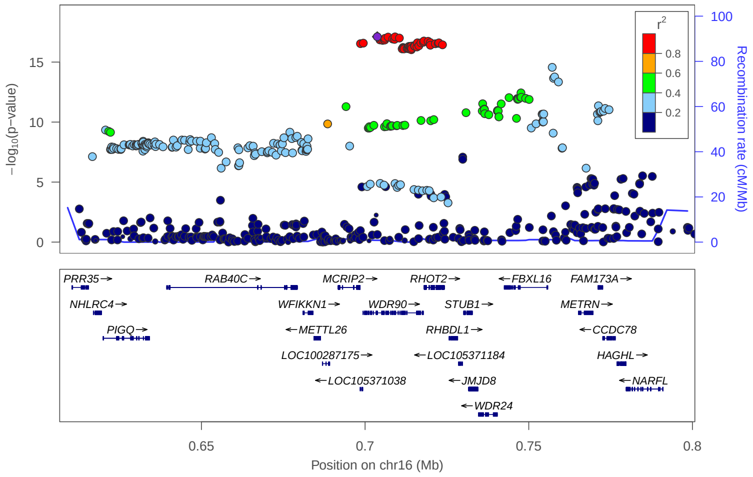


### locus 470 rs11461440

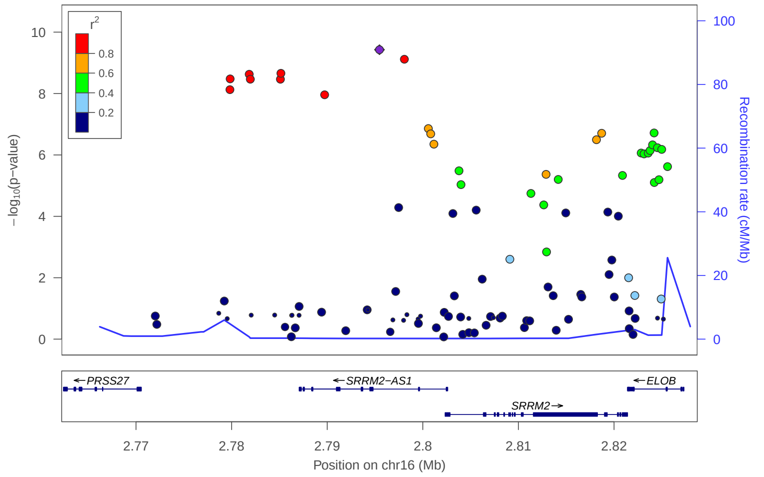


### locus 471 rs11077334

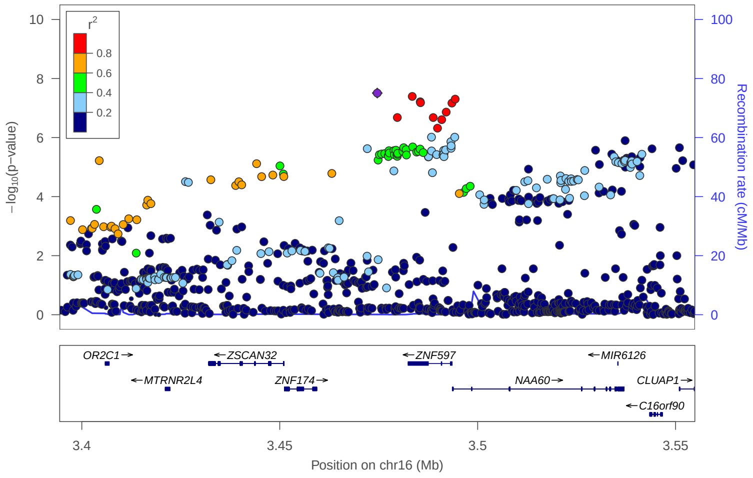


### locus 472 rs12928387

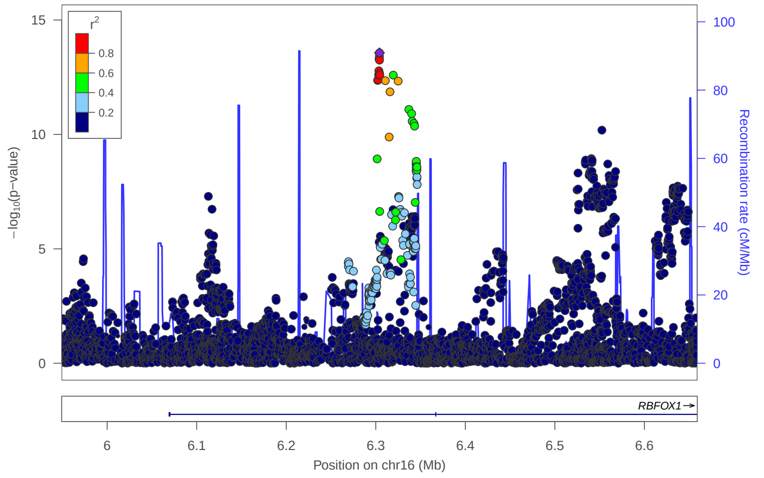


### locus 473 rs12924275

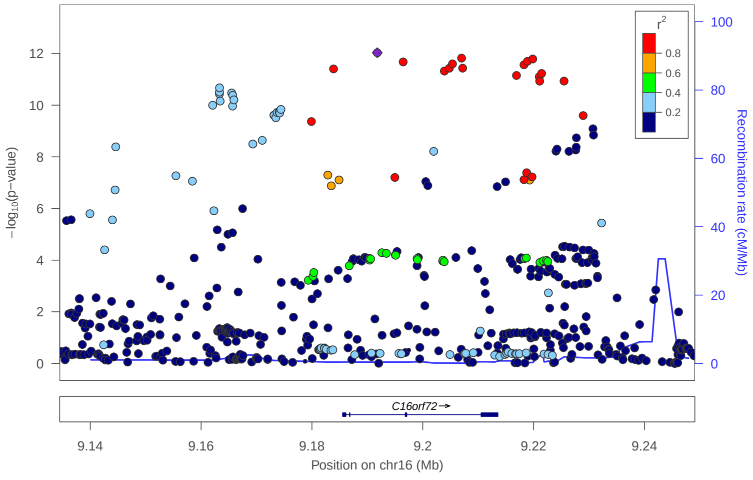


### locus 474 rs111505982

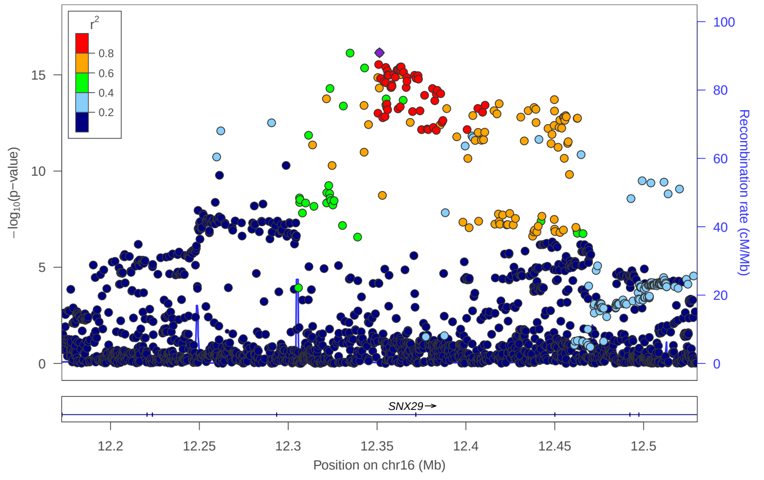

### locus 475 rs111536178

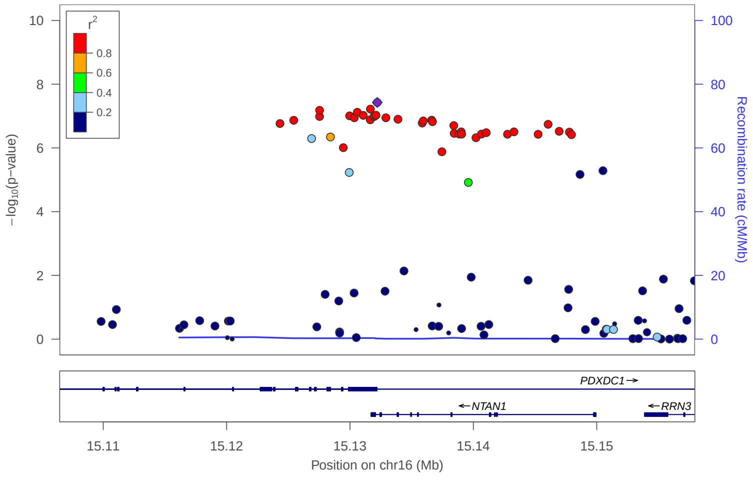


### locus 476 rs8054082

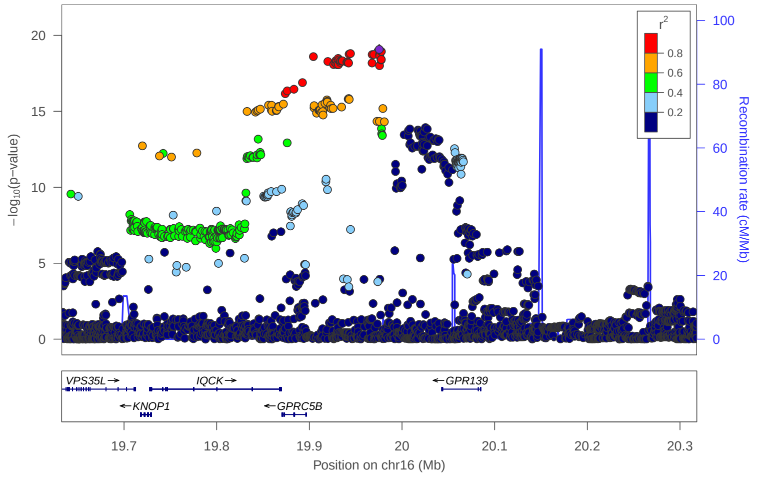


### locus 477 rs148514776

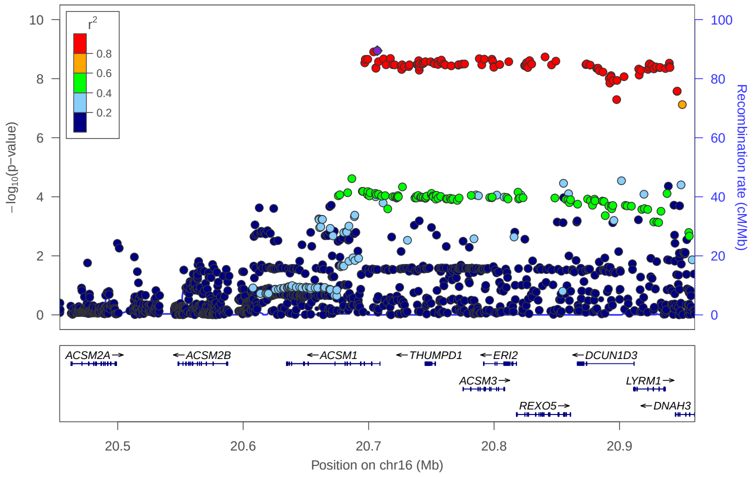


### locus 478 rs3814883

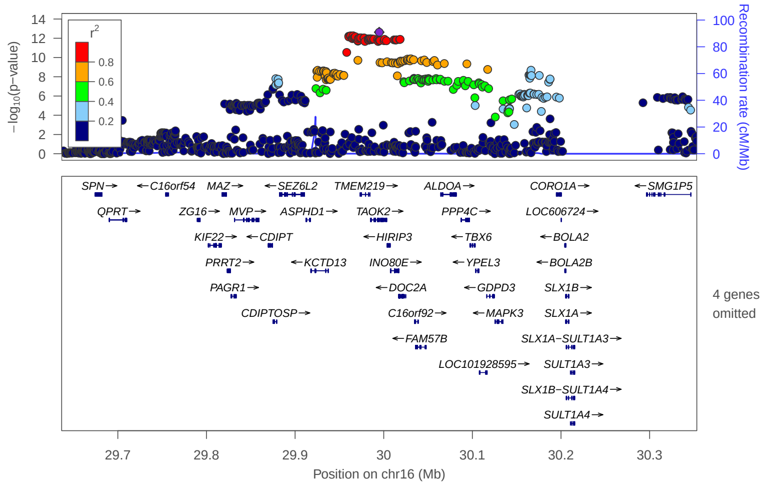


### locus 479 rs2356265

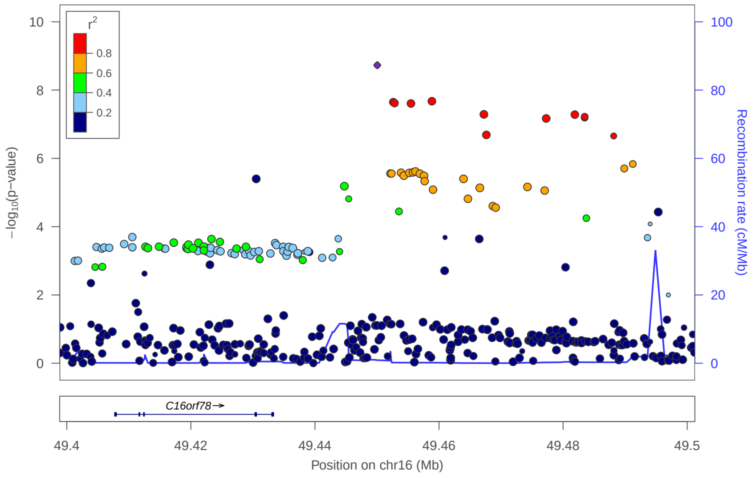


### locus 480 rs1015438

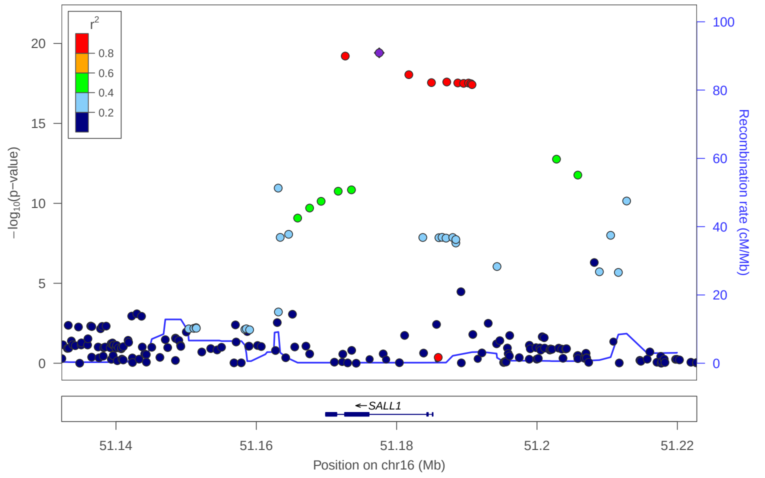


### locus 481 rs59937052

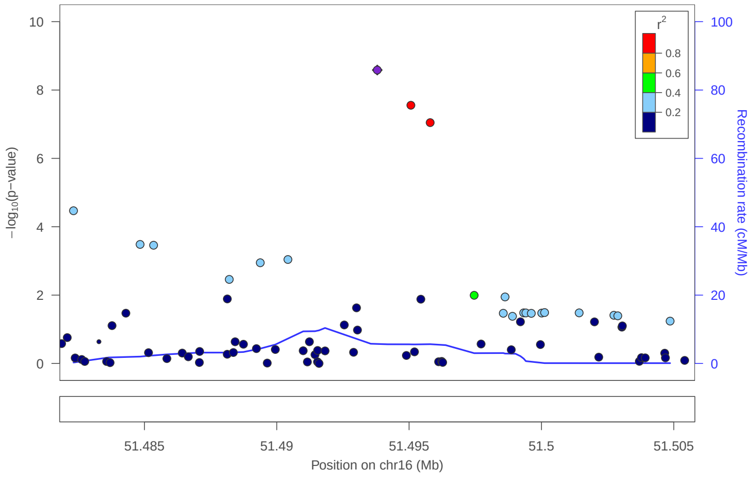


### locus 482 rs12927162

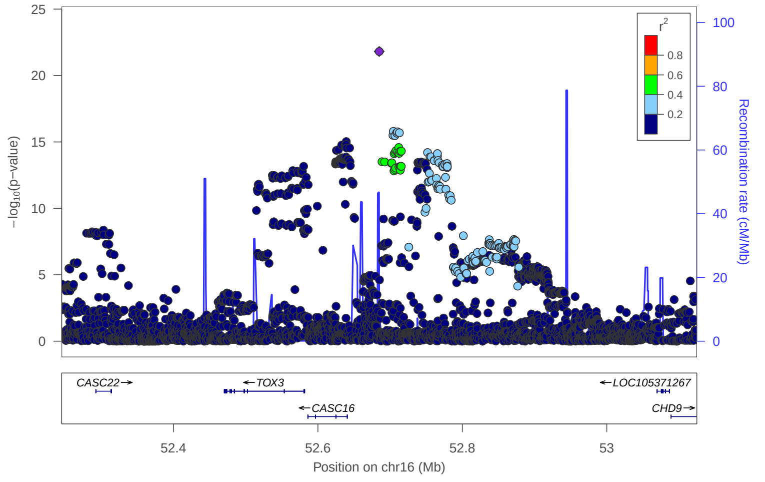

### locus 483 rs1548911

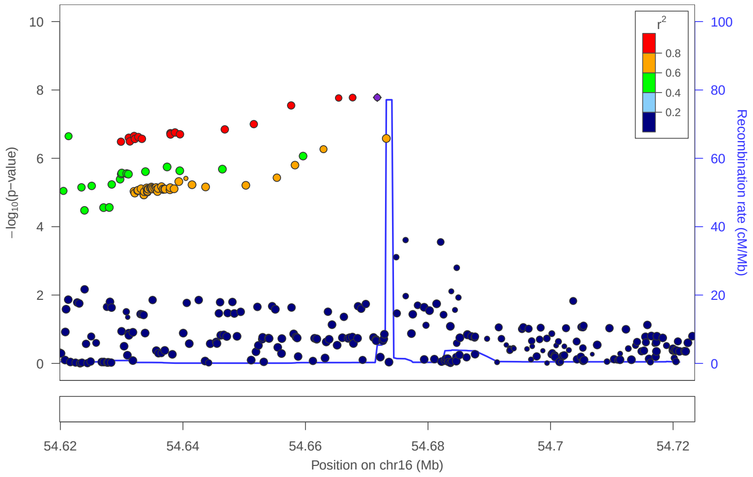


### locus 484 rs2398144

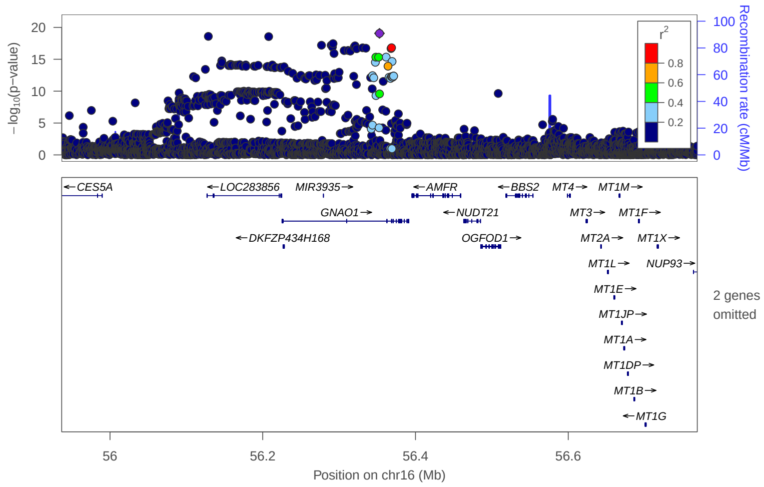


### locus 485 rs34160647

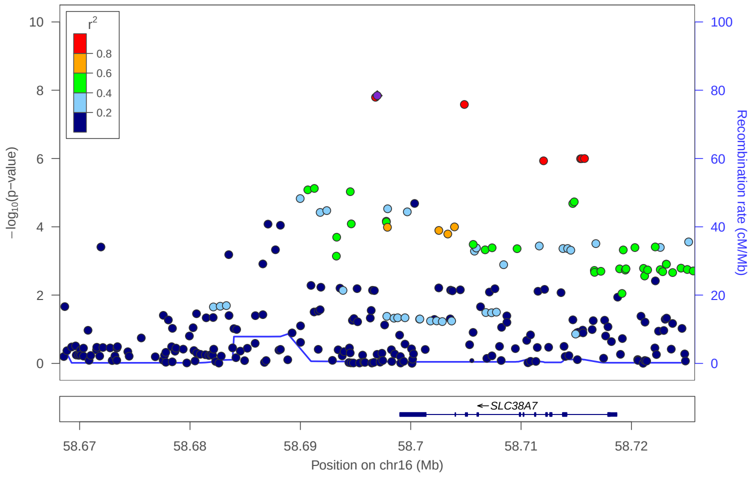


### locus 486 rs7203385

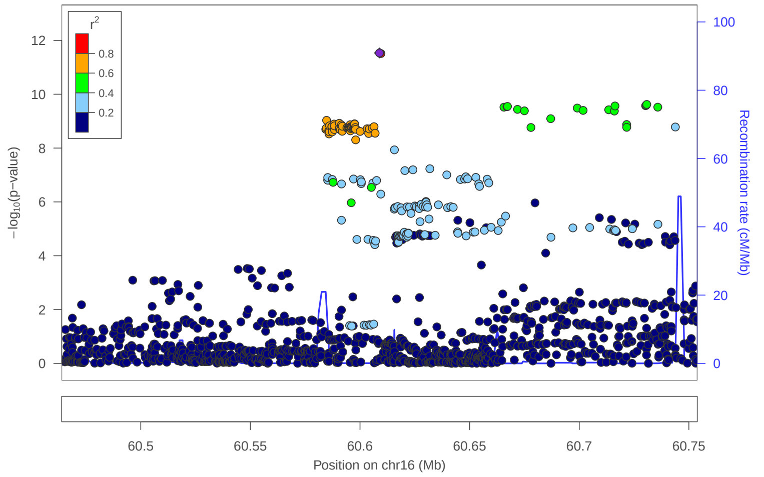


### locus 487 rs11863753

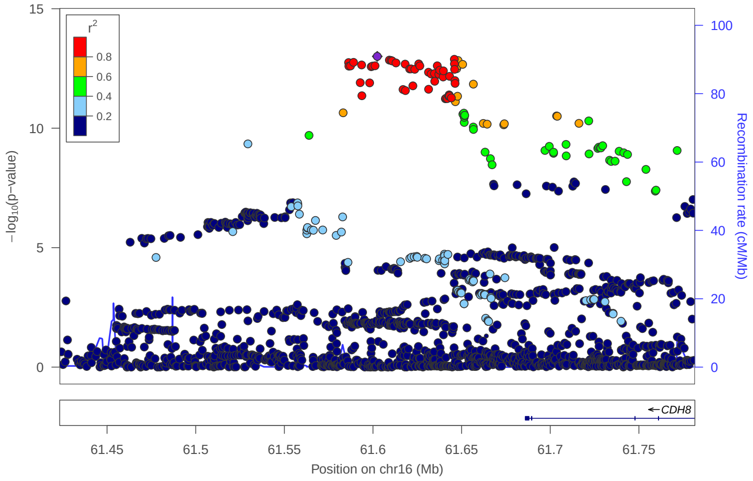


### locus 488 rs12598089

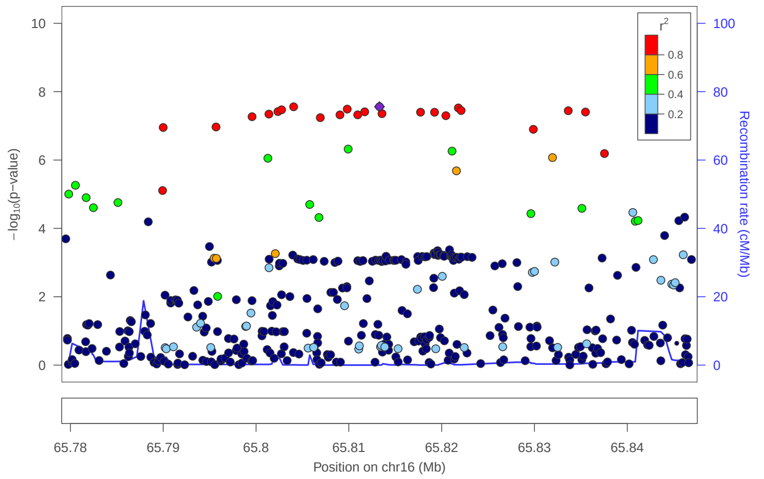


### locus 489 rs9926159

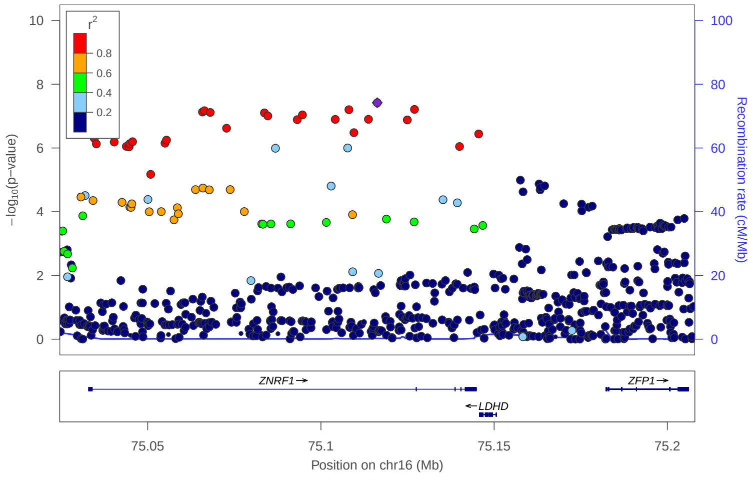


### locus 490 rs7184998

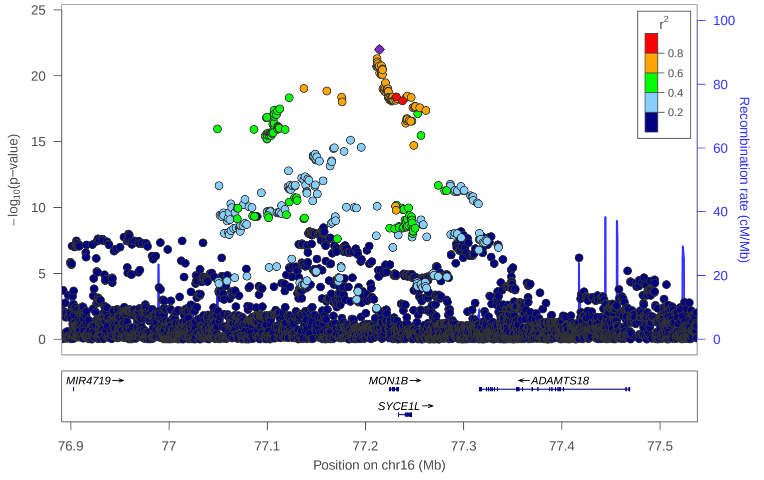

locus 491 rs79637296

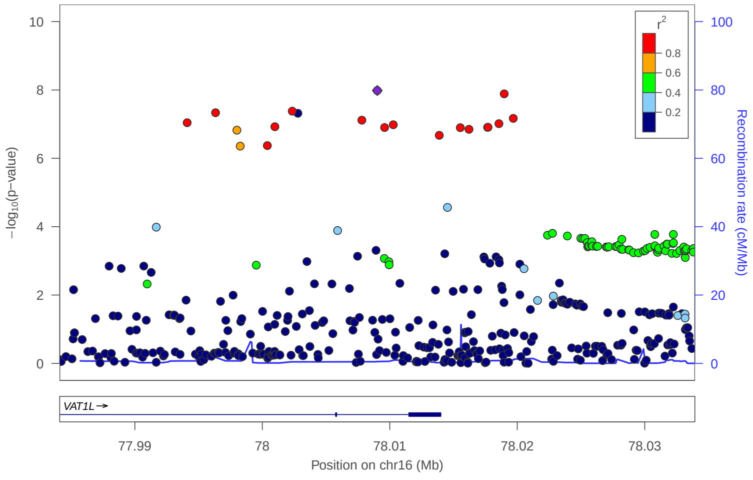


locus 492 rs11150494

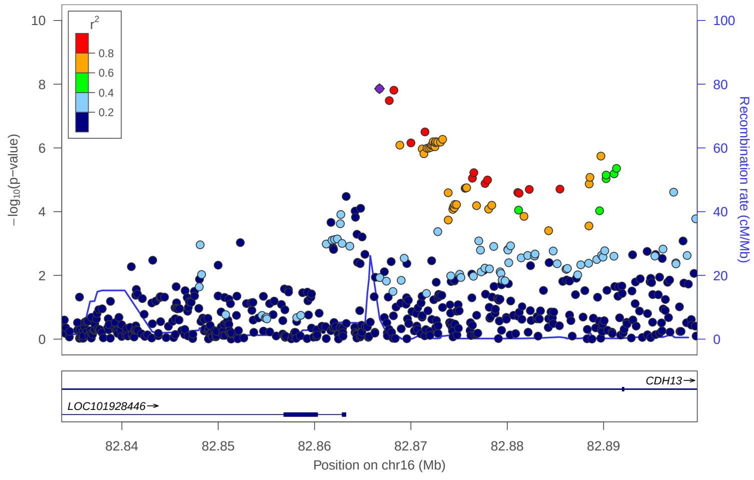


locus 493 rs8063871

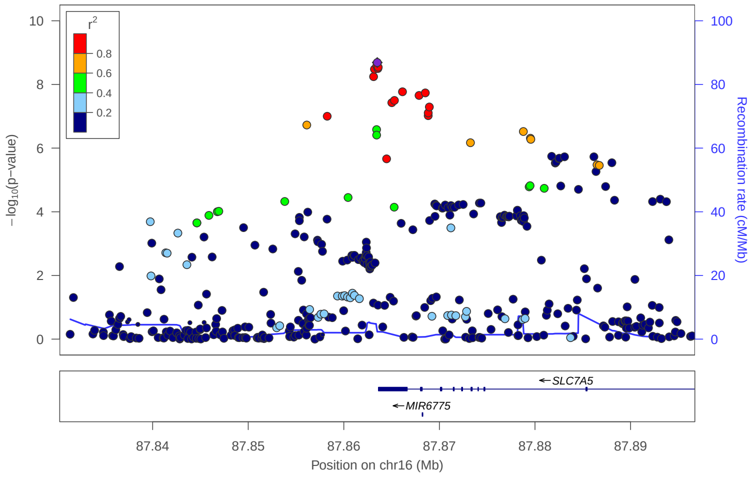


locus 494 rs8069356

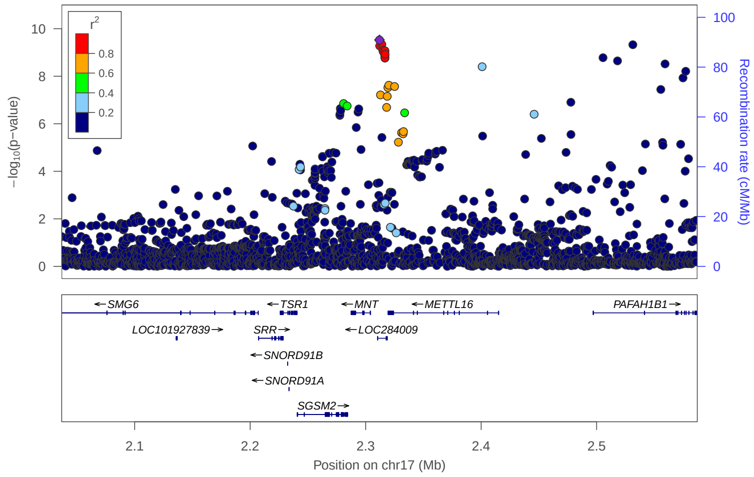


locus 495 rs8066969

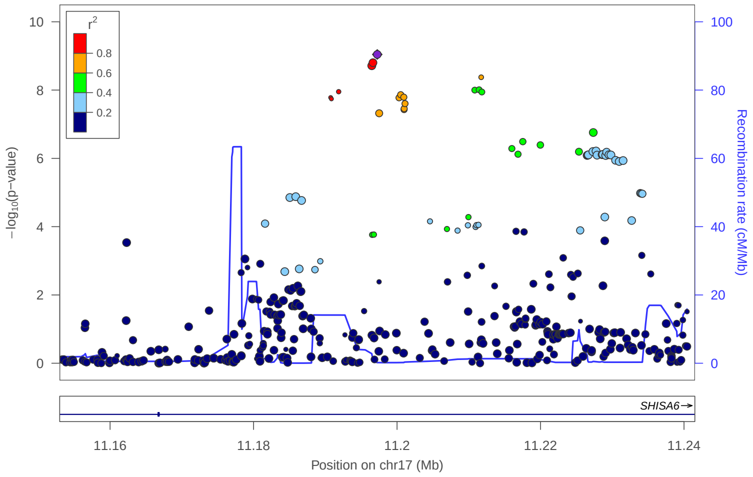


locus 496 rs2232839

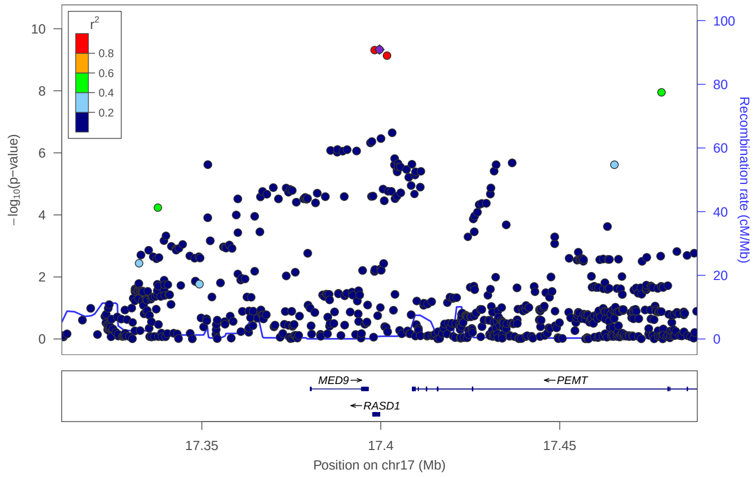


locus 497 rs34490907

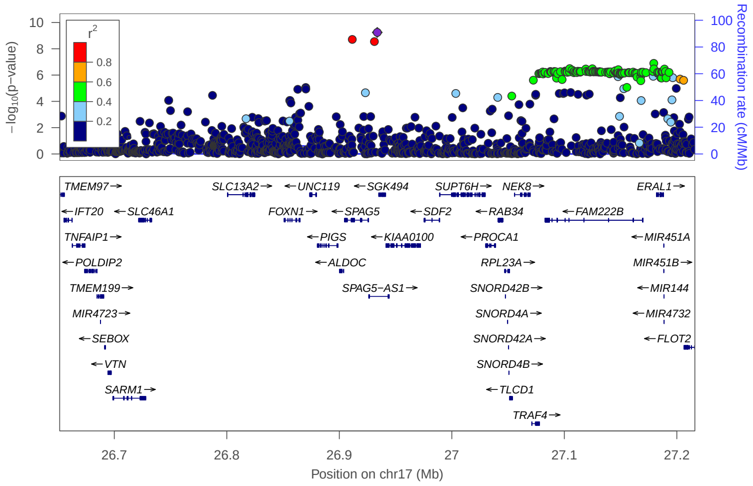


locus 498 rs8076005

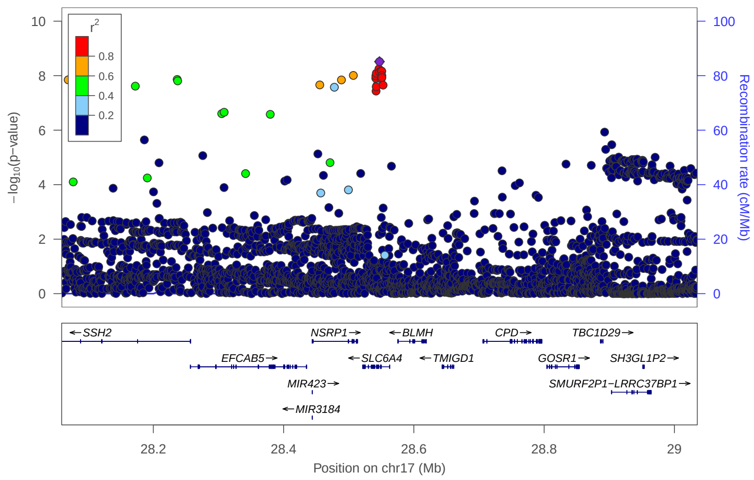

### locus 499 rs3815156

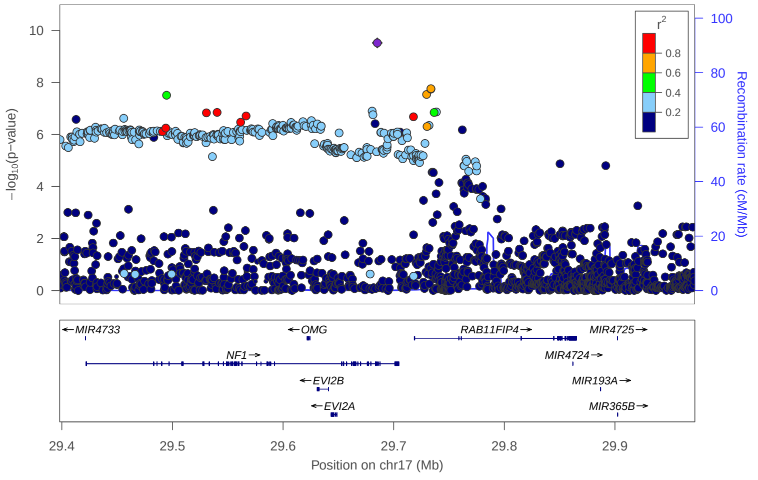


### locus 500 rs12947676

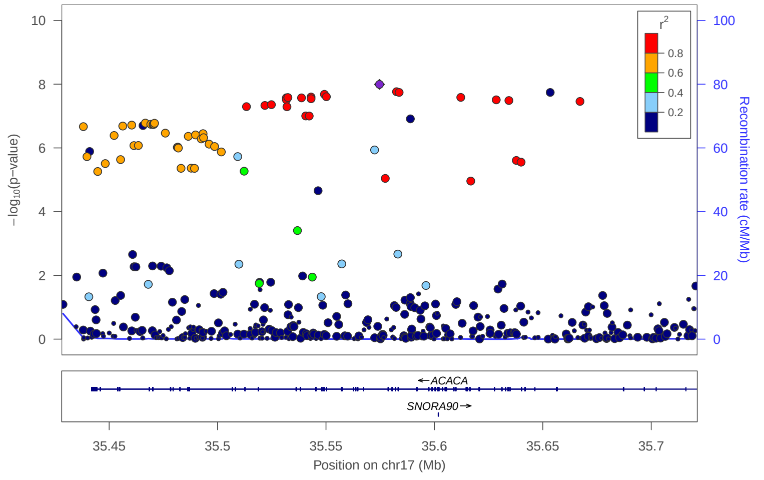


### locus 501 rs11871043

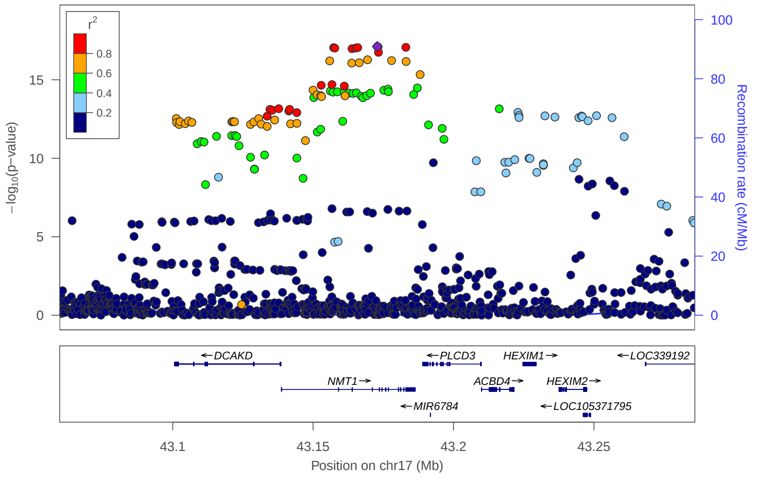


### locus 502 rs4643373

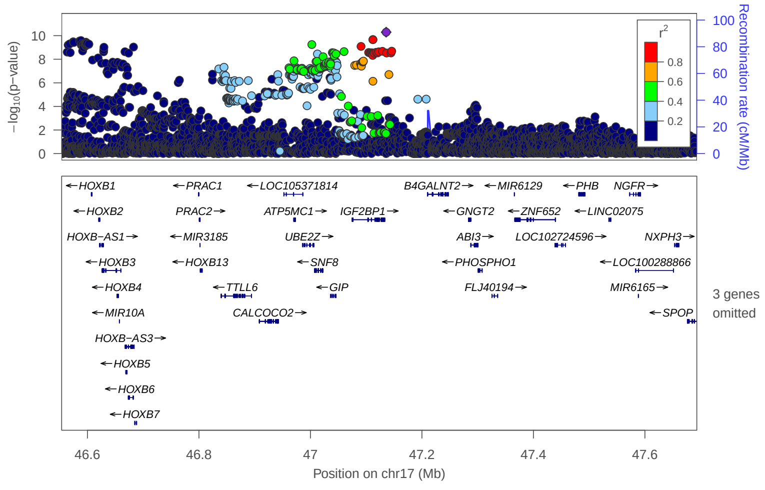


### locus 503 rs9889282

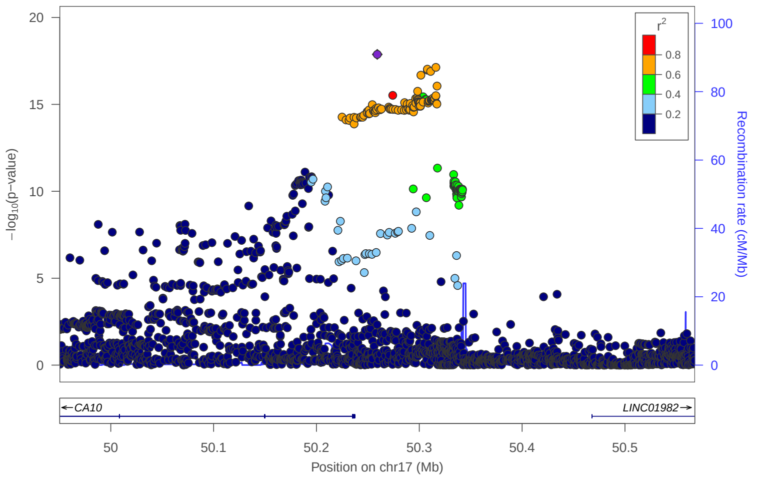


### locus 504 rs8076183

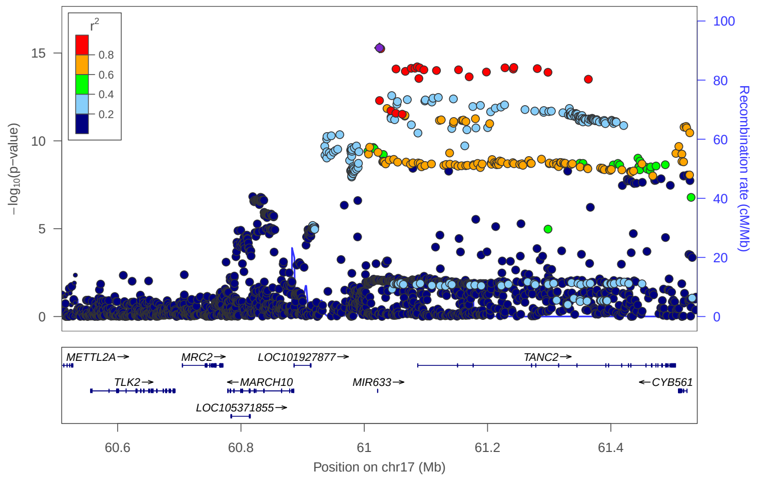


### locus 505 rs6504568

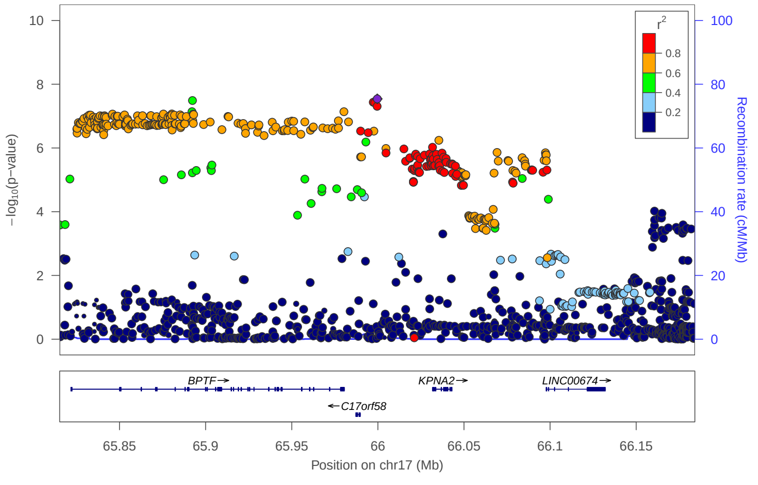


### locus 506 rs323406

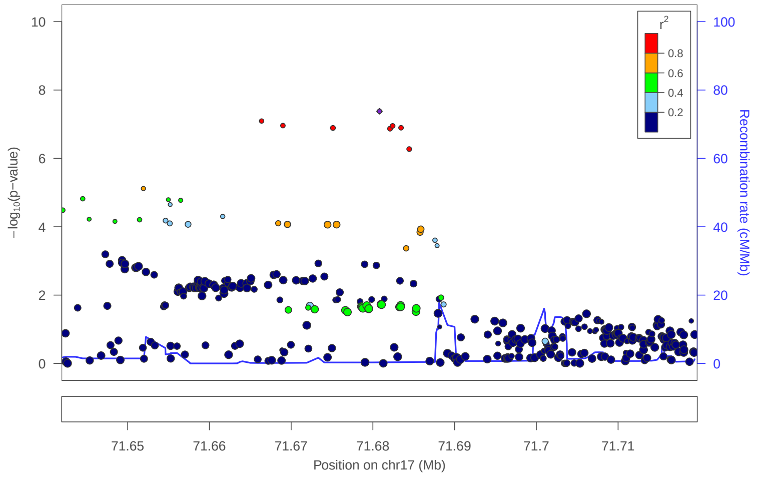

locus 507 rs56324203

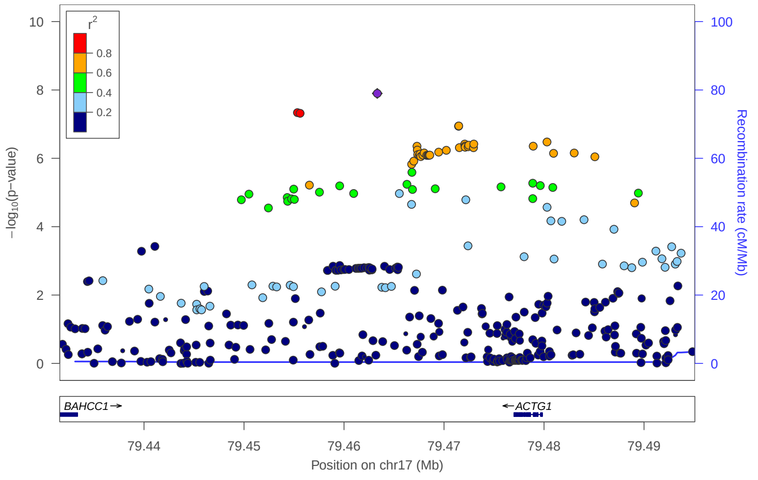


locus 508 rs8074498

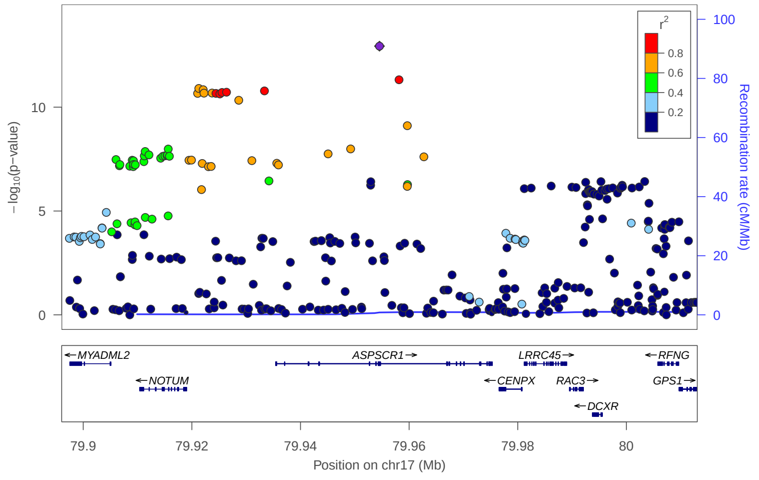


locus 509 rs113722141

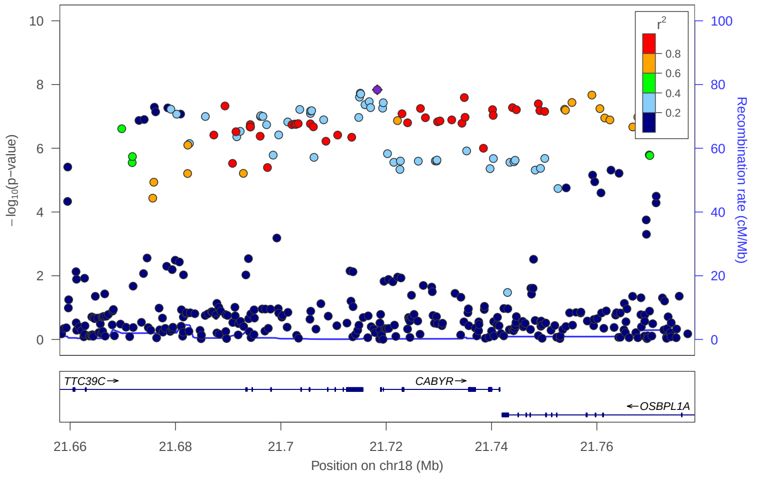


locus 510 rs4432322

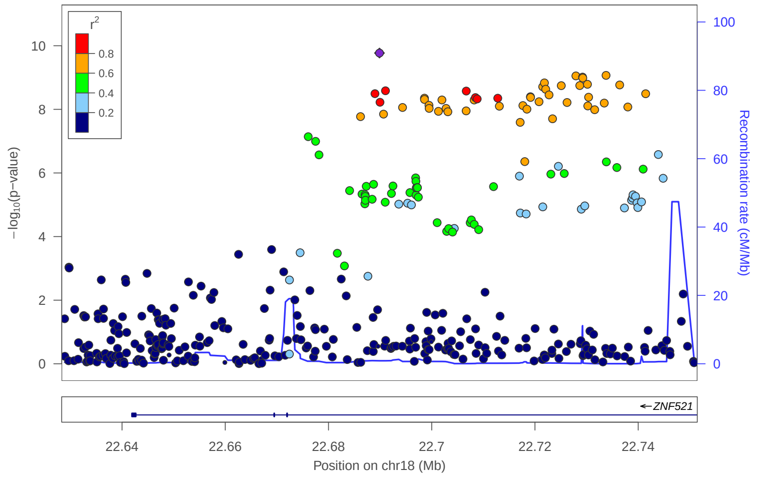


locus 511 rs790316

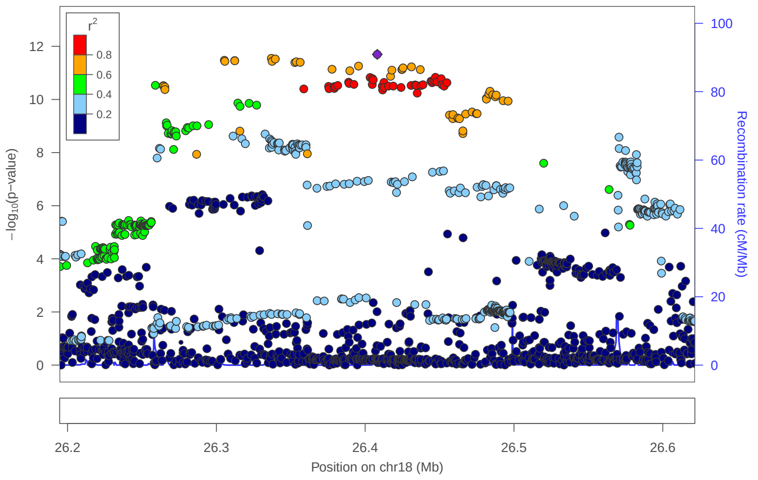


locus 512 rs12605642

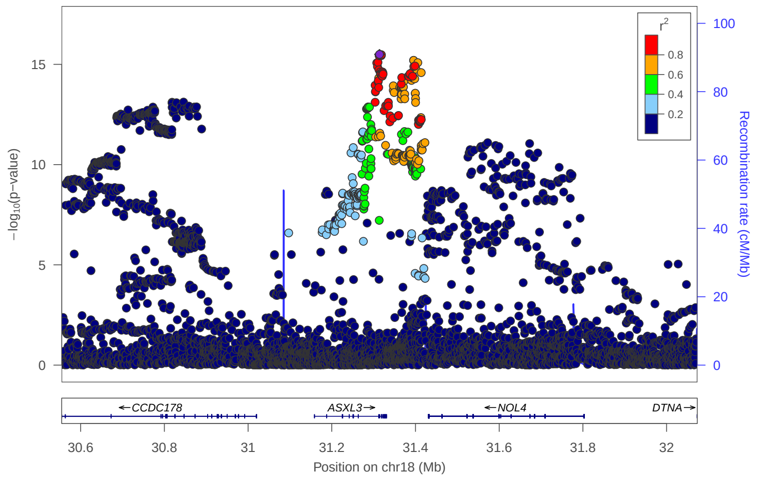


locus 513 rs62092949

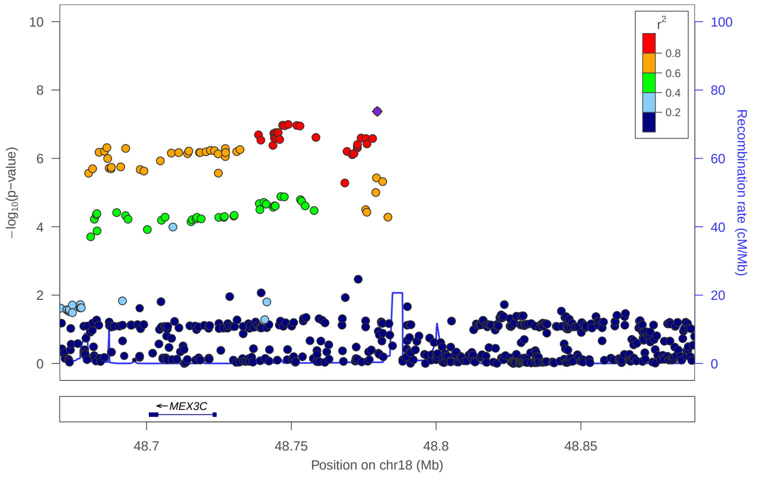


locus 514 rs35541683

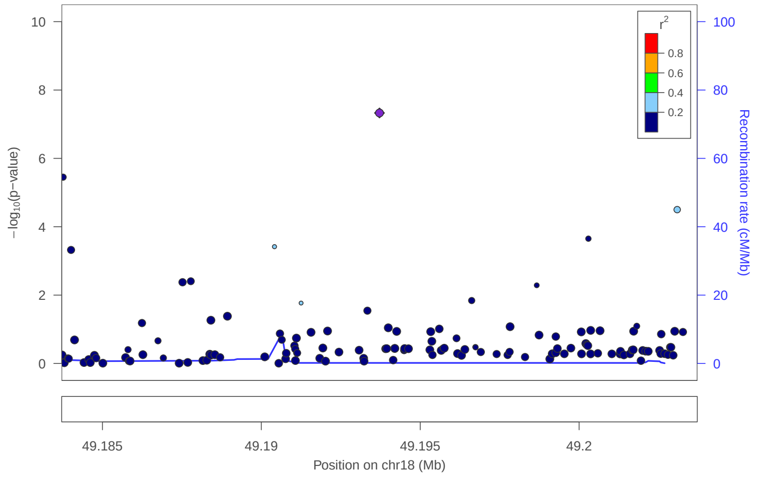

locus 515 rs10502966

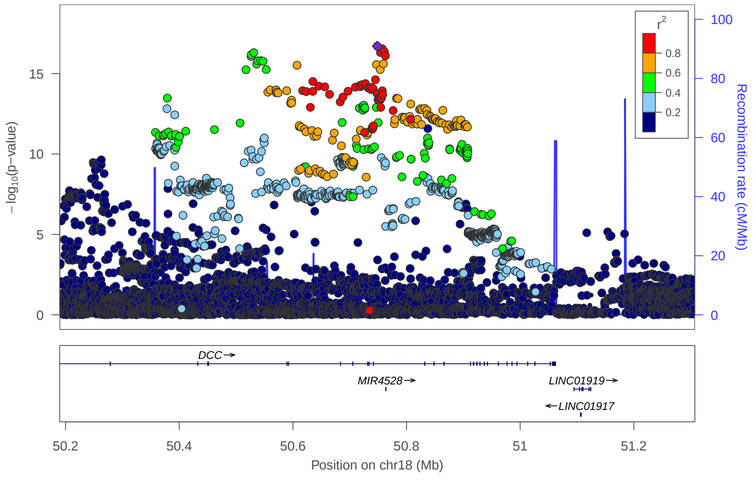


locus 516 rs8766

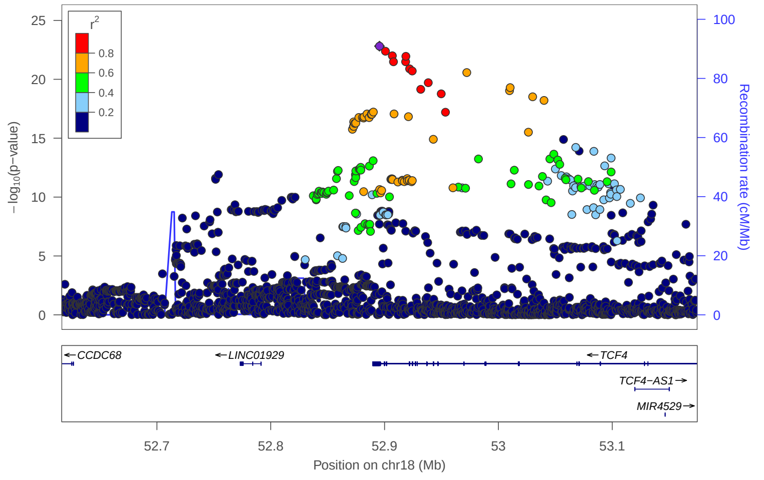


locus 517 rs2271731

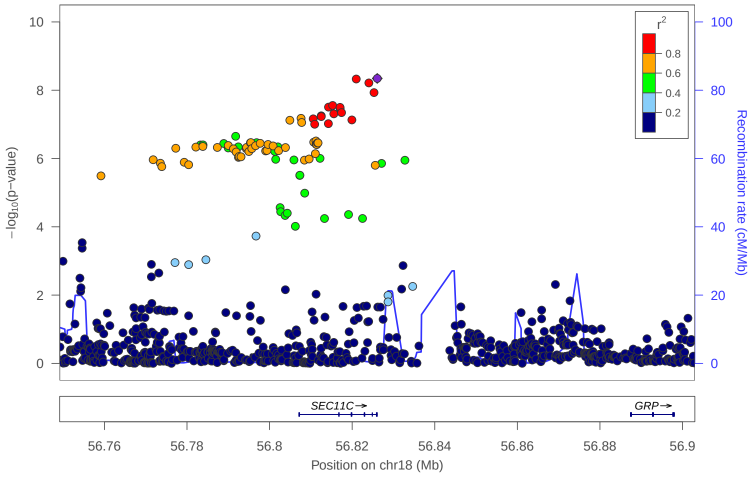


locus 518 rs8084405

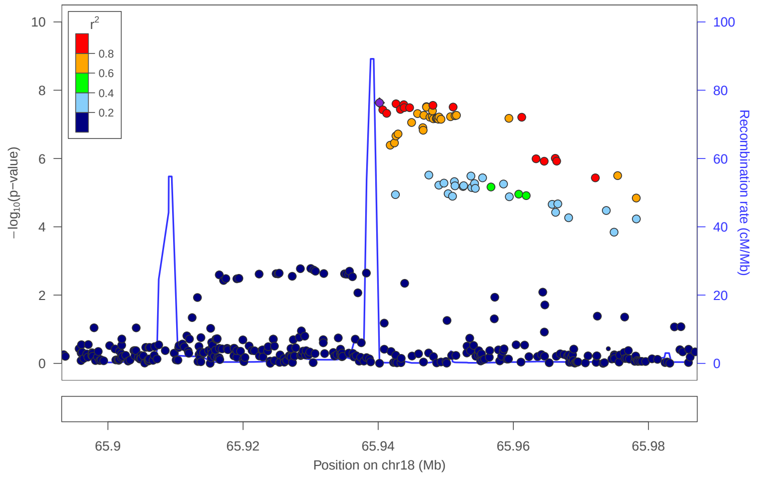


locus 519 rs516890

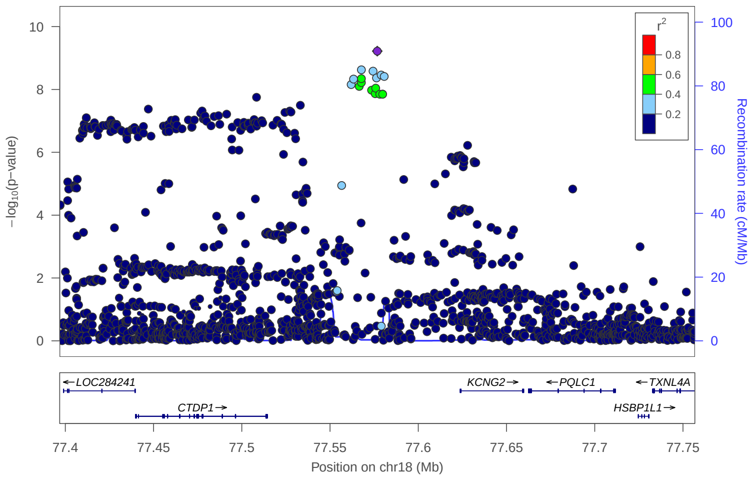


locus 520 rs12983032

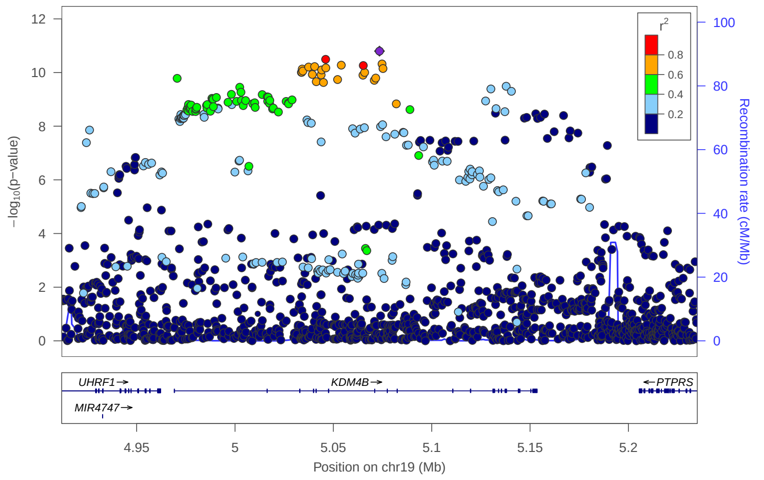


locus 521 rs12611068

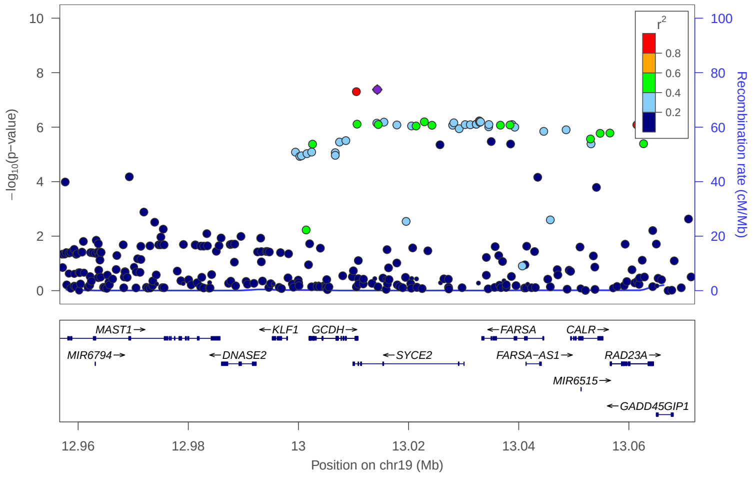


locus 522 rs12973258

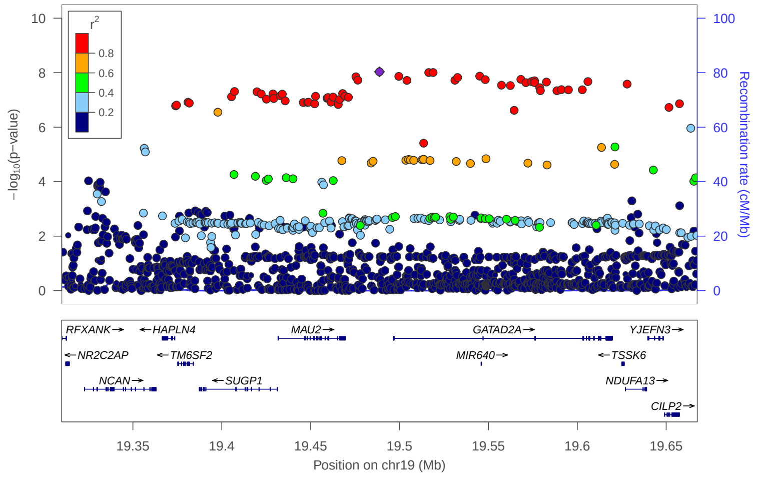

locus 523 rs11375063

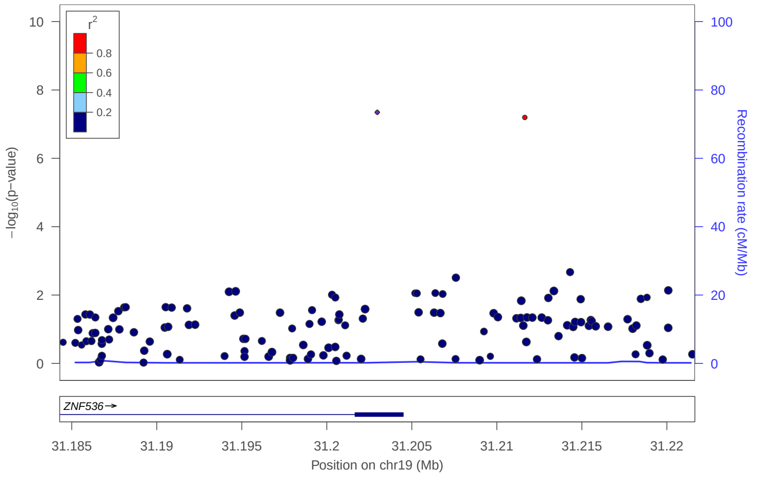


locus 524 rs10421599

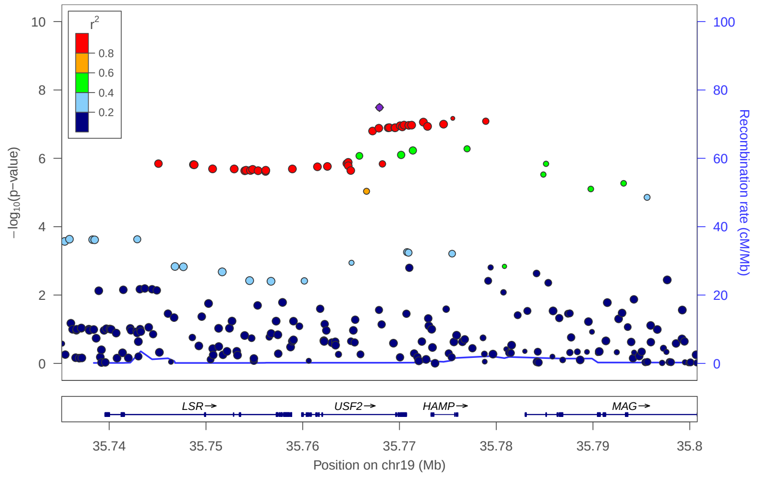


locus 525 rs73034109

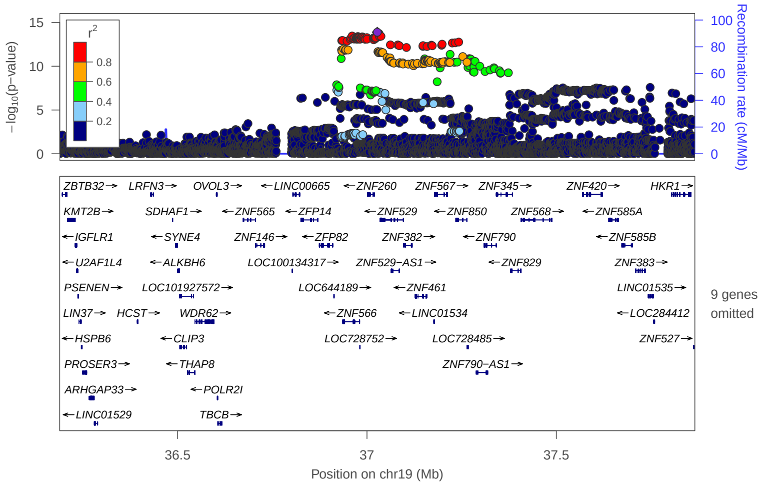


locus 526 rs4803217

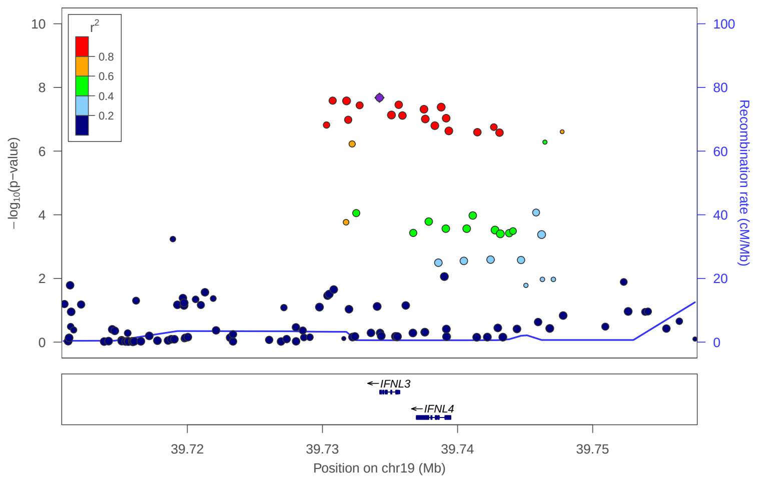


locus 527 rs429358

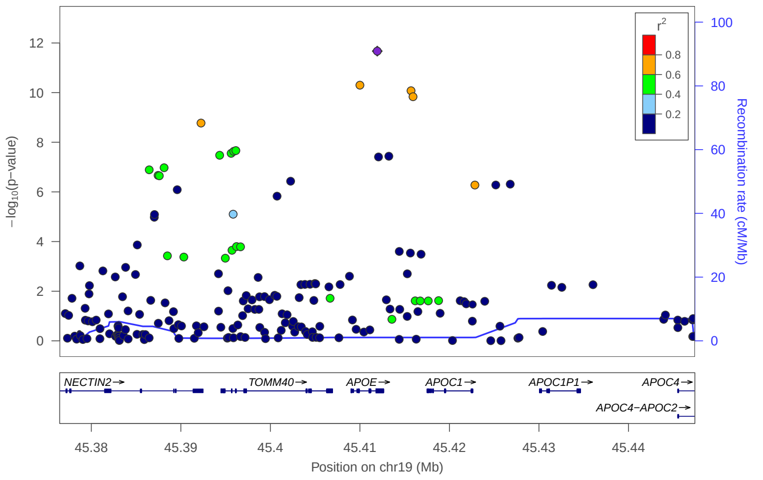


locus 529 rs57314044

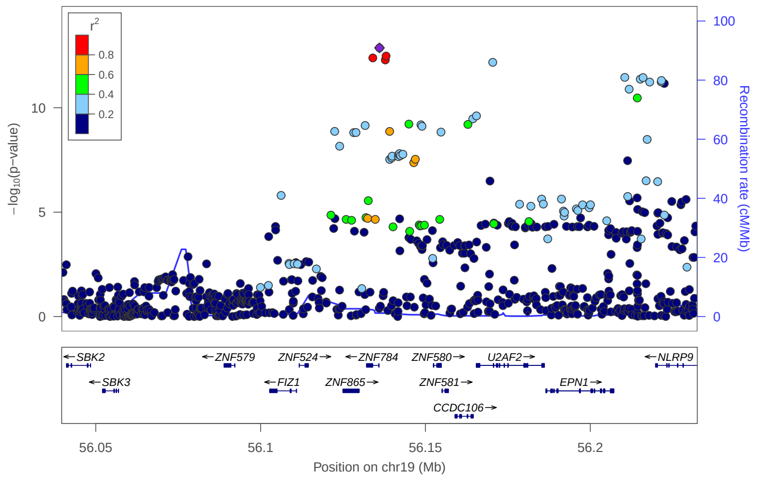


locus 530 rs8110135

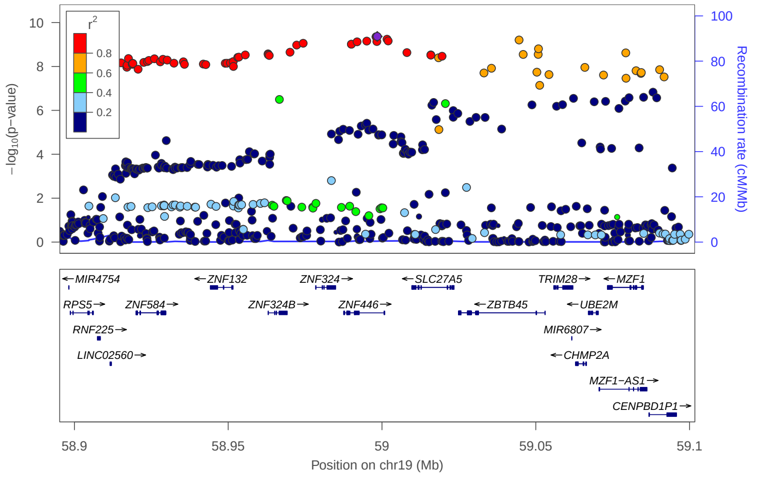


locus 531 rs56353032

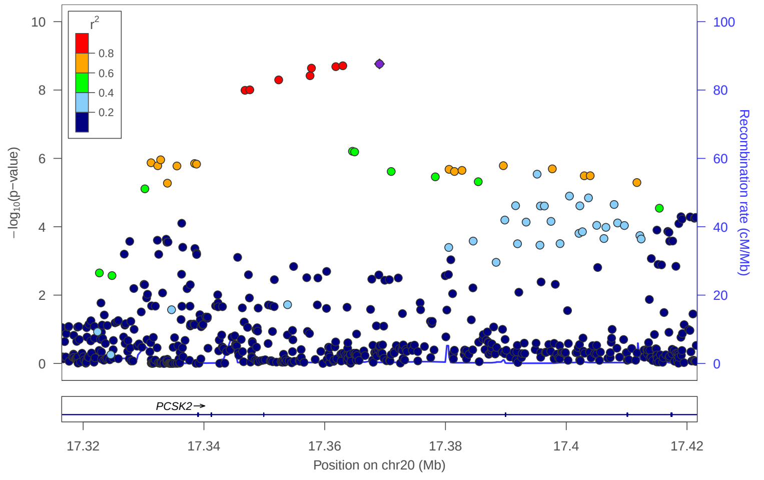

locus 532 rs6035826

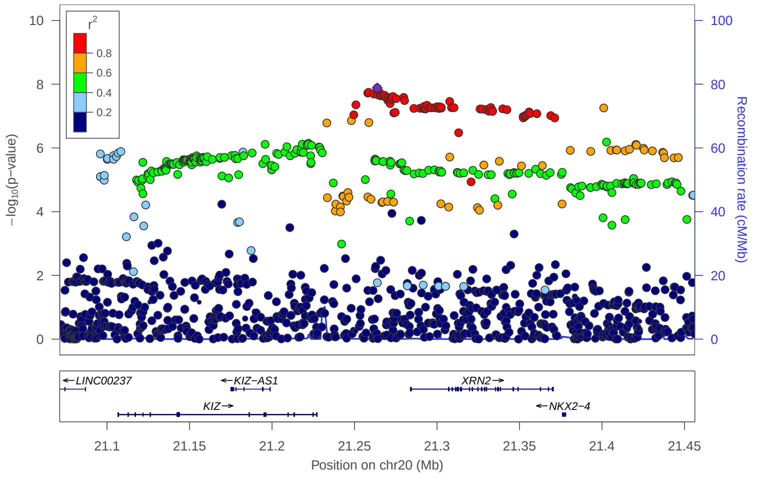


locus 533 rs760997

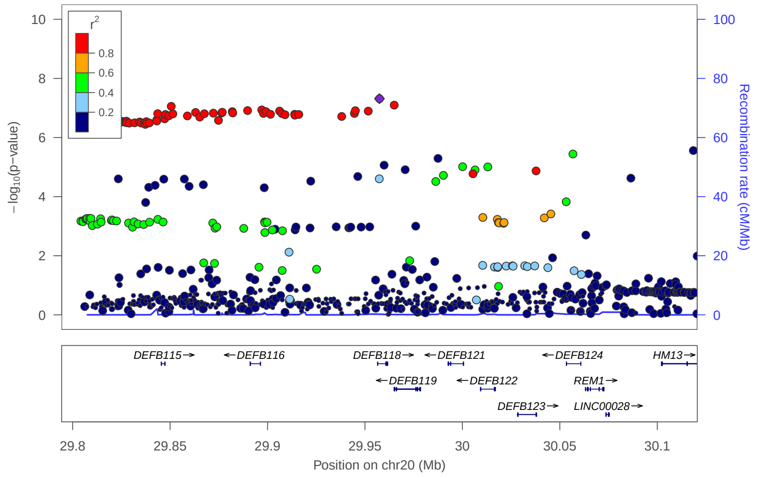


locus 534 rs293567

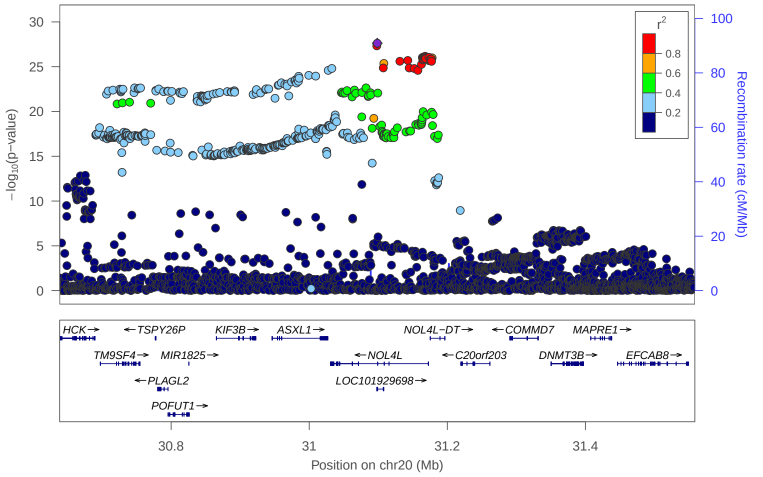


locus 535 rs13038787

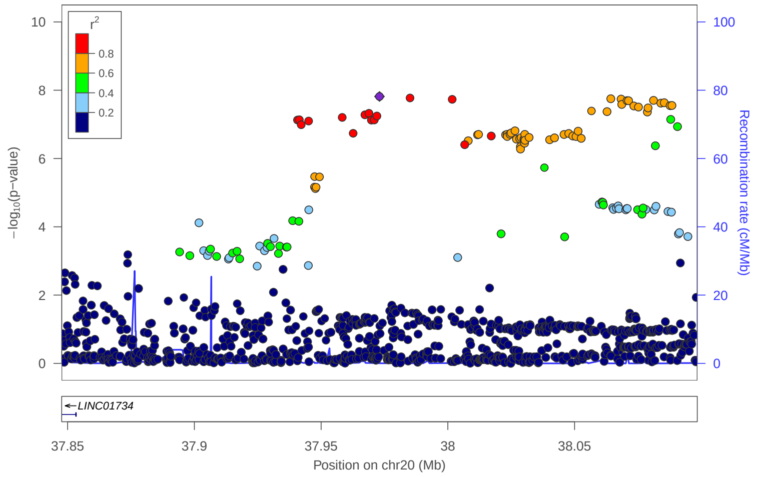


locus 536 rs6030790

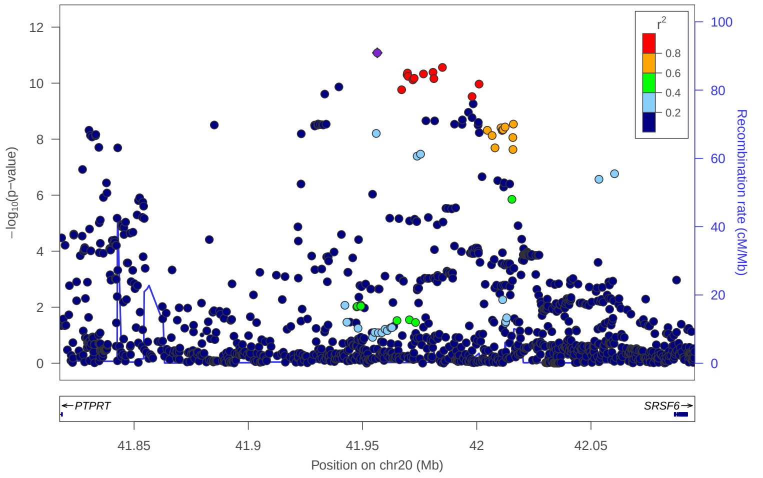


locus 537 rs1883832

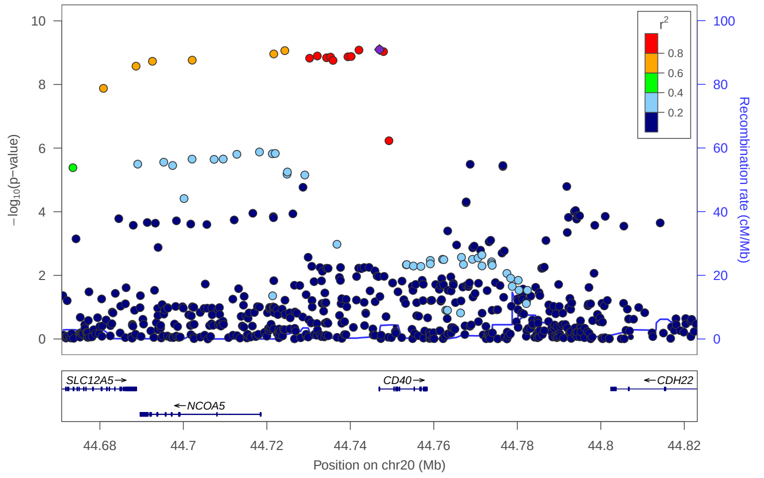


locus 538 rs910187

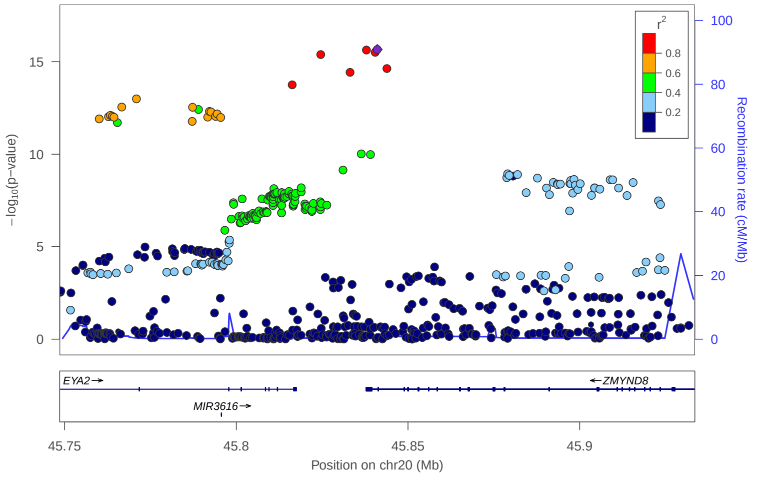


locus 539 rs6019663

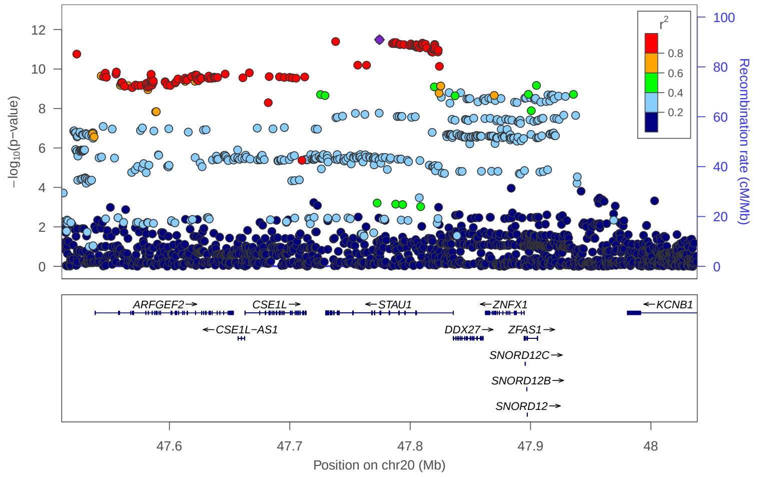

locus 540 rs13037010

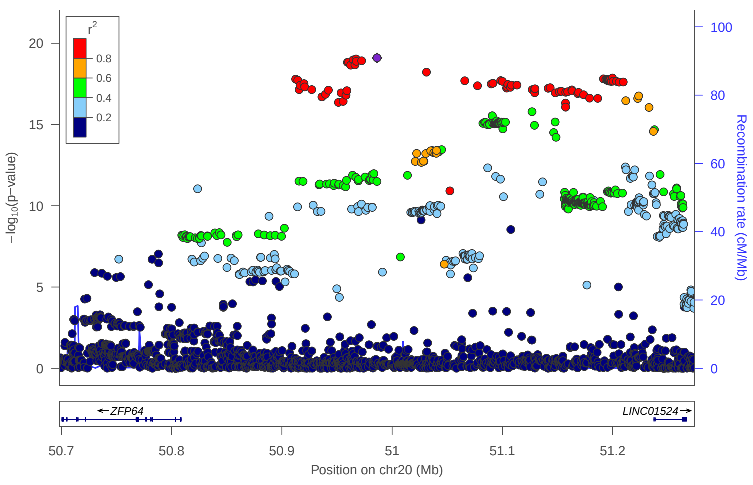


locus 541 rs6024441

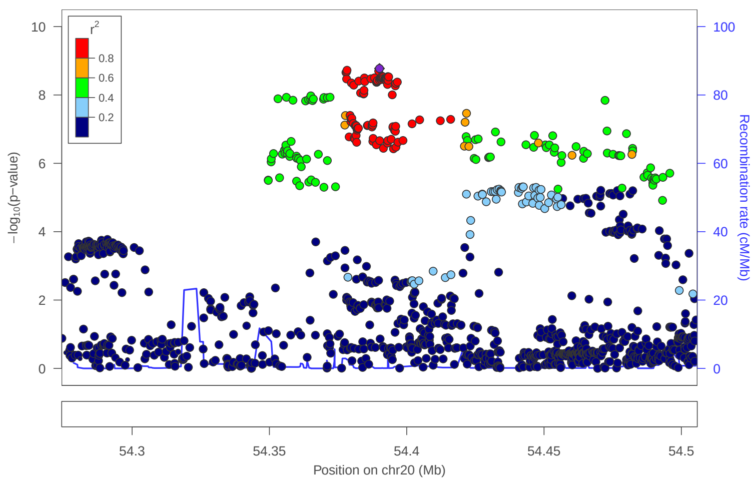


locus 542 rs2236202

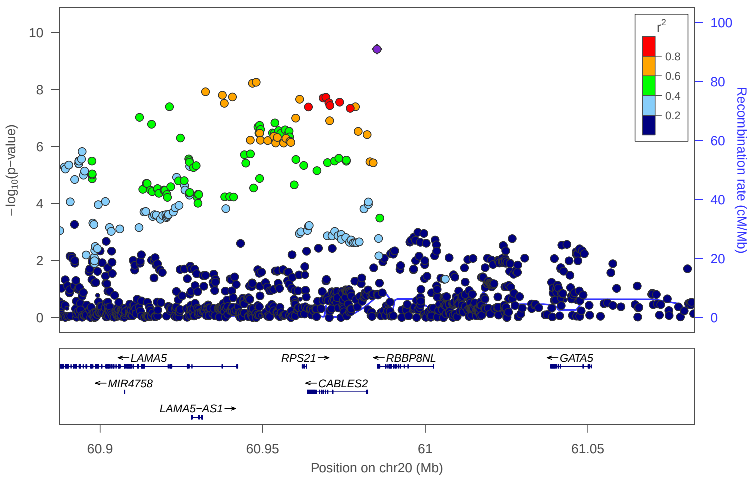


locus 543 rs67047102

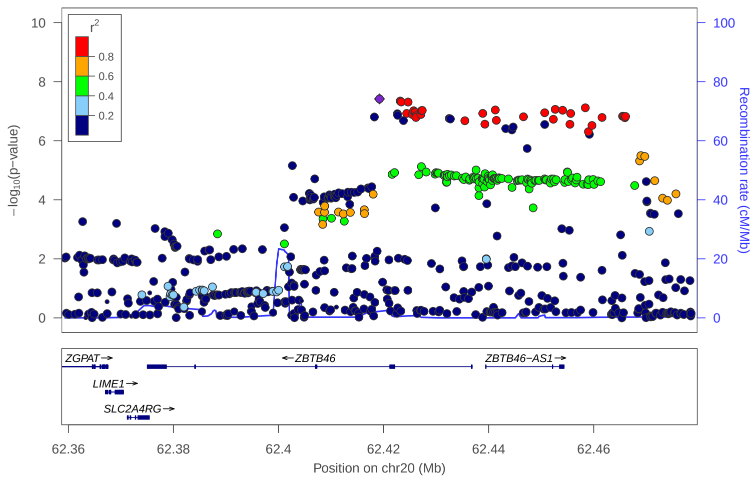


locus 544 rs965494

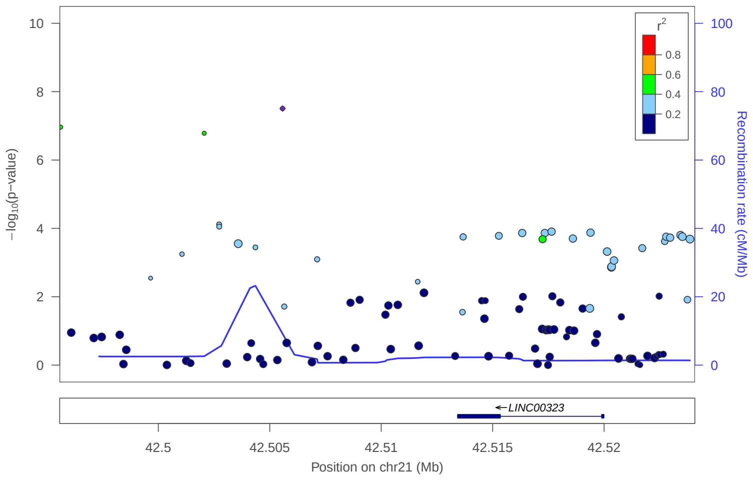


locus 545 rs10368

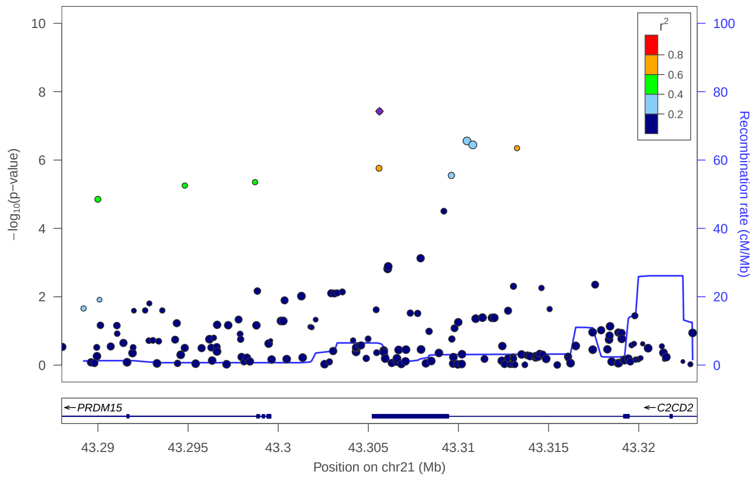


locus 546 rs2003273

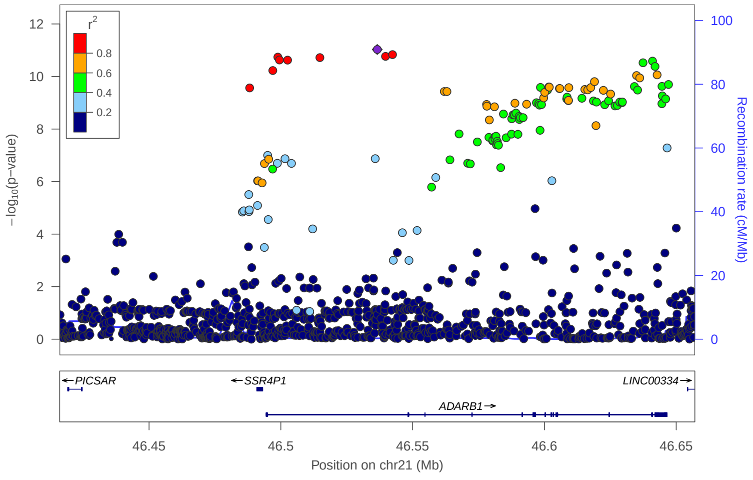


locus 547 rs147970828

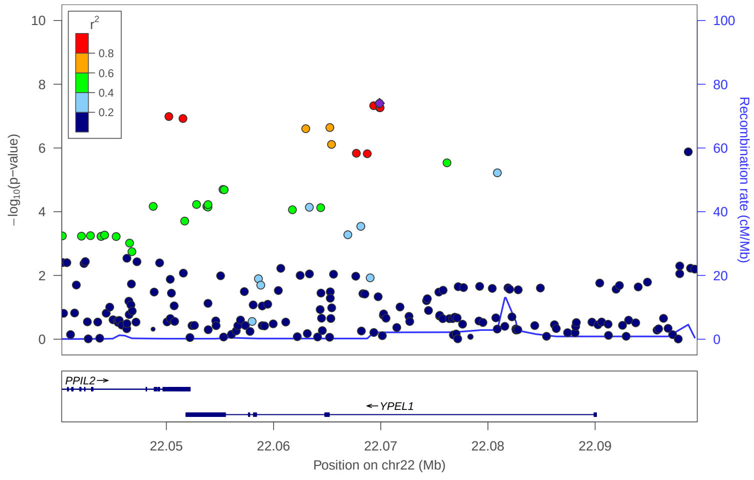

locus 548 rs136310

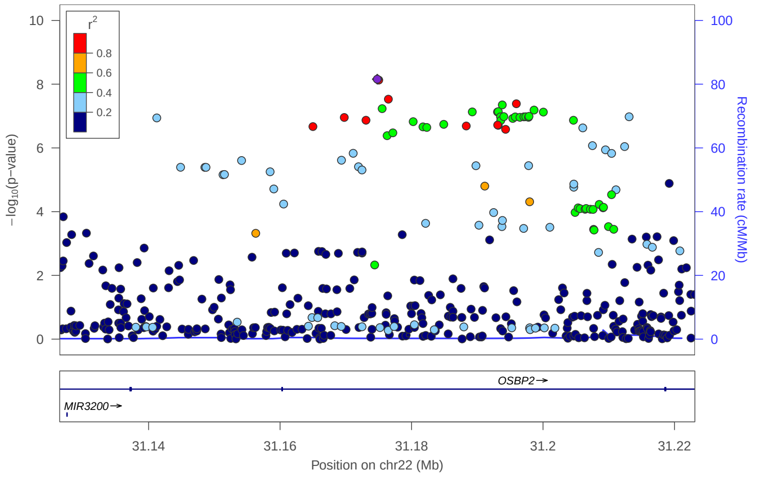


locus 549 rs9607581

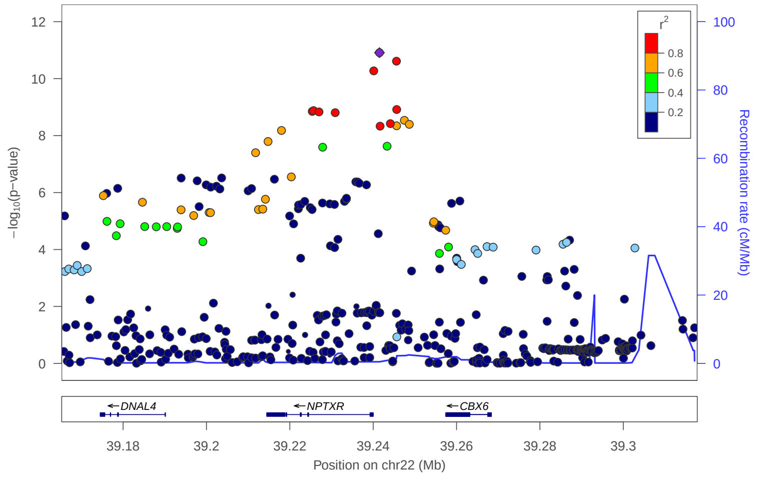


locus 550 rs145611631

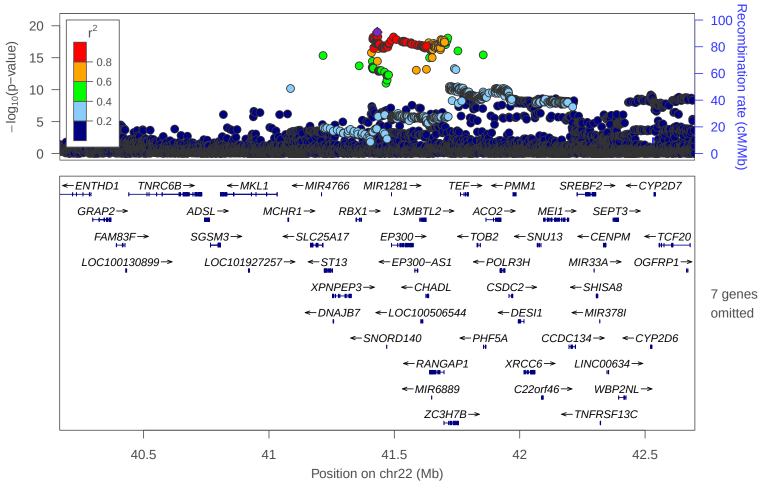


locus 551 rs12010000

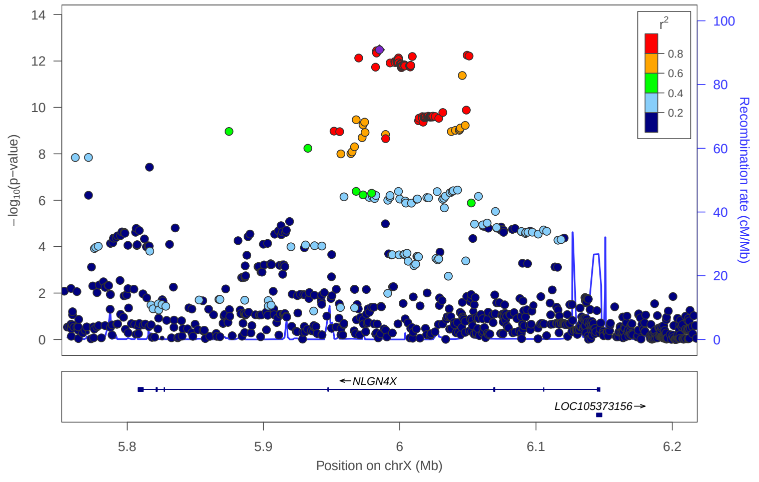


locus 552 rs62590551

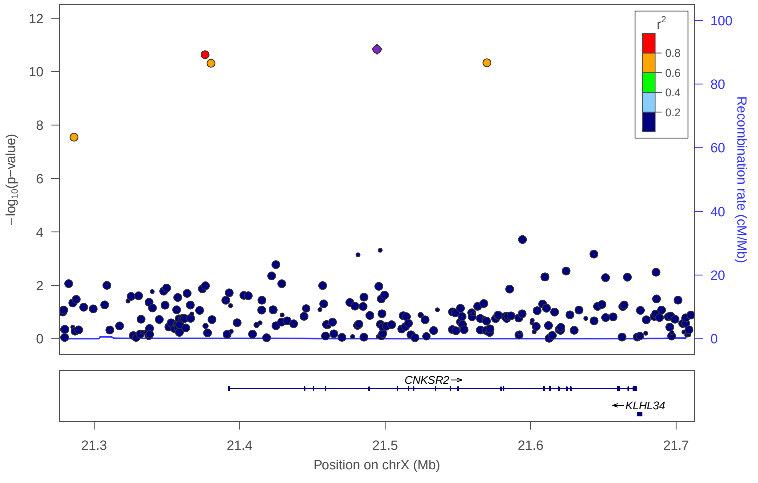


locus 553 rs6629928

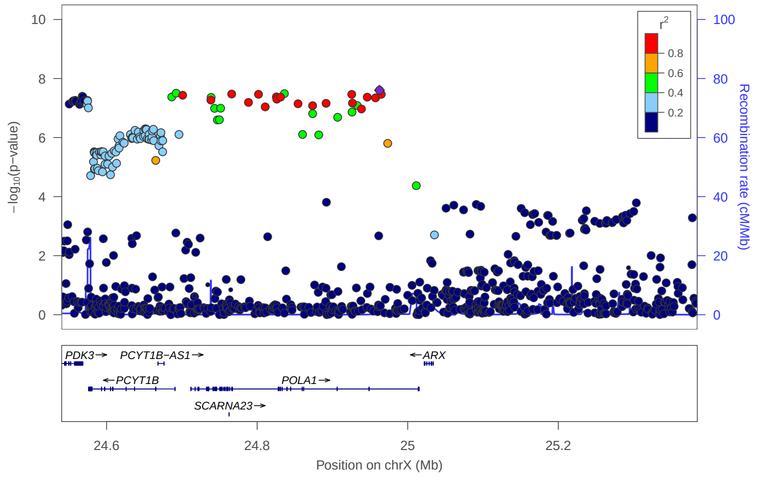


locus 555 rs4828253

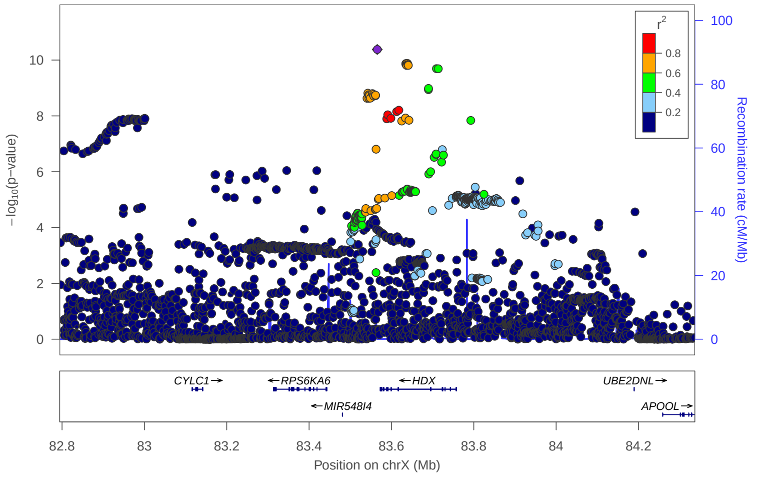


locus 556 rs5932880

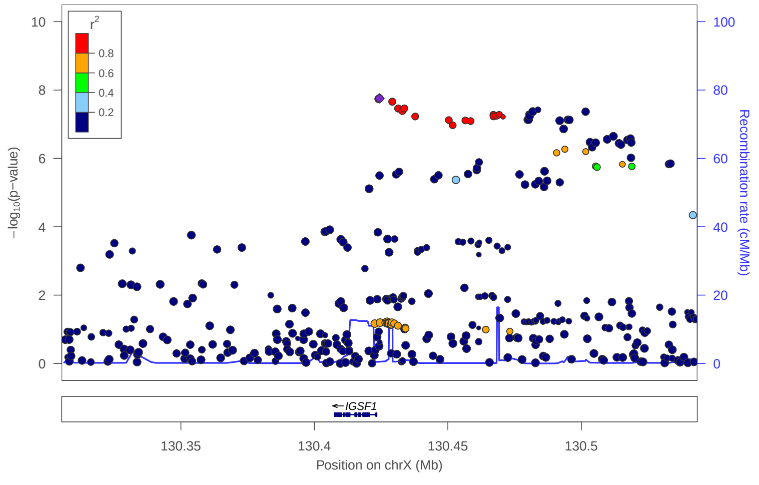

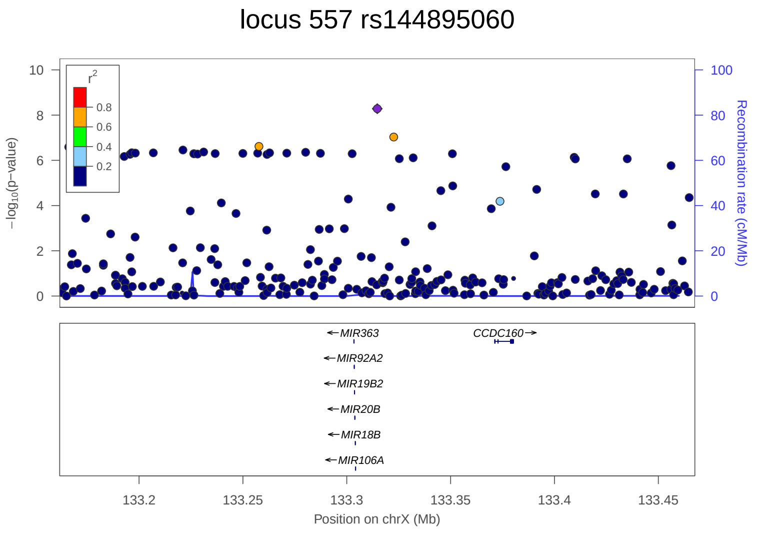

locus 557 rs144895060
r2
0.8
0.6
0.4
0.2
−log10(p−value)
Recombination rate (cM/Mb)
MIR363
CCDC160
MIR92A2
MIR19B2
MIR20B
MIR18B
MIR106A
133.2
133.25
133.3
133.35
133.4
133.45
Position on chrX (Mb)


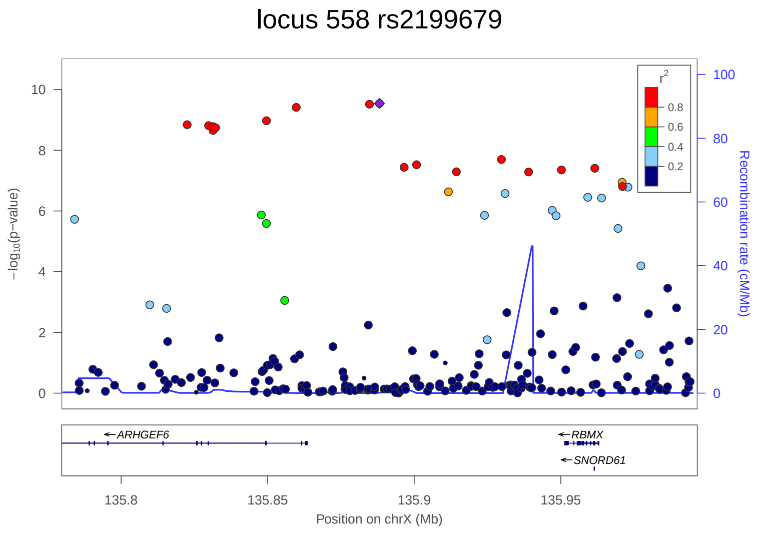

locus 558 rs2199679
r2
0.8
0.6
0.4
0.2
−log10(p−value)
Recombination rate (cM/Mb)
ARHGEF6
RBMX
SNORD61
135.8
135.85
135.9
135.95
Position on chrX (Mb)
