## Supplementary figures and images for "Genome-wide meta-analysis of insomnia in over 2.3 million individuals implicates involvement of specific biological pathways through gene-prioritization"

### Suppl Fig 3

locus 37 rs138122366

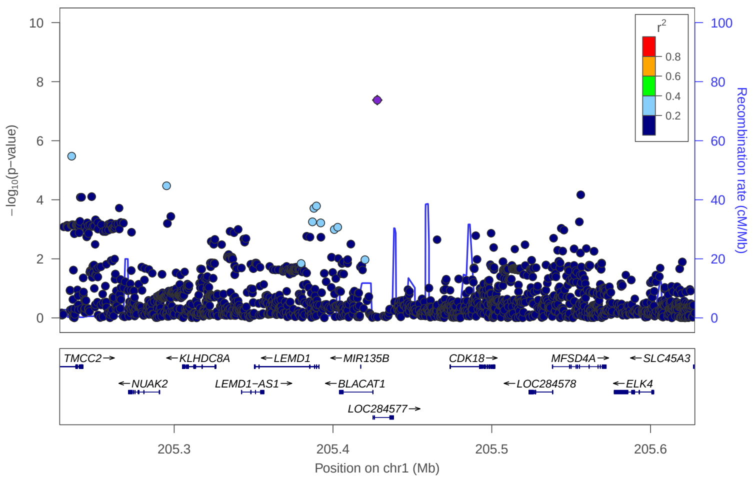


locus 227 rs2076060

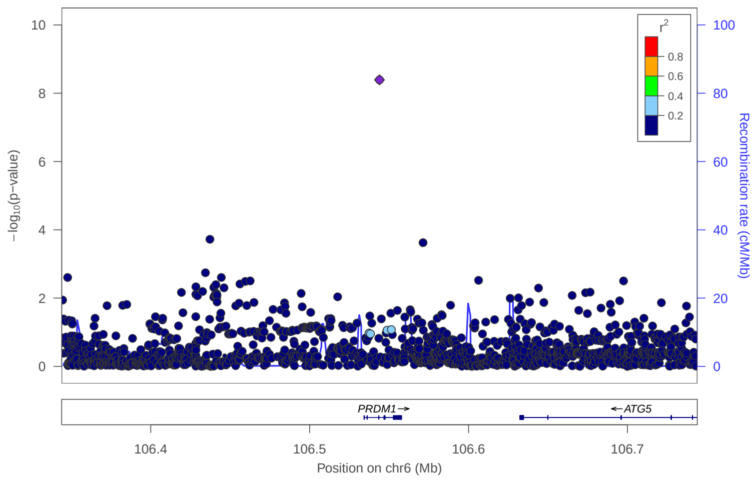


locus 528 rs10406993

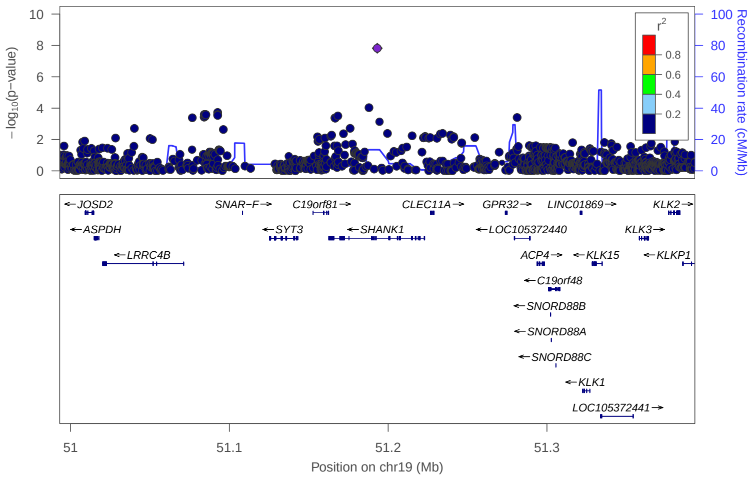


locus 554 rs17315269

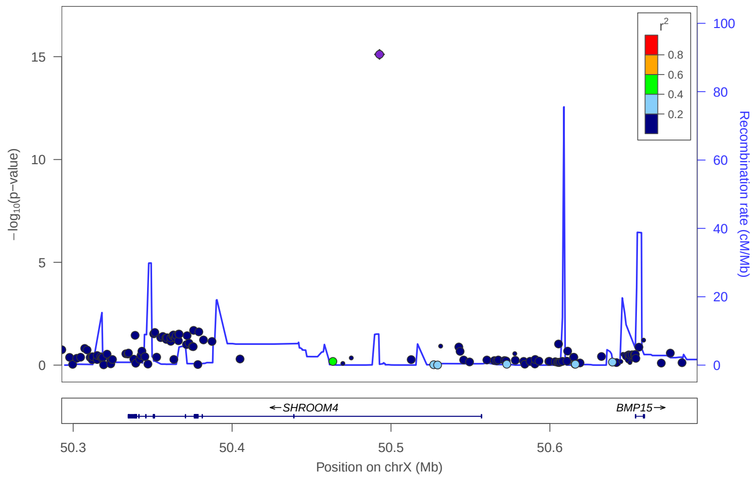
